# Cardiovascular outcomes and fracture risk after the discontinuation of preventative medications in older patients with complex health needs: a self-controlled case series analysis

**DOI:** 10.1101/2024.03.08.24303980

**Authors:** Daniel Prieto-Alhambra, Francesco Dernie, Antonella Delmestri, Trishna Rathod-Mistry, Eng Hooi Tan, Annika M. Jödicke

## Abstract

**Objective:** To assess the effect of stopping statins, antihypertensives, and bisphosphonates on the risk of cardiovascular events and fractures in older patients with complex health needs (CHN).

**Methods:** Patients aged >65 years, registered in CPRD GOLD for ≥1 year before study start (01/01/2010) and with CHN (non-elective hospitalisation/s, frailty or polypharmacy) were selected.

Self-controlled case series (SCCS) analyses were subsequently conducted among people who did not use the respective preventative treatment in the year before study start.

Incidence rate ratios (IRR) were calculated for myocardial infarction (MI) and stroke [antihypertensives, statins] and fractures [bisphosphonates] comparing event rates for the respective outcomes during treatment vs. post-discontinuation periods.

**Results:** 198,039 people were included to the CHN cohort. Among those, 6,245 individuals were included for the analysis of bisphosphonate discontinuation and fracture risk; 738 and 669 persons for the analysis of antihypertensive therapy discontinuation and MI/stroke risk; and 1,408 and 1,361 people for statin discontinuation and MI/stroke risk.

Risk of MI was substantially increased following discontinuation of antihypertensives (IRR 2.6 [95%CI 1.56-4.33]) and statins (IRR 1.75 [1.16-2.62]). No significant association for treatment discontinuation and stroke risk was seen. Likewise, no increased fracture risk was seen after discontinuing bisphosphonates. However, discontinuation among people with >1 year history of bisphosphonate therapy pointed towards increased fracture risk.

**Conclusions:** Our study showed risks associated with discontinuing preventative medications in people with CHN, likely explained by the continued efficacy of these medications. Further research focussing on the risk-benefit of these treatments for most vulnerable older adults is needed.

**Key points:**

- Self-controlled case series showed substantially increased risk for myocardial infarction following discontinuation of antihypertensives and statins.
- Fracture risk was not increased during treatment with bisphosphonates vs. post-discontinuation, but in people with >1 year of treatment a trend towards increased risk was seen.
- Future research is required to study further conditions, medications, and sequalae in older patients with complex health needs.

## Introduction

As life expectancy increases, a growing proportion of the UK population will be living with multimorbidity, defined as the presence of two or more long-term health conditions^1^. Studies have found that the overall rate of multimorbidity in England and Scotland is between 23-27%^2,3^. These figures rise dramatically with age, with between 55-74% of 65-84 year olds, and >80% of >85 years old, experiencing multimorbidity and are predicted to increase in the coming decades^4^.

Alongside rising multimorbidity comes cumulative prescription of medications to treat each individual condition. Rates of polypharmacy increased in the UK between 1995-2010, with the proportion prescribed >5 drugs doubling, and >10 drugs tripling^5^. Treatment decisions and efficacy are also impacted by frailty, which increases considerably with age^6^ and is estimated affect more than 40% of people aged ≥65 years^7^. Historically, guidelines have struggled to help clinicians navigate the management of these increasingly frequent patients with complex health needs^8^.

A set of medications frequently prescribed to patients with complex health needs are ‘preventative’ drugs such as statins, antihypertensives, and bisphosphonates^9^. A recent multimorbidity guideline published by the National Institute for Health and Care Excellence (NICE) recommended that further research is urgently required into the benefits or risks of stopping such medications, taking into account the significant potential burden of side effects these drugs may cause, and that this patient cohort is less likely to have been eligible or been included in previously published trials^10^.

In this study we aimed to address this research gap by conducting a self-controlled case series (SCCS) analysis^11^ to establish the effect of stopping statins, antihypertensives and bisphosphonates on the risk of cardiovascular events and fractures, in older patients with complex health needs.

## Methods

### Data source

We obtained pseudonymised electronic health records from the Clinical Practice Research Datalink (CPRD) GOLD, a primary care dataset covering >17 million people in the UK^12^. General practitioners act as gatekeepers to the UK healthcare system, and are therefore responsible for longitudinal care, including repeat prescriptions, and the recording of all conditions, health events and outcomes. CPRD GOLD includes data on socio-demographics, comorbidities, prescriptions, laboratory tests, clinical measurements, lifestyle factors, and referrals^13^. In addition, CPRD GOLD^14^ was linked to Hospital Episode Statistics (HES) inpatient care records, and to the Office for National Statistics mortality data.

### Study Design

We conducted separate SCCS analyses for each study outcome: fracture, myocardial infarction (MI), and stroke. SCCS is a case-only study design, where event rates during exposed and unexposed time windows are compared within a patient’s follow-up, including only those who experienced the event of interest^11,15,16^. As patients act as “controls” for themselves, time-fixed confounding is resolved by design^11^. To estimate the potential effect of the discontinuation of preventative medicines, we studied the risk for each of the study outcomes during exposed time (*treatment period*) and during the time immediately after therapy was stopped (*post-discontinuation period*) by dividing the latter over the former. The study design is illustrated in Supplement 1 Figure S1.

### Study population

The source population comprised all patients aged >65 years registered with an “up-to-standard” practice^13^ with HES linkage for ≥1 year before the start of study period (01/01/2010), and who remained registered for at least 1 day of follow-up. People who were exposed to any of the study drugs (statins, antihypertensives, or bisphosphonates) in the year before study start were excluded for each of the study-specific analyses.

Out of the source population, we identified patients with complex health needs as defined by 3 different indicators^9^: patients with unplanned hospitalisations (*hospitalisation* cohort); patients with an electronic frailty index deficit count^17^ of ≥ 3 (*frailty* cohort); and patients with ≥10 different medicines prescribed in the previous year (*polypharmacy* cohort). Patients included in any of these 3 cohorts were also included in a complex health needs (*CHN*) cohort. Only those with one of the outcomes of interest (cases) within the study period (01/01/2010 – end of follow-up) were included. A population flowchart is provided in Figure 1. For the SCCS, only people with exposure to the respective preventative treatment were analysed.

**Figure 1:**
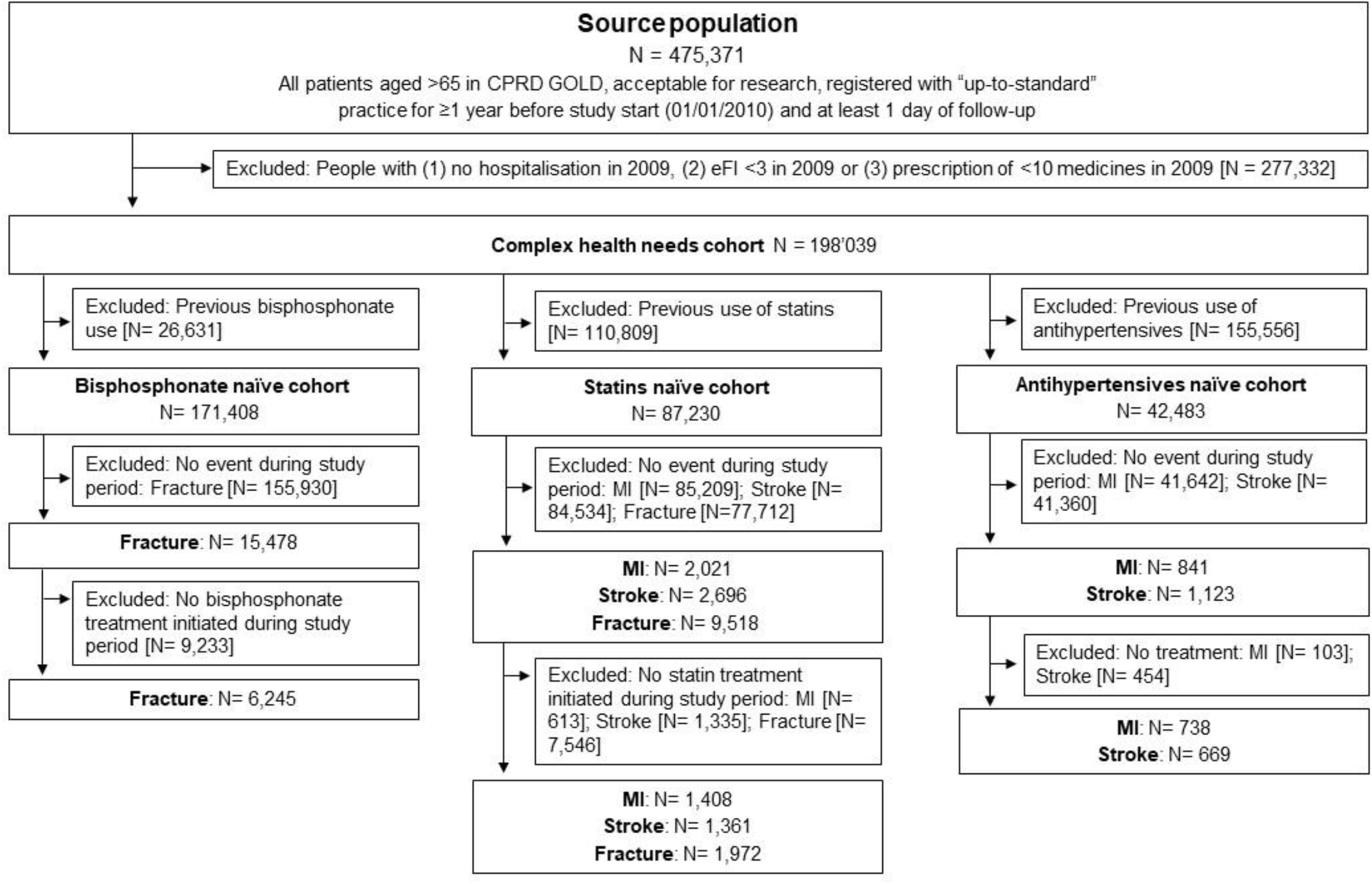
**Study inclusion flowchart** for the complex heath needs cohort (CHN). Among those people with events, only those who were treated with the respective preventative treatments were included for the respective SCCS analyses.

### Exposure and follow-up

Patients’ follow-up time was divided into two subsequent periods. First, a *treatment period* was defined, elapsing from the day after start of therapy initiation (first prescription after a 1-year washout) until date of therapy discontinuation. Treatment periods were defined using repeat prescriptions of a same drug group with a maximum refill gap of 90 days between them^18^. A grace period of 90 days was added to the end date to account for non-compliance and stockpiling. Secondly, a *post-discontinuation period* was defined, going from day 1 after therapy discontinuation and until the earliest of death, transfer out of the practice, or date of data extraction (13/09/2019).

The main analysis focussed on the observed event rates in the *post-discontinuation* compared to the *treatment period* to assess the impact of therapy discontinuation. The time from start of study period (01/01/2010) until the date of therapy initiation was excluded. The 30-day period before therapy initiation was used to test key assumptions of the SCCS method (see Statistical Analyses).

Three exposures were studied separately, based on primary care prescriptions (product-specific codes) [Supplement 2]. Antihypertensive drug use included angiotensin-converting-enzyme inhibitors (ACE-I), angiotensin-II receptor blockers (ARB), beta blockers (BB), calcium-channel blockers (CCB), diuretics, renin inhibitors and other antihypertensives, as well as their respective combinations. Statins included atorvastatin, simvastatin, fluvastatin, pravastatin, rosuvastatin, and cerivastatin. Finally, bisphosphonates included all oral bisphosphonates recorded in the study period: alendronate, risedronate, ibandronate, clodronate, etidronate, and tiludronate.

### Outcomes

Outcomes of interest were fractures, MI, and stroke, defined using previously published code lists/algorithms [Supplement 2]. Multiple recordings of the respective outcomes within 30 days were considered duplicates and removed from the dataset.

### Statistical analyses

SCCS analyses were conducted for each of the study outcomes and for each of the medicines of interest separately. Incidence rate ratios (IRR) were derived using conditional Poisson regression to compare event rates during the *post-discontinuation period* vs the *treatment period*. Therefore, an IRR>1 would imply an increased risk of a given health outcome in the time period following the discontinuation of the preventative treatment under study. Age was considered as a time-varying confounder and was adjusted for using 1-year bands. Event rates per 1,000 person-years and corresponding 95% confidence intervals were calculated with Poisson regression.

We conducted three sensitivity analyses. First, we restricted analyses of bisphosphonate discontinuation on fracture risk to subjects who used bisphosphonate/s for at least 1 year, as pivotal trial data demonstrate no fracture protection is achieved amongst those who use them for less than 1 year^19,20^. Secondly, we excluded patients with no follow-up time available after therapy discontinuation to test the impact of such a restriction. Lastly, follow-up time was *censored* for patients if they restarted the respective treatment after previous discontinuation to avoid misclassification of events in the post-discontinuation period in case of re-exposure to preventative treatments.

The association between statin use and fracture risk was studied as a negative exposure analysis to identify residual confounding.

We tested the key assumptions of the SCCS methods^11^ among people in the CHN cohort. Assumption (1), *the occurrence of an event should not affect subsequent exposures*, was tested graphically by investigating the distribution of events relative to treatment start and discontinuation date(s). For assumption (2), *events do not influence the length of observation periods*, follow-up duration was compared between people with/without events and stratified for exposure. Lastly, the relevance of assumption (3), *recurrences of an event should be independent or rare*, was assessed calculating the number of events per person.

All analyses were conducted using R version 4.1.3. SCCS analyses were conducted using the SCCS package^21^. Curator software^22^ was used to perform pre-analytical data curation.

## Results

Out of a source population of 475,371 people, 198,039 were identified as CHN based on their definition in the hospitalization (n= 90,597), frailty (n= 110,225) or polypharmacy (n= 116,076) sub-populations. Of these, a total of 6,245 were included for the analysis of bisphosphonate discontinuation and fracture risk. This number reduced to 3,091 when the analysis was restricted to subjects who used bisphosphonates for at least 1 year (sensitivity analysis). Similarly, 738 and 669 were studied for the analysis of antihypertensive therapy discontinuation, and MI and stroke risk respectively. Finally, 1,408 and 1,361 people were analysed for the study of statin discontinuation and MI and stroke respectively; a total of 1,972 patients were included for the negative exposure analysis based on the association between statin discontinuation and fracture risk. Figure 1 shows a population flowchart, with Supplementary Figures S2, S3, and S4 depicting similar information for the hospitalization, frailty and polypharmacy sub-populations.

Figure 2 shows the results from age-adjusted SCCS analyses. A total of 1,262 fractures were observed during 9,796 person-years of treatment with bisphosphonates, equivalent to an event rate of 128.8/1,000 person-years [95%CI 121.7-136.0]. This compared to 942 fractures during 6,428 person-years of follow-up post-discontinuation (event rate of 146.6/1,000 person-years [137.2-156.1]). The resulting age-adjusted IRR for the effect of bisphosphonate discontinuation on fracture risk was 0.94 [95%CI 0.81-1.1]. In the sensitivity analysis restricted to those who used bisphosphonates for at least 1 year, a total of 966 fractures were observed during 8,338 person-years of bisphosphonate therapy (event rate of 115.9/1,000 person-years [108.6-123.3]), compared to 280/1,887 person-years post-discontinuation (event rate of 148.4/1,000 person-years [131.0-166.3]). This equated to an age-adjusted IRR of 1.26 [1.0-1.58].

**Figure 2:**
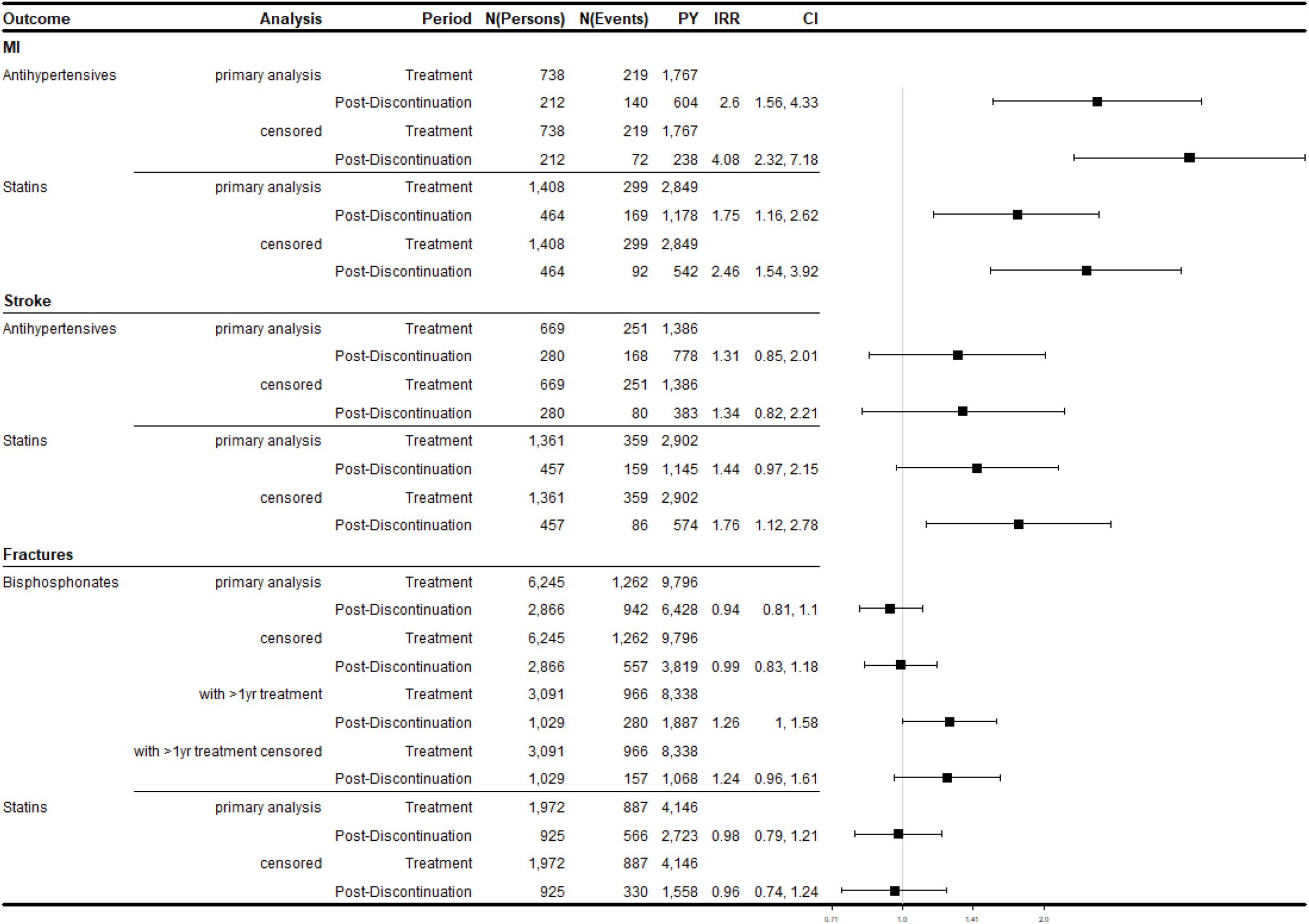
**Self-controlled case series** in the complex heath needs cohort (CHN). N(Persons) = Number of persons, N(Events) = Number of events, PY = patient years, IRR = Incidence rate ratio, CI = 95% confidence intervals, “censored” = people’s follow-up time was censored at the time of treatment restart after discontinuation.

Regarding antihypertensive therapy discontinuation, a total of 212 MI events were seen during 1,767 person-years of treatment (event rate 123.9/1,000 person-years [107.5-140.9]), compared to 140 MIs in 604 person-years of post-discontinuation observation time (event rate 231.8/1,000 person-years [193.5-271.7]). This resulted in an age-adjusted IRR of 2.6 [1.56-4.33]. Similarly, 251 stroke events were observed during 1,386 person-years of treatment (event rate 181.1/1,000 person-years [158.7-204.2]), compared to 168 strokes in 778 person-years of post-discontinuation time (event rate 215.9/1,000 person-years [183.3-249.8]), equivalent to an age-adjusted IRR of 1.31 [0.85-2.01] for the effect of antihypertensive therapy discontinuation on stroke risk.

Finally, a total of 299 MIs were seen during 2,849 person-years of statin treatment observation (event rate 105.0/1,000 person-years [93.06-117.2]), compared to 169 MIs in 1,178 person-years of post-discontinuation person-time (event rate 143.5/1,000 person-years [121.9-165.9]). This resulted in an age-adjusted IRR of 1.75 [1.16-2.62] for the association between statin discontinuation and MI risk. In line with this, 359 vs 159 stroke events were seen in 2,902 vs 1,145 person-years on statins vs post-discontinuation time, equivalent to event rates of 123.7/1,000 person-years [110.9-136.0] vs 138.9/1,000 person-years [117.3-161.3]. This led to an age-adjusted IRR of 1.44 [0.97-2.15] for the association between statin discontinuation and stroke risk. No association was observed between statin discontinuation and fracture risk, with fracture rates of 213.9/1,000 person-years [199.9-228.3] vs 211.8/1,000 person-years [189.0-235.3] during statin treatment vs post-discontinuation, and an age-adjusted IRR of 0.98 [0.79-1.21].

### Subgroups, Sensitivity analyses, and SCCS assumptions

All analyses were conducted separately for the hospitalization, frailty, and polypharmacy subpopulations [Supplementary Tables S5-S7]. The results were largely consistent across the different cohorts, with different point estimates for age-adjusted IRR but similar direction/s of association.

Sensitivity analyses censoring a person’s follow-up time at the time they restarted the respective treatment were consistent with the main results [Figure 2]. Additional sensitivity analyses only including people with at least one day of follow-up available after therapy discontinuation are reported in Figures S8-S11. The observed associations were overall in line with the main results above, with effect sizes slightly attenuated towards the null for MI, and no significant association for statin discontinuation and stroke risk.

Results from SCCS assumption test broadly indicated that the assumptions were not violated. Figures S12 and S13 in the Supplement illustrate the distribution of events around the date of treatment discontinuation, whereas Tables S1 highlights that the duration of follow-up was comparable between people with and without events. Recurrent events were rare, with 94% and 92% of people with MI, 83% of people with fractures and 91% of people with stroke having only one event during follow-up (Table S2).

## Discussion

### Statement of principal findings

We identified increased risks of severe adverse events following discontinuation of preventative treatments in patients with complex health needs, as defined by frailty, non-elective hospitalisation/s, and polypharmacy. Overall, the risks of MI increased following cessation of antihypertensive treatment or statins in our complex health needs group. Conversely, no consistent association of treatment discontinuation and stroke risk was consistently seen across our cohorts. Finally, the risk of fractures increased by around 25% after stopping bisphosphonates only in those who had taken these treatments for at least one year, although this was only significant among the frailty sub-group.

### Strengths and Limitations

Our study has several strengths. CPRD GOLD is frequently used for pharmacoepidemiological studies^23^ and provides reliable, good quality data for research^13^. Patients included in CPRD GOLD are broadly representative of the UK general population across key metrics such as age, sex, and ethnicity^24^. Our use of the SCCS method also provided a number of benefits compared to other epidemiological study designs^11^. This method allows transient exposures to be considered, which is important with long-term medications that may be halted, switched, or recommenced. Furthermore, given individuals act as their own controls, characteristics which remain constant over time, such as sex and ethnicity, are already accounted for. Lastly, we used negative exposure analysis to assess residual confounding.

Our study also had some limitations. As the population under study was made of older people with complex health needs, follow-up after treatment discontinuation was not available for a substantial proportion of the population (>50%). The reasons for stopping medications remain speculative and could not be evaluated in this study. For those for whom post-discontinuation follow-up was available, the main analysis did not consider potential restart of the treatment during the post-discontinuation period, potentially leading to exposure misclassification. We therefore conducted sensitivity analyses in which we censored follow-up at the time of restart of preventative treatment. Likewise, we did not consider changes in co-medication, e.g. medication for secondary prevention of cardiovascular events.

### Research in context

To our knowledge, this is the first study to use the SCCS method to investigate this question. However, previous studies have used alternative study designs to investigate the effects of discontinuing preventative medications in similar populations.

In 2021, a systematic review and meta-analysis of four randomised control trials investigated the effects of discontinuing bisphosphonates in patients aged >60 years, reporting an increased risk of vertebral fractures (HR 2.04, 95%CI 1.39-2.99)^25^. Our study also showed a increased risk of fractures among the frailty sub-group, although smaller in effect size.

A 2020 Cochrane review explored the effect of antihypertensive withdrawal in older people on rates of mortality and myocardial infarction, but could not provide any firm conclusions given the relatively small sample sizes and low or very low certainty of evidence provided by the six included randomised control trials^26^. Recently, the OPTIMISE trial investigated the effect of deprescribing antihypertensive medications among patients aged >80 years^27^. There was no difference in the rate of serious adverse events, including myocardial infarction and stroke, although this finding was limited by small event counts. In addition, the Opti-med study^28^ and TONE trial^29^ explored deprescribing antihypertensives among frail or older adults, but did not investigate rates of cardiovascular events. In contrast, our results provide evidence of significantly increased risks of stroke and MI following cessation of antihypertensive medications.

Several cohort studies have compared rates of cardiovascular events following either continuation or discontinuation of statins^30-32^. A study among patients >75 years in France reported increased risks of coronary (HR 1.46, 95%CI 1.21-1.75) and cerebrovascular (HR 1.26, 95% 1.05-1.51) events after discontinuation^32^. Similarly, increased risk for MI and stroke were found in a study from Denmark which compared the same age group by statin indication (primary or secondary prevention)^30^. While our results showed an increase in MI following discontinuation of statins, we did not see the same effect for stroke.

The studies discussed above included relatively older patients, who often had high rates of concurrent medication use and comorbidities. However, they did not restrict inclusion beyond age, making direct comparison with our formally defined complex health needs groups more difficult.

A further study investigated statin discontinuation among Italian adults >65 years old who specifically received ‘polypharmacy’, defined as use of statins with antihypertensives, antidiabetics and antiplatelet agents before the follow-up period^31^. No increased risks for hospitalisation with cerebrovascular disease (HR 1.15, 95% CI 0.95-1.38) or ischaemic heart disease (HR 1.08, 95%CI 0.94-1.23) was reported, although an increased risk of a composite cardiovascular outcome score was seen. Our study, however, did not find increased risks of stroke and MI following discontinuation of statins in our differently defined polypharmacy cohort.

Our study highlights the risks associated with discontinuing preventative medications, which are likely explained by the continued efficacy of these medications in groups with complex health needs. Recent large-scale studies have shown that pharmacological management of blood pressure is effective well into old age^33^. Most studies to date have investigated preventative medication discontinuation in older age groups, rather than explicitly in groups with complex health needs. Whilst there is an important association between age and complex health needs, older age groups are not homogenous, and modern clinical practitioners and policy-makers will increasingly need to take into account other key factors, such as the complex health needs we explore in this study, when making individual treatment decisions^34,35^.

The current lack of evidence surrounding medication discontinuation in these groups has led to unclear, equivocal, or contradictory guidelines. For example, a systematic review of statin guidelines found that several guidelines recommend consideration of discontinuation in people with complex health needs, whilst others did not^36^. Other guidelines, such as NICE’s multimorbidity guideline^10^, instead recommend further research to help shape future guideline development. Our study provides novel insights which could contribute to improved care strategies in these groups in the future.

### Unanswered questions and future research

Our study focusses on key preventative medications and outcomes, but future research will allow further conditions, medications and sequalae to be studied in these key patient populations. Exploration of other facets of medication discontinuation, such as economic analyses, will help to build a fuller picture of the cost-benefits of treatment strategies.

## Supporting information

Supplemental File 2 - Code Lists

## Data Availability

Data were obtained from CPRD under the Oxford University CPRD license. Direct data
sharing is not allowed. Data access can be obtained from CPRD, conditional on approval
through CPRD s Research Data Governance Process.

## Notes

### Contributions

AJ, TRM and DPA designed the study. AD extracted and prepared the dataset. AJ analysed the data. EHT and TRM provided statistical advice. AJ, DPA and FD interpreted the results. AJ, DPA and FD drafted the manuscript. All authors critically reviewed and approved the final version of the manuscript.

### Ethical Approval

The study protocol was approved by the Independent Scientific Advisory Committee for MHRA Database Research (RDG), including amendments (Protocol No 19_132A2). It will be made available to the journal reviewers upon request.

### Data sharing

Data were obtained from CPRD under the Oxford University CPRD license. Direct data sharing is not allowed. Data access can be obtained from CPRD, conditional on approval through CPRD’s Research Data Governance Process.

## Acknowledgement

This study is based in part on data from the Clinical Practice Research Datalink obtained under license from the UK Medicines and Healthcare products Regulatory Agency. The data is provided by patients and collected by the NHS as part of their care and support. The interpretation and conclusions contained in this study are those of the authors alone.

