## Supplemental File 1 - Figures for "Cardiovascular outcomes and fracture risk after the discontinuation of preventative medications in older patients with complex health needs: a self-controlled case series analysis"

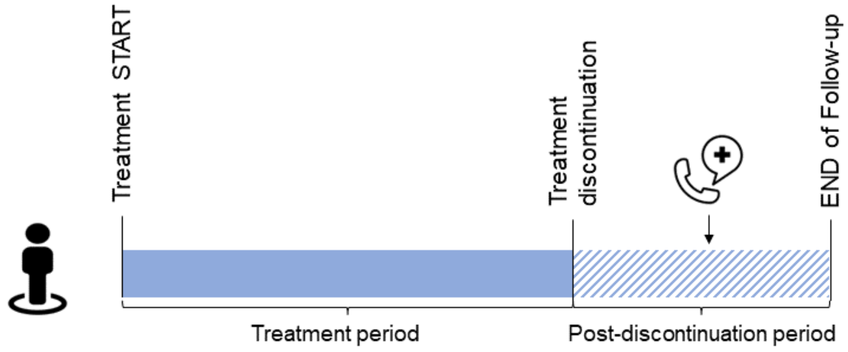


**Figure S1: Self-controlled case series design** Within each patient's follow-up, event rates during exposed (*treatment period*) and unexposed (*post-discontinuation*) time windows are compared.

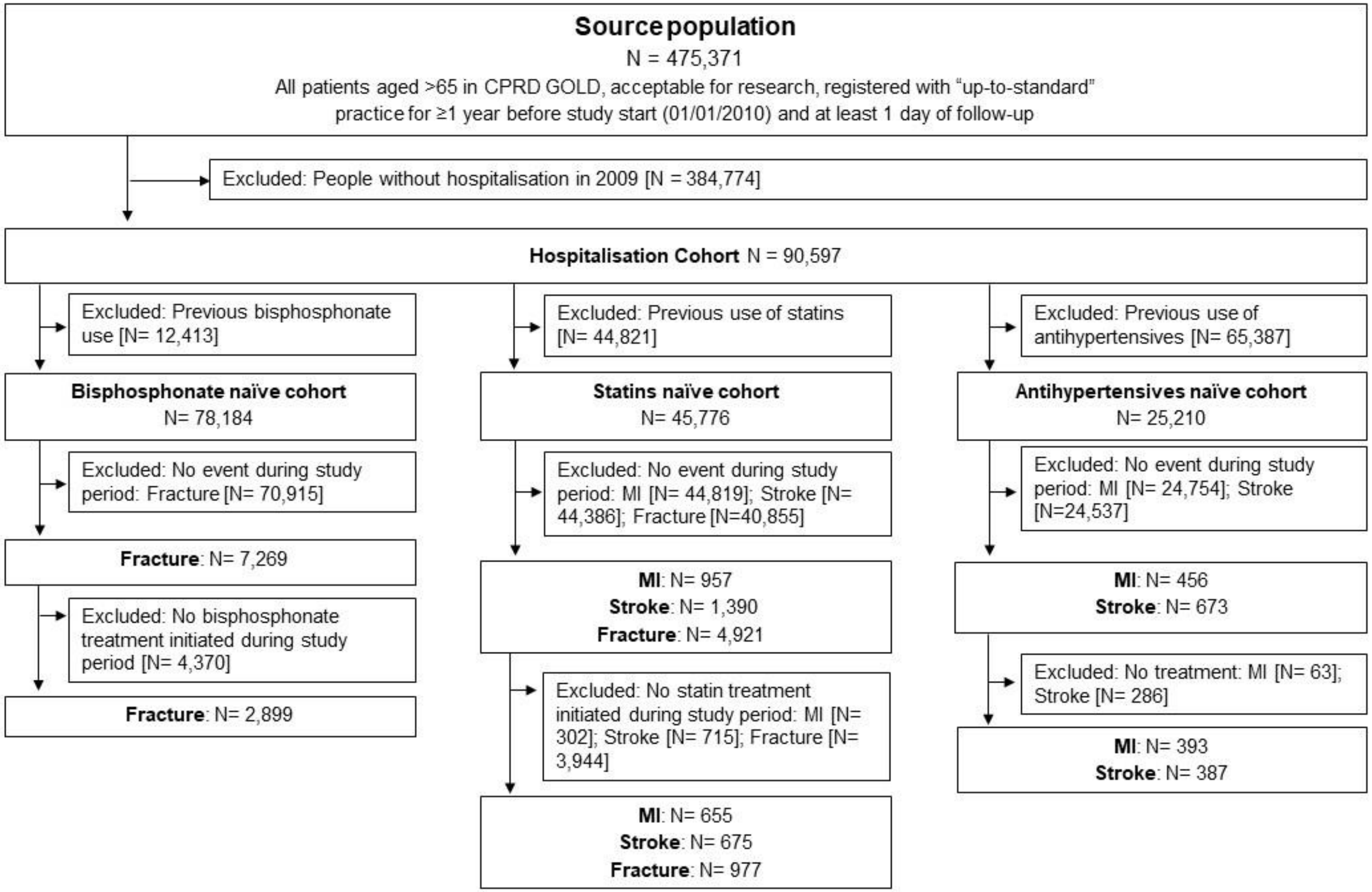


**Figure S2: Study inclusion flowchart** for the hospitalisation cohort

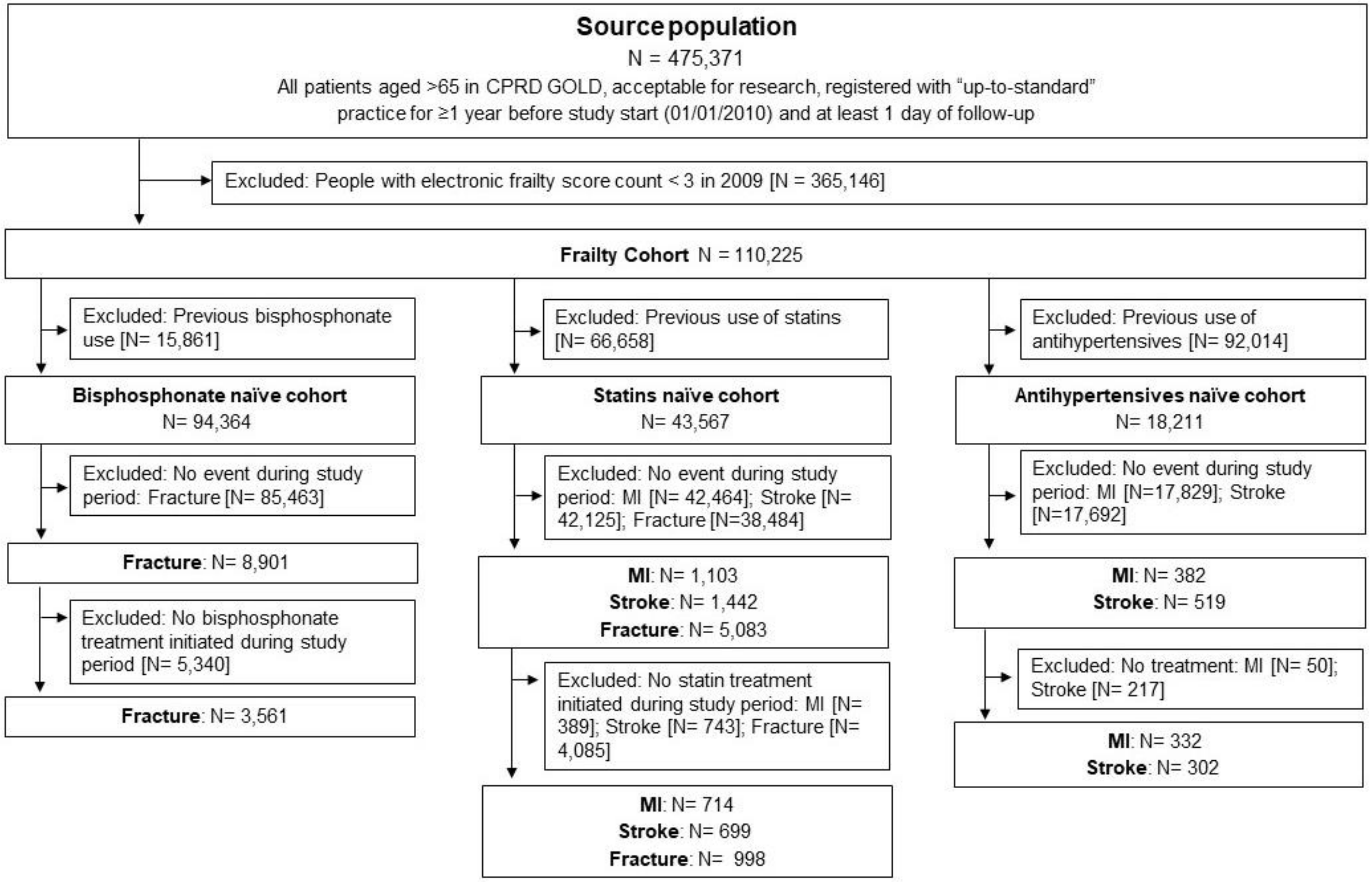


**Figure S3: Study inclusion flowchart** for the frailty cohort

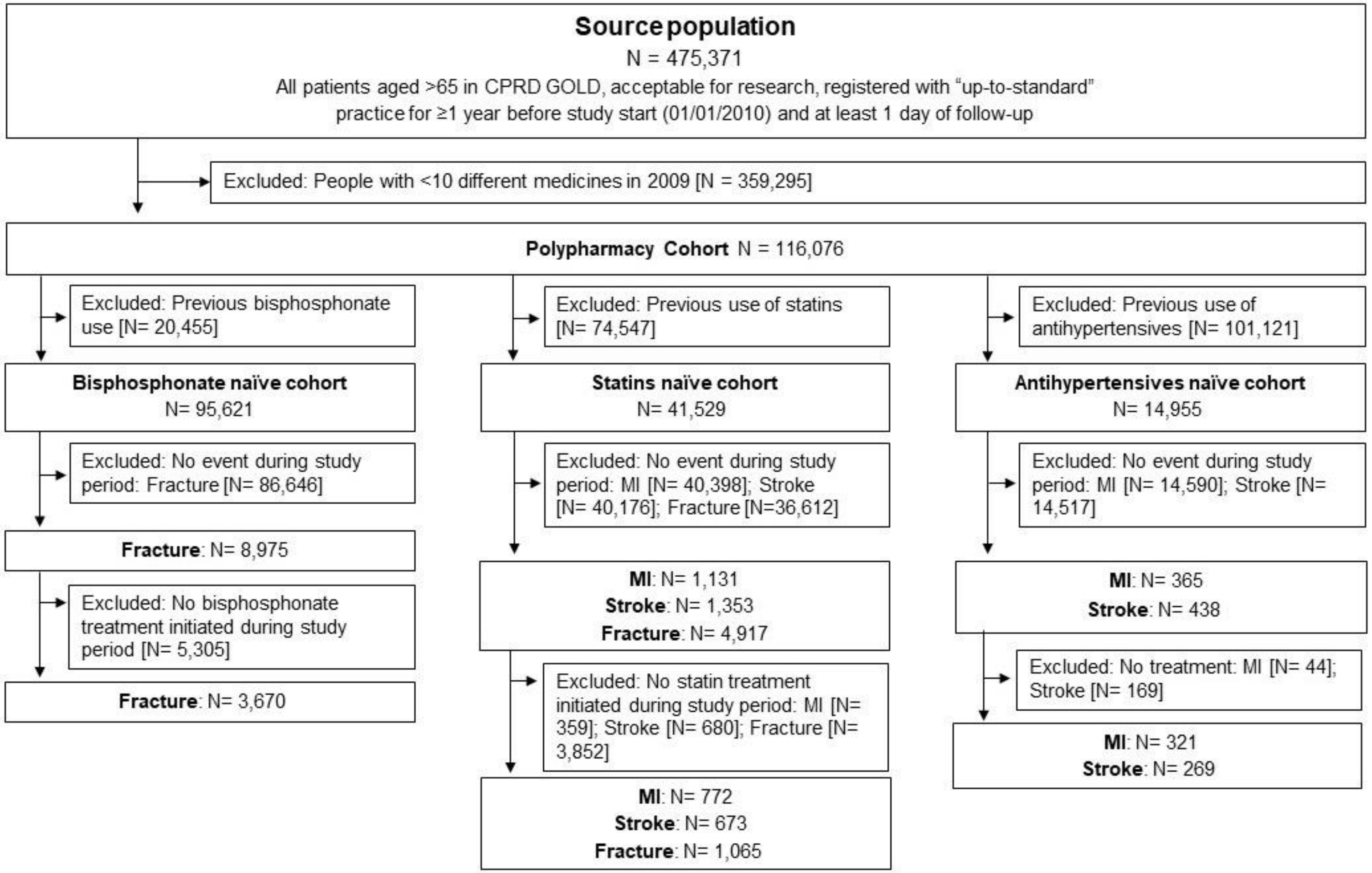


**Figure S4: Study inclusion flowchart** for the polypharmacy cohort

| Outcome | Analysis | Period | N(Persons) | N(Events) | PY | IRR | CI |
|---|---|---|---|---|---|---|---|
| **MI** | | | | | | | |
| Antihypertensives | primary analysis | Treatment | 393 | 122 | 941 | | |
| | | Post-Discontinuation | 106 | 65 | 307 | 1.86 | 0.93, 3.73 |
| | censored | Treatment | 393 | 122 | 941 | | |
| | | Post-Discontinuation | 106 | 32 | 132 | 3.41 | 1.6, 7.25 |
| Statins | primary analysis | Treatment | 655 | 141 | 1,300 | | |
| | | Post-Discontinuation | 213 | 71 | 505 | 1.28 | 0.71, 2.3 |
| | censored | Treatment | 655 | 141 | 1,300 | | |
| | | Post-Discontinuation | 213 | 36 | 236 | 1.77 | 0.91, 3.43 |
| **Stroke** | | | | | | | |
| Antihypertensives | primary analysis | Treatment | 387 | 146 | 807 | | |
| | | Post-Discontinuation | 148 | 84 | 410 | 1.12 | 0.62, 2.02 |
| | censored | Treatment | 387 | 146 | 807 | | |
| | | Post-Discontinuation | 148 | 52 | 209 | 1.47 | 0.76, 2.84 |
| Statins | primary analysis | Treatment | 675 | 166 | 1,414 | | |
| | | Post-Discontinuation | 215 | 64 | 462 | 1.88 | 1.04, 3.38 |
| | censored | Treatment | 675 | 166 | 1,414 | | |
| | | Post-Discontinuation | 215 | 36 | 234 | 2.12 | 1.08, 4.15 |
| **Fractures** | | | | | | | |
| Bisphosphonates | primary analysis | Treatment | 2,899 | 624 | 4,545 | | |
| | | Post-Discontinuation | 1,331 | 466 | 3,039 | 1.01 | 0.8, 1.26 |
| | censored | Treatment | 2,899 | 624 | 4,545 | | |
| | | Post-Discontinuation | 1,331 | 278 | 1,819 | 1.06 | 0.83, 1.36 |
| | with >1yr treatment | Treatment | 1,396 | 486 | 3,858 | | |
| | | Post-Discontinuation | 468 | 136 | 898 | 1.13 | 0.82, 1.56 |
| | with >1yr treatment censored | Treatment | 1,396 | 486 | 3,858 | | |
| | | Post-Discontinuation | 468 | 80 | 512 | 1.16 | 0.81, 1.66 |
| Statins | primary analysis | Treatment | 977 | 431 | 2,040 | | |
| | | Post-Discontinuation | 444 | 263 | 1,225 | 1.03 | 0.76, 1.41 |
| | censored | Treatment | 977 | 431 | 2,040 | | |
| | | Post-Discontinuation | 444 | 155 | 673 | 1.05 | 0.73, 1.52 |

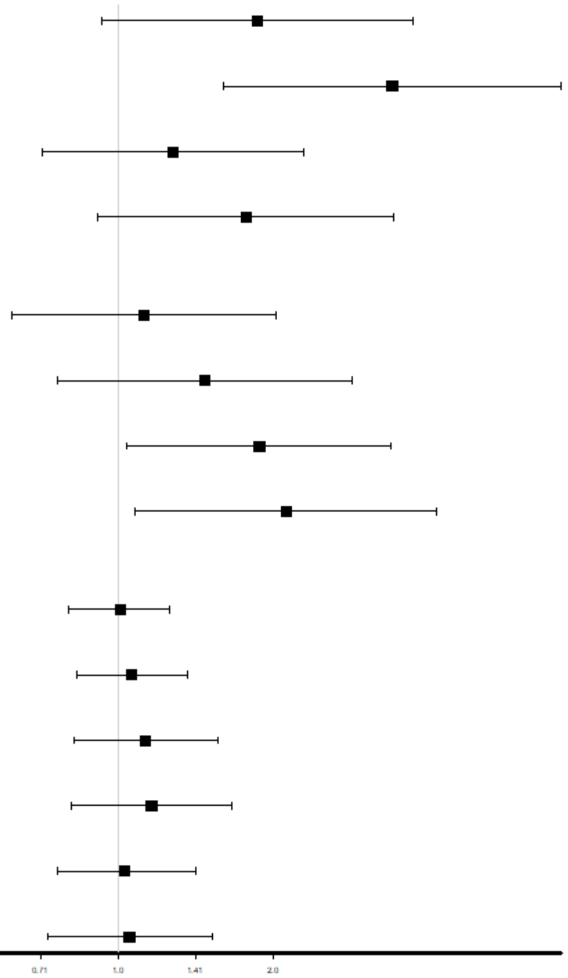

**Figure S5: Self-controlled case series** in the hospitalisation cohort.
N(Persons) = Number of persons, N(Events) = Number of events, PY = patient years, IRR = Incidence rate ratio, CI = 95% confidence intervals

| Outcome | Analysis | Period | N(Persons) | N(Events) | PY | IRR | CI |
|---|---|---|---|---|---|---|---|
| **MI** | | | | | | | |
| Antihypertensives | primary analysis | Treatment | 332 | 95 | 719 | | |
| | | Post-Discontinuation | 102 | 75 | 301 | 5.21 | 2.11, 12.85 |
| | censored | Treatment | 332 | 95 | 719 | | |
| | | Post-Discontinuation | 102 | 35 | 108 | 11.34 | 3.71, 34.65 |
| Statins | primary analysis | Treatment | 714 | 156 | 1,370 | | |
| | | Post-Discontinuation | 239 | 102 | 556 | 2.14 | 1.18, 3.88 |
| | censored | Treatment | 714 | 156 | 1,370 | | |
| | | Post-Discontinuation | 239 | 55 | 254 | 3.66 | 1.85, 7.23 |
| **Stroke** | | | | | | | |
| Antihypertensives | primary analysis | Treatment | 302 | 107 | 580 | | |
| | | Post-Discontinuation | 141 | 93 | 371 | 2.43 | 1.2, 4.92 |
| | censored | Treatment | 302 | 107 | 580 | | |
| | | Post-Discontinuation | 141 | 41 | 177 | 2 | 0.92, 4.36 |
| Statins | primary analysis | Treatment | 699 | 193 | 1,426 | | |
| | | Post-Discontinuation | 237 | 82 | 604 | 1.19 | 0.68, 2.05 |
| | censored | Treatment | 699 | 193 | 1,426 | | |
| | | Post-Discontinuation | 237 | 40 | 326 | 1.18 | 0.62, 2.26 |
| **Fractures** | | | | | | | |
| Bisphosphonates | primary analysis | Treatment | 3,561 | 737 | 5,425 | | |
| | | Post-Discontinuation | 1,597 | 549 | 3,558 | 1 | 0.81, 1.22 |
| | censored | Treatment | 3,561 | 737 | 5,425 | | |
| | | Post-Discontinuation | 1,597 | 327 | 2,086 | 1.09 | 0.87, 1.38 |
| | with >1yr treatment | Treatment | 1,735 | 563 | 4,575 | | |
| | | Post-Discontinuation | 560 | 154 | 1,014 | 1.4 | 1.03, 1.91 |
| | with >1yr treatment censored | Treatment | 1,735 | 563 | 4,575 | | |
| | | Post-Discontinuation | 560 | 90 | 581 | 1.46 | 1.03, 2.06 |
| Statins | primary analysis | Treatment | 998 | 455 | 2,025 | | |
| | | Post-Discontinuation | 469 | 299 | 1,362 | 0.94 | 0.7, 1.27 |
| | censored | Treatment | 998 | 455 | 2,025 | | |
| | | Post-Discontinuation | 469 | 176 | 781 | 1.01 | 0.7, 1.46 |

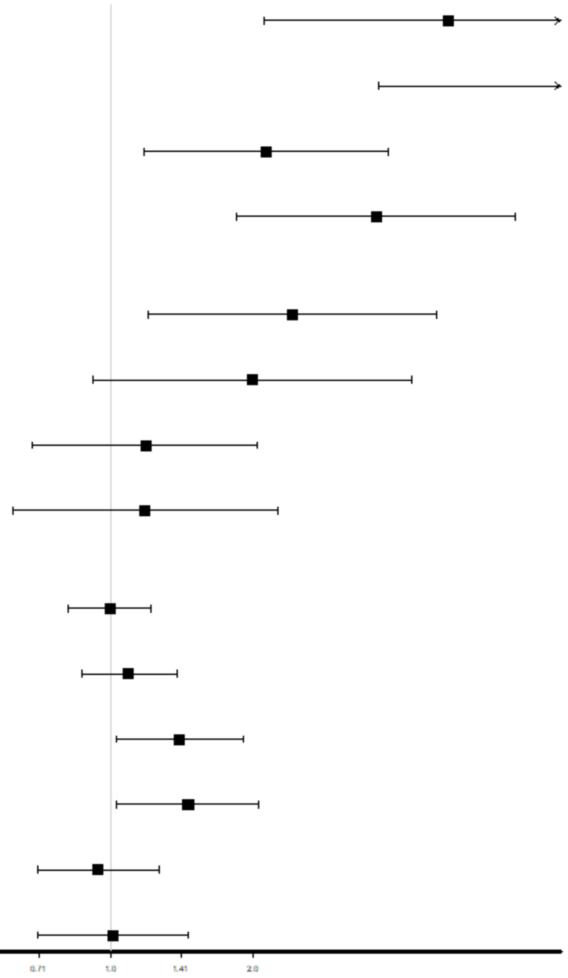


**Figure S6: Self-controlled case series** in the frailty cohort.
N(Persons) = Number of persons, N(Events) = Number of events, PY = patient years, IRR = Incidence rate ratio, CI = 95% confidence intervals

| Outcome | Analysis | Period | N(Persons) | N(Events) | PY | IRR | CI |
|---|---|---|---|---|---|---|---|
| **MI** | | | | | | | |
| Antihypertensives | primary analysis | Treatment | 321 | 88 | 757 | | |
| | | Post-Discontinuation | 107 | 81 | 332 | 2.78 | 1.34, 5.79 |
| | censored | Treatment | 321 | 88 | 757 | | |
| | | Post-Discontinuation | 107 | 36 | 125 | 3.9 | 1.68, 9.06 |
| Statins | primary analysis | Treatment | 772 | 167 | 1,523 | | |
| | | Post-Discontinuation | 263 | 94 | 678 | 1.25 | 0.75, 2.07 |
| | censored | Treatment | 772 | 167 | 1,523 | | |
| | | Post-Discontinuation | 263 | 51 | 319 | 1.56 | 0.88, 2.78 |
| **Stroke** | | | | | | | |
| Antihypertensives | primary analysis | Treatment | 269 | 105 | 546 | | |
| | | Post-Discontinuation | 127 | 81 | 334 | 1.17 | 0.61, 2.25 |
| | censored | Treatment | 269 | 105 | 546 | | |
| | | Post-Discontinuation | 127 | 33 | 172 | 1.22 | 0.56, 2.65 |
| Statins | primary analysis | Treatment | 673 | 186 | 1,427 | | |
| | | Post-Discontinuation | 230 | 79 | 584 | 1.03 | 0.58, 1.82 |
| | censored | Treatment | 673 | 186 | 1,427 | | |
| | | Post-Discontinuation | 230 | 43 | 297 | 1.52 | 0.78, 2.95 |
| **Fractures** | | | | | | | |
| Bisphosphonates | primary analysis | Treatment | 3,670 | 767 | 5,699 | | |
| | | Post-Discontinuation | 1,747 | 624 | 3,992 | 0.98 | 0.81, 1.2 |
| | censored | Treatment | 3,670 | 767 | 5,699 | | |
| | | Post-Discontinuation | 1,747 | 366 | 2,333 | 1.01 | 0.81, 1.26 |
| | with >1yr treatment | Treatment | 1,821 | 575 | 4,849 | | |
| | | Post-Discontinuation | 649 | 190 | 1,241 | 1.33 | 0.99, 1.78 |
| | with >1yr treatment censored | Treatment | 1,821 | 575 | 4,849 | | |
| | | Post-Discontinuation | 649 | 110 | 708 | 1.32 | 0.95, 1.83 |
| Statins | primary analysis | Treatment | 1,065 | 487 | 2,177 | | |
| | | Post-Discontinuation | 518 | 312 | 1,534 | 0.9 | 0.68, 1.19 |
| | censored | Treatment | 1,065 | 487 | 2,177 | | |
| | | Post-Discontinuation | 518 | 175 | 872 | 0.87 | 0.62, 1.22 |

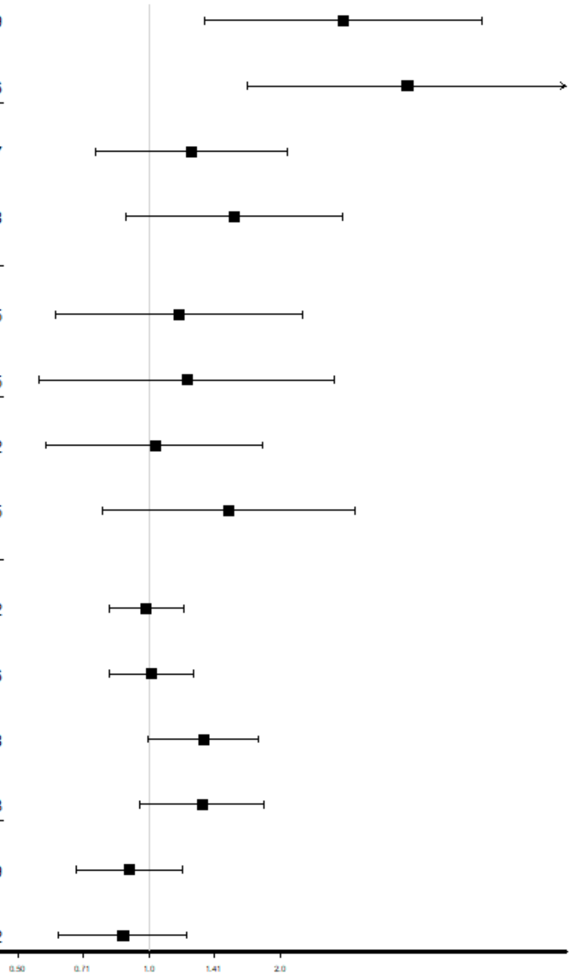


**Figure S7: Self-controlled case series** in the polypharmacy cohort.
N(Persons) = Number of persons, N(Events) = Number of events, PY = patient years, IRR = Incidence rate ratio, CI = 95% confidence intervals

| Outcome | Analysis | Period | N(Persons) | N(Events) | PY | IRR | CI |
|---|---|---|---|---|---|---|---|
| **MI** | | | | | | | |
| Antihypertensives | with discontinuation | Treatment | 212 | 34 | 260 | | |
| | | Post-Discontinuation | 212 | 140 | 604 | 2.11 | 1.22, 3.64 |
| | with discontinuation, censored | Treatment | 212 | 34 | 260 | | |
| | | Post-Discontinuation | 212 | 72 | 238 | 2.35 | 1.28, 4.33 |
| Statins | with discontinuation | Treatment | 464 | 53 | 590 | | |
| | | Post-Discontinuation | 464 | 169 | 1,178 | 1.55 | 1, 2.38 |
| | with discontinuation, censored | Treatment | 464 | 53 | 590 | | |
| | | Post-Discontinuation | 464 | 92 | 542 | 1.73 | 1.05, 2.86 |
| **Stroke** | | | | | | | |
| Antihypertensives | with discontinuation | Treatment | 280 | 48 | 300 | | |
| | | Post-Discontinuation | 280 | 168 | 778 | 1.37 | 0.86, 2.17 |
| | with discontinuation, censored | Treatment | 280 | 48 | 300 | | |
| | | Post-Discontinuation | 280 | 80 | 383 | 1.27 | 0.73, 2.21 |
| Statins | with discontinuation | Treatment | 457 | 65 | 656 | | |
| | | Post-Discontinuation | 457 | 159 | 1,145 | 1.3 | 0.84, 2 |
| | with discontinuation, censored | Treatment | 457 | 65 | 656 | | |
| | | Post-Discontinuation | 457 | 86 | 574 | 1.31 | 0.79, 2.17 |
| **Fractures** | | | | | | | |
| Bisphosphonates | with discontinuation | Treatment | 2,866 | 463 | 3,448 | | |
| | | Post-Discontinuation | 2,866 | 942 | 6,428 | 0.87 | 0.74, 1.03 |
| | with discontinuation, censored | Treatment | 2,866 | 463 | 3,448 | | |
| | | Post-Discontinuation | 2,866 | 557 | 3,819 | 0.84 | 0.7, 1.02 |
| Statins | with discontinuation | Treatment | 925 | 230 | 1,153 | | |
| | | Post-Discontinuation | 925 | 566 | 2,723 | 0.92 | 0.73, 1.16 |
| | with discontinuation, censored | Treatment | 925 | 230 | 1,153 | | |
| | | Post-Discontinuation | 925 | 330 | 1,558 | 0.86 | 0.65, 1.14 |

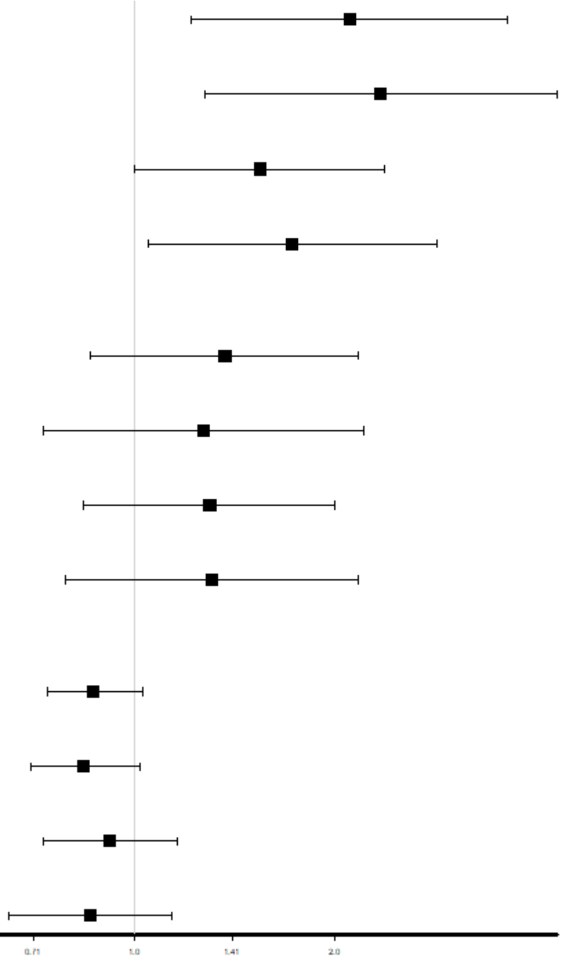

**Figure S8: Self-controlled case series** in the complex health needs cohort (CHN).
N(Persons) = Number of persons, N(Events) = Number of events, PY = patient years, IRR = Incidence rate ratio, CI = 95% confidence intervals

| Outcome | Analysis | Period | N(Persons) | N(Events) | PY | IRR | CI |
|---|---|---|---|---|---|---|---|
| **MI** | | | | | | | |
| Antihypertensives | with discontinuation | Treatment | 106 | 18 | 131 | | |
| | | Post-Discontinuation | 106 | 65 | 307 | 1.41 | 0.66, 3.03 |
| | with discontinuation, censored | Treatment | 106 | 18 | 131 | | |
| | | Post-Discontinuation | 106 | 32 | 132 | 2.24 | 0.93, 5.43 |
| Statins | with discontinuation | Treatment | 213 | 29 | 285 | | |
| | | Post-Discontinuation | 213 | 71 | 505 | 1.24 | 0.66, 2.33 |
| | with discontinuation, censored | Treatment | 213 | 29 | 285 | | |
| | | Post-Discontinuation | 213 | 36 | 236 | 1.49 | 0.72, 3.11 |
| **Stroke** | | | | | | | |
| Antihypertensives | with discontinuation | Treatment | 148 | 26 | 151 | | |
| | | Post-Discontinuation | 148 | 84 | 410 | 1.24 | 0.65, 2.39 |
| | with discontinuation, censored | Treatment | 148 | 26 | 151 | | |
| | | Post-Discontinuation | 148 | 52 | 209 | 1.32 | 0.63, 2.77 |
| Statins | with discontinuation | Treatment | 215 | 33 | 319 | | |
| | | Post-Discontinuation | 215 | 64 | 462 | 1.65 | 0.85, 3.2 |
| | with discontinuation, censored | Treatment | 215 | 33 | 319 | | |
| | | Post-Discontinuation | 215 | 36 | 234 | 1.53 | 0.73, 3.23 |
| **Fractures** | | | | | | | |
| Bisphosphonates | with discontinuation | Treatment | 1,331 | 232 | 1,603 | | |
| | | Post-Discontinuation | 1,331 | 466 | 3,039 | 0.92 | 0.73, 1.17 |
| | with discontinuation, censored | Treatment | 1,331 | 232 | 1,603 | | |
| | | Post-Discontinuation | 1,331 | 278 | 1,819 | 0.89 | 0.68, 1.16 |
| Statins | with discontinuation | Treatment | 444 | 109 | 547 | | |
| | | Post-Discontinuation | 444 | 263 | 1,225 | 0.99 | 0.71, 1.4 |
| | with discontinuation, censored | Treatment | 444 | 109 | 547 | | |
| | | Post-Discontinuation | 444 | 155 | 673 | 1.04 | 0.69, 1.57 |

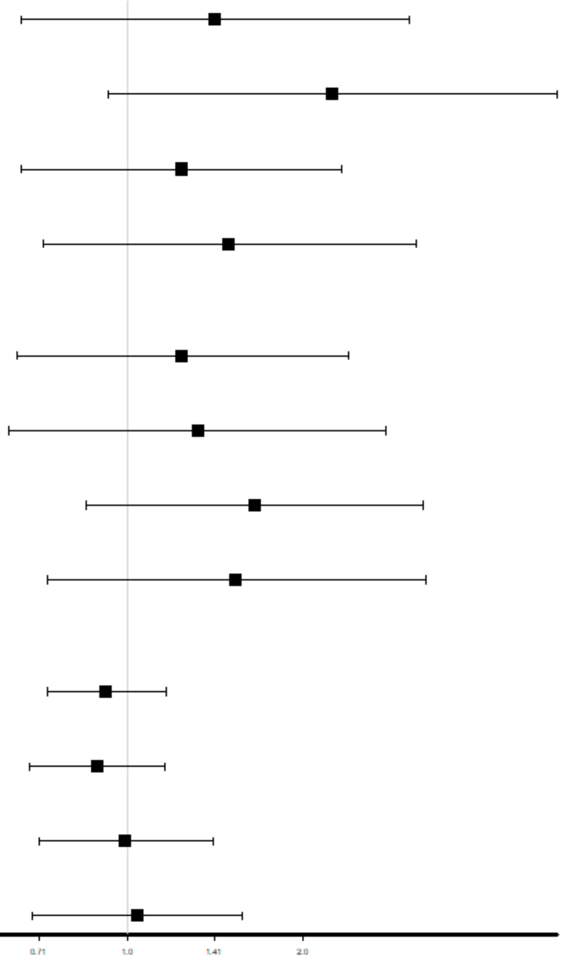

**Figure S9: Self-controlled case series** in the hospitalisation cohort.
N(Persons) = Number of persons, N(Events) = Number of events, PY = patient years, IRR = Incidence rate ratio, CI = 95% confidence intervals

| Outcome | Analysis | Period | N(Persons) | N(Events) | PY | IRR | CI |
|---|---|---|---|---|---|---|---|
| **MI** | | | | | | | |
| Antihypertensives | with discontinuation | Treatment | 102 | 10 | 107 | | |
| | | Post-Discontinuation | 102 | 75 | 301 | 5.17 | 1.9, 14.03 |
| | with discontinuation, censored | Treatment | 102 | 10 | 107 | | |
| | | Post-Discontinuation | 102 | 35 | 108 | 7.89 | 2.56, 24.28 |
| Statins | with discontinuation | Treatment | 239 | 22 | 290 | | |
| | | Post-Discontinuation | 239 | 102 | 556 | 1.87 | 1, 3.5 |
| | with discontinuation, censored | Treatment | 239 | 22 | 290 | | |
| | | Post-Discontinuation | 239 | 55 | 254 | 2.6 | 1.23, 5.52 |
| **Stroke** | | | | | | | |
| Antihypertensives | with discontinuation | Treatment | 141 | 16 | 133 | | |
| | | Post-Discontinuation | 141 | 93 | 371 | 2.43 | 1.16, 5.09 |
| | with discontinuation, censored | Treatment | 141 | 16 | 133 | | |
| | | Post-Discontinuation | 141 | 41 | 177 | 1.58 | 0.66, 3.77 |
| Statins | with discontinuation | Treatment | 237 | 38 | 337 | | |
| | | Post-Discontinuation | 237 | 82 | 604 | 0.93 | 0.5, 1.71 |
| | with discontinuation, censored | Treatment | 237 | 38 | 337 | | |
| | | Post-Discontinuation | 237 | 40 | 326 | 0.88 | 0.42, 1.85 |
| **Fractures** | | | | | | | |
| Bisphosphonates | with discontinuation | Treatment | 1,597 | 258 | 1,852 | | |
| | | Post-Discontinuation | 1,597 | 549 | 3,558 | 0.91 | 0.73, 1.13 |
| | with discontinuation, censored | Treatment | 1,597 | 258 | 1,852 | | |
| | | Post-Discontinuation | 1,597 | 327 | 2,086 | 0.91 | 0.71, 1.17 |
| Statins | with discontinuation | Treatment | 469 | 117 | 589 | | |
| | | Post-Discontinuation | 469 | 299 | 1,362 | 0.86 | 0.62, 1.19 |
| | with discontinuation, censored | Treatment | 469 | 117 | 589 | | |
| | | Post-Discontinuation | 469 | 176 | 781 | 0.83 | 0.55, 1.24 |

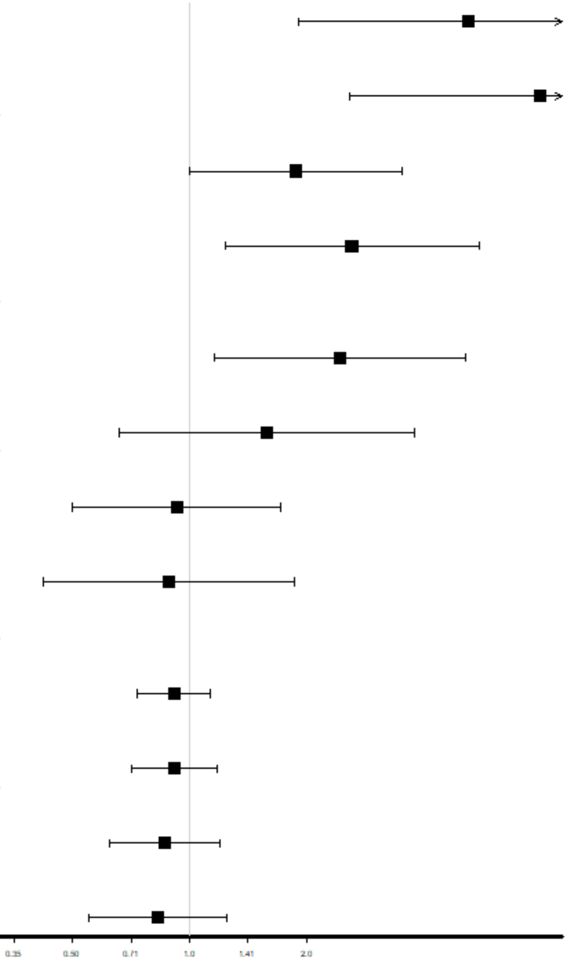

**Figure S10: Self-controlled case series** in the frailty cohort.
N(Persons) = Number of persons, N(Events) = Number of events, PY = patient years, IRR = Incidence rate ratio, CI = 95% confidence intervals

| Outcome | Analysis | Period | N(Persons) | N(Events) | PY | IRR | CI |
| --- | --- | --- | --- | --- | --- | --- | --- |
| **MI** | | | | | | | |
| Antihypertensives | with discontinuation | Treatment | 107 | 16 | 128 | | |
| | | Post-Discontinuation | 107 | 81 | 332 | 2.63 | 1.2, 5.77 |
| | with discontinuation, censored | Treatment | 107 | 16 | 128 | | |
| | | Post-Discontinuation | 107 | 36 | 125 | 3.17 | 1.25, 8.04 |
| Statins | with discontinuation | Treatment | 263 | 37 | 322 | | |
| | | Post-Discontinuation | 263 | 94 | 678 | 1.18 | 0.7, 2.01 |
| | with discontinuation, censored | Treatment | 263 | 37 | 322 | | |
| | | Post-Discontinuation | 263 | 51 | 319 | 1.29 | 0.68, 2.42 |
| **Stroke** | | | | | | | |
| Antihypertensives | with discontinuation | Treatment | 127 | 22 | 139 | | |
| | | Post-Discontinuation | 127 | 81 | 334 | 1.19 | 0.59, 2.4 |
| | with discontinuation, censored | Treatment | 127 | 22 | 139 | | |
| | | Post-Discontinuation | 127 | 33 | 172 | 1.02 | 0.43, 2.44 |
| Statins | with discontinuation | Treatment | 230 | 32 | 337 | | |
| | | Post-Discontinuation | 230 | 79 | 584 | 0.97 | 0.52, 1.82 |
| | with discontinuation, censored | Treatment | 230 | 32 | 337 | | |
| | | Post-Discontinuation | 230 | 43 | 297 | 1.16 | 0.55, 2.47 |
| **Fractures** | | | | | | | |
| Bisphosphonates | with discontinuation | Treatment | 1,747 | 280 | 2,110 | | |
| | | Post-Discontinuation | 1,747 | 624 | 3,992 | 0.91 | 0.74, 1.12 |
| | with discontinuation, censored | Treatment | 1,747 | 280 | 2,110 | | |
| | | Post-Discontinuation | 1,747 | 366 | 2,333 | 0.83 | 0.65, 1.05 |
| Statins | with discontinuation | Treatment | 518 | 133 | 625 | | |
| | | Post-Discontinuation | 518 | 312 | 1,534 | 0.9 | 0.66, 1.22 |
| | with discontinuation, censored | Treatment | 518 | 133 | 625 | | |
| | | Post-Discontinuation | 518 | 175 | 872 | 0.8 | 0.55, 1.17 |

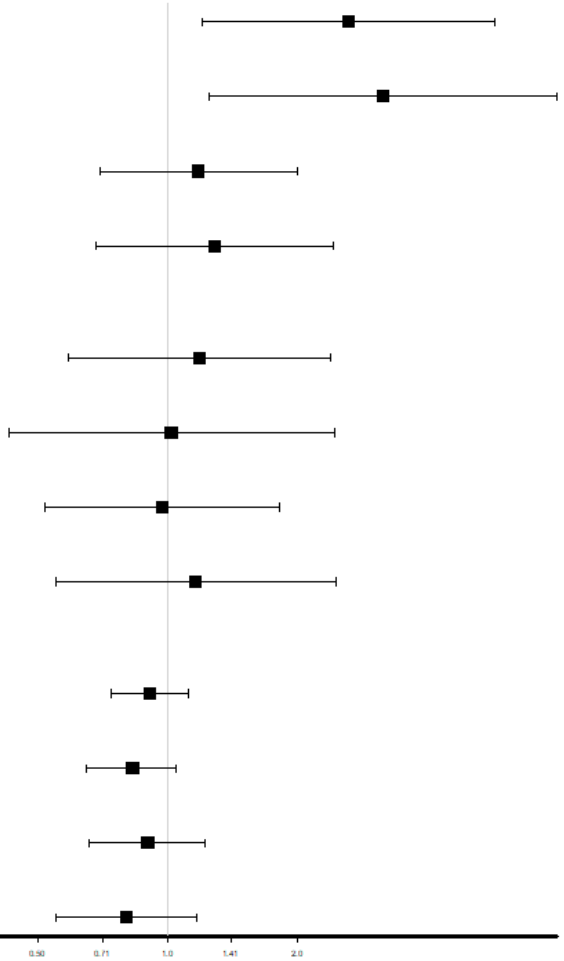


**Figure S11: Self-controlled case series** in the polypharmacy cohort.
N(Persons) = Number of persons, N(Events) = Number of events, PY = patient years, IRR = Incidence rate ratio, CI = 95% confidence intervals

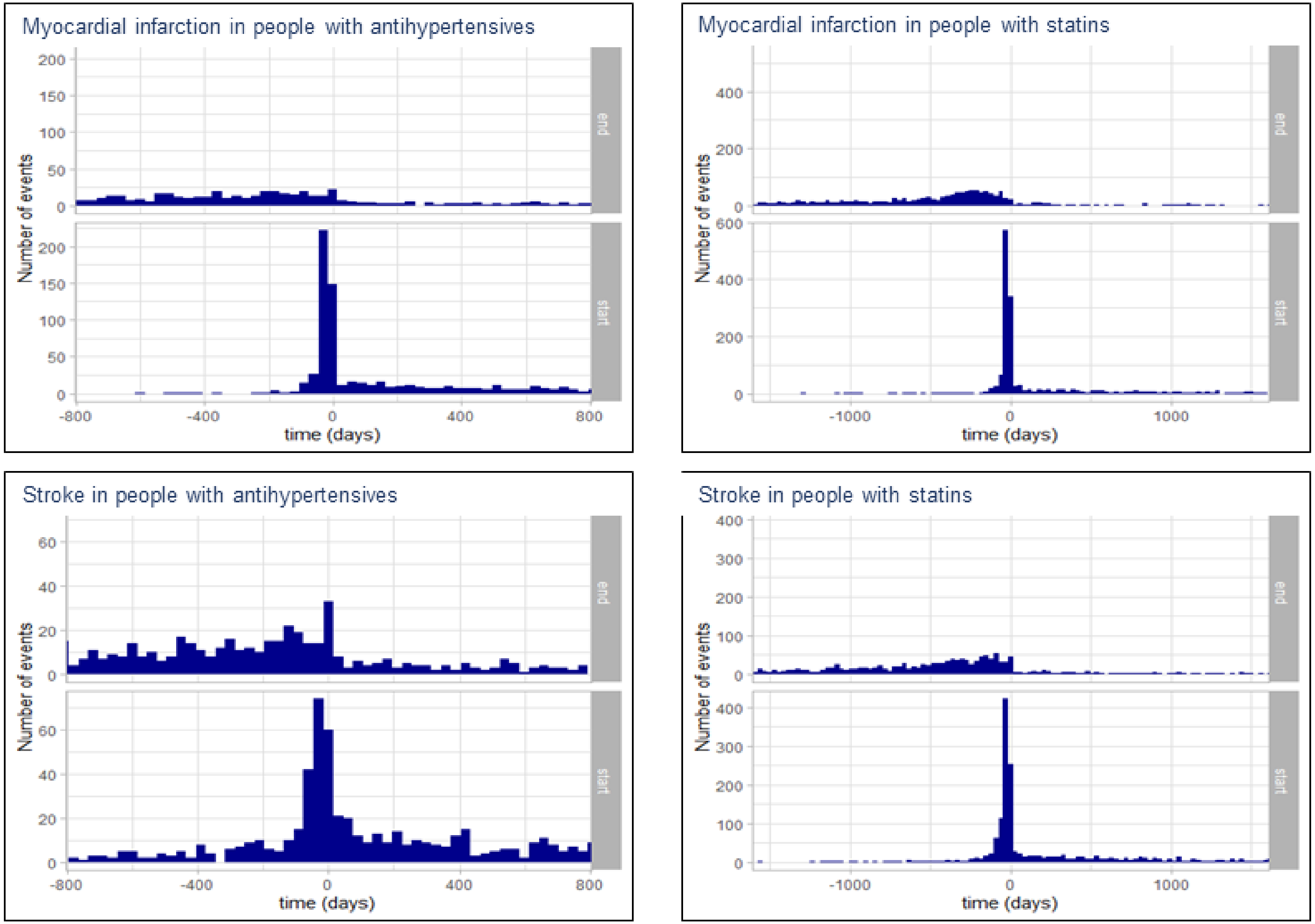


**Figure S12: Assumption testing for cardiovascular events** in CHN cohort. Distribution of number of events relative to treatment end and start for antihypertensives and statins.

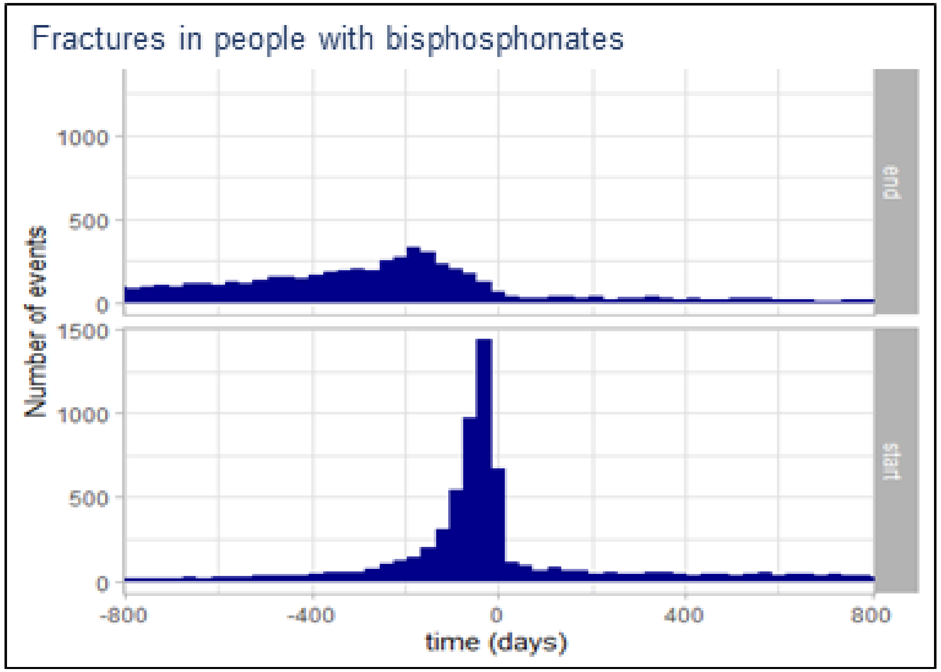


**Figure S13: Assumption testing for fractures** in CHN cohort. Distribution of number of events relative to treatment end and start of bisphosphonates.

**Table S1: Follow-up duration in year for people with and without events** in CHN cohort without previous use of respective preventative medication

| | **1$^{st}$ Qu.** [years] | **Median** [years] | **3$^{rd}$ Qu.** [years] |
|---|---|---|---|
| **Antihypertensives-naïve people** | | | |
| with MI | 3.20 | 5.24 | 6.91 |
| without MI | 1.81 | 4.21 | 5.93 |
| with stroke | 2.77 | 4.78 | 6.65 |
| without stroke | 1.80 | 4.21 | 5.93 |
| **Bisphosphonate-naïve people** | | | |
| with fractures | 3.44 | 5.23 | 6.74 |
| without fractures | 1.76 | 3.99 | 5.83 |
| **Statin-naïve people** | | | |
| with MI | 2.83 | 4.57 | 6.28 |
| without MI | 1.68 | 3.81 | 5.78 |
| with stroke | 2.66 | 4.54 | 6.28 |
| without stroke | 1.68 | 3.81 | 5.77 |

**Table S2: Number of events per person** in CHN cohort without previous use of respective preventative medication.

| | Number of events per person | | | | | |
|---|---|---|---|---|---|---|
| | **1** | **2** | **3** | **4** | **5** | **6** |
| **Myocardial infarction** | | | | | | |
| In antihypertensives-naïve people | 841 | 51 | <5 | <5 | | |
| In statin-naïve people | 2,021 | 152 | 17 | | | |
| **Fractures** | | | | | | |
| In bisphosphonate-naïve people | 15,478 | 2,565 | 448 | 88 | 22 | 5 |
| **Stroke** | | | | | | |
| In antihypertensives-naïve people | 1,123 | 104 | 9 | <5 | | |
| In statin-naïve people | 2,696 | 247 | 18 | <5 | | |
