## Supplemental File 2 - Code Lists for "Cardiovascular outcomes and fracture risk after the discontinuation of preventative medications in older patients with complex health needs: a self-controlled case series analysis"

#### Code lists: Stroke

| readcode | readterm |
|---|---|
| G61z.00 | Intracerebral haemorrhage NOS |
| G61..00 | Intracerebral haemorrhage |
| G61..11 | CVA - cerebrovascular accid due to intracerebral haemorrhage |
| G613.00 | Cerebellar haemorrhage |
| G61..12 | Stroke due to intracerebral haemorrhage |
| G61X100 | Right sided intracerebral haemorrhage, unspecified |
| G602.00 | Subarachnoid haemorrhage from middle cerebral artery |
| G61X000 | Left sided intracerebral haemorrhage, unspecified |
| G617.00 | Intracerebral haemorrhage, intraventricular |
| G61X.00 | Intracerebral haemorrhage in hemisphere, unspecified |
| Gyu6200 | [X]Other intracerebral haemorrhage |
| G618.00 | Intracerebral haemorrhage, multiple localized |
| Gyu6F00 | [X]Intracerebral haemorrhage in hemisphere, unspecified |
| G619.00 | Lobar cerebral haemorrhage |
| G64..12 | Infarction - cerebral |
| G64z.00 | Cerebral infarction NOS |
| G64z.12 | Cerebellar infarction |
| G64..13 | Stroke due to cerebral arterial occlusion |
| G64z200 | Left sided cerebral infarction |
| G64z300 | Right sided cerebral infarction |
| G671z00 | Generalised ischaemic cerebrovascular disease NOS |
| G64z.11 | Brainstem infarction NOS |
| G63y100 | Cerebral infarction due to embolism of precerebral arteries |
| G64z000 | Brainstem infarction |
| G64z400 | Infarction of basal ganglia |
| G641000 | Cerebral infarction due to embolism of cerebral arteries |
| G6X..00 | Cerebrl infarctn due/unspcf occlusn or sten/cerebrl artrs |
| G640000 | Cerebral infarction due to thrombosis of cerebral arteries |
| G671.00 | Generalised ischaemic cerebrovascular disease NOS |
| G6W..00 | Cereb infarct due unsp occlus/stenos precerebr arteries |
| L440.12 | Stroke in the puerperium |
| G63..11 | Infarction - precerebral |
| Gyu6300 | [X]Cerebrl infarctn due/unspcf occlusn or sten/cerebrl artrs |
| Gyu6G00 | [X]Cereb infarct due unsp occlus/stenos precerebr arteries |
| CVA unspecified | February 2009 |
| G66..00 | Stroke and cerebrovascular accident unspecified |
| G66..13 | CVA - Cerebrovascular accident unspecified |

| | |
|---|---|
| G66..12 | Stroke unspecified |
| G667.00 | Left sided CVA |
| G663.00 | Brain stem stroke syndrome |
| G668.00 | Right sided CVA |
| G664.00 | Cerebellar stroke syndrome |
| G660.00 | Middle cerebral artery syndrome |
| G662.00 | Posterior cerebral artery syndrome |
| G661.00 | Anterior cerebral artery syndrome |
| G665.00 | Pure motor lacunar syndrome |
| G666.00 | Pure sensory lacunar syndrome |
| Gyu6400 | [X]Other cerebral infarction |
| 8Hd6.00 | Admission to stroke unit |

### Code lists: Myocardial infarction

| Readcode | Description |
|---|---|
| G30..00 | Acute myocardial infarction |
| G30..14 | Heart attack |
| G30..15 | MI - acute myocardial infarction |
| G308.00 | Inferior myocardial infarction NOS |
| G307.00 | Acute subendocardial infarction |
| G301.00 | Other specified anterior myocardial infarction |
| 323..00 | ECG: myocardial infarction |
| G302.00 | Acute inferolateral infarction |
| G307100 | Acute non-ST segment elevation myocardial infarction |
| G300.00 | Acute anterolateral infarction |
| G30X000 | Acute ST segment elevation myocardial infarction |
| G30..11 | Attack - heart |
| G30z.00 | Acute myocardial infarction NOS |
| G301z00 | Anterior myocardial infarction NOS |
| G305.00 | Lateral myocardial infarction NOS |
| G30..17 | Silent myocardial infarction |
| G301100 | Acute anteroseptal infarction |
| G35..00 | Subsequent myocardial infarction |
| G310.00 | Postmyocardial infarction syndrome |
| G361.00 | Atrial septal defect/curr comp folow acut myocardal infarct |
| G304.00 | Posterior myocardial infarction NOS |
| G360.00 | Haemopericardium/current comp folow acut myocard infarct |
| G366.00 | Thrombosis atrium,auric append&vent/curr comp foll acute MI |
| G303.00 | Acute inferoposterior infarction |
| G30X.00 | Acute transmural myocardial infarction of unspecif site |

| | |
|---|---|
| G30..13 | Cardiac rupture following myocardial infarction (MI) |
| G38..00 | Postoperative myocardial infarction |
| G30B.00 | Acute posterolateral myocardial infarction |
| G30y.00 | Other acute myocardial infarction |
| G36..00 | Certain current complication follow acute myocardial infarct |
| G362.00 | Ventric septal defect/curr comp fol acut myocardal infarctn |
| G351.00 | Subsequent myocardial infarction of inferior wall |
| G301000 | Acute anteroapical infarction |
| G30y200 | Acute septal infarction |
| G384.00 | Postoperative subendocardial myocardial infarction |
| G350.00 | Subsequent myocardial infarction of anterior wall |
| G30yz00 | Other acute myocardial infarction NOS |
| G380.00 | Postoperative transmural myocardial infarction anterior wall |
| G35X.00 | Subsequent myocardial infarction of unspecified site |
| G381.00 | Postoperative transmural myocardial infarction inferior wall |
| G311011 | MI - myocardial infarction aborted |
| 323Z.00 | ECG: myocardial infarct NOS |
| G363.00 | Ruptur cardiac wall w'out haemopericard/cur comp fol ac MI |
| G364.00 | Ruptur chordae tendinae/curr comp fol acute myocard infarct |
| G311000 | Myocardial infarction aborted |
| 889A.00 | Diab mellit insulin-glucose infus acute myocardial infarct |
| G306.00 | True posterior myocardial infarction |
| G38z.00 | Postoperative myocardial infarction, unspecified |
| G365.00 | Rupture papillary muscle/curr comp fol acute myocard infarct |
| G353.00 | Subsequent myocardial infarction of other sites |
| Gyu3400 | [X]Acute transmural myocardial infarction of unspecif site |
| Gyu3600 | [X]Subsequent myocardial infarction of unspecified site |
| G383.00 | Postoperative transmural myocardial infarction unspec site |
| Gyu3500 | [X]Subsequent myocardial infarction of other sites |

#### Outcome Events: Fractures

| READ Code | Description |
|---|---|
| S34..00 | Fracture of ankle |
| S34x.00 | Closed fracture ankle, unspecified |
| S348.00 | Fracture of medial malleolus |
| 7K1L800 | Closed reduction of fracture of ankle |
| S340.00 | Closed fracture ankle, medial malleolus |
| S349.00 | Fracture of lateral malleolus |
| S342000 | Closed fracture ankle, lateral malleolus, low |
| S344.00 | Closed fracture ankle, bimalleolar |

| | |
|---|---|
| S342.00 | Closed fracture ankle, lateral malleolus |
| S34z.00 | Fracture of ankle, NOS |
| S346.00 | Closed fracture ankle, trimalleolar |
| S344.11 | Dupuytren's fracture, fibula |
| S4G..00 | Fracture-dislocation or subluxation ankle |
| S4G0.00 | Closed fracture-dislocation, ankle joint |
| S342100 | Closed fracture ankle, lateral malleolus, high |
| S344000 | Closed fracture ankle, bimalleolar, low fibular fracture |
| S346100 | Closed fracture ankle, trimalleolar, high fibular fracture |
| S344100 | Closed fracture ankle, bimalleolar, high fibular fracture |
| S346000 | Closed fracture ankle, trimalleolar, low fibular fracture |
| S4G2.00 | Closed fracture-subluxation, ankle joint |
| S312300 | Closed fracture distal femur, supracondylar |
| S312200 | Closed fracture of femur, lower epiphysis |
| S312.11 | Closed fracture of femur, distal end |
| S312.00 | Closed fracture distal femur |
| S4F4.00 | Closed fracture-dislocation, patello-femoral joint |
| S312500 | Closed fracture distal femur, lateral condyle |
| S4F6.00 | Closed fracture-subluxation, patello-femoral joint |
| S312400 | Closed fracture distal femur, medial condyle |
| S312000 | Closed fracture of distal femur, unspecified |
| S312600 | Closed fracture distal femur, bicondylar (T-Y fracture) |
| S312x00 | Closed fracture distal femur, comminuted/intra-articular |
| S312z00 | Closed fracture of distal femur not otherwise specified |
| S302200 | Closed fracture proximal femur, subtrochanteric |
| S310100 | Closed fracture shaft of femur |
| S314.00 | Fracture of shaft of femur |
| S305.00 | Subtrochanteric fracture |
| S30..11 | Hip fracture |
| S30..00 | Fracture of neck of femur |
| S302.00 | Closed fracture of proximal femur, pertrochanteric |
| 7K1L400 | Closed reduction of fracture of hip |
| S302400 | Closed fracture of femur, intertrochanteric |
| 7K1J000 | Cls red+int fxn proximal femoral #+screw/nail device alone |
| 7K1D01E | DHS - Dynamic hip screw primary fixation of neck of femur |
| S30y.11 | Hip fracture NOS |
| 7K1D01F | Dynamic hip screw primary fixation of neck of femur |
| S300500 | Cls # prox femur, subcapital, Garden grade unspec. |
| S30y.00 | Closed fracture of neck of femur NOS |
| S302000 | Cls # proximal femur, trochanteric section, unspecified |
| S302011 | Closed fracture of femur, greater trochanter |
| S30w.00 | Closed fracture of unspecified proximal femur |
| S304.00 | Pertrochanteric fracture |

| | |
|---|---|
| S300700 | Closed fracture proximal femur, subcapital, Garden grade II |
| S300900 | Closed fracture proximal femur, subcapital, Garden grade IV |
| S300600 | Closed fracture proximal femur, subcapital, Garden grade I |
| 7K1J500 | Primary int fxn(no red) prox fem #+screw/nail device alone |
| S300400 | Closed fracture head of femur |
| S300800 | Closed fracture proximal femur, subcapital, Garden grade III |
| S300.00 | Closed fracture proximal femur, transcervical |
| 7K1J700 | Primary int fxn(no red) prox fem #+screw/nail+plate device |
| 7K1Jd00 | Closed reduction of intracapsular # NOF internal fixat DHS |
| S300000 | Cls # prox femur, intracapsular section, unspecified |
| 7K1J012 | Cl red intracaps fract neck femur fix - Smith-Petersen nail |
| 7K1J600 | Primary int fxn(no red) prox fem #+scrw/nail+intramed device |
| S302z00 | Cls # of proximal femur, pertrochanteric section, NOS |
| S302100 | Closed fracture proximal femur, intertrochanteric, two part |
| S300A00 | Closed fracture of femur, upper epiphysis |
| 7K1JD00 | Primary cls red+int fxn prox fem #+screw/nail+plate device |
| S302012 | Closed fracture of femur, lesser trochanter |
| S300y00 | Closed fracture proximal femur, other transcervical |
| S302300 | Cls # proximal femur, intertrochanteric, comminuted |
| S300311 | Closed fracture, base of neck of femur |
| S300300 | Closed fracture proximal femur, basicervical |
| 7K1J011 | Cl red intracaps frac neck femur fix-Garden cannulated screw |
| 7K1JC00 | Prim cls rd+int fxn prox fem #+screw/nail+intramdulry device |
| 7K1JB00 | Primary cls red+int fxn prox fem #+screw/nail device alone |
| 7K1J013 | Cls red+int fxn prox femoral #+Richard's cannulat hip screw |
| S300z00 | Closed fracture proximal femur, transcervical, NOS |
| S300200 | Closed fracture proximal femur, midcervical section |
| S300y11 | Closed fracture of femur, subcapital |
| S300100 | Closed fracture proximal femur, transepiphyseal |
| 7K1K300 | Primary external fixation(without reduction) prox femoral # |
| 7K1K500 | Primary cls reduction+external fixation proximal femoral # |
| 7K1Y000 | Remanip intracap fract neck fem and fix using nail or screw |
| S10B600 | Multiple fractures of lumbar spine and pelvis |
| S10B.00 | Fracture of lumbar spine and pelvis |
| S32..00 | Fracture of patella |
| S20..00 | Fracture of clavicle |
| S22..00 | Fracture of humerus |
| S31z.00 | Fracture of femur, NOS |
| S339.00 | Fracture of fibula alone |
| S23x211 | Fracture of ulna NOS |
| S21..00 | Fracture of scapula |
| S28..11 | Ill-defined fracture of arm |
| S224.11 | Elbow fracture - closed |

| | |
|---|---|
| S228.00 | Fracture of lower end of humerus |
| S237.00 | Fracture of upper end of radius |
| S3...11 | Leg fracture |
| S28z.00 | Ill-defined fractures of upper limb NOS |
| S122.00 | Closed fracture sternum |
| S210300 | Closed fracture scapula, glenoid |
| S20..11 | Collar bone fracture |
| S33x100 | Closed fracture of fibula, unspecified part, NOS |
| S352300 | Closed fracture cuboid |
| S242300 | Multiple fractures of metacarpal bones |
| S210400 | Closed fracture scapula, blade |
| S2...11 | Arm fracture |
| S2...00 | Fracture of upper limb |
| 7K1J.00 | Closed (or no) reduction of fracture and internal fixation |
| S312100 | Closed fracture of femoral condyle, unspecified |
| S339000 | Closed fracture of distal fibula |
| S310.00 | Closed fracture of femur, shaft or unspecified part |
| S224100 | Closed fracture distal humerus, supracondylar |
| S230600 | Closed fracture radius, head |
| 7K1LE00 | Closed reduction of fracture of elbow |
| S31..00 | Other fracture of femur |
| S350.11 | Heel bone fracture |
| S4A0.00 | Closed fracture-dislocation shoulder |
| S315.00 | Fracture of lower end of femur |
| ZV67400 | [V]Fracture follow-up |
| S224600 | Closed fracture distal humerus, lateral epicondyle |
| 7K1L600 | Closed reduction of fracture of knee |
| 7K1LJ00 | Closed reduction of fracture of thumb |
| S3...00 | Fracture of lower limb |
| S3x3.00 | Multiple fractures of lower leg |
| S4B0000 | Closed fracture-dislocation elbow joint |
| S230100 | Closed fracture olecranon, extra-articular |
| S22z.00 | Fracture of humerus NOS |
| S21..11 | Shoulder blade fracture |
| S3X..00 | Fracture of lower leg, part unspecified |
| 7K1LB00 | Closed reduction of fracture of hallux |
| N331M00 | Fragility fracture due to unspecified osteoporosis |
| S128.00 | Fracture of sternum |
| S230B00 | Closed fracture olecranon, intra-articular |
| S224.00 | Closed fracture of the distal humerus |
| S23x.00 | Closed fracture of radius and ulna, unspecified part |
| S210100 | Closed fracture scapula, acromion |
| S292.00 | Multiple fractures of clavicle, scapula and humerus |

| | |
|---|---|
| S240700 | Closed fracture capitate |
| S23x100 | Closed fracture of radius (alone), unspecified |
| S224200 | Closed fracture distal humerus, lateral condyle |
| NyuB800 | [X]Unspecified osteoporosis with pathological fracture |
| 7K1L500 | Closed reduction of fracture of femur |
| S222000 | Closed fracture of humerus NOS |
| S310012 | Upper leg fracture NOS |
| S4A2100 | Closed fracture-subluxation acromio-clavicular joint |
| 7K1K.00 | Closed (or no) reduction of fracture and external fixation |
| N331600 | Idiopathic osteoporosis with pathological fracture |
| S210.00 | Closed fracture of scapula |
| S4A0100 | Closed fracture-dislocation acromio-clavicular joint |
| S12z.12 | Sternum fracture NOS |
| S20z.00 | Fracture of clavicle NOS |
| S200z00 | Closed fracture of clavicle NOS |
| S200200 | Closed fracture clavicle, shaft |
| S224800 | Closed fracture distal humerus, capitellum |
| S332100 | Closed fracture shaft of fibula |
| S224700 | Closed fracture distal humerus, medial epicondyle |
| S200300 | Closed fracture clavicle, lateral end |
| S2z..00 | Fracture of upper limb NOS |
| S352111 | Closed fracture of astragalus |
| S227.00 | Fracture of shaft of humerus |
| S29..11 | Multiple fractures of arm |
| S224400 | Closed fracture of distal humerus, condyle(s) unspecified |
| S320.00 | Closed fracture of the patella |
| S236.00 | Fracture of upper end of ulna |
| N331300 | Osteoporosis of disuse with pathological fracture |
| S224z00 | Closed fracture of distal humerus, not otherwise specified |
| S330100 | Closed fracture proximal fibula |
| S4F2.00 | Closed fracture-subluxation, knee joint |
| S4B0.00 | Closed fracture-dislocation elbow |
| S4A0000 | Closed fracture-dislocation shoulder joint |
| S224000 | Closed fracture of elbow, unspecified part |
| S200.00 | Closed fracture of clavicle |
| S21z.00 | Fracture of scapula NOS |
| S2A..00 | Fracture of upper limb, level unspecified |
| S230900 | Closed fracture of the proximal radius |
| S37..00 | Fracture of lower limb, level unspecified |
| S230000 | Closed fracture of proximal forearm, unspecified part |
| S230800 | Closed fracture proximal radius, comminuted |
| S230400 | Closed fracture of proximal ulna, comminuted |
| S230500 | Closed fracture of the proximal ulna |

| | |
|---|---|
| S350100 | Closed fracture calcaneus, intra-articular |
| S210200 | Closed fracture scapula, coracoid |
| S32z.00 | Fracture of patella, NOS |
| S232300 | Closed fracture radius and ulna, middle |
| S33x.11 | Lower leg fracture NOS |
| S4A2.00 | Closed fracture-subluxation shoulder |
| S352H00 | Closed fracture of cuneiforms |
| S210600 | Closed fracture scapula, neck |
| S352800 | Closed fracture talus, head |
| S310000 | Closed fracture of femur, unspecified part |
| S210000 | Closed fracture of scapula, unspecified part |
| 7K1LN00 | Closed reduction of fracture of upper limb |
| N331B00 | Postmenopausal osteoporosis with pathological fracture |
| S224300 | Closed fracture distal humerus, medial condyle |
| S370.00 | Closed fracture of lower limb, level unspecified |
| S4F0.00 | Closed fracture-dislocation, knee joint |
| S292000 | Closed multiple fractures of clavicle, scapula and humerus |
| S320400 | Closed fracture patella, comminuted (stellate) |
| S232100 | Closed fracture of the radial shaft |
| S230.00 | Closed fracture of proximal radius and ulna |
| S320200 | Closed fracture patella, distal pole |
| S230A00 | Closed fracture radius and ulna, proximal |
| S200100 | Closed fracture clavicle, medial end |
| S200000 | Closed fracture of clavicle, unspecified part |
| S352A00 | Closed fracture talus, body |
| N331500 | Drug-induced osteoporosis with pathological fracture |
| S352900 | Closed fracture talus, neck |
| S280.00 | Closed ill-defined fractures of upper limb |
| ZV66400 | [V]Convalescence after treatment of fracture |
| S210500 | Closed fracture scapula, spine |
| S28..00 | Ill-defined fractures of upper limb |
| S320000 | Closed fracture patella, transverse |
| S352600 | Closed fracture lateral cuneiform |
| S320100 | Closed fracture patella, proximal pole |
| S224500 | Closed fracture of distal humerus, trochlea |
| S310z00 | Closed fracture of shaft or unspecified part, NOS |
| S330900 | Closed fracture fibula, neck |
| S330800 | Closed fracture fibula, head |
| Zw01.00 | [Q] Fractures involving the epiphyseal plate |
| S224900 | Closed fracture distal humerus, bicondylar (T-Y fracture) |
| S12X.00 | Fracture of bony thorax, part unspecified |
| S320300 | Closed fracture patella, vertical |
| 7K1LC00 | Closed reduction of fracture of lower limb |

| | |
|---|---|
| SR16000 | Closed fracture inv thorax wth low back and pelvis and limbs |
| NyuB000 | [X]Other osteoporosis with pathological fracture |
| S210z00 | Closed fracture of scapula NOS |
| S350000 | Closed fracture calcaneus, extra-articular |
| S4B2.00 | Closed fracture-subluxation elbow |
| S4A2000 | Closed fracture-subluxation shoulder joint |
| S4J2000 | Closed fracture-subluxation of sternum |
| S224x00 | Closed fracture of distal humerus, multiple |
| S4J0000 | Closed fracture-dislocation of sternum |
| S4B2000 | Closed fracture-subluxation elbow joint |
| S29..13 | Multiple fractures of sternum |
| Syu7200 | [X]Fractures of other parts of femur |
| SR1z000 | [X]Closed multiple fractures unspecified |
| SR12000 | Closed fractures involving multiple regions of one upp limb |
| N331N00 | Fragility fracture |
| N331M11 | Minimal trauma fracture due to unspecified osteoporosis |
| N331N11 | Minimal trauma fracture |
| Zw02500 | [Q] Refracture |
| S294000 | Cl fractures involving multiple regions of both upper limbs |
| S12y000 | Closed fracture of other parts of bony thorax |
| Syu8300 | [X]Fractures of other parts of lower leg |
| Syu4200 | [X]Multiple fractures of clavicle, scapula and humerus |
| Syu9400 | [X]Fracture of other tarsal bones |
| S12y.00 | Fracture of other parts of bony thorax |
| Syu8D00 | [X]Fracture of lower leg, part unspecified |
| SR15000 | Cl fractures involving multiple regions upper with lower lmb |
| S106000 | Closed compression fracture sacrum |
| S130.00 | Closed fracture acetabulum |
| S130z00 | Closed fracture acetabulum NOS |
| S130200 | Closed fracture acetabulum, anterior column |
| S130000 | Closed fracture acetabulum, anterior lip alone |
| S130600 | Closed fracture acetabulum, double column unspecified |
| S130400 | Closed fracture acetabulum, floor |
| S130300 | Closed fracture acetabulum, posterior column |
| S130100 | Closed fracture acetabulum, posterior lip alone |
| S134800 | Closed fracture dislocation of sacro-iliac joint |
| S134000 | Closed fracture of ilium, unspecified |
| S13y.00 | Closed fracture of pelvis NOS |
| S134500 | Closed fracture pelvis, anterior inferior iliac spine |
| S134400 | Closed fracture pelvis, anterior superior iliac spine |
| S108.00 | Closed fracture pelvis, coccyx |
| S134600 | Closed fracture pelvis, iliac wing |
| S134300 | Closed fracture pelvis, ischial tuberosity |

| | |
|---|---|
| S134100 | Closed fracture pelvis, ischium |
| S132100 | Closed fracture pelvis, multiple pubic rami - stable |
| S132200 | Closed fracture pelvis, multiple pubic rami - unstable |
| S132000 | Closed fracture pelvis, single pubic ramus |
| S132.00 | Closed fracture pubis |
| S132z00 | Closed fracture pubis NOS |
| S106.00 | Closed fracture sacrum |
| S4J0100 | Closed fracture-dislocation of pelvis |
| S4J2100 | Closed fracture-subluxation of pelvis |
| S134700 | Closed vertical fracture of ilium |
| S106100 | Closed vertical fracture of sacrum |
| S10B400 | Fracture of acetabulum |
| S10B200 | Fracture of coccyx |
| S10B300 | Fracture of ilium |
| S10B500 | Fracture of pubis |
| S10B100 | Fracture of sacrum |
| S13..00 | Fracture or disruption of pelvis |
| S134.00 | Other or multiple closed fracture of pelvis |
| S134z00 | Other or multiple closed fracture of pelvis NOS |
| S130y00 | Other specified closed fracture acetabulum |
| S132y00 | Other specified closed fracture pubis |
| S118.00 | Closed fracture of coccyx with spinal cord lesion |
| S116.00 | Closed fracture of sacrum with spinal cord lesion |
| S116z00 | Closed fracture of sacrum with spinal cord lesion NOS |
| S118z00 | Closed fracture of coccyx with spinal cord lesion NOS |
| S120900 | Closed fracture multiple ribs |
| S120800 | Closed fracture of eight or more ribs |
| S120500 | Closed fracture of five ribs |
| S120400 | Closed fracture of four ribs |
| S120100 | Closed fracture of one rib |
| S120z00 | Closed fracture of rib(s) NOS |
| S120000 | Closed fracture of rib, unspecified |
| S120700 | Closed fracture of seven ribs |
| S120600 | Closed fracture of six ribs |
| S120300 | Closed fracture of three ribs |
| S120200 | Closed fracture of two ribs |
| S120.00 | Closed fracture rib |
| S127100 | Cough fracture of ribs |
| S127000 | Multiple fractures of ribs |
| S29..12 | Multiple rib fractures |
| S12z.11 | Rib fracture NOS |
| S127.00 | Fracture of rib |
| S120A00 | Cough fracture |

| | |
|---|---|
| S12..00 | Fracture of rib(s), sternum, larynx and trachea |
| S12z.00 | Fracture of rib(s), sternum, larynx or trachea NOS |
| S226.00 | Fracture of upper end of humerus |
| 7K1LF00 | Closed reduction of fracture of humerus |
| 7K1LG00 | Closed reduction of fracture of shoulder |
| S220300 | Closed fracture proximal humerus, greater tuberosity |
| S220.00 | Closed fracture of the proximal humerus |
| S220100 | Closed fracture proximal humerus, neck |
| S222100 | Closed fracture of humerus, shaft |
| S220400 | Closed fracture proximal humerus, head |
| S220700 | Closed fracture proximal humerus, four part |
| S220200 | Closed fracture of proximal humerus, anatomical neck |
| S222.00 | Closed fracture of humerus, shaft or unspecified part |
| S220z00 | Closed fracture of proximal humerus not otherwise specified |
| S220600 | Closed fracture proximal humerus, three part |
| S220000 | Closed fracture of proximal humerus, unspecified part |
| S220500 | Closed fracture of humerus, upper epiphysis |
| Syu4300 | [X]Fracture of other parts of shoulder and upper arm |
| Syu4400 | [X]Fracture of shoulder and upper arm, unspecified |
| S222z00 | Closed fracture of humerus, shaft or unspecified part NOS |
| S104.00 | Closed fracture lumbar vertebra |
| S104000 | Closed fracture lumbar vertebra, burst |
| S104500 | Closed fracture lumbar vertebra, posterior arch |
| S104300 | Closed fracture lumbar vertebra, spinous process |
| S104200 | Closed fracture lumbar vertebra, spondylolysis |
| S104400 | Closed fracture lumbar vertebra, transverse process |
| S104600 | Closed fracture lumbar vertebra, tricolumnar |
| S104100 | Closed fracture lumbar vertebra, wedge |
| S114.00 | Closed fracture of lumbar spine with spinal cord lesion |
| S11x.00 | Closed fracture of spine with spinal cord lesion unspecified |
| S10x.00 | Closed fracture of spine, unspecified, |
| S112z00 | Closed fracture of thoracic spine with cord lesion NOS |
| S112.00 | Closed fracture of thoracic spine with spinal cord lesion |
| S102.00 | Closed fracture thoracic vertebra |
| S102z00 | Closed fracture thoracic vertebra not otherwise specified |
| S102000 | Closed fracture thoracic vertebra, burst |
| S102500 | Closed fracture thoracic vertebra, posterior arch |
| S102300 | Closed fracture thoracic vertebra, spinous process |
| S102200 | Closed fracture thoracic vertebra, spondylolysis |
| S102400 | Closed fracture thoracic vertebra, transverse process |
| S102100 | Closed fracture thoracic vertebra, wedge |
| S150000 | Closed multiple fractures of thoracic spine |
| S114500 | Closed spinal fracture with cauda equina lesion |

| | |
|---|---|
| S114100 | Closed spinal fracture with complete lumbar cord lesion |
| S114000 | Closed spinal fracture with unspecified lumbar cord lesion |
| S112700 | Cls spinal fracture with complete thorac cord lesion, T7-12 |
| S112A00 | Cls spinal fracture with posterior thorac cord lesion, T7-12 |
| S112600 | Cls spinal fracture with unspec thoracic cord lesion, T7-12 |
| S112100 | Cls spinal fracture wth complete thoracic cord lesion,T1-6 |
| N1y1.00 | Fatigue fracture of vertebra |
| S150.00 | Multiple fractures of thoracic spine |
| N331A00 | Osteoporosis + pathological fracture cervical vertebrae |
| N331800 | Osteoporosis + pathological fracture lumbar vertebrae |
| N331900 | Osteoporosis + pathological fracture thoracic vertebrae |
| 7J41500 | Balloon kyphoplasty of fracture of spine |
| S112000 | Cls spinal fracture with unspec thoracic cord lesion,T1-6 |
| S10B000 | Fracture of lumbar vertebra |
| S10..00 | Fracture of spine without mention of spinal cord injury |
| S10z.00 | Fracture of spine without mention of spinal cord lesion NOS |
| S15..00 | Fracture of thoracic vertebra |
| S10..12 | Fracture of vertebra without spinal cord lesion |
| S102y00 | Other specified closed fracture thoracic vertebra |
| N331100 | Pathological fracture of lumbar vertebra |
| N331000 | Pathological fracture of thoracic vertebra |
| 7J42600 | Primary bedrest stabilisation of spinal fracture |
| 7J42900 | Primary cast stabilisation of spinal fracture |
| 7J41300 | Vertebroplasty of fracture of spine |
| S33x000 | Closed fracture of tibia, unspecified part, NOS |
| S33..00 | Fracture of tibia and fibula |
| S33x200 | Closed fracture of tibia and fibula, unspecified part |
| 7K1L700 | Closed reduction of fracture of tibia and or fibula |
| S337.00 | Fracture of shaft of tibia |
| S334100 | Closed fracture distal tibia, intra-articular |
| S338.00 | Fracture of lower end of tibia |
| S33z.00 | Fracture of tibia and fibula, NOS |
| S334.00 | Closed fracture distal tibia |
| S33x.00 | Closed fracture of tibia and fibula, unspecified part, NOS |
| S332.00 | Closed fracture of tibia/fibula, shaft |
| S332200 | Closed fracture of tibia and fibula, shaft |
| S332000 | Closed fracture shaft of tibia |
| S334000 | Closed fracture distal tibia, extra-articular |
| S33xz00 | Closed fracture of tibia and fibula, unspecified part, NOS |
| S332z00 | Closed fracture of tibia and fibula, shaft, NOS |
| S33A.00 | Fracture of tibia |
| S33C.00 | Closed fracture of distal tibia and fibula |
| S336.00 | Fracture of upper end of tibia |

| | |
|---|---|
| S330300 | Closed fracture proximal tibia, medial condyle (plateau) |
| S330400 | Closed fracture proximal tibia, lateral condyle (plateau) |
| S330012 | Closed fracture of tibial tuberosity |
| S330000 | Closed fracture of the proximal tibia |
| S330600 | Closed fracture spine, tibia |
| S330700 | Closed fracture tubercle, tibia |
| S330500 | Closed fracture proximal tibia, bicondylar |
| S330z00 | Closed fracture of tibia and fibula, proximal NOS |
| S330.00 | Closed fracture of tibia and fibula, proximal |
| S330011 | Closed fracture of tibial condyles |
| S330200 | Closed fracture of tibia and fibula, proximal |
| S336000 | Fracture tibial plateau |
| Syu5400 | [X]Fracture of forearm, unspecified |
| Syu5300 | [X]Fracture of other parts of forearm |
| S234A00 | Closd dorsal Barton's fracture |
| S234F00 | Closed Barton's fracture |
| S234100 | Closed Colles' fracture |
| S234A11 | Closed dorsal Barton's fracture-dislocation |
| S234D00 | Closed fracture distal radius, extra-articular, other type |
| S234C00 | Closed fracture distal radius, intra-articular, die-punch |
| S234E00 | Closed fracture distal radius, intra-articular, other type |
| S234500 | Closed fracture distal ulna, unspecified |
| S234z00 | Closed fracture of forearm, lower end, NOS |
| S234000 | Closed fracture of forearm, lower end, unspecified |
| S23x000 | Closed fracture of forearm, unspecified |
| S234.00 | Closed fracture of radius and ulna, lower end |
| S234200 | Closed fracture of the distal radius, unspecified |
| S23x300 | Closed fracture of the radius and ulna |
| S23x200 | Closed fracture of ulna (alone), unspecified |
| S230200 | Closed fracture of ulna, coronoid |
| S234400 | Closed fracture of ulna, lower epiphysis |
| S234300 | Closed fracture of ulna, styloid process |
| S234B00 | Closed fracture radial styloid |
| S234600 | Closed fracture radius and ulna, distal |
| S4C0000 | Closed fracture-dislocation distal radio-ulnar joint |
| S4C0100 | Closed fracture-dislocation radiocarpal joint |
| READ.00 | Closed fracture-subluxation of the wrist |
| S4C2100 | Closed fracture-subluxation radiocarpal joint |
| S4C2000 | Closed fracture-subluxation, distal radio-ulnar jt |
| S234800 | Closed Galeazzi fracture |
| S230300 | Closed Monteggia's fracture |
| S234700 | Closed Smith's fracture |
| S234211 | Dupuytren's fracture, radius - closed |

| | |
|---|---|
| S23..11 | Forearm fracture |
| S293.00 | Multiple fractures of forearm |
| S234111 | Smith's fracture - closed |
| S234.11 | Wrist fracture - closed |
| S234A12 | Closed dorsal Barton fracture-subluxation |
| S240F00 | Closed fracture carpal bones, multiple |
| S4C0.00 | Closed fracture dislocation of wrist |
| S240.00 | Closed fracture of carpal bone |
| S240z00 | Closed fracture of carpal bone NOS |
| S240000 | Closed fracture of carpal bone, unspecified |
| S240y00 | Closed fracture of other carpal bone |
| S230z00 | Closed fracture of proximal forearm not otherwise specified |
| S23xz00 | Closed fracture of radius and ulna, NOS |
| S232.00 | Closed fracture of radius and ulna, shaft |
| S232z00 | Closed fracture of radius and ulna, shaft, NOS |
| S232000 | Closed fracture of radius, shaft, unspecified |
| S232200 | Closed fracture of the ulnar shaft |
| S230700 | Closed fracture radius, neck |
| S4C0200 | Closed fracture-dislocation mid carpal |
| S4B0100 | Closed fracture-dislocation superior radio-ulnar joint |
| S4C0300 | Closed fracture-dislocation, carpometacarpal joint |
| S4C2200 | Closed fracture-subluxation mid carpal |
| S4C2y00 | Closed fracture-subluxation other carpal |
| S4B2100 | Closed fracture-subluxation superior radio-ulnar joint |
| S4C2300 | Closed fracture-subluxation, carpometacarpal joint |
| 7K1LL00 | Closed reduction of fracture of radius and or ulna |
| 7K1LM00 | Closed reduction of fracture of wrist |
| S234912 | Closed volar Barton fracture-subluxation |
| S234900 | Closed volar Barton's fracture |
| S234911 | Closed volar Barton's fracture-dislocation |
| S242.00 | Fracture at wrist and hand level |
| S24..00 | Fracture of carpal bone |
| S24z.00 | Fracture of carpal bone NOS |
| S23C.00 | Fracture of lower end of both ulna and radius |
| S23B.00 | Fracture of lower end of radius |
| S23..00 | Fracture of radius and ulna |
| S23z.00 | Fracture of radius and ulna, NOS |
| S23x111 | Fracture of radius NOS |
| S239.00 | Fracture of shaft of radius |
| S238.00 | Fracture of shaft of ulna |
| S23A.00 | Fracture of shafts of both ulna and radius |
| S4C..00 | Fracture-dislocation or subluxation of wrist |
| S234G00 | Greenstick fracture of distal radius |

| | |
|---|---|
| S24..11 | Hand fracture - carpal bone |
| Syu6300 | [X]Fracture of other carpal bone(s) |
| Syu6500 | [X]Fracture of other & unspecified parts of wrist and hand |

### Code lists: Preventative medication

| Product code | dmd code | Drug substance name | Product name |
|---|---|---|---|
| **ANTIHYPERTENSIVES** | | | |
| **ACE-inhibitors** | | | |
| 4103 | 318925003 | Trandolapril | Trandolapril 1mg capsules |
| 5047 | 318926002 | Trandolapril | Trandolapril 2mg capsules |
| 7419 | 318924004 | Trandolapril | Trandolapril 500microgram capsules |
| 8025 | 346811000001101 | Trandolapril | Gopten 1mg capsules (Abbott Laboratories Ltd) |
| 8026 | 273111000001109 | Trandolapril | Gopten 2mg capsules (Abbott Laboratories Ltd) |
| 9948 | 410958005 | Trandolapril | Trandolapril 4mg capsules |
| 16710 | 253511000001102 | Trandolapril | Gopten 500microgram capsules (Abbott Laboratories Ltd) |
| 28902 | 140511000001108 | Trandolapril | Odrik 2mg capsules (Aventis Pharma) |
| 29130 | 5651611000001103 | Trandolapril | Gopten 4mg capsules (Abbott Laboratories Ltd) |
| 31307 | 227511000001106 | Trandolapril | Odrik 500microgram capsules (Aventis Pharma) |
| 31810 | 432911000001102 | Trandolapril | Odrik 1mg capsules (Aventis Pharma) |
| 54345 | 15162111000001105 | Trandolapril | Trandolapril 4mg capsules (Arrow Generics Ltd) |
| 60757 | 13433411000001109 | Trandolapril | Trandolapril 500microgram capsules (Teva UK Ltd) |
| 65389 | 5372611000001104 | Trandolapril | Gopten 500microgram capsules (Waymade Healthcare Plc) |
| 65570 | 13494811000001100 | Trandolapril | Trandolapril 4mg capsules (Teva UK Ltd) |
| 66623 | 13470311000001106 | Trandolapril | Trandolapril 2mg capsules (A A H Pharmaceuticals Ltd) |
| 74209 | 14407611000001105 | Trandolapril | Trandolapril 1mg capsules (Actavis UK Ltd) |
| 80 | 318902004 | Ramipril | Ramipril 5mg capsules |
| 82 | 318906001 | Ramipril | Ramipril 10mg capsules |
| 147 | 318900007 | Ramipril | Ramipril 1.25mg capsules |
| 654 | 212915001000027107 | Ramipril | Ramipril 2.5/ 5mg/ 10mg capsule |
| 709 | 318901006 | Ramipril | Ramipril 2.5mg capsules |
| 756 | 408052000 | Ramipril | Ramipril 10mg tablets |
| 761 | 408040007 | Ramipril | Ramipril 1.25mg tablets |
| 5275 | 835411000001105 | Ramipril | Tritace 2.5mg capsules (Sanofi) |
| 5735 | 802311000001101 | Ramipril | Tritace 5mg capsules (Sanofi) |
| 6261 | 5010511000001106 | Ramipril | Tritace 1.25mg tablets (Sanofi) |
| 6288 | 408051007 | Ramipril | Ramipril 5mg tablets |

| | | | |
|---|---|---|---|
| 6314 | 408050008 | Ramipril | Ramipril 2.5mg tablets |
| 6362 | 5011111000001108 | Ramipril | Tritace 5mg tablets (Sanofi) |
| 6364 | 5010811000001109 | Ramipril | Tritace 2.5mg tablets (Sanofi) |
| 9646 | 111611000001109 | Ramipril | Tritace 1.25mg capsules (Aventis Pharma) |
| 9693 | 43711000001100 | Ramipril | Tritace 10mg capsules (Sanofi) |
| 9915 | 5011411000001103 | Ramipril | Tritace 10mg tablets (Sanofi) |
| 11937 | 8720711000001102 | Ramipril | Ramipril 2.5mg/ 5ml oral suspension |
| 28586 | 7948911000001100 | Ramipril | Lopace 5mg capsules (Discovery Pharmaceuticals) |
| 29627 | 7948711000001102 | Ramipril | Lopace 2.5mg capsules (Discovery Pharmaceuticals) |
| 32857 | 5587511000001109 | Ramipril | Ramipril 1.25mg capsules (Teva UK Ltd) |
| 32934 | 7949111000001105 | Ramipril | Lopace 10mg capsules (Discovery Pharmaceuticals) |
| 33811 | 7817811000001108 | Ramipril | Ramipril 2.5mg capsules (Ranbaxy (UK) Ltd) |
| 33894 | 5589711000001107 | Ramipril | Ramipril 10mg capsules (Teva UK Ltd) |
| 34357 | 5629611000001101 | Ramipril | Ramipril 10mg capsules (Genus Pharmaceuticals Ltd) |
| 34382 | 5631411000001102 | Ramipril | Ramipril 5mg capsules (Zentiva) |
| 34390 | 5629311000001106 | Ramipril | Ramipril 5mg capsules (Genus Pharmaceuticals Ltd) |
| 34412 | 5588511000001108 | Ramipril | Ramipril 5mg capsules (Teva UK Ltd) |
| 34429 | 7433211000001103 | Ramipril | Ramipril 5mg capsules (Mylan) |
| 34431 | 5631211000001101 | Ramipril | Ramipril 2.5mg capsules (Zentiva) |
| 34432 | 5628711000001101 | Ramipril | Ramipril 2.5mg capsules (Genus Pharmaceuticals Ltd) |
| 34490 | 5588011000001100 | Ramipril | Ramipril 2.5mg capsules (Teva UK Ltd) |
| 34505 | 5623511000001102 | Ramipril | Ramipril 2.5mg capsules (Sandoz Ltd) |
| 34528 | 5875911000001104 | Ramipril | Ramipril 2.5mg capsules (A A H Pharmaceuticals Ltd) |
| 34539 | 5624411000001103 | Ramipril | Ramipril 5mg capsules (Sandoz Ltd) |
| 34540 | 5878511000001103 | Ramipril | Ramipril 5mg capsules (A A H Pharmaceuticals Ltd) |
| 34567 | 7433011000001108 | Ramipril | Ramipril 2.5mg capsules (Mylan) |
| 34583 | 14776501000027101 | Ramipril | Ramipril 10mg Capsule (Dexcel-Pharma Ltd) |
| 34589 | 14772501000027109 | Ramipril | Ramipril 5mg Capsule (Dexcel-Pharma Ltd) |
| 34651 | 7433511000001100 | Ramipril | Ramipril 10mg capsules (Mylan) |
| 34652 | 14715501000027104 | Ramipril | Ramipril 5mg Capsule (Sovereign Medical Ltd) |
| 34657 | 5631611000001104 | Ramipril | Ramipril 10mg capsules (Zentiva) |
| 34698 | 7338611000001103 | Ramipril | Ramipril 1.25mg capsules (Zentiva) |
| 34710 | 5624611000001100 | Ramipril | Ramipril 10mg capsules (Sandoz Ltd) |
| 34732 | 14768501000027107 | Ramipril | Ramipril 2.5mg Capsule (Dexcel-Pharma Ltd) |
| 34877 | 14721501000027107 | Ramipril | Ramipril 10mg Capsule (Sovereign Medical Ltd) |
| 34893 | 15347501000027100 | Ramipril | Ramipril 10mg Capsule (IVAX Pharmaceuticals UK Ltd) |
| 34943 | 5880511000001109 | Ramipril | Ramipril 10mg capsules (A A H Pharmaceuticals Ltd) |
| 35007 | 8720511000001107 | Ramipril | Ramipril 10mg/ 5ml oral suspension |
| 38308 | 25153501000027101 | Ramipril | Ramipril 2.5/ 5mg/ 10mg tablet |
| 39355 | 23705501000027109 | Ramipril | Tritace 10mg Tablet (Sterwin Medicines) |
| 40384 | 5885911000001106 | Ramipril | Ramipril 10mg tablets (A A H Pharmaceuticals Ltd) |
| 42081 | 23702501000027108 | Ramipril | Tritace 1.25mg Tablet (Sterwin Medicines) |
| 45264 | 8266111000001102 | Ramipril | Ramipril 1.25mg capsules (Actavis UK Ltd) |

| | | | |
|---|---|---|---|
| 45340 | 15766501000027100 | Ramipril | Ramipril 10mg Capsule (Actavis UK Ltd) |
| 45554 | 8720811000001105 | Ramipril | Ramipril 5mg/ 5ml oral solution |
| 46890 | 8720911000001100 | Ramipril | Ramipril 5mg/ 5ml oral suspension |
| 47021 | 19877111000001100 | Ramipril | Ramipril 2.5mg/ 5ml oral solution sugar free |
| 47998 | 8266411000001107 | Ramipril | Ramipril 2.5mg capsules (Actavis UK Ltd) |
| 48008 | 8267111000001104 | Ramipril | Ramipril 5mg capsules (Actavis UK Ltd) |
| 48053 | 9805611000001106 | Ramipril | Ramipril 2.5mg capsules (Almus Pharmaceuticals Ltd) |
| 49164 | 8267511000001108 | Ramipril | Ramipril 10mg capsules (Actavis UK Ltd) |
| 50509 | 8720411000001108 | Ramipril | Ramipril 10mg/ 5ml oral solution |
| 51701 | 16065011000001105 | Ramipril | Ramipril 5mg capsules (Bristol Laboratories Ltd) |
| 51714 | 5921211000001103 | Ramipril | Ramipril 2.5mg capsules (Alliance Healthcare (Distribution) Ltd) |
| 52197 | 15183911000001103 | Ramipril | Ramipril 5mg capsules (Sigma Pharmaceuticals Plc) |
| 52399 | 5632211000001108 | Ramipril | Ramipril 1.25mg capsules (Kent Pharmaceuticals Ltd) |
| 52407 | 5632811000001109 | Ramipril | Ramipril 10mg capsules (Kent Pharmaceuticals Ltd) |
| 53612 | 5937511000001108 | Ramipril | Ramipril 10mg tablets (Alliance Healthcare (Distribution) Ltd) |
| 53621 | 16064811000001100 | Ramipril | Ramipril 2.5mg capsules (Bristol Laboratories Ltd) |
| 54298 | 10412211000001105 | Ramipril | Ramipril 2.5mg capsules (Arrow Generics Ltd) |
| 54620 | 15183111000001101 | Ramipril | Ramipril 2.5mg capsules (Sigma Pharmaceuticals Plc) |
| 54941 | 5923011000001109 | Ramipril | Ramipril 5mg capsules (Alliance Healthcare (Distribution) Ltd) |
| 55299 | 5873711000001102 | Ramipril | Ramipril 1.25mg capsules (A A H Pharmaceuticals Ltd) |
| 55798 | 21878111000001106 | Ramipril | Ramipril 5mg capsules (Waymade Healthcare Plc) |
| 56013 | 21877911000001108 | Ramipril | Ramipril 2.5mg capsules (Waymade Healthcare Plc) |
| 56038 | 20005711000001100 | Ramipril | Ramipril 10mg tablets (Pfizer Ltd) |
| 56129 | 5632611000001105 | Ramipril | Ramipril 5mg capsules (Kent Pharmaceuticals Ltd) |
| 56148 | 7562211000001109 | Ramipril | Ramipril 1.25mg tablets (Kent Pharmaceuticals Ltd) |
| 56169 | 10412711000001103 | Ramipril | Ramipril 10mg capsules (Arrow Generics Ltd) |
| 56356 | 5925111000001106 | Ramipril | Ramipril 10mg capsules (Alliance Healthcare (Distribution) Ltd) |
| 56704 | 5919811000001106 | Ramipril | Ramipril 1.25mg capsules (Alliance Healthcare (Distribution) Ltd) |
| 56763 | 17921311000001109 | Ramipril | Ramipril 10mg capsules (Phoenix Healthcare Distribution Ltd) |
| 56855 | 15182211000001101 | Ramipril | Ramipril 10mg capsules (Sigma Pharmaceuticals Plc) |
| 57073 | 21877711000001106 | Ramipril | Ramipril 1.25mg capsules (Waymade Healthcare Plc) |
| 57235 | 11533411000001106 | Ramipril | Ramipril 1.25mg tablets (Sandoz Ltd) |
| 57346 | 21878311000001108 | Ramipril | Ramipril 10mg capsules (Waymade Healthcare Plc) |
| 57658 | 5881111000001106 | Ramipril | Ramipril 1.25mg tablets (A A H Pharmaceuticals Ltd) |
| 57864 | 15184311000001102 | Ramipril | Ramipril 5mg tablets (Sigma Pharmaceuticals Plc) |
| 59557 | 5632411000001107 | Ramipril | Ramipril 2.5mg capsules (Kent Pharmaceuticals Ltd) |
| 59603 | 17920911000001103 | Ramipril | Ramipril 2.5mg capsules (Phoenix Healthcare Distribution Ltd) |
| 59788 | 16065411000001101 | Ramipril | Ramipril 10mg capsules (Bristol Laboratories Ltd) |
| 60730 | 17921111000001107 | Ramipril | Ramipril 5mg capsules (Phoenix Healthcare Distribution Ltd) |
| 61067 | 11407911000001108 | Ramipril | Ramipril 5mg capsules (Almus Pharmaceuticals Ltd) |
| 61339 | 11408211000001100 | Ramipril | Ramipril 10mg capsules (Almus Pharmaceuticals Ltd) |
| 61499 | 10720911000001101 | Ramipril | Ramipril 2.5mg tablets (Actavis UK Ltd) |
| 61694 | 7339211000001105 | Ramipril | Ramipril 5mg tablets (Zentiva) |

| | | | |
|---|---|---|---|
| 61985 | 5590011000001101 | Ramipril | Ramipril 1.25mg tablets (Teva UK Ltd) |
| 62036 | 21879211000001105 | Ramipril | Ramipril 5mg tablets (Waymade Healthcare Plc) |
| 62039 | 7338811000001104 | Ramipril | Ramipril 1.25mg tablets (Zentiva) |
| 62918 | 8720611000001106 | Ramipril | Ramipril 2.5mg/ 5ml oral solution |
| 62958 | 5591311000001108 | Ramipril | Ramipril 5mg tablets (Teva UK Ltd) |
| 63010 | 17922811000001109 | Ramipril | Ramipril 10mg tablets (Phoenix Healthcare Distribution Ltd) |
| 63442 | 5590811000001107 | Ramipril | Ramipril 2.5mg tablets (Teva UK Ltd) |
| 64055 | 21878511000001102 | Ramipril | Ramipril 2.5mg/ 5ml oral solution sugar free (Waymade Healthcare Plc) |
| 65443 | 147275001000027104 | Ramipril | Ramipril 1.25mg Tablet (Sovereign Medical Ltd) |
| 65599 | 5885211000001102 | Ramipril | Ramipril 5mg tablets (A A H Pharmaceuticals Ltd) |
| 65749 | 28780011000001109 | Ramipril | Ramipril 5mg capsules (Ennogen Pharma Ltd) |
| 65936 | 30087011000001109 | Ramipril | Ramipril 5mg capsules (DE Pharmaceuticals) |
| 66162 | 5591911000001109 | Ramipril | Ramipril 10mg tablets (Teva UK Ltd) |
| 66329 | 244065001000027109 | Ramipril | Ramipril oral solution |
| 66669 | 30087411000001100 | Ramipril | Ramipril 10mg capsules (DE Pharmaceuticals) |
| 67719 | 30866711000001105 | Ramipril | Ramipril 5mg capsules (Mawdsley-Brooks & Company Ltd) |
| 67741 | 9806011000001108 | Ramipril | Ramipril 1.25mg capsules (Almus Pharmaceuticals Ltd) |
| 68192 | 32497211000001100 | Ramipril | Ramipril 5mg capsules (Brown & Burk UK Ltd) |
| 68372 | 10721111000001105 | Ramipril | Ramipril 5mg tablets (Actavis UK Ltd) |
| 68480 | 30867211000001101 | Ramipril | Ramipril 10mg capsules (Mawdsley-Brooks & Company Ltd) |
| 69288 | 32497411000001101 | Ramipril | Ramipril 10mg capsules (Brown & Burk UK Ltd) |
| 70072 | 30865311000001101 | Ramipril | Ramipril 1.25mg capsules (Mawdsley-Brooks & Company Ltd) |
| 70709 | 10721411000001100 | Ramipril | Ramipril 10mg tablets (Actavis UK Ltd) |
| 71025 | 19834511000001104 | Ramipril | Ramipril 1.25mg tablets (APC Pharmaceuticals & Chemicals (Europe) Ltd) |
| 71040 | 20005511000001105 | Ramipril | Ramipril 5mg tablets (Pfizer Ltd) |
| 71068 | 19834711000001109 | Ramipril | Ramipril 2.5mg tablets (APC Pharmaceuticals & Chemicals (Europe) Ltd) |
| 71491 | 28779811000001103 | Ramipril | Ramipril 2.5mg capsules (Ennogen Pharma Ltd) |
| 72341 | 34747411000001106 | Ramipril | Ramipril 2.5mg capsules (Wockhardt UK Ltd) |
| 72842 | 34747811000001108 | Ramipril | Ramipril 10mg capsules (Wockhardt UK Ltd) |
| 73459 | 20007111000001105 | Ramipril | Ramipril 2.5mg/ 5ml oral solution sugar free (A A H Pharmaceuticals Ltd) |
| 73484 | 237045001000027106 | Ramipril | Tritace 5mg Tablet (Sterwin Medicines) |
| 74066 | 13762311000001106 | Ramipril | Ramipril 10mg tablets (Tillomed Laboratories Ltd) |
| 74618 | 30866311000001106 | Ramipril | Ramipril 2.5mg capsules (Mawdsley-Brooks & Company Ltd) |
| 74632 | 147375001000027107 | Ramipril | Ramipril 5mg Tablet (Sovereign Medical Ltd) |
| 75410 | | Ramipril | Ramipril 2.5mg capsules (Brown & Burk UK Ltd) |
| 76548 | | Ramipril | Ramipril 1.25mg capsules (Phoenix Healthcare Distribution Ltd) |
| 76567 | | Ramipril | Ramipril 1.25mg tablets (Sigma Pharmaceuticals Plc) |
| 76784 | | Ramipril | Ramipril 5mg capsules (Wockhardt UK Ltd) |
| 77129 | | Ramipril | Ramipril 10mg capsules (Ennogen Pharma Ltd) |
| 77486 | | Ramipril | Ramipril 2.5mg tablets (Alliance Healthcare (Distribution) Ltd) |
| 77615 | | Ramipril | Ramipril 1.25mg/ 5ml oral solution |
| 3929 | 318886000 | Quinapril | Quinapril 10mg tablets |
| 5159 | 318887009 | Quinapril | Quinapril 20mg tablets |

| | | | |
|---|---|---|---|
| 6765 | 318885001 | Quinapril | Quinapril 5mg tablets |
| 7314 | 582611000001106 | Quinapril | Accupro 5mg tablets (Pfizer Ltd) |
| 9731 | 318894007 | Quinapril | Quinapril 40mg tablets |
| 14477 | 231111000001106 | Quinapril | Accupro 10mg tablets (Pfizer Ltd) |
| 14478 | 829111000001100 | Quinapril | Accupro 20mg tablets (Pfizer Ltd) |
| 15096 | 86411000001109 | Quinapril | Accupro 40mg tablets (Pfizer Ltd) |
| 38854 | 252775001000027101 | Quinapril | Quinapril 20mg/ 5ml oral solution |
| 38899 | 9208111000001100 | Quinapril | Quinil 10mg tablets (Tillomed Laboratories Ltd) |
| 40355 | 9207711000001100 | Quinapril | Quinil 5mg tablets (Tillomed Laboratories Ltd) |
| 42285 | 9208711000001104 | Quinapril | Quinil 40mg tablets (Tillomed Laboratories Ltd) |
| 46365 | 9208411000001105 | Quinapril | Quinil 20mg tablets (Tillomed Laboratories Ltd) |
| 61292 | 7392611000001105 | Quinapril | Quinapril 40mg tablets (Mylan) |
| 77116 | | Quinapril | Quinapril 5mg tablets (Alliance Healthcare (Distribution) Ltd) |
| 56079 | 21939811000001102 | Perindopril tosilate | Perindopril tosilate 10mg tablets |
| 57333 | 21940111000001100 | Perindopril tosilate | Perindopril tosilate 5mg tablets |
| 57944 | 21939911000001107 | Perindopril tosilate | Perindopril tosilate 2.5mg tablets |
| 97 | 318897000 | Perindopril erbumine | Perindopril erbumine 4mg tablets |
| 593 | 318896009 | Perindopril erbumine | Perindopril erbumine 2mg tablets |
| 5612 | 48211000001104 | Perindopril erbumine | Coversyl 2mg tablets (Servier Laboratories Ltd) |
| 5800 | 902211000001100 | Perindopril erbumine | Coversyl 4mg tablets (Servier Laboratories Ltd) |
| 6078 | 374667004 | Perindopril erbumine | Perindopril erbumine 8mg tablets |
| 11983 | 8671311000001103 | Perindopril erbumine | Perindopril erbumine 4mg/ 5ml oral suspension |
| 14960 | 3803711000001101 | Perindopril erbumine | Coversyl 8mg tablets (Servier Laboratories Ltd) |
| 33095 | 10828711000001103 | Perindopril erbumine | Perindopril erbumine 4mg tablets (A A H Pharmaceuticals Ltd) |
| 35731 | 10829111000001106 | Perindopril erbumine | Perindopril erbumine 8mg tablets (A A H Pharmaceuticals Ltd) |
| 38285 | 11879911000001102 | Perindopril erbumine | Perindopril erbumine 4mg tablets (Teva UK Ltd) |
| 38510 | 13767811000001104 | Perindopril erbumine | Perindopril erbumine 4mg tablets (Apotex UK Ltd) |
| 43012 | 244345001000027102 | Perindopril Erbumine | Perindopril erbumine oral solution |
| 43813 | 12498511000001100 | Perindopril erbumine | Perindopril erbumine 2mg tablets (Actavis UK Ltd) |
| 45319 | 10828511000001108 | Perindopril erbumine | Perindopril erbumine 2mg tablets (A A H Pharmaceuticals Ltd) |
| 45938 | 11880111000001100 | Perindopril erbumine | Perindopril erbumine 8mg tablets (Teva UK Ltd) |
| 48049 | 16631411000001104 | Perindopril erbumine | Perindopril erbumine 2mg tablets (Mylan) |
| 48180 | 13652011000001107 | Perindopril erbumine | Perindopril erbumine 4mg tablets (Sandoz Ltd) |
| 48214 | 12498711000001105 | Perindopril erbumine | Perindopril erbumine 4mg tablets (Actavis UK Ltd) |
| 49491 | 14126011000001109 | Perindopril erbumine | Perindopril erbumine 2mg tablets (Consilient Health Ltd) |
| 50402 | 193395001000027104 | Perindopril erbumine | Perindopril 2mg Tablet (Servier Laboratories Ltd) |
| 53058 | 20319211000001109 | Perindopril erbumine | Perindopril erbumine 8mg tablets (Sandoz Ltd) |
| 54733 | 17218111000001108 | Perindopril erbumine | Perindopril erbumine 8mg tablets (Consilient Health Ltd) |
| 54899 | 11879711000001104 | Perindopril erbumine | Perindopril erbumine 2mg tablets (Teva UK Ltd) |
| 54942 | 16631011000001108 | Perindopril erbumine | Perindopril erbumine 8mg tablets (Mylan) |
| 54986 | 14057411000001109 | Perindopril erbumine | Perindopril erbumine 8mg/ 5ml oral suspension |
| 56162 | 14126311000001107 | Perindopril erbumine | Perindopril erbumine 4mg tablets (Consilient Health Ltd) |
| 56472 | 10743211000001103 | Perindopril erbumine | Perindopril erbumine 4mg tablets (Kent Pharmaceuticals Ltd) |

| | | | |
|---|---|---|---|
| 56473 | 15156311000001100 | Perindopril erbumine | Perindopril erbumine 2mg tablets (Sigma Pharmaceuticals Plc) |
| 56506 | 5535211000001106 | Perindopril erbumine | Coversyl 2mg tablets (Dowelhurst Ltd) |
| 56508 | 5538211000001104 | Perindopril erbumine | Coversyl 4mg tablets (Dowelhurst Ltd) |
| 56516 | 13651711000001102 | Perindopril erbumine | Perindopril erbumine 2mg tablets (Sandoz Ltd) |
| 57701 | 12498911000001107 | Perindopril erbumine | Perindopril erbumine 8mg tablets (Actavis UK Ltd) |
| 57801 | 15639211000001104 | Perindopril erbumine | Perindopril erbumine 4mg tablets (Glenmark Pharmaceuticals Europe Ltd) |
| 58843 | 10742711000001102 | Perindopril erbumine | Perindopril erbumine 2mg tablets (Kent Pharmaceuticals Ltd) |
| 58874 | 18426311000001107 | Perindopril erbumine | Perindopril erbumine 2mg tablets (Somex Pharma) |
| 59770 | 22378411000001100 | Perindopril erbumine | Perindopril erbumine 4mg tablets (Aurobindo Pharma Ltd) |
| 59790 | 20168911000001102 | Perindopril erbumine | Perindopril erbumine 8mg tablets (Accord Healthcare Ltd) |
| 59972 | 12060711000001101 | Perindopril erbumine | Perindopril erbumine 2mg tablets (Alliance Healthcare (Distribution) Ltd) |
| 60065 | 15156711000001101 | Perindopril erbumine | Perindopril erbumine 4mg tablets (Sigma Pharmaceuticals Plc) |
| 61117 | 23471511000001106 | Perindopril erbumine | Perindopril erbumine 4mg/ 5ml oral solution |
| 61270 | 21296211000001103 | Perindopril erbumine | Perindopril erbumine 4mg tablets (Accord Healthcare Ltd) |
| 61693 | 22378211000001104 | Perindopril erbumine | Perindopril erbumine 8mg tablets (Aurobindo Pharma Ltd) |
| 64602 | 21845111000001102 | Perindopril erbumine | Perindopril erbumine 2mg tablets (Waymade Healthcare Plc) |
| 65273 | 16631211000001103 | Perindopril erbumine | Perindopril erbumine 4mg tablets (Mylan) |
| 66060 | 30075311000001109 | Perindopril erbumine | Perindopril erbumine 2mg tablets (DE Pharmaceuticals) |
| 67269 | 5352011000001109 | Perindopril erbumine | Coversyl 2mg tablets (Waymade Healthcare Plc) |
| 67789 | 21295411000001108 | Perindopril erbumine | Perindopril erbumine 2mg tablets (Accord Healthcare Ltd) |
| 68021 | 12061211000001102 | Perindopril erbumine | Perindopril erbumine 4mg tablets (Alliance Healthcare (Distribution) Ltd) |
| 68381 | 30856611000001105 | Perindopril erbumine | Perindopril erbumine 4mg tablets (Mawdsley-Brooks & Company Ltd) |
| 68759 | 22378611000001102 | Perindopril erbumine | Perindopril erbumine 2mg tablets (Aurobindo Pharma Ltd) |
| 69016 | 30075711000001108 | Perindopril erbumine | Perindopril erbumine 8mg tablets (DE Pharmaceuticals) |
| 70916 | 15639611000001102 | Perindopril erbumine | Perindopril erbumine 8mg tablets (Glenmark Pharmaceuticals Europe Ltd) |
| 70917 | 15639011000001109 | Perindopril erbumine | Perindopril erbumine 2mg tablets (Glenmark Pharmaceuticals Europe Ltd) |
| 71004 | 21296511000001100 | Perindopril erbumine | Perindopril erbumine 8mg tablets (Accord Healthcare Ltd) |
| 72295 | 196145001000027107 | Perindopril erbumine | Perindopril 2mg Tablet (Neo Laboratories Ltd) |
| 72941 | 13011311000001104 | Perindopril erbumine | Perindopril erbumine 1mg/ 5ml oral suspension |
| 75021 | 16213911000001100 | Perindopril erbumine | Perindopril erbumine 4mg tablets (Ranbaxy (UK) Ltd) |
| 75024 | 17955711000001106 | Perindopril erbumine | Perindopril erbumine 4mg tablets (Phoenix Healthcare Distribution Ltd) |
| 75847 | | Perindopril erbumine | Perindopril erbumine 8mg/ 5ml oral solution |
| 77665 | | Perindopril erbumine | Perindopril erbumine 8mg tablets (Somex Pharma) |
| 37930 | 13454411000001108 | Perindopril arginine | Perindopril arginine 5mg tablets |
| 37964 | 13454211000001109 | Perindopril arginine | Perindopril arginine 2.5mg tablets |
| 37965 | 13444911000001103 | Perindopril arginine | Coversyl Arginine 5mg tablets (Servier Laboratories Ltd) |
| 37971 | 13454111000001103 | Perindopril arginine | Perindopril arginine 10mg tablets |
| 38026 | 13444611000001109 | Perindopril arginine | Coversyl Arginine 10mg tablets (Servier Laboratories Ltd) |
| 38034 | 13445211000001108 | Perindopril arginine | Coversyl Arginine 2.5mg tablets (Servier Laboratories Ltd) |
| 50347 | 15416811000001102 | Perindopril arginine | Coversyl Arginine 5mg tablets (Waymade Healthcare Plc) |
| 51807 | 19720911000001102 | Perindopril arginine | Coversyl Arginine 5mg tablets (DE Pharmaceuticals) |
| 15121 | 318934008 | Moexipril | Moexipril 7.5mg tablets |
| 17120 | 318935009 | Moexipril | Moexipril 15mg tablets |

| | | | |
|---|---|---|---|
| 28724 | 4041111000001109 | Moexipril | Perdix 7.5mg tablets (UCB Pharma Ltd) |
| 28725 | 4040311000001101 | Moexipril | Perdix 15mg tablets (UCB Pharma Ltd) |
| 65 | 318859000 | Lisinopril | Lisinopril 10mg tablets |
| 69 | 318860005 | Lisinopril | Lisinopril 20mg tablets |
| 78 | 318858008 | Lisinopril | Lisinopril 5mg tablets |
| 277 | 318857003 | Lisinopril | Lisinopril 2.5mg tablets |
| 3720 | 593111000001108 | Lisinopril | Zestril 2.5mg tablets (AstraZeneca UK Ltd) |
| 6806 | 825311000001100 | Lisinopril | Zestril 10mg tablets (AstraZeneca UK Ltd) |
| 6807 | 823211000001109 | Lisinopril | Zestril 5mg tablets (AstraZeneca UK Ltd) |
| 8268 | 891711000001107 | Lisinopril | Zestril 20mg tablets (AstraZeneca UK Ltd) |
| 10882 | 321011000001103 | Lisinopril | Carace 2.5mg tablets (Bristol-Myers Squibb Pharmaceuticals Ltd) |
| 11987 | 8622511000001100 | Lisinopril | Lisinopril 5mg/ 5ml oral solution |
| 12313 | 56711000001109 | Lisinopril | Carace 20mg tablets (Bristol-Myers Squibb Pharmaceuticals Ltd) |
| 14387 | 778011000001109 | Lisinopril | Carace 5mg tablets (Bristol-Myers Squibb Pharmaceuticals Ltd) |
| 16701 | 315211000001104 | Lisinopril | Carace 10mg tablets (Bristol-Myers Squibb Pharmaceuticals Ltd) |
| 19198 | 487411000001102 | Lisinopril | Lisinopril 20mg tablets (Teva UK Ltd) |
| 19204 | 528611000001100 | Lisinopril | Lisinopril 5mg tablets (Teva UK Ltd) |
| 19223 | 640511000001107 | Lisinopril | Lisinopril 10mg tablets (Teva UK Ltd) |
| 20975 | 8622911000001107 | Lisinopril | Lisinopril 7.5mg/ 5ml oral suspension |
| 30921 | 60211000001105 | Lisinopril | Lisinopril 2.5mg tablets (Teva UK Ltd) |
| 32597 | 145811000001100 | Lisinopril | Lisinopril 10mg tablets (Sandoz Ltd) |
| 33977 | 656411000001108 | Lisinopril | Lisinopril 10mg tablets (Mylan) |
| 34471 | 35611000001109 | Lisinopril | Lisinopril 5mg tablets (Mylan) |
| 34696 | 576511000001101 | Lisinopril | Lisinopril 20mg tablets (Mylan) |
| 34799 | 360711000001100 | Lisinopril | Lisinopril 20mg tablets (Zentiva) |
| 37778 | 8622611000001101 | Lisinopril | Lisinopril 5mg/ 5ml oral suspension |
| 41522 | 7315011000001107 | Lisinopril | Lisopress 20mg tablets (Teva UK Ltd) |
| 41532 | 7314511000001100 | Lisinopril | Lisopress 5mg tablets (Teva UK Ltd) |
| 41538 | 7314011000001108 | Lisinopril | Lisopress 2.5mg tablets (Teva UK Ltd) |
| 41573 | 7314811000001102 | Lisinopril | Lisopress 10mg tablets (Teva UK Ltd) |
| 43412 | 874311000001100 | Lisinopril | Lisinopril 2.5mg tablets (A A H Pharmaceuticals Ltd) |
| 43413 | 668811000001102 | Lisinopril | Lisinopril 20mg tablets (A A H Pharmaceuticals Ltd) |
| 43416 | 532711000001109 | Lisinopril | Lisinopril 10mg tablets (A A H Pharmaceuticals Ltd) |
| 43418 | 18111000001103 | Lisinopril | Lisinopril 5mg tablets (A A H Pharmaceuticals Ltd) |
| 43566 | 877611000001104 | Lisinopril | Lisinopril 2.5mg tablets (Sandoz Ltd) |
| 45300 | 247511000001109 | Lisinopril | Lisinopril 10mg tablets (Actavis UK Ltd) |
| 45324 | 460111000001109 | Lisinopril | Lisinopril 20mg tablets (Actavis UK Ltd) |
| 45337 | 34511000001109 | Lisinopril | Lisinopril 5mg tablets (Actavis UK Ltd) |
| 45816 | 9810211000001103 | Lisinopril | Lisinopril 5mg tablets (Almus Pharmaceuticals Ltd) |
| 46975 | 683211000001108 | Lisinopril | Lisinopril 5mg tablets (Sandoz Ltd) |
| 46979 | 637111000001101 | Lisinopril | Lisinopril 20mg tablets (Sandoz Ltd) |
| 47159 | 9810411000001104 | Lisinopril | Lisinopril 10mg tablets (Almus Pharmaceuticals Ltd) |
| 51433 | 13754411000001103 | Lisinopril | Lisinopril 20mg tablets (Tillomed Laboratories Ltd) |

| | | | |
|---|---|---|---|
| 52088 | 17937211000001108 | Lisinopril | Lisinopril 5mg tablets (Phoenix Healthcare Distribution Ltd) |
| 53271 | 912911000001107 | Lisinopril | Lisinopril 10mg tablets (Alliance Healthcare (Distribution) Ltd) |
| 53551 | 17937611000001105 | Lisinopril | Lisinopril 20mg tablets (Phoenix Healthcare Distribution Ltd) |
| 53820 | 10386011000001109 | Lisinopril | Lisinopril 5mg tablets (Arrow Generics Ltd) |
| 54037 | 10290011000001104 | Lisinopril | Lisinopril 10mg tablets (Relonchem Ltd) |
| 54283 | 8594211000001106 | Lisinopril | Lisinopril 5mg/ 5ml oral suspension (Special Order) |
| 54288 | 10386411000001100 | Lisinopril | Lisinopril 10mg tablets (Arrow Generics Ltd) |
| 54512 | 243735001000027100 | Lisinopril | Lisinopril Oral solution |
| 54928 | 16060611000001106 | Lisinopril | Lisinopril 10mg tablets (Bristol Laboratories Ltd) |
| 55002 | 18463211000001101 | Lisinopril | Lisinopril 20mg tablets (Accord Healthcare Ltd) |
| 55456 | 72111000001106 | Lisinopril | Lisinopril 5mg tablets (Alliance Healthcare (Distribution) Ltd) |
| 55588 | 15107811000001101 | Lisinopril | Lisinopril 20mg tablets (Sigma Pharmaceuticals Plc) |
| 55639 | 18463011000001106 | Lisinopril | Lisinopril 10mg tablets (Accord Healthcare Ltd) |
| 55896 | 460011000001108 | Lisinopril | Lisinopril 2.5mg tablets (Actavis UK Ltd) |
| 56279 | 8622111000001109 | Lisinopril | Lisinopril 2.5mg/ 5ml oral solution |
| 56505 | 16450011000001100 | Lisinopril | Zestril 5mg tablets (Lexon (UK) Ltd) |
| 56510 | 14767811000001102 | Lisinopril | Zestril 20mg tablets (Sigma Pharmaceuticals Plc) |
| 57048 | 499711000001101 | Lisinopril | Lisinopril 10mg tablets (Zentiva) |
| 57588 | 17480011000001107 | Lisinopril | Zestril 2.5mg tablets (Mawdsley-Brooks & Company Ltd) |
| 58258 | 8622211000001103 | Lisinopril | Lisinopril 2.5mg/ 5ml oral suspension |
| 58294 | 18463411000001102 | Lisinopril | Lisinopril 5mg tablets (Accord Healthcare Ltd) |
| 58451 | 9809811000001108 | Lisinopril | Lisinopril 2.5mg tablets (Almus Pharmaceuticals Ltd) |
| 58461 | 709811000001102 | Lisinopril | Lisinopril 2.5mg tablets (Kent Pharmaceuticals Ltd) |
| 58682 | 5253711000001106 | Lisinopril | Lisinopril 2.5mg tablets (Mylan) |
| 58863 | 17937411000001107 | Lisinopril | Lisinopril 10mg tablets (Phoenix Healthcare Distribution Ltd) |
| 58871 | 22072611000001100 | Lisinopril | Lisinopril 10mg tablets (Waymade Healthcare Plc) |
| 59109 | 13754011000001107 | Lisinopril | Lisinopril 5mg tablets (Tillomed Laboratories Ltd) |
| 59111 | 580411000001107 | Lisinopril | Lisinopril 20mg tablets (Alliance Healthcare (Distribution) Ltd) |
| 60010 | 832711000001108 | Lisinopril | Lisinopril 10mg tablets (Kent Pharmaceuticals Ltd) |
| 60097 | 461611000001106 | Lisinopril | Lisinopril 2.5mg tablets (Zentiva) |
| 60232 | 574111000001103 | Lisinopril | Lisinopril 5mg tablets (Zentiva) |
| 60309 | 10291711000001105 | Lisinopril | Lisinopril 5mg tablets (Relonchem Ltd) |
| 61262 | 16060811000001105 | Lisinopril | Lisinopril 20mg tablets (Bristol Laboratories Ltd) |
| 62564 | 8621911000001101 | Lisinopril | Lisinopril 10mg/ 5ml oral solution |
| 63030 | 24367411000001102 | Lisinopril | Lisinopril 10mg tablets (DE Pharmaceuticals) |
| 63559 | 607011000001108 | Lisinopril | Lisinopril 20mg tablets (Kent Pharmaceuticals Ltd) |
| 63824 | 8622011000001108 | Lisinopril | Lisinopril 10mg/ 5ml oral suspension |
| 64902 | 30251811000001106 | Lisinopril | Lisinopril 5mg/ 5ml oral solution sugar free |
| 65102 | 15107411000001103 | Lisinopril | Lisinopril 10mg tablets (Sigma Pharmaceuticals Plc) |
| 65416 | 27377411000001101 | Lisinopril | Lisinopril 5mg tablets (Lupin Healthcare (UK) Ltd) |
| 65536 | 337211000001101 | Lisinopril | Lisinopril 2.5mg tablets (Alliance Healthcare (Distribution) Ltd) |
| 65983 | 27377211000001100 | Lisinopril | Lisinopril 2.5mg tablets (Lupin Healthcare (UK) Ltd) |
| 65985 | 24368011000001107 | Lisinopril | Lisinopril 2.5mg tablets (DE Pharmaceuticals) |

| | | | |
|---|---|---|---|
| 66558 | 24368811000001101 | Lisinopril | Lisinopril 5mg tablets (DE Pharmaceuticals) |
| 66622 | 24368611000001100 | Lisinopril | Lisinopril 20mg tablets (DE Pharmaceuticals) |
| 66772 | 22072211000001102 | Lisinopril | Lisinopril 2.5mg tablets (Waymade Healthcare Plc) |
| 67075 | 30221411000001105 | Lisinopril | Lisinopril 2.5mg tablets (Mawdsley-Brooks & Company Ltd) |
| 67194 | 16060211000001109 | Lisinopril | Lisinopril 2.5mg tablets (Bristol Laboratories Ltd) |
| 67795 | 9810811000001102 | Lisinopril | Lisinopril 20mg tablets (Almus Pharmaceuticals Ltd) |
| 68094 | 15108011000001108 | Lisinopril | Lisinopril 5mg tablets (Sigma Pharmaceuticals Plc) |
| 68247 | 16060411000001108 | Lisinopril | Lisinopril 5mg tablets (Bristol Laboratories Ltd) |
| 69074 | 10291011000001108 | Lisinopril | Lisinopril 20mg tablets (Relonchem Ltd) |
| 69269 | 22072811000001101 | Lisinopril | Lisinopril 20mg tablets (Waymade Healthcare Plc) |
| 70667 | 30221611000001108 | Lisinopril | Lisinopril 5mg tablets (Mawdsley-Brooks & Company Ltd) |
| 71562 | 129245001000027105 | Lisinopril | Lisinopril 10mg Tablet (Niche Generics Ltd) |
| 72038 | 10290411000001108 | Lisinopril | Lisinopril 2.5mg tablets (Relonchem Ltd) |
| 72336 | 22072411000001103 | Lisinopril | Lisinopril 5mg tablets (Waymade Healthcare Plc) |
| 73672 | 35165411000001106 | Lisinopril | Lisinopril 10mg tablets (Aurobindo Pharma Ltd) |
| 73716 | 35391311000001107 | Lisinopril | Lisinopril 2.5mg tablets (Crescent Pharma Ltd) |
| 74155 | 35165211000001107 | Lisinopril | Lisinopril 5mg tablets (Aurobindo Pharma Ltd) |
| 76100 | | Lisinopril | Lisinopril 5mg tablets (Kent Pharmaceuticals Ltd) |
| 76128 | | Lisinopril | Lisinopril 20mg tablets (Lupin Healthcare (UK) Ltd) |
| 77378 | | Lisinopril | Zestril 10mg tablets (Dowelhurst Ltd) |
| 77401 | | Lisinopril | Zestril 2.5mg tablets (Waymade Healthcare Plc) |
| 77402 | | Lisinopril | Zestril 5mg tablets (Dowelhurst Ltd) |
| 77407 | | Lisinopril | Zestril 5mg tablets (Waymade Healthcare Plc) |
| 6408 | 797911000001103 | Imidapril | Tanatril 5mg tablets (Mitsubishi Tanabe Pharma Europe Ltd) |
| 12815 | 680811000001102 | Imidapril | Tanatril 10mg tablets (Mitsubishi Tanabe Pharma Europe Ltd) |
| 12858 | 318944005 | Imidapril | Imidapril 10mg tablets |
| 16924 | 318943004 | Imidapril | Imidapril 5mg tablets |
| 18219 | 318942009 | Imidapril | Imidapril 20mg tablets |
| 32560 | 533511000001106 | Imidapril | Tanatril 20mg tablets (Mitsubishi Tanabe Pharma Europe Ltd) |
| 633 | 318909008 | Fosinopril sodium | Fosinopril 10mg tablets |
| 4571 | 462511000001104 | Fosinopril sodium | Staril 10mg tablets (Bristol-Myers Squibb Pharmaceuticals Ltd) |
| 5861 | 318910003 | Fosinopril sodium | Fosinopril 20mg tablets |
| 13589 | 348111000001107 | Fosinopril sodium | Staril 20mg tablets (Bristol-Myers Squibb Pharmaceuticals Ltd) |
| 67307 | 5543711000001104 | Fosinopril sodium | Staril 20mg tablets (Dowelhurst Ltd) |
| 196 | 318851002 | Enalapril maleate | Enalapril 5mg tablets |
| 448 | 318850001 | Enalapril maleate | Enalapril 2.5mg tablets |
| 1299 | 318853004 | Enalapril maleate | Enalapril 10mg tablets |
| 1904 | 318855006 | Enalapril maleate | Enalapril 20mg tablets |
| 8105 | 302311000001104 | Enalapril maleate | Innovace 20mg tablets (Merck Sharp & Dohme Ltd) |
| 8106 | 749611000001107 | Enalapril maleate | Innovace 2.5mg tablets (Merck Sharp & Dohme Ltd) |
| 8800 | 730211000001104 | Enalapril maleate | Innovace 5mg tablets (Merck Sharp & Dohme Ltd) |
| 8830 | 316111000001104 | Enalapril maleate | Innovace 10mg tablets (Merck Sharp & Dohme Ltd) |
| 11197 | 233035001000027101 | Enalapril Maleate | Innovace melt 5mg Wafer (Merck Sharp & Dohme Ltd) |

| | | | |
|---|---|---|---|
| 13755 | 206205001000027109 | Enalapril Maleate | Enalapril 10mg wafer |
| 15085 | 82345001000027107 | Enalapril Maleate | Innovace Titration pack (Merck Sharp & Dohme Ltd) |
| 16708 | 98645001000027107 | Enalapril Maleate | Enalapril titration pack |
| 19208 | 676011000001102 | Enalapril maleate | Enalapril 10mg tablets (Actavis UK Ltd) |
| 20188 | 98655001000027105 | Enalapril Maleate | Enalapril 2.5mg wafer |
| 22439 | 222975001000027100 | Enalapril maleate | Ednyt 20mg Tablet (Dominion Pharma) |
| 22708 | 206195001000027109 | Enalapril Maleate | Enalapril 5mg wafer |
| 23252 | 761411000001101 | Enalapril maleate | Pralenal 10 tablets (Opus Pharmaceuticals Ltd) |
| 24041 | 206215001000027106 | Enalapril Maleate | Enalapril 20mg wafer |
| 27871 | 233045001000027100 | Enalapril Maleate | Innovace melt 10mg Wafer (Merck Sharp & Dohme Ltd) |
| 28127 | 795111000001105 | Enalapril maleate | Enalapril 2.5mg tablets (Teva UK Ltd) |
| 29530 | 233025001000027103 | Enalapril Maleate | Innovace melt 2.5mg Wafer (Merck Sharp & Dohme Ltd) |
| 31587 | 233005001000027105 | Enalapril Maleate | Innovace melt 20mg Wafer (Merck Sharp & Dohme Ltd) |
| 31716 | 665211000001105 | Enalapril maleate | Enalapril 20mg tablets (Actavis UK Ltd) |
| 32241 | 225811000001104 | Enalapril maleate | Enalapril 10mg tablets (A A H Pharmaceuticals Ltd) |
| 33057 | 195715001000027106 | Enalapril maleate | Ednyt 5mg Tablet (Dominion Pharma) |
| 33078 | 526511000001100 | Enalapril maleate | Enalapril 20mg tablets (A A H Pharmaceuticals Ltd) |
| 34400 | 61665001000027109 | Enalapril maleate | Enalapril 5mg Tablet (Dowelhurst Ltd) |
| 34453 | 923711000001107 | Enalapril maleate | Enalapril 20mg tablets (Mylan) |
| 34712 | 233511000001103 | Enalapril maleate | Enalapril 20mg tablets (Kent Pharmaceuticals Ltd) |
| 34768 | 381411000001100 | Enalapril maleate | Enalapril 20mg tablets (IVAX Pharmaceuticals UK Ltd) |
| 34798 | 728411000001104 | Enalapril maleate | Enalapril 20mg tablets (Sandoz Ltd) |
| 34952 | 401711000001103 | Enalapril maleate | Enalapril 10mg tablets (Mylan) |
| 34953 | 346311000001105 | Enalapril maleate | Enalapril 20mg tablets (Zentiva) |
| 35794 | 26111000001109 | Enalapril maleate | Enalapril 5mg tablets (A A H Pharmaceuticals Ltd) |
| 36753 | 195725001000027102 | Enalapril maleate | Ednyt 10mg Tablet (Dominion Pharma) |
| 37080 | 8486911000001103 | Enalapril maleate | Enalapril 5mg/ 5ml oral solution |
| 37087 | 8487011000001104 | Enalapril maleate | Enalapril 5mg/ 5ml oral suspension |
| 41417 | 4811000001104 | Enalapril maleate | Enalapril 2.5mg tablets (A A H Pharmaceuticals Ltd) |
| 41694 | 456011000001101 | Enalapril maleate | Enalapril 2.5mg tablets (IVAX Pharmaceuticals UK Ltd) |
| 41746 | 450811000001105 | Enalapril maleate | Enalapril 10mg tablets (Sandoz Ltd) |
| 42723 | 578511000001100 | Enalapril maleate | Pralenal 5 tablets (Opus Pharmaceuticals Ltd) |
| 42894 | 115211000001105 | Enalapril maleate | Enalapril 10mg tablets (Teva UK Ltd) |
| 42901 | 319411000001102 | Enalapril maleate | Enalapril 5mg tablets (Teva UK Ltd) |
| 42902 | 695511000001106 | Enalapril maleate | Enalapril 20mg tablets (Teva UK Ltd) |
| 42908 | 856011000001106 | Enalapril maleate | Enalapril 5mg tablets (IVAX Pharmaceuticals UK Ltd) |
| 43411 | 51511000001108 | Enalapril maleate | Enalapril 5mg tablets (Sandoz Ltd) |
| 43563 | 102611000001100 | Enalapril maleate | Enalapril 2.5mg tablets (Zentiva) |
| 44657 | 195705001000027109 | Enalapril maleate | Ednyt 2.5mg Tablet (Dominion Pharma) |
| 45217 | 50211000001104 | Enalapril maleate | Enalapril 5mg tablets (Kent Pharmaceuticals Ltd) |
| 46974 | 766411000001107 | Enalapril maleate | Enalapril 5mg tablets (Mylan) |
| 50334 | 8486811000001108 | Enalapril maleate | Enalapril 4mg/ 5ml oral suspension |
| 50780 | 12086111000001102 | Enalapril maleate | Enalapril 2mg/ 5ml oral solution |

| | | | |
|---|---|---|---|
| 50863 | 8463911000001106 | Enalapril maleate | Enalapril 5mg/ 5ml oral solution (Drug Tariff Special Order) |
| 52010 | 688811000001104 | Enalapril maleate | Enalapril 10mg tablets (Alliance Healthcare (Distribution) Ltd) |
| 52882 | 20092911000001108 | Enalapril maleate | Enalapril 5mg/ 5ml oral suspension sugar free |
| 53719 | 367611000001108 | Enalapril maleate | Enalapril 20mg tablets (Alliance Healthcare (Distribution) Ltd) |
| 53915 | 11553911000001104 | Enalapril maleate | Enalapril 5mg tablets (Dexcel-Pharma Ltd) |
| 55903 | 11554111000001100 | Enalapril maleate | Enalapril 10mg tablets (Dexcel-Pharma Ltd) |
| 57378 | 12086211000001108 | Enalapril maleate | Enalapril 2mg/ 5ml oral suspension |
| 57882 | 8489611000001101 | Enalapril maleate | Enalapril 2.5mg/ 5ml oral suspension |
| 58751 | 8485611000001104 | Enalapril maleate | Enalapril 1.25mg/ 5ml oral suspension |
| 59996 | 19191211000001101 | Enalapril maleate | Enalapril 20mg tablets (Milpharm Ltd) |
| 60143 | 19727011000001100 | Enalapril maleate | Enalapril 5mg tablets (Medreich Plc) |
| 61133 | 17901611000001106 | Enalapril maleate | Enalapril 10mg tablets (Phoenix Healthcare Distribution Ltd) |
| 62860 | 23881911000001104 | Enalapril maleate | Enalapril 5mg tablets (DE Pharmaceuticals) |
| 63322 | 9794411000001107 | Enalapril maleate | Enalapril 10mg tablets (Almus Pharmaceuticals Ltd) |
| 64062 | 12085811000001101 | Enalapril maleate | Enalapril 1mg/ 5ml oral suspension |
| 64877 | 11553711000001101 | Enalapril maleate | Enalapril 2.5mg tablets (Dexcel-Pharma Ltd) |
| 66895 | 8485811000001100 | Enalapril maleate | Enalapril 10mg/ 5ml oral suspension |
| 68496 | 127311000001103 | Enalapril maleate | Enalapril 10mg tablets (Kent Pharmaceuticals Ltd) |
| 71668 | 9794611000001105 | Enalapril maleate | Enalapril 20mg tablets (Almus Pharmaceuticals Ltd) |
| 71737 | 9793911000001101 | Enalapril maleate | Enalapril 2.5mg tablets (Almus Pharmaceuticals Ltd) |
| 72017 | 8485511000001103 | Enalapril maleate | Enalapril 1.25mg/ 5ml oral solution |
| 73389 | 23881711000001101 | Enalapril maleate | Enalapril 2.5mg tablets (DE Pharmaceuticals) |
| 73617 | 12085411000001103 | Enalapril maleate | Enalapril 1.5mg/ 5ml oral suspension |
| 74237 | 12085911000001106 | Enalapril maleate | Enalapril 25mg/ 5ml oral solution |
| 76486 | | Enalapril maleate | Enalapril 2.5mg tablets (Mylan) |
| 76619 | | Enalapril maleate | Enalapril 20mg tablets (Dexcel-Pharma Ltd) |
| 77271 | | Enalapril maleate | Enalapril 20mg Tablet (Neo Laboratories Ltd) |
| 77315 | | Enalapril maleate | Enalapril 2.5mg/ 5ml oral solution |
| 77316 | | Enalapril maleate | Enalapril 4mg/ 5ml oral solution |
| 77345 | | Enalapril maleate | Innovace 10mg tablets (Dowelhurst Ltd) |
| 77690 | | Enalapril maleate | Enalapril 10mg tablets (IVAX Pharmaceuticals UK Ltd) |
| 77831 | | Enalapril maleate | Enalapril 500micrograms/ 5ml oral solution |
| 12411 | 318915008 | Cilazapril monohydrate | Cilazapril 500microgram tablets |
| 12412 | 318917000 | Cilazapril monohydrate | Cilazapril 2.5mg tablets |
| 12574 | 318916009 | Cilazapril monohydrate | Cilazapril 1mg tablets |
| 13026 | 318923005 | Cilazapril monohydrate | Cilazapril 5mg tablets |
| 16196 | 3671311000001104 | Cilazapril monohydrate | Vascace 5mg tablets (Roche Products Ltd) |
| 16197 | 3672411000001101 | Cilazapril monohydrate | Vascace 2.5mg tablets (Roche Products Ltd) |
| 16212 | 3669711000001105 | Cilazapril monohydrate | Vascace 1mg tablets (Roche Products Ltd) |
| 21053 | 3740211000001108 | Cilazapril monohydrate | Vascace 500microgram tablets (Roche Products Ltd) |
| 15605 | 11055001000027102 | Cilazapril | Cilazapril 250micrograms tablets |
| 23642 | 20325001000027107 | Cilazapril | Vascace 0.25mg Tablet (Roche Products Ltd) |
| 1121 | 318820009 | Captopril | Captopril 12.5mg tablets |

| | | | |
|---|---|---|---|
| 1143 | 318821008 | Captopril | Captopril 25mg tablets |
| 1144 | 455611000001103 | Captopril | Capoten 25mg tablets (Bristol-Myers Squibb Pharmaceuticals Ltd) |
| 1807 | 318824000 | Captopril | Captopril 50mg tablets |
| 3069 | 517711000001103 | Captopril | Acepril 25mg tablets (Bristol-Myers Squibb Pharmaceuticals Ltd) |
| 3310 | 134511000001103 | Captopril | Capoten 12.5mg tablets (Bristol-Myers Squibb Pharmaceuticals Ltd) |
| 3839 | 386611000001109 | Captopril | Capoten 50mg tablets (Bristol-Myers Squibb Pharmaceuticals Ltd) |
| 15958 | 219545001000027103 | Captopril | Captopril 2mg tablets |
| 17624 | 8351811000001102 | Captopril | Captopril 5mg/ 5ml oral suspension |
| 17633 | 236435001000027104 | Captopril | Captopril 3mg/ 5ml oral solution |
| 18269 | 660111000001107 | Captopril | Acepril 12.5mg tablets (Bristol-Myers Squibb Pharmaceuticals Ltd) |
| 18325 | 814111000001108 | Captopril | Acepril 50mg tablets (Bristol-Myers Squibb Pharmaceuticals Ltd) |
| 20849 | 673711000001107 | Captopril | Tensopril 12.5mg tablets (Teva UK Ltd) |
| 21943 | 777711000001105 | Captopril | Kaplon 12.5mg tablets (Teva UK Ltd) |
| 23478 | 653111000001108 | Captopril | Tensopril 50mg tablets (Teva UK Ltd) |
| 24482 | 760211000001106 | Captopril | Captomex 50mg tablets (Actavis UK Ltd) |
| 25998 | 372511000001104 | Captopril | Captomex 12.5mg tablets (Actavis UK Ltd) |
| 26995 | 477111000001100 | Captopril | Kaplon 25mg tablets (Teva UK Ltd) |
| 28486 | 8352011000001100 | Captopril | Captopril 6.25mg/ 5ml oral suspension |
| 28820 | 562011000001101 | Captopril | Captomex 25mg tablets (Actavis UK Ltd) |
| 30039 | 597511000001109 | Captopril | Tensopril 25mg tablets (Teva UK Ltd) |
| 32048 | 572511000001102 | Captopril | Kaplon 50mg tablets (Teva UK Ltd) |
| 32514 | 817411000001108 | Captopril | Ecopace 25mg tablets (AMCo) |
| 33336 | 160135001000027103 | Captopril | Captopril 5mg/ 5ml Oral suspension (Eldon Laboratories) |
| 33646 | 68395001000027109 | Captopril | Captopril 12.5mg Tablet (Generics (UK) Ltd) |
| 34544 | 68435001000027105 | Captopril | Captopril 12.5mg Tablet (IVAX Pharmaceuticals UK Ltd) |
| 34562 | 68445001000027106 | Captopril | Captopril 25mg Tablet (IVAX Pharmaceuticals UK Ltd) |
| 34719 | 68415001000027104 | Captopril | Captopril 50mg Tablet (Generics (UK) Ltd) |
| 34936 | 68705001000027105 | Captopril | Captopril 25mg Tablet (Lagap) |
| 34937 | 68455001000027109 | Captopril | Captopril 50mg Tablet (IVAX Pharmaceuticals UK Ltd) |
| 35302 | 8347211000001105 | Captopril | Captopril 12.5mg/ 5ml oral suspension |
| 36742 | 248405001000027101 | Captopril | Captopril 2mg/ 5ml oral suspension |
| 37655 | 424411000001101 | Captopril | Captopril 25mg tablets (Teva UK Ltd) |
| 39512 | 256275001000027106 | Captopril | Captopril 25mg/ 5ml oral suspension |
| 41617 | 501411000001104 | Captopril | Captopril 25mg tablets (Actavis UK Ltd) |
| 41633 | 605411000001105 | Captopril | Captopril 12.5mg tablets (Actavis UK Ltd) |
| 41743 | 3911000001107 | Captopril | Captopril 50mg tablets (Teva UK Ltd) |
| 43432 | 245485001000027106 | Captopril | Captopril 6.25mg tablets |
| 43507 | 68405001000027101 | Captopril | Captopril 25mg Tablet (Generics (UK) Ltd) |
| 43649 | 303411000001102 | Captopril | Captopril 25mg tablets (A A H Pharmaceuticals Ltd) |
| 44527 | 7659911000001107 | Captopril | Captopril 5mg/ ml oral solution sugar free |
| 45228 | 243265001000027100 | Captopril | Captopril capsules |
| 46851 | 8351611000001101 | Captopril | Captopril 5mg/ 5ml oral solution |
| 46951 | 226411000001105 | Captopril | Captopril 12.5mg tablets (A A H Pharmaceuticals Ltd) |

| | | | |
|---|---|---|---|
| 46957 | 13741311000001109 | Captopril | Captopril 12.5mg tablets (Tillomed Laboratories Ltd) |
| 52293 | 8791811000001109 | Captopril | Captopril 2mg capsules |
| 52499 | 8348511000001107 | Captopril | Captopril 25mg/ 5ml oral solution |
| 54544 | 8348611000001106 | Captopril | Captopril 25mg/ 5ml oral suspension |
| 56509 | 5528711000001108 | Captopril | Capoten 12.5mg tablets (Dowelhurst Ltd) |
| 56850 | 221411000001104 | Captopril | Ecopace 12.5mg tablets (AMCo) |
| 58195 | 8347111000001104 | Captopril | Captopril 12.5mg/ 5ml oral solution |
| 59699 | 23707511000001102 | Captopril | Captopril 5mg/ 5ml oral solution sugar free |
| 59915 | 23707311000001108 | Captopril | Captopril 25mg/ 5ml oral solution sugar free |
| 60349 | 23681711000001107 | Captopril | Noyada 25mg/ 5ml oral solution (Martindale Pharmaceuticals Ltd) |
| 60823 | 23682011000001102 | Captopril | Noyada 5mg/ 5ml oral solution (Martindale Pharmaceuticals Ltd) |
| 64739 | 8317011000001108 | Captopril | Captopril 25mg/ 5ml oral solution (Special Order) |
| 66597 | 8346811000001109 | Captopril | Captopril 10mg/ 5ml oral suspension |
| 69192 | 243255001000027109 | Captopril | Captopril oral solution |
| 69599 | 8351111000001109 | Captopril | Captopril 500micrograms/ 5ml oral suspension |
| 69600 | 8347911000001101 | Captopril | Captopril 1mg/ 5ml oral suspension |
| 70994 | 654711000001106 | Captopril | Captopril 12.5mg tablets (Sandoz Ltd) |
| 71277 | 8353911000001108 | Captopril | Captopril 7.5mg/ 5ml oral suspension |
| 73659 | 291911000001104 | Captopril | Captopril 50mg tablets (Kent Pharmaceuticals Ltd) |
| 74417 | 8348311000001101 | Captopril | Captopril 20mg/ 5ml oral suspension |
| 74627 | 68765001000027106 | Captopril | Captopril 25mg Tablet (C P Pharmaceuticals Ltd) |
| 76433 | | Captopril | Captopril 1.5mg capsules |
| 77046 | | Captopril | Captopril 3mg oral powder sachets |
| 77361 | | Captopril | Capoten 25mg tablets (Waymade Healthcare Plc) |
| 77364 | | Captopril | Capoten 25mg tablets (Stephar (U.K.) Ltd) |
| 77400 | | Captopril | Capoten 25mg tablets (Mawdsley-Brooks & Company Ltd) |
| 77410 | | Captopril | Capoten 50mg tablets (Dowelhurst Ltd) |
| 77415 | | Captopril | Capoten 50mg tablets (Waymade Healthcare Plc) |
| 77597 | | Captopril | Captopril 50mg/ 5ml oral suspension |
| 217 | 118915001000027101 | | CAPTOPRIL 4 MG/ ML LIQ |
| 2927 | 98615001000027100 | | PERINDOPRIL/ TERT-BUTYLAMINE 2 MG TAB |
| 3509 | 82805001000027100 | | ENALAPRIL MALEATE 40 MG TAB |
| 6200 | 3042211000001107 | | Tritace titration pack capsules (Sanofi) |
| 8923 | 91075001000027109 | | CAPTOPRIL 100 MG TAB |
| 22004 | | | CARACE (SPECIAL COMPLIANCE PACK) |
| 22882 | | | RAMIPRIL |
| 23382 | | | CARACE (SPECIAL COMPLIANCE PACK) |
| 24214 | 178845001000027106 | | TRITACE 1.25 MG TAB |
| 24693 | | | CARACE (SPECIAL COMPLIANCE PACK) |
| 27890 | | | ENALAPRIL MALEATE |
| 29964 | 172005001000027105 | | TRITACE 2.5 MG TAB |
| 31288 | | | TRITACE |
| 39421 | 13600911000001104 | | Tritace titration pack tablets (Sanofi) |

| | | | |
|---|---|---|---|
| 63594 | 13610111000001104 | | Generic Tritace titration pack tablets |
| **ACE-inhibitors in combination with calcium channel blockers** | | | |
| 18223 | 232125001000027103 | Verapamil / Trandolapril | Trandolapril with verapamil 2mg + 180mg Modified-release capsule |
| 19690 | 36149211000001102 | Verapamil / Trandolapril | Verapamil 180mg modified-release / Trandolapril 2mg capsules |
| 20579 | 3691211000001101 | Verapamil / Trandolapril | Tarka modified-release capsules (Abbott Laboratories Ltd) |
| 60684 | 23984911000001108 | Perindopril erbumine/ Amlodipine besilate | Perindopril erbumine 4mg / Amlodipine 10mg tablets |
| 11567 | 216655001000027100 | Felodipine/ Ramipril | Ramipril 5mg with felodipine 5mg modified-release tablet |
| 11965 | 216645001000027103 | Felodipine/ Ramipril | Ramipril 2.5mg with felodipine 2.5mg modified-release tablet |
| 17006 | 3887911000001109 | Felodipine/ Ramipril | Triapin 5mg/ 5mg modified-release tablets (Sanofi) |
| 17474 | 318177008 | Felodipine/ Ramipril | Felodipine 5mg modified-release / Ramipril 5mg tablets |
| 21162 | 318176004 | Felodipine/ Ramipril | Felodipine 2.5mg modified-release / Ramipril 2.5mg tablets |
| 28438 | 4093211000001109 | Felodipine/ Ramipril | Triapin 2.5mg/ 2.5mg modified-release tablets (Sanofi) |
| 60067 | 23985011000001108 | Amlodipine besilate/ Perindopril erbumine | Perindopril erbumine 4mg / Amlodipine 5mg tablets |
| 60744 | 23985211000001103 | Amlodipine besilate/ Perindopril erbumine | Perindopril erbumine 8mg / Amlodipine 5mg tablets |
| 63149 | 23985111000001109 | Amlodipine besilate/ Perindopril erbumine | Perindopril erbumine 8mg / Amlodipine 10mg tablets |
| **ACE- inhibitors in combination with diuretics** | | | |
| 6794 | 3437611000001100 | Perindopril erbumine/ Indapamide | Perindopril erbumine 4mg / Indapamide 1.25mg tablets |
| 14228 | 562511000001109 | Perindopril erbumine/ Indapamide | Coversyl Plus tablets (Servier Laboratories Ltd) |
| 48098 | 263725001000027105 | Perindopril Erbumine/ Indapamide | Perindopril arginine 4mg with Indapamide 1.25mg tablet |
| 50607 | 246725001000027107 | Perindopril Erbumine/ Indapamide | Perindopril arginine 2mg with Indapamide 625 micrograms tablet |
| 37908 | 13444311000001104 | Perindopril arginine/ Indapamide | Coversyl Arginine Plus 5mg/ 1.25mg tablets (Servier Laboratories Ltd) |
| 37978 | 13454311000001101 | Perindopril arginine/ Indapamide | Perindopril arginine 5mg / Indapamide 1.25mg tablets |
| 51258 | 18571111000001108 | Perindopril arginine/ Indapamide | Coversyl Arginine Plus 5mg/ 1.25mg tablets (DE Pharmaceuticals) |
| 2982 | 27665001000027103 | Lisinopril/ Hydrochlorothiazide | Zestoretic 20- 20mg+12.5mg Tablet (AstraZeneca UK Ltd) |
| 6359 | 27675001000027105 | Lisinopril/ Hydrochlorothiazide | Zestoretic 10- 10mg+12.5mg Tablet (AstraZeneca UK Ltd) |
| 6468 | 318880006 | Lisinopril/ Hydrochlorothiazide | Lisinopril 20mg / Hydrochlorothiazide 12.5mg tablets |
| 6786 | 318884002 | Lisinopril/ Hydrochlorothiazide | Lisinopril 10mg / Hydrochlorothiazide 12.5mg tablets |
| 9764 | 178295001000027105 | Lisinopril/ Hydrochlorothiazide | Carace 20 Tablet (Bristol-Myers Squibb Pharmaceuticals Ltd) |
| 17655 | 178305001000027103 | Lisinopril/ Hydrochlorothiazide | Carace 10 Tablet (Bristol-Myers Squibb Pharmaceuticals Ltd) |
| 21231 | 7385711000001106 | Lisinopril/ Hydrochlorothiazide | Caralpha 20mg/ 12.5mg tablets (Actavis UK Ltd) |
| 33353 | 7334911000001102 | Lisinopril/ Hydrochlorothiazide | Lisinopril 20mg / Hydrochlorothiazide 12.5mg tablets (Teva UK Ltd) |
| 37710 | 7334711000001104 | Lisinopril/ Hydrochlorothiazide | Lisinopril 10mg / Hydrochlorothiazide 12.5mg tablets (Teva UK Ltd) |
| 38995 | 3143111000001100 | Lisinopril/ Hydrochlorothiazide | Zestoretic 20 tablets (AstraZeneca UK Ltd) |
| 39137 | 3144311000001101 | Lisinopril/ Hydrochlorothiazide | Zestoretic 10 tablets (AstraZeneca UK Ltd) |
| 39147 | 3143511000001109 | Lisinopril/ Hydrochlorothiazide | Carace 20 Plus tablets (Merck Sharp & Dohme Ltd) |
| 39242 | 3144511000001107 | Lisinopril/ Hydrochlorothiazide | Carace 10 Plus tablets (Merck Sharp & Dohme Ltd) |
| 54201 | 13845711000001107 | Lisinopril/ Hydrochlorothiazide | Lisinopril 20mg / Hydrochlorothiazide 12.5mg tablets (Almus Pharmaceuticals Ltd) |

| | | | |
|---|---|---|---|
| 55399 | 7495911000001104 | Lisinopril/ Hydrochlorothiazide | Lisinopril 20mg / Hydrochlorothiazide 12.5mg tablets (A A H Pharmaceuticals Ltd) |
| 56244 | 13754811000001101 | Lisinopril/ Hydrochlorothiazide | Lisinopril 20mg / Hydrochlorothiazide 12.5mg tablets (Tillomed Laboratories Ltd) |
| 57539 | 14767411000001104 | Lisinopril/ Hydrochlorothiazide | Zestoretic 10 tablets (Sigma Pharmaceuticals Plc) |
| 67767 | 13845511000001102 | Lisinopril/ Hydrochlorothiazide | Lisinopril 10mg / Hydrochlorothiazide 12.5mg tablets (Almus Pharmaceuticals Ltd) |
| 71115 | 10483611000001104 | Lisinopril/ Hydrochlorothiazide | Zestoretic 10 tablets (Waymade Healthcare Plc) |
| 74008 | 5426911000001100 | Lisinopril/ Hydrochlorothiazide | Zestoretic 20 tablets (Waymade Healthcare Plc) |
| 74040 | 16449811000001104 | Lisinopril/ Hydrochlorothiazide | Zestoretic 20 tablets (Lexon (UK) Ltd) |
| 74874 | 13983811000001102 | Lisinopril/ Hydrochlorothiazide | Zestoretic 20 tablets (DE Pharmaceuticals) |
| 56157 | 21940011000001101 | Indapamide/ Perindopril tosilate | Perindopril tosilate 5mg / Indapamide 1.25mg tablets |
| 15031 | 260211000001104 | Hydrochlorothiazide/ Quinapril | Accuretic 12.5mg/ 10mg tablets (Pfizer Ltd) |
| 15108 | 318892006 | Hydrochlorothiazide/ Quinapril | Quinapril 10mg / Hydrochlorothiazide 12.5mg tablets |
| 3203 | 15255501000027101 | Hydrochlorothiazide/ Captopril | Capozide LS Tablet (E R Squibb and Sons Ltd) |
| 11561 | 318806002 | Hydrochlorothiazide/ Captopril | Co-zidocapt 12.5mg/ 25mg tablets |
| 39227 | 546711000001107 | Hydrochlorothiazide/ Captopril | Capozide LS 12.5mg/ 25mg tablets (Bristol-Myers Squibb Pharmaceuticals Ltd) |
| 1021 | 146811000001108 | Enalapril maleate/ Hydrochlorothiazide | Innozide 20mg/ 12.5mg tablets (Merck Sharp & Dohme Ltd) |
| 5189 | 318849001 | Enalapril maleate/ Hydrochlorothiazide | Enalapril 20mg / Hydrochlorothiazide 12.5mg tablets |
| 76920 | | Enalapril maleate/ Hydrochlorothiazide | Enalapril 20mg / Hydrochlorothiazide 12.5mg tablets (Tillomed Laboratories Ltd) |
| 1520 | 17311000001102 | Captopril/ Hydrochlorothiazide | Capozide 25mg/ 50mg tablets (Bristol-Myers Squibb Pharmaceuticals Ltd) |
| 10902 | 15251501000027106 | Captopril/ Hydrochlorothiazide | Captopril 50mg with Hydrochlorothiazide 25mg tablets |
| 11133 | 15930501000027103 | Captopril/ Hydrochlorothiazide | Hydrochlorothiazide with captopril 25mg with 50mg Tablet |
| 11351 | 318807006 | Captopril/ Hydrochlorothiazide | Co-zidocapt 25mg/ 50mg tablets |
| 11641 | 15252501000027102 | Captopril/ Hydrochlorothiazide | Captopril 25mg with Hydrochlorothiazide 12.5mg tablets |
| 15135 | 15931501000027101 | Captopril/ Hydrochlorothiazide | Hydrochlorothiazide with captopril 12.5mg with 25mg Tablet |
| 18263 | 263311000001101 | Captopril/ Hydrochlorothiazide | Acezide 25mg/ 50mg tablets (Bristol-Myers Squibb Pharmaceuticals Ltd) |
| 32166 | 21477501000027102 | Captopril/ Hydrochlorothiazide | Capto-co 25mg+50mg Tablet (IVAX Pharmaceuticals UK Ltd) |
| 77457 | | Captopril/ Hydrochlorothiazide | Capozide 25mg/ 50mg tablets (Dowelhurst Ltd) |
| **Angiotensin II receptor blocker** | | | |
| 575 | 318961008 | Valsartan | Valsartan 40mg capsules |
| 3222 | 318962001 | Valsartan | Valsartan 80mg capsules |
| 4645 | 318963006 | Valsartan | Valsartan 160mg capsules |
| 6518 | 117011000001107 | Valsartan | Diovan 160mg capsules (Novartis Pharmaceuticals UK Ltd) |
| 11251 | 777611000001101 | Valsartan | Diovan 40mg capsules (Novartis Pharmaceuticals UK Ltd) |
| 11252 | 554511000001105 | Valsartan | Diovan 80mg capsules (Novartis Pharmaceuticals UK Ltd) |
| 14943 | 416515008 | Valsartan | Valsartan 40mg tablets |
| 24359 | 8263211000001101 | Valsartan | Diovan 40mg tablets (Novartis Pharmaceuticals UK Ltd) |
| 37573 | 376487009 | Valsartan | Valsartan 320mg tablets |
| 38395 | 375034009 | Valsartan | Valsartan 80mg tablets |
| 39199 | 13143311000001102 | Valsartan | Diovan 320mg tablets (Novartis Pharmaceuticals UK Ltd) |
| 44778 | 375035005 | Valsartan | Valsartan 160mg tablets |
| 45600 | 26165501000027107 | Valsartan | Diovan 160mg Tablet (Novartis Pharmaceuticals UK Ltd) |

| | | | |
|---|---|---|---|
| 53833 | 19827711000001100 | Valsartan | Valsartan 160mg capsules (Mylan) |
| 54726 | 19631111000001104 | Valsartan | Valsartan 40mg capsules (Teva UK Ltd) |
| 55187 | 20021711000001108 | Valsartan | Valsartan 160mg capsules (Arrow Generics Ltd) |
| 55821 | 19630911000001108 | Valsartan | Valsartan 160mg capsules (Teva UK Ltd) |
| 58669 | 19631111000001104 | Valsartan | Valsartan 40mg capsules (Teva UK Ltd) |
| 58910 | 14690811000001105 | Valsartan | Valsartan 80mg capsules (Sigma Pharmaceuticals Plc) |
| 59029 | 20007411000001100 | Valsartan | Valsartan 3mg/ ml oral solution |
| 59448 | 19803311000001101 | Valsartan | Valsartan 80mg capsules (A A H Pharmaceuticals Ltd) |
| 60076 | 21923911000001107 | Valsartan | Valsartan 160mg capsules (Waymade Healthcare Plc) |
| 61442 | 19630911000001108 | Valsartan | Valsartan 160mg capsules (Teva UK Ltd) |
| 67663 | 19683111000001105 | Valsartan | Valsartan 160mg capsules (Dexcel-Pharma Ltd) |
| 68948 | 19688111000001101 | Valsartan | Valsartan 160mg capsules (Actavis UK Ltd) |
| 70628 | 20001711000001102 | Valsartan | Diovan 3mg/ 1ml oral solution (Novartis Pharmaceuticals UK Ltd) |
| 71028 | 20021411000001102 | Valsartan | Valsartan 80mg capsules (Arrow Generics Ltd) |
| 74013 | 13866611000001102 | Valsartan | Diovan 160mg capsules (DE Pharmaceuticals) |
| 74055 | 15775511000001107 | Valsartan | Valsartan 40mg/ 5ml oral solution |
| 74057 | 15775611000001106 | Valsartan | Valsartan 40mg/ 5ml oral suspension |
| 75409 | | Valsartan | Valsartan 80mg capsules (Teva UK Ltd) |
| 5988 | 318986004 | Telmisartan | Telmisartan 40mg tablets |
| 6243 | 134463001 | Telmisartan | Telmisartan 20mg tablets |
| 12874 | 318987008 | Telmisartan | Telmisartan 80mg tablets |
| 13821 | 924911000001106 | Telmisartan | Micardis 40mg tablets (Boehringer Ingelheim Ltd) |
| 17545 | 527411000001102 | Telmisartan | Micardis 80mg tablets (Boehringer Ingelheim Ltd) |
| 17686 | 648711000001100 | Telmisartan | Micardis 20mg tablets (Boehringer Ingelheim Ltd) |
| 61177 | 14625011000001101 | Telmisartan | Telmisartan 20mg tablets (Sigma Pharmaceuticals Plc) |
| 65274 | 23669311000001109 | Telmisartan | Telmisartan 40mg tablets (Actavis UK Ltd) |
| 70251 | 23613311000001107 | Telmisartan | Telmisartan 20mg tablets (Teva UK Ltd) |
| 6217 | 408055003 | Olmesartan medoxomil | Olmesartan medoxomil 10mg tablets |
| 6285 | 385542009 | Olmesartan medoxomil | Olmesartan medoxomil 20mg tablets |
| 6351 | 385543004 | Olmesartan medoxomil | Olmesartan medoxomil 40mg tablets |
| 14983 | 4624011000001101 | Olmesartan medoxomil | Olmetec 10mg tablets (Daiichi Sankyo UK Ltd) |
| 18910 | 4624311000001103 | Olmesartan medoxomil | Olmetec 20mg tablets (Daiichi Sankyo UK Ltd) |
| 20117 | 4624611000001108 | Olmesartan medoxomil | Olmetec 40mg tablets (Daiichi Sankyo UK Ltd) |
| 39786 | 14680711000001103 | Olmesartan medoxomil | Olmesartan medoxomil 10mg/ 5ml oral suspension |
| 520 | 318955005 | Losartan potassium | Losartan 25mg tablets |
| 624 | 407784004 | Losartan potassium | Losartan 100mg tablets |
| 1780 | 318956006 | Losartan potassium | Losartan 50mg tablets |
| 4226 | 266511000001104 | Losartan potassium | Cozaar 25mg tablets (Merck Sharp & Dohme Ltd) |
| 5723 | 53611000001106 | Losartan potassium | Cozaar 50mg tablets (Merck Sharp & Dohme Ltd) |
| 14965 | 245811000001102 | Losartan potassium | Cozaar 100mg tablets (Merck Sharp & Dohme Ltd) |
| 39944 | 15148111000001100 | Losartan potassium | Losartan 12.5mg tablets |
| 40571 | 15138911000001101 | Losartan potassium | Cozaar 12.5mg tablets (Merck Sharp & Dohme Ltd) |
| 40711 | 15507411000001105 | Losartan potassium | Losartan 2.5mg/ ml oral suspension sugar free |

| | | | |
|---|---|---|---|
| 41232 | 15506811000001105 | Losartan potassium | Cozaar 2.5mg/ ml oral suspension (Merck Sharp & Dohme Ltd) |
| 47006 | 17024511000001106 | Losartan potassium | Losartan 100mg tablets (Teva UK Ltd) |
| 48398 | 16998411000001102 | Losartan potassium | Losartan 25mg tablets (Dexcel-Pharma Ltd) |
| 49492 | 16971611000001102 | Losartan potassium | Losartan 25mg tablets (Mylan) |
| 49588 | 16732611000001102 | Losartan potassium | Losartan 100mg tablets (A A H Pharmaceuticals Ltd) |
| 50971 | 17015511000001104 | Losartan potassium | Losartan 25mg tablets (A A H Pharmaceuticals Ltd) |
| 51186 | 17660711000001105 | Losartan potassium | Losartan 25mg tablets (Arrow Generics Ltd) |
| 51601 | 16749111000001100 | Losartan potassium | Losartan 50mg tablets (Actavis UK Ltd) |
| 52427 | 18226311000001106 | Losartan potassium | Cozaar 100mg tablets (Necessity Supplies Ltd) |
| 52658 | 15451211000001101 | Losartan potassium | Losartan 100mg/ 5ml oral suspension |
| 52659 | 14159411000001106 | Losartan potassium | Losartan 50mg/ 5ml oral solution |
| 52886 | 17015311000001105 | Losartan potassium | Losartan 12.5mg tablets (A A H Pharmaceuticals Ltd) |
| 54049 | 18464011000001108 | Losartan potassium | Losartan 50mg tablets (Accord Healthcare Ltd) |
| 54057 | 17024311000001100 | Losartan potassium | Losartan 50mg tablets (Teva UK Ltd) |
| 54404 | 16749311000001103 | Losartan potassium | Losartan 100mg tablets (Actavis UK Ltd) |
| 54735 | 17026111000001109 | Losartan potassium | Losartan 50mg tablets (Alliance Healthcare (Distribution) Ltd) |
| 54740 | 16748711000001108 | Losartan potassium | Losartan 25mg tablets (Actavis UK Ltd) |
| 54843 | 16998911000001105 | Losartan potassium | Losartan 50mg tablets (Dexcel-Pharma Ltd) |
| 55296 | 16971811000001103 | Losartan potassium | Losartan 50mg tablets (Mylan) |
| 55446 | 21037511000001102 | Losartan potassium | Losartan 100mg tablets (Bristol Laboratories Ltd) |
| 55718 | 18140111000001105 | Losartan potassium | Losartan 25mg tablets (Phoenix Healthcare Distribution Ltd) |
| 56104 | 16732411000001100 | Losartan potassium | Losartan 50mg tablets (A A H Pharmaceuticals Ltd) |
| 56970 | 18169911000001109 | Losartan potassium | Losartan 100mg tablets (Pfizer Ltd) |
| 57028 | 16972011000001101 | Losartan potassium | Losartan 100mg tablets (Mylan) |
| 58274 | 18463811000001100 | Losartan potassium | Losartan 25mg tablets (Accord Healthcare Ltd) |
| 58649 | 21033711000001102 | Losartan potassium | Losartan 25mg tablets (Bristol Laboratories Ltd) |
| 58967 | 17025711000001102 | Losartan potassium | Losartan 12.5mg tablets (Alliance Healthcare (Distribution) Ltd) |
| 59086 | 20571411000001104 | Losartan potassium | Losartan 25mg tablets (Wockhardt UK Ltd) |
| 59271 | 20308011000001106 | Losartan potassium | Losartan 25mg tablets (Sandoz Ltd) |
| 59340 | 16998111000001107 | Losartan potassium | Losartan 12.5mg tablets (Dexcel-Pharma Ltd) |
| 59351 | 18170211000001103 | Losartan potassium | Losartan 50mg tablets (Pfizer Ltd) |
| 59750 | 22338211000001101 | Losartan potassium | Losartan 50mg tablets (Aptil Pharma Ltd) |
| 59903 | 14159511000001105 | Losartan potassium | Losartan 50mg/ 5ml oral suspension |
| 60506 | 16999411000001105 | Losartan potassium | Losartan 100mg tablets (Dexcel-Pharma Ltd) |
| 61053 | 17026311000001106 | Losartan potassium | Losartan 100mg tablets (Alliance Healthcare (Distribution) Ltd) |
| 61288 | 18463611000001104 | Losartan potassium | Losartan 100mg tablets (Accord Healthcare Ltd) |
| 61495 | 22338011000001106 | Losartan potassium | Losartan 25mg tablets (Aptil Pharma Ltd) |
| 61754 | 14204111000001106 | Losartan potassium | Losartan 25mg/ 5ml oral suspension |
| 62388 | 24372011000001100 | Losartan potassium | Losartan 12.5mg tablets (DE Pharmaceuticals) |
| 63222 | 18170411000001104 | Losartan potassium | Losartan 25mg tablets (Pfizer Ltd) |
| 63918 | 17024111000001102 | Losartan potassium | Losartan 25mg tablets (Teva UK Ltd) |
| 64888 | 29898911000001106 | Losartan potassium | Losartan 12.5mg tablets (Sigma Pharmaceuticals Plc) |
| 65094 | 20308411000001102 | Losartan potassium | Losartan 50mg tablets (Sandoz Ltd) |

| | | | |
|---|---|---|---|
| 66114 | 30811311000001102 | Losartan potassium | Losartan 12.5mg tablets (Mawdsley-Brooks & Company Ltd) |
| 66551 | 21036211000001102 | Losartan potassium | Losartan 50mg tablets (Bristol Laboratories Ltd) |
| 67902 | 20308611000001104 | Losartan potassium | Losartan 100mg tablets (Sandoz Ltd) |
| 68340 | 32661211000001105 | Losartan potassium | Losartan 12.5mg tablets (Consilient Health Ltd) |
| 68603 | 20571811000001102 | Losartan potassium | Losartan 50mg tablets (Wockhardt UK Ltd) |
| 69667 | 15451111000001107 | Losartan potassium | Losartan 100mg/ 5ml oral solution |
| 69858 | 17025911000001100 | Losartan potassium | Losartan 25mg tablets (Alliance Healthcare (Distribution) Ltd) |
| 70325 | 32617311000001107 | Losartan potassium | Losartan 50mg tablets (Almus Pharmaceuticals Ltd) |
| 70765 | 32616911000001105 | Losartan potassium | Losartan 100mg tablets (Almus Pharmaceuticals Ltd) |
| 71910 | 18494111000001106 | Losartan potassium | Losartan 50mg tablets (Necessity Supplies Ltd) |
| 74243 | 35657011000001102 | Losartan potassium | Losartan 50mg tablets (Consilient Health Ltd) |
| 74589 | 32753911000001104 | Losartan potassium | Losartan 25mg tablets (Genesis Pharmaceuticals Ltd) |
| 74904 | 24372211000001105 | Losartan potassium | Losartan 25mg tablets (DE Pharmaceuticals) |
| 77196 | | Losartan potassium | Losartan 25mg tablets (Kent Pharmaceuticals Ltd) |
| 77408 | | Losartan potassium | Cozaar 50mg tablets (Waymade Healthcare Plc) |
| 828 | 318968002 | Irbesartan | Irbesartan 75mg tablets |
| 1293 | 318969005 | Irbesartan | Irbesartan 150mg tablets |
| 2971 | 318970006 | Irbesartan | Irbesartan 300mg tablets |
| 7338 | 434511000001104 | Irbesartan | Aprovel 75mg tablets (Sanofi) |
| 9196 | 859711000001103 | Irbesartan | Aprovel 150mg tablets (Sanofi) |
| 11348 | 323211000001107 | Irbesartan | Aprovel 300mg tablets (Sanofi) |
| 36939 | 12639511000001103 | Irbesartan | Irbesartan 300mg/ 5ml oral suspension |
| 52972 | 14261011000001108 | Irbesartan | Irbesartan 300mg tablets (Sigma Pharmaceuticals Plc) |
| 55017 | 21285511000001106 | Irbesartan | Irbesartan 300mg tablets (Accord Healthcare Ltd) |
| 58108 | 21230711000001102 | Irbesartan | Irbesartan 150mg tablets (A A H Pharmaceuticals Ltd) |
| 58201 | 21219211000001108 | Irbesartan | Irbesartan 150mg tablets (Actavis UK Ltd) |
| 59393 | 21993211000001106 | Irbesartan | Irbesartan 300mg tablets (Sandoz Ltd) |
| 60597 | 21112911000001107 | Irbesartan | Irbesartan 150mg tablets (Teva UK Ltd) |
| 61781 | 21113111000001103 | Irbesartan | Irbesartan 300mg tablets (Teva UK Ltd) |
| 62415 | 21100511000001103 | Irbesartan | Irbesartan 300mg tablets (A A H Pharmaceuticals Ltd) |
| 63385 | 21522211000001102 | Irbesartan | Sabervel 75mg tablets (Aspire Pharma Ltd) |
| 63411 | 21123811000001104 | Irbesartan | Irbesartan 300mg tablets (Alliance Healthcare (Distribution) Ltd) |
| 63717 | 24177711000001109 | Irbesartan | Irbesartan 300mg tablets (DE Pharmaceuticals) |
| 65065 | 21230511000001107 | Irbesartan | Irbesartan 75mg tablets (A A H Pharmaceuticals Ltd) |
| 70431 | 27588011000001102 | Irbesartan | Irbesartan 150mg tablets (Lupin Healthcare (UK) Ltd) |
| 70955 | 194235001000027100 | Irbesartan | Irbesartan 300mg/ 5ml Oral suspension (Martindale Pharmaceuticals Ltd) |
| 71019 | 21623611000001102 | Irbesartan | Irbesartan 75mg tablets (Dr Reddy's Laboratories (UK) Ltd) |
| 71096 | 21624111000001107 | Irbesartan | Irbesartan 150mg tablets (Dr Reddy's Laboratories (UK) Ltd) |
| 71215 | 21219011000001103 | Irbesartan | Irbesartan 75mg tablets (Actavis UK Ltd) |
| 72000 | 21123311000001108 | Irbesartan | Irbesartan 150mg tablets (Alliance Healthcare (Distribution) Ltd) |
| 76202 | | Irbesartan | Irbesartan 150mg/ 5ml oral suspension |
| 76797 | | Irbesartan | Ifirmasta 75mg tablets (Consilient Health Ltd) |
| 77350 | | Irbesartan | Aprovel 150mg tablets (Dowelhurst Ltd) |

| | | | |
|---|---|---|---|
| 77443 | | Irbesartan | Aprovel 300mg tablets (Mawdsley-Brooks & Company Ltd) |
| 78012 | | Irbesartan | Ifirmasta 150mg tablets (Consilient Health Ltd) |
| 6939 | 318994006 | Eprosartan mesilate | Eprosartan 300mg tablets |
| 9745 | 401211000001105 | Eprosartan mesilate | Teveten 300mg tablets (Mylan) |
| 12836 | 318996008 | Eprosartan mesilate | Eprosartan 600mg tablets |
| 13123 | 318995007 | Eprosartan mesilate | Eprosartan 400mg tablets |
| 16285 | 151411000001103 | Eprosartan mesilate | Teveten 400mg tablets (Abbott Healthcare Products Ltd) |
| 16371 | 872011000001109 | Eprosartan mesilate | Teveten 600mg tablets (Mylan) |
| 63337 | 21229711000001107 | Eprosartan mesilate | Eprosartan 600mg tablets (A A H Pharmaceuticals Ltd) |
| 529 | 318977009 | Candesartan cilexetil | Candesartan 2mg tablets |
| 531 | 318978004 | Candesartan cilexetil | Candesartan 4mg tablets |
| 4155 | 97311000001103 | Candesartan cilexetil | Amias 2mg tablets (Takeda UK Ltd) |
| 4685 | 857411000001100 | Candesartan cilexetil | Amias 4mg tablets (Takeda UK Ltd) |
| 4741 | 318980005 | Candesartan cilexetil | Candesartan 16mg tablets |
| 4818 | 318979007 | Candesartan cilexetil | Candesartan 8mg tablets |
| 5013 | 36011000001106 | Candesartan cilexetil | Amias 8mg tablets (Takeda UK Ltd) |
| 5117 | 908511000001100 | Candesartan cilexetil | Amias 16mg tablets (Takeda UK Ltd) |
| 7043 | 376998003 | Candesartan cilexetil | Candesartan 32mg tablets |
| 31072 | 8983911000001107 | Candesartan cilexetil | Amias 32mg tablets (Takeda UK Ltd) |
| 50185 | 20476311000001109 | Candesartan cilexetil | Candesartan 8mg tablets (Teva UK Ltd) |
| 51117 | 13840411000001108 | Candesartan cilexetil | Candesartan 8mg tablets (DE Pharmaceuticals) |
| 51519 | 20495911000001109 | Candesartan cilexetil | Candesartan 8mg tablets (A A H Pharmaceuticals Ltd) |
| 51647 | 16353011000001102 | Candesartan cilexetil | Candesartan 4mg tablets (Mawdsley-Brooks & Company Ltd) |
| 52208 | 20496211000001106 | Candesartan cilexetil | Candesartan 16mg tablets (A A H Pharmaceuticals Ltd) |
| 52559 | 20913011000001106 | Candesartan cilexetil | Candesartan 8mg tablets (Zentiva) |
| 53680 | 20476511000001103 | Candesartan cilexetil | Candesartan 16mg tablets (Teva UK Ltd) |
| 53755 | 20476011000001106 | Candesartan cilexetil | Candesartan 4mg tablets (Teva UK Ltd) |
| 54326 | 20476711000001108 | Candesartan cilexetil | Candesartan 32mg tablets (Teva UK Ltd) |
| 54414 | 20530511000001105 | Candesartan cilexetil | Candesartan 16mg tablets (Consilient Health Ltd) |
| 57026 | 21813511000001100 | Candesartan cilexetil | Candesartan 8mg tablets (Waymade Healthcare Plc) |
| 57266 | 21666411000001103 | Candesartan cilexetil | Candesartan 2mg tablets (Actavis UK Ltd) |
| 57273 | 20483811000001104 | Candesartan cilexetil | Candesartan 8mg tablets (Actavis UK Ltd) |
| 57977 | 20509411000001107 | Candesartan cilexetil | Candesartan 16mg tablets (Alliance Healthcare (Distribution) Ltd) |
| 58646 | 20483511000001102 | Candesartan cilexetil | Candesartan 4mg tablets (Actavis UK Ltd) |
| 59690 | 20530311000001104 | Candesartan cilexetil | Candesartan 8mg tablets (Consilient Health Ltd) |
| 59802 | 20595811000001108 | Candesartan cilexetil | Candesartan 2mg tablets (Teva UK Ltd) |
| 62035 | 21813311000001106 | Candesartan cilexetil | Candesartan 16mg tablets (Waymade Healthcare Plc) |
| 62140 | 24505711000001100 | Candesartan cilexetil | Candesartan 4mg tablets (Sandoz Ltd) |
| 64359 | 22626711000001100 | Candesartan cilexetil | Candesartan 4mg tablets (DE Pharmaceuticals) |
| 65228 | 16353811000001108 | Candesartan cilexetil | Candesartan 16mg tablets (Mawdsley-Brooks & Company Ltd) |
| 65479 | 20493511000001101 | Candesartan cilexetil | Candesartan 2mg tablets (A A H Pharmaceuticals Ltd) |
| 66624 | 24506211000001101 | Candesartan cilexetil | Candesartan 16mg tablets (Sandoz Ltd) |
| 66958 | 24506011000001106 | Candesartan cilexetil | Candesartan 8mg tablets (Sandoz Ltd) |

| | | | |
|---|---|---|---|
| 67929 | 32554811000001106 | Candesartan cilexetil | Candesartan 16mg tablets (Tillomed Laboratories Ltd) |
| 68647 | 32746311000001107 | Candesartan cilexetil | Candesartan 8mg tablets (Genesis Pharmaceuticals Ltd) |
| 68718 | 20495511000001102 | Candesartan cilexetil | Candesartan 4mg tablets (A A H Pharmaceuticals Ltd) |
| 68751 | 32746511000001101 | Candesartan cilexetil | Candesartan 16mg tablets (Genesis Pharmaceuticals Ltd) |
| 69802 | 33582711000001106 | Candesartan cilexetil | Candesartan 4mg tablets (Mylan) |
| 70455 | 15825311000001103 | Candesartan cilexetil | Candesartan 4mg/ 5ml oral suspension |
| 70805 | 34007811000001104 | Candesartan cilexetil | Candesartan 8mg tablets (Crescent Pharma Ltd) |
| 71080 | 20912811000001108 | Candesartan cilexetil | Candesartan 4mg tablets (Zentiva) |
| 72215 | 20530011000001102 | Candesartan cilexetil | Candesartan 4mg tablets (Consilient Health Ltd) |
| 72924 | 23646611000001101 | Candesartan cilexetil | Candesartan 4mg tablets (Phoenix Healthcare Distribution Ltd) |
| 73528 | 35139711000001105 | Candesartan cilexetil | Candesartan 4mg tablets (Milpharm Ltd) |
| 73811 | 22947511000001109 | Candesartan cilexetil | Candesartan 2mg tablets (Ranbaxy (UK) Ltd) |
| 73813 | 21812911000001100 | Candesartan cilexetil | Candesartan 4mg tablets (Waymade Healthcare Plc) |
| 75407 | | Candesartan cilexetil | Candesartan 32mg tablets (Sandoz Ltd) |
| 75418 | | Candesartan cilexetil | Candesartan 16mg tablets (Mylan) |
| 76070 | | Candesartan cilexetil | Candesartan 8mg tablets (Almus Pharmaceuticals Ltd) |
| 76209 | | Candesartan cilexetil | Candesartan 8mg tablets (Mylan) |
| 77128 | | Candesartan cilexetil | Candesartan 16mg tablets (Actavis UK Ltd) |
| 51368 | 449333009 | Azilsartan medoxomil | Azilsartan medoxomil 80mg tablets |
| 51897 | 20350911000001100 | Azilsartan medoxomil | Edarbi 20mg tablets (Takeda UK Ltd) |
| 56606 | 449109006 | Azilsartan medoxomil | Azilsartan medoxomil 40mg tablets |
| 76119 | | Azilsartan medoxomil | Azilsartan medoxomil 20mg tablets |
| 76121 | | Azilsartan medoxomil | Edarbi 40mg tablets (Takeda UK Ltd) |
| 31160 | | | IRBESARTAN |
| **Angiotensin II receptor blockers in combination with calcium channel blockers** | | | |
| 35096 | 11160711000001108 | Valsartan/ Amlodipine besilate | Exforge 10mg/ 160mg tablets (Novartis Pharmaceuticals UK Ltd) |
| 35189 | 11160111000001107 | Valsartan/ Amlodipine besilate | Amlodipine 10mg / Valsartan 160mg tablets |
| 35317 | 11161811000001108 | Valsartan/ Amlodipine besilate | Exforge 5mg/ 80mg tablets (Novartis Pharmaceuticals UK Ltd) |
| 35329 | 11160311000001109 | Valsartan/ Amlodipine besilate | Amlodipine 5mg / Valsartan 80mg tablets |
| 35343 | 11160211000001101 | Valsartan/ Amlodipine besilate | Amlodipine 5mg / Valsartan 160mg tablets |
| 35697 | 11161511000001105 | Valsartan/ Amlodipine besilate | Exforge 5mg/ 160mg tablets (Novartis Pharmaceuticals UK Ltd) |
| 47727 | 18987311000001103 | Olmesartan medoxomil/ Hydrochlorothiazide/ Amlodipine besilate | Sevikar HCT 40mg/ 5mg/ 25mg tablets (Daiichi Sankyo UK Ltd) |
| 53220 | 18987611000001108 | Olmesartan medoxomil/ Hydrochlorothiazide/ Amlodipine besilate | Sevikar HCT 40mg/ 10mg/ 25mg tablets (Daiichi Sankyo UK Ltd) |
| 39984 | 15773211000001105 | Olmesartan medoxomil/ Amlodipine besilate | Sevikar 20mg/ 5mg tablets (Daiichi Sankyo UK Ltd) |
| 40316 | 429502004 | Olmesartan medoxomil/ Amlodipine besilate | Olmesartan medoxomil 20mg / Amlodipine 5mg tablets |
| 40639 | 429503009 | Olmesartan medoxomil/ Amlodipine besilate | Olmesartan medoxomil 40mg / Amlodipine 5mg tablets |

| | | | |
|---|---|---|---|
| 40668 | 429678006 | Olmesartan medoxomil/ Amlodipine besilate | Olmesartan medoxomil 40mg / Amlodipine 10mg tablets |
| 41203 | 15772611000001102 | Olmesartan medoxomil/ Amlodipine besilate | Sevikar 40mg/ 10mg tablets (Daiichi Sankyo UK Ltd) |
| 41205 | 15772911000001108 | Olmesartan medoxomil/ Amlodipine besilate | Sevikar 40mg/ 5mg tablets (Daiichi Sankyo UK Ltd) |
| 46355 | 18986411000001108 | Hydrochlorothiazide/ Olmesartan medoxomil/ Amlodipine besilate | Sevikar HCT 20mg/ 5mg/ 12.5mg tablets (Daiichi Sankyo UK Ltd) |
| 60780 | 18987811000001107 | Hydrochlorothiazide/ Olmesartan medoxomil/ Amlodipine besilate | Generic Sevikar HCT 20mg/ 5mg/ 12.5mg tablets |
| 46687 | 26494500100002710З | Amlodipine/ Hydrochlorothiazide/ Olmesartan Medoxomil | Olmesartan medoxomil with amlodipine and hydrochlorothiazide 20mg + 5mg + 12.5mg Tablet |
| 46715 | 264965001000027109 | Amlodipine/ Hydrochlorothiazide/ Olmesartan Medoxomil | Olmesartan medoxomil with amlodipine and hydrochlorothiazide 40mg + 10mg + 12.5mg Tablet |
| 46792 | 264955001000027100 | Amlodipine/ Hydrochlorothiazide/ Olmesartan Medoxomil | Olmesartan medoxomil with amlodipine and hydrochlorothiazide 40mg + 5mg + 12.5mg Tablet |
| 47467 | 264975001000027102 | Amlodipine/ Hydrochlorothiazide/ Olmesartan Medoxomil | Olmesartan medoxomil with amlodipine and hydrochlorothiazide 40mg + 5mg + 25mg Tablet |
| 55358 | 264985001000027107 | Amlodipine/ Hydrochlorothiazide/ Olmesartan Medoxomil | Olmesartan medoxomil with amlodipine and hydrochlorothiazide 40mg + 10mg + 25mg Tablet |
| 35173 | 246845001000027107 | Amlodipine Besilate/ Valsartan | Valsartan 160mg with amlodipine 5mg tablets |
| 35174 | 246835001000027106 | Amlodipine Besilate/ Valsartan | Valsartan 80mg with amlodipine 5mg tablets |
| 35304 | 246855001000027105 | Amlodipine Besilate/ Valsartan | Valsartan 160mg with amlodipine 10mg tablets |
| 47573 | 18986711000001102 | Amlodipine besilate/ Hydrochlorothiazide/ Olmesartan medoxomil | Sevikar HCT 40mg/ 5mg/ 12.5mg tablets (Daiichi Sankyo UK Ltd) |
| 47616 | 18987011000001101 | Amlodipine besilate/ Hydrochlorothiazide/ Olmesartan medoxomil | Sevikar HCT 40mg/ 10mg/ 12.5mg tablets (Daiichi Sankyo UK Ltd) |
| 60007 | 18987911000001102 | Amlodipine besilate/ Hydrochlorothiazide/ Olmesartan medoxomil | Generic Sevikar HCT 40mg/ 10mg/ 12.5mg tablets |
| **Angiotensin II receptor blockers and diuretics** | | | |
| 764 | 8150111000001108 | Valsartan/ Hydrochlorothiazide | Co-Diovan 80mg/ 12.5mg tablets (Novartis Pharmaceuticals UK Ltd) |
| 6877 | 7668611000001104 | Valsartan/ Hydrochlorothiazide | Co-Diovan 160mg/ 12.5mg tablets (Novartis Pharmaceuticals UK Ltd) |
| 11864 | 395521005 | Valsartan/ Hydrochlorothiazide | Valsartan 160mg / Hydrochlorothiazide 12.5mg tablets |
| 14283 | 409298002 | Valsartan/ Hydrochlorothiazide | Valsartan 160mg / Hydrochlorothiazide 25mg tablets |
| 16060 | 377488008 | Valsartan/ Hydrochlorothiazide | Valsartan 80mg / Hydrochlorothiazide 12.5mg tablets |
| 23456 | 238185001000027100 | Valsartan/ Hydrochlorothiazide | Hydrochlorothiazide with valsartan 25mg with 160mg Tablet |
| 24268 | 239025001000027101 | Valsartan/ Hydrochlorothiazide | Hydrochlorothiazide with valsartan 12.5mg with 80mg Tablet |
| 24484 | 238175001000027105 | Valsartan/ Hydrochlorothiazide | Hydrochlorothiazide with valsartan 12.5mg with 160mg Tablet |
| 25382 | 7668911000001105 | Valsartan/ Hydrochlorothiazide | Co-Diovan 160mg/ 25mg tablets (Novartis Pharmaceuticals UK Ltd) |
| 52858 | 14206911000001101 | Valsartan/ Hydrochlorothiazide | Co-Diovan 80mg/ 12.5mg tablets (Sigma Pharmaceuticals Plc) |

| | | | |
|---|---|---|---|
| 67664 | 19631911000001101 | Valsartan/ Hydrochlorothiazide | Valsartan 160mg / Hydrochlorothiazide 12.5mg tablets (Teva UK Ltd) |
| 72086 | 19688511000001105 | Valsartan/ Hydrochlorothiazide | Valsartan 160mg / Hydrochlorothiazide 12.5mg tablets (Actavis UK Ltd) |
| 14870 | 407855002 | Telmisartan/ Hydrochlorothiazide | Telmisartan 40mg / Hydrochlorothiazide 12.5mg tablets |
| 16161 | 407856001 | Telmisartan/ Hydrochlorothiazide | Telmisartan 80mg / Hydrochlorothiazide 12.5mg tablets |
| 17689 | 3806911000001105 | Telmisartan/ Hydrochlorothiazide | MicardisPlus 80mg/ 12.5mg tablets (Boehringer Ingelheim Ltd) |
| 18202 | 3806311000001109 | Telmisartan/ Hydrochlorothiazide | MicardisPlus 40mg/ 12.5mg tablets (Boehringer Ingelheim Ltd) |
| 62376 | 24412511000001109 | Telmisartan/ Hydrochlorothiazide | Actelsar HCT 80mg/ 12.5mg tablets (Actavis UK Ltd) |
| 63890 | 10540911000001100 | Telmisartan/ Hydrochlorothiazide | MicardisPlus 80mg/ 12.5mg tablets (Waymade Healthcare Plc) |
| 66997 | 29749811000001100 | Telmisartan/ Hydrochlorothiazide | MicardisPlus 40mg/ 12.5mg tablets (Waymade Healthcare Plc) |
| 76840 | | Telmisartan/ Hydrochlorothiazide | Actelsar HCT 40mg/ 12.5mg tablets (Actavis UK Ltd) |
| 43322 | 409185001 | Olmesartan medoxomil/ Hydrochlorothiazide | Olmesartan medoxomil 40mg / Hydrochlorothiazide 12.5mg tablets |
| 43915 | 17220911000001102 | Olmesartan medoxomil/ Hydrochlorothiazide | Olmetec Plus 40mg/ 12.5mg tablets (Daiichi Sankyo UK Ltd) |
| 4540 | 255911000001105 | Losartan potassium/ Hydrochlorothiazide | Cozaar-Comp 50mg/ 12.5mg tablets (Merck Sharp & Dohme Ltd) |
| 6437 | 318959004 | Losartan potassium/ Hydrochlorothiazide | Losartan 50mg / Hydrochlorothiazide 12.5mg tablets |
| 10323 | 395497004 | Losartan potassium/ Hydrochlorothiazide | Losartan 100mg / Hydrochlorothiazide 25mg tablets |
| 14738 | 209115001000027101 | Losartan Potassium/ Hydrochlorothiazide | Hydrochlorothiazide with losartan 12.5mg with 50mg Tablet |
| 21423 | 9566911000001105 | Losartan potassium/ Hydrochlorothiazide | Cozaar-Comp 100mg/ 25mg tablets (Merck Sharp & Dohme Ltd) |
| 24632 | 241785001000027103 | Losartan Potassium/ Hydrochlorothiazide | Hydrochlorothiazide with losartan 25mg with 100mg Tablet |
| 38367 | 250095001000027100 | Losartan Potassium/ Hydrochlorothiazide | Hydrochlorothiazide with losartan 12.5mg with 100mg Tablet |
| 52189 | 17015111000001108 | Losartan potassium/ Hydrochlorothiazide | Losartan 100mg / Hydrochlorothiazide 25mg tablets (A A H Pharmaceuticals Ltd) |
| 55160 | 14210911000001101 | Losartan potassium/ Hydrochlorothiazide | Cozaar-Comp 50mg/ 12.5mg tablets (Sigma Pharmaceuticals Plc) |
| 56204 | 18165111000001109 | Losartan potassium/ Hydrochlorothiazide | Losartan 50mg / Hydrochlorothiazide 12.5mg tablets (Actavis UK Ltd) |
| 56975 | 17014911000001107 | Losartan potassium/ Hydrochlorothiazide | Losartan 50mg / Hydrochlorothiazide 12.5mg tablets (A A H Pharmaceuticals Ltd) |
| 57796 | 19541911000001108 | Losartan potassium/ Hydrochlorothiazide | Cozaar-Comp 50mg/ 12.5mg tablets (DE Pharmaceuticals) |
| 62911 | 17024711000001101 | Losartan potassium/ Hydrochlorothiazide | Losartan 50mg / Hydrochlorothiazide 12.5mg tablets (Teva UK Ltd) |
| 66598 | 27475411000001100 | Losartan potassium/ Hydrochlorothiazide | Losartan 50mg / Hydrochlorothiazide 12.5mg tablets (Lupin Healthcare (UK) Ltd) |
| 70754 | 18552511000001109 | Losartan potassium/ Hydrochlorothiazide | Losartan 50mg / Hydrochlorothiazide 12.5mg tablets (Ranbaxy (UK) Ltd) |

| | | | |
|---|---|---|---|
| 71682 | 30826311000001104 | Losartan potassium/ Hydrochlorothiazide | Losartan 100mg / Hydrochlorothiazide 25mg tablets (Mawdsley-Brooks & Company Ltd) |
| 74215 | 24371611000001101 | Losartan potassium/ Hydrochlorothiazide | Losartan 50mg / Hydrochlorothiazide 12.5mg tablets (DE Pharmaceuticals) |
| 11469 | 134460003 | Irbesartan/ Hydrochlorothiazide | Irbesartan 300mg / Hydrochlorothiazide 12.5mg tablets |
| 11526 | 682711000001109 | Irbesartan/ Hydrochlorothiazide | CoAprovel 300mg/ 12.5mg tablets (Sanofi) |
| 35196 | 10968611000001106 | Irbesartan/ Hydrochlorothiazide | CoAprovel 300mg/ 25mg tablets (Sanofi) |
| 35481 | 10970311000001105 | Irbesartan/ Hydrochlorothiazide | Irbesartan 300mg / Hydrochlorothiazide 25mg tablets |
| 62337 | 23472611000001104 | Irbesartan/ Hydrochlorothiazide | Irbesartan 300mg / Hydrochlorothiazide 12.5mg tablets (Actavis UK Ltd) |
| 77641 | | Irbesartan/ Hydrochlorothiazide | CoAprovel 300mg/ 12.5mg tablets (Mawdsley-Brooks & Company Ltd) |
| 38459 | 13731911000001109 | Hydrochlorothiazide/ Telmisartan | Telmisartan 80mg / Hydrochlorothiazide 25mg tablets |
| 38889 | 13719711000001103 | Hydrochlorothiazide/ Telmisartan | MicardisPlus 80mg/ 25mg tablets (Boehringer Ingelheim Ltd) |
| 71748 | 24413611000001103 | Hydrochlorothiazide/ Telmisartan | Actelsar HCT 80mg/ 25mg tablets (Actavis UK Ltd) |
| 18200 | 409184002 | Hydrochlorothiazide/ Olmesartan medoxomil | Olmesartan medoxomil 20mg / Hydrochlorothiazide 12.5mg tablets |
| 18903 | 10270711000001105 | Hydrochlorothiazide/ Olmesartan medoxomil | Olmesartan medoxomil 20mg / Hydrochlorothiazide 25mg tablets |
| 27520 | 10261811000001100 | Hydrochlorothiazide/ Olmesartan medoxomil | Olmetec Plus 20mg/ 25mg tablets (Daiichi Sankyo UK Ltd) |
| 29634 | 10261511000001103 | Hydrochlorothiazide/ Olmesartan medoxomil | Olmetec Plus 20mg/ 12.5mg tablets (Daiichi Sankyo UK Ltd) |
| 35380 | 242645001000027102 | Hydrochlorothiazide/ Olmesartan Medoxomil | Hydrochlorothiazide with olmesartan medoxomil 12.5mg with 20mg tablet |
| 39021 | 242635001000027103 | Hydrochlorothiazide/ Olmesartan Medoxomil | Hydrochlorothiazide with olmesartan medoxomil 25mg with 20mg tablet |
| 37650 | 13112711000001103 | Hydrochlorothiazide/ Losartan potassium | Losartan 100mg / Hydrochlorothiazide 12.5mg tablets |
| 37747 | 13094111000001102 | Hydrochlorothiazide/ Losartan potassium | Cozaar-Comp 100mg/ 12.5mg tablets (Merck Sharp & Dohme Ltd) |
| 48039 | 19485511000001100 | Hydrochlorothiazide/ Losartan potassium | Losartan 100mg / Hydrochlorothiazide 12.5mg tablets (Teva UK Ltd) |
| 70922 | 18618811000001108 | Hydrochlorothiazide/ Losartan potassium | Losartan 100mg / Hydrochlorothiazide 12.5mg tablets (Phoenix Healthcare Distribution Ltd) |
| 71618 | 18276311000001107 | Hydrochlorothiazide/ Losartan potassium | Losartan 100mg / Hydrochlorothiazide 12.5mg tablets (A A H Pharmaceuticals Ltd) |
| 10316 | 792411000001108 | Hydrochlorothiazide/ Irbesartan | CoAprovel 150mg/ 12.5mg tablets (Sanofi) |
| 11448 | 134461004 | Hydrochlorothiazide/ Irbesartan | Irbesartan 150mg / Hydrochlorothiazide 12.5mg tablets |
| **Other fixed combinations of antihypertensives** | | | |
| 47727 | 18987311000001103 | Olmesartan medoxomil/ Hydrochlorothiazide/ Amlodipine besilate | Sevikar HCT 40mg/ 5mg/ 25mg tablets (Daiichi Sankyo UK Ltd) |

| | | | |
|---|---|---|---|
| 53220 | 18987611000001108 | Olmesartan medoxomil/ Hydrochlorothiazide/ Amlodipine besilate | Sevikar HCT 40mg/ 10mg/ 25mg tablets (Daiichi Sankyo UK Ltd) |
| 46355 | 18986411000001108 | Hydrochlorothiazide/ Olmesartan medoxomil/ Amlodipine besilate | Sevikar HCT 20mg/ 5mg/ 12.5mg tablets (Daiichi Sankyo UK Ltd) |
| 60780 | 18987811000001107 | Hydrochlorothiazide/ Olmesartan medoxomil/ Amlodipine besilate | Generic Sevikar HCT 20mg/ 5mg/ 12.5mg tablets |
| 18606 | 216735001000027106 | Diltiazem / Hydrochlorothiazide | Diltiazem and hydrochlorothiazide 150mg+12.5mg modified-release capsules |
| 23505 | 216725001000027109 | Diltiazem / Hydrochlorothiazide | Adizem xl plus 150mg+12.5mg Modified-release capsule (Napp Pharmaceuticals Ltd) |
| 4406 | 48465001000027107 | Benzthiazide/ methoserpidine | Decaserpyl plus Tablet (Roussel Laboratories Ltd) |
| 29696 | 124515001000027108 | Benzthiazide/ methoserpidine | Methoserpidine with benzthiazide Tablet |
| 46687 | 264945001000027103 | Amlodipine/ Hydrochlorothiazide/ Olmesartan Medoxomil | Olmesartan medoxomil with amlodipine and hydrochlorothiazide 20mg + 5mg + 12.5mg Tablet |
| 46715 | 264965001000027109 | Amlodipine/ Hydrochlorothiazide/ Olmesartan Medoxomil | Olmesartan medoxomil with amlodipine and hydrochlorothiazide 40mg + 10mg + 12.5mg Tablet |
| 46792 | 264955001000027100 | Amlodipine/ Hydrochlorothiazide/ Olmesartan Medoxomil | Olmesartan medoxomil with amlodipine and hydrochlorothiazide 40mg + 5mg + 12.5mg Tablet |
| 47467 | 264975001000027102 | Amlodipine/ Hydrochlorothiazide/ Olmesartan Medoxomil | Olmesartan medoxomil with amlodipine and hydrochlorothiazide 40mg + 5mg + 25mg Tablet |
| 55358 | 264985001000027107 | Amlodipine/ Hydrochlorothiazide/ Olmesartan Medoxomil | Olmesartan medoxomil with amlodipine and hydrochlorothiazide 40mg + 10mg + 25mg Tablet |
| 47573 | 18986711000001102 | Amlodipine besilate/ Hydrochlorothiazide/ Olmesartan medoxomil | Sevikar HCT 40mg/ 5mg/ 12.5mg tablets (Daiichi Sankyo UK Ltd) |
| 47616 | 18987011000001101 | Amlodipine besilate/ Hydrochlorothiazide/ Olmesartan medoxomil | Sevikar HCT 40mg/ 10mg/ 12.5mg tablets (Daiichi Sankyo UK Ltd) |
| 60007 | 18987911000001102 | Amlodipine besilate/ Hydrochlorothiazide/ Olmesartan medoxomil | Generic Sevikar HCT 40mg/ 10mg/ 12.5mg tablets |
| **Other antihypertensives** | | | |
| 29757 | 36143911000001108 | Trimetaphan camsilate | Trimetaphan camsilate 250mg/ 5ml solution for injection ampoules |
| 36612 | 375899001 | Sodium nitroprusside dihydrate | Sodium nitroprusside 50mg powder for solution for infusion vials |
| 37085 | 11394311000001108 | Sitaxentan sodium | Sitaxentan 100mg tablets |
| 40899 | 11392011000001104 | Sitaxentan sodium | Thelin 100mg tablets (Pfizer Ltd) |
| 64930 | 24408611000001108 | Riociguat | Riociguat 2mg tablets |
| 74720 | 24399711000001103 | Riociguat | Adempas 2mg tablets (Merck Sharp & Dohme Ltd) |
| 75040 | 24408411000001105 | Riociguat | Riociguat 1mg tablets |
| 75041 | 24408311000001103 | Riociguat | Riociguat 1.5mg tablets |
| 76902 | | Riociguat | Riociguat 500microgram tablets |
| 20656 | 76555001000027105 | Reserpine | Serpasil 250microgram Tablet (Novartis Pharmaceuticals UK Ltd) |
| 20690 | 139815001000027109 | Reserpine | Reserpine 250micrograms tablet |
| 21502 | 189705001000027101 | Reserpine | Serpasil 100microgram Tablet (Novartis Pharmaceuticals UK Ltd) |

| | | | |
|---|---|---|---|
| 22853 | 13980500100002710 7 | Reserpine | Reserpine 100micrograms tablet |
| 591 | 318768008 | Prazosin | Prazosin 1mg tablets |
| 726 | 318769000 | Prazosin | Prazosin 2mg tablets |
| 1292 | 347411000001101 | Prazosin | Hypovase 1mg tablets (Pfizer Ltd) |
| 1455 | 318767003 | Prazosin | Prazosin 500microgram tablets |
| 3715 | 318770004 | Prazosin | Prazosin 5mg tablets |
| 4111 | 321311000001100 | Prazosin | Hypovase 500microgram tablets (Pfizer Ltd) |
| 5183 | 150911000001104 | Prazosin | Hypovase 2mg tablets (Pfizer Ltd) |
| 8198 | 14555001000027108 | Prazosin | Hypovase 5mg Tablet (Pfizer Ltd) |
| 8863 | 15362500100002710 1 | Prazosin | Hypovase benign prostatic hyperplasia 1mg Tablet (Pfizer Ltd) |
| 13610 | 570011000001106 | Prazosin | Alphavase 2 tablets (Ashbourne Pharmaceuticals Ltd) |
| 19823 | 74711000001102 | Prazosin | Alphavase 5 tablets (Ashbourne Pharmaceuticals Ltd) |
| 23459 | 15365500100002710 0 | Prazosin | Hypovase benign prostatic hyperplasia 2mg Tablet (Pfizer Ltd) |
| 25047 | 15361500100002710 5 | Prazosin | Hypovase benign prostatic hyperplasia 500microgram Tablet (Pfizer Ltd) |
| 26237 | 30515001000027104 | Prazosin | Alphavase 500microgram Tablet (Ashbourne Pharmaceuticals Ltd) |
| 26238 | 934411000001100 | Prazosin | Alphavase 1 tablets (Ashbourne Pharmaceuticals Ltd) |
| 26693 | 15360500100002710 8 | Prazosin | Hypovase benign prostatic hyperplasia bd BD Starter pack (Pfizer Ltd) |
| 41651 | 52515001000027104 | Prazosin | Prazosin 500microgram Tablet (Approved Prescription Services Ltd) |
| 41652 | 89711000001108 | Prazosin | Prazosin 500microgram tablets (A A H Pharmaceuticals Ltd) |
| 41721 | 778311000001107 | Prazosin | Prazosin 1mg tablets (A A H Pharmaceuticals Ltd) |
| 43547 | 898711000001102 | Prazosin | Prazosin 500microgram tablets (IVAX Pharmaceuticals UK Ltd) |
| 46922 | 559411000001105 | Prazosin | Prazosin 1mg tablets (IVAX Pharmaceuticals UK Ltd) |
| 55826 | 739211000001106 | Prazosin | Prazosin 5mg tablets (A A H Pharmaceuticals Ltd) |
| 60316 | 52525001000027108 | Prazosin | Prazosin 1mg Tablet (Approved Prescription Services Ltd) |
| 4993 | 318707000 | Moxonidine | Moxonidine 200microgram tablets |
| 7174 | 318708005 | Moxonidine | Moxonidine 400microgram tablets |
| 9749 | 142811000001107 | Moxonidine | Physiotens 400microgram tablets (Mylan) |
| 9876 | 522011000001109 | Moxonidine | Physiotens 300microgram tablets (Mylan) |
| 10253 | 408604009 | Moxonidine | Moxonidine 300microgram tablets |
| 11177 | 41111000001102 | Moxonidine | Physiotens 200microgram tablets (Mylan) |
| 33322 | 8098911000001106 | Moxonidine | Moxonidine 200microgram tablets (Sandoz Ltd) |
| 40310 | 8390411000001102 | Moxonidine | Moxonidine 200microgram tablets (Teva UK Ltd) |
| 43531 | 8099411000001106 | Moxonidine | Moxonidine 400microgram tablets (Sandoz Ltd) |
| 60898 | 8171411000001109 | Moxonidine | Moxonidine 200microgram tablets (Mylan) |
| 61036 | 21407711000001105 | Moxonidine | Physiotens 300microgram tablets (Actavis UK Ltd) |
| 62853 | 8936111000001107 | Moxonidine | Moxonidine 200microgram tablets (A A H Pharmaceuticals Ltd) |
| 63938 | 8099111000001101 | Moxonidine | Moxonidine 300microgram tablets (Sandoz Ltd) |
| 67665 | 8265311000001105 | Moxonidine | Moxonidine 400microgram tablets (Actavis UK Ltd) |
| 67808 | 21406211000001100 | Moxonidine | Physiotens 200microgram tablets (Actavis UK Ltd) |
| 76479 | | Moxonidine | Moxonidine 400microgram tablets (A A H Pharmaceuticals Ltd) |
| 2967 | 318656009 | Minoxidil | Minoxidil 5mg tablets |
| 2968 | 318657000 | Minoxidil | Minoxidil 10mg tablets |
| 2970 | 318655008 | Minoxidil | Minoxidil 2.5mg tablets |

| | | | |
|---|---|---|---|
| 9463 | 3666411000001106 | Minoxidil | Loniten 5mg tablets (Pfizer Ltd) |
| 9697 | 3667011000001104 | Minoxidil | Loniten 2.5mg tablets (Pfizer Ltd) |
| 14495 | 3666711000001100 | Minoxidil | Loniten 10mg tablets (Pfizer Ltd) |
| 22454 | 167805001000027109 | Metirosine | Demser 250mg Capsule (Merck Sharp & Dohme Ltd) |
| 33788 | 167775001000027100 | Metirosine | Metirosine 250mg Capsule |
| 7416 | 51165001000027107 | Methyldopate | Aldomet 50mg/ ml Injection (Merck Sharp & Dohme Ltd) |
| 26919 | 125025001000027109 | Methyldopate | Methyldopa 50mg/ ml Injection |
| 21346 | 125115001000027106 | Methyldopa / Hydrochlorothiazide | Hydromet Tablet (MSD Thomas Morson Pharmaceuticals) |
| 28738 | 125055001000027105 | Methyldopa / Hydrochlorothiazide | Methyldopa with hydrochlorothiazide Tablet |
| 1707 | 318672001 | Methyldopa | Methyldopa 250mg tablets |
| 3049 | 318671008 | Methyldopa | Methyldopa 125mg tablets |
| 3070 | 318673006 | Methyldopa | Methyldopa 500mg tablets |
| 7626 | 655001000027106 | Methyldopa | Aldomet 250mg Tablet (Merck Sharp & Dohme Ltd) |
| 7642 | 665001000027102 | Methyldopa | Aldomet 500mg Tablet (Merck Sharp & Dohme Ltd) |
| 8033 | 645001000027108 | Methyldopa | Aldomet 125mg Tablet (Merck Sharp & Dohme Ltd) |
| 9225 | 204605001000027101 | Methyldopa | Methyldopa 250mg Capsule |
| 14390 | 51145001000027101 | Methyldopa | Aldomet 250mg/ 5ml Liquid (Merck Sharp & Dohme Ltd) |
| 18252 | 29695001000027106 | Methyldopa | Metalpha 250mg Tablet (Ashbourne Pharmaceuticals Ltd) |
| 23761 | 8667311000001100 | Methyldopa | Methyldopa 250mg/ 5ml oral suspension |
| 24196 | 54625001000027106 | Methyldopa | Dopamet 250mg Tablet (Berk Pharmaceuticals Ltd) |
| 25275 | 29705001000027102 | Methyldopa | Metalpha 500mg Tablet (Ashbourne Pharmaceuticals Ltd) |
| 25289 | 54635001000027108 | Methyldopa | Dopamet 500mg Tablet (Berk Pharmaceuticals Ltd) |
| 29570 | 54615001000027102 | Methyldopa | Dopamet 125mg Tablet (Berk Pharmaceuticals Ltd) |
| 32913 | 874511000001106 | Methyldopa | Methyldopa 250mg tablets (Actavis UK Ltd) |
| 41661 | 15965001000027104 | Methyldopa | Methyldopa 250mg Tablet (C P Pharmaceuticals Ltd) |
| 43988 | 253711000001107 | Methyldopa | Aldomet 250mg tablets (Aspen Pharma Trading Ltd) |
| 43989 | 73611000001108 | Methyldopa | Aldomet 500mg tablets (Aspen Pharma Trading Ltd) |
| 62513 | 17204211000001105 | Methyldopa | Methyldopa 250mg tablets (Sovereign Medical Ltd) |
| 71110 | 17895811000001103 | Methyldopa | Aldomet 500mg tablets (Sigma Pharmaceuticals Plc) |
| 71385 | 17895611000001102 | Methyldopa | Aldomet 250mg tablets (Sigma Pharmaceuticals Plc) |
| 72819 | 17940911000001101 | Methyldopa | Methyldopa 250mg tablets (Phoenix Healthcare Distribution Ltd) |
| 73640 | 87911000001103 | Methyldopa | Methyldopa 250mg tablets (Sandoz Ltd) |
| 77825 | | Methyldopa | Methyldopa 250mg tablets (A A H Pharmaceuticals Ltd) |
| 10713 | 6855001000027101 | Methoserpidine | Decaserpyl 5mg Tablet (Roussel Laboratories Ltd) |
| 10714 | 124465001000027102 | Methoserpidine | Methoserpidine 5mg Tablet |
| 25393 | 6865001000027105 | Methoserpidine | Decaserpyl 10mg Tablet (Roussel Laboratories Ltd) |
| 29187 | 124475001000027109 | Methoserpidine | Methoserpidine 10mg Tablet |
| 63780 | 23707811000001104 | Macitentan | Macitentan 10mg tablets |
| 54940 | 14696611000001108 | Ketanserin | Ketanserin 20mg tablets |
| 2117 | 3354611000001100 | Indoramin | Doralese Tiltab 20mg tablets (Chemidex Pharma Ltd) |
| 2816 | 318739007 | Indoramin | Indoramin 20mg tablets |
| 5815 | 318740009 | Indoramin | Indoramin 25mg tablets |
| 9019 | 118045001000027102 | Indoramin | Indoramin 50mg Tablet |

| | | | |
|---|---|---|---|
| 11394 | 2415001000027104 | Indoramin | Baratol 25mg Tablet (Shire Pharmaceuticals Ltd) |
| 16198 | 2425001000027108 | Indoramin | Baratol 50mg Tablet (Shire Pharmaceuticals Ltd) |
| 40256 | 3689111000001107 | Indoramin | Baratol 25mg tablets (Amdipharm Plc) |
| 504 | 34193811000001100 | Hydralazine | Hydralazine 20mg powder for solution for injection ampoules |
| 573 | 318649003 | Hydralazine | Hydralazine 25mg tablets |
| 1296 | 318650003 | Hydralazine | Hydralazine 50mg tablets |
| 2362 | 657011000001101 | Hydralazine | Apresoline 25mg tablets (AMCo) |
| 2680 | 1655001000027106 | Hydralazine | Apresoline 50mg Tablet (Sovereign Medical Ltd) |
| 13317 | 3925011000001100 | Hydralazine | Apresoline 20mg powder for solution for injection ampoules (AMCo) |
| 18861 | 8528611000001105 | Hydralazine | Hydralazine 10mg/ 5ml oral suspension |
| 31220 | 193711000001103 | Hydralazine | Hydralazine 25mg tablets (A A H Pharmaceuticals Ltd) |
| 41639 | 147111000001103 | Hydralazine | Hydralazine 50mg tablets (Actavis UK Ltd) |
| 43500 | 364711000001106 | Hydralazine | Hydralazine 25mg tablets (Actavis UK Ltd) |
| 59512 | 8528711000001101 | Hydralazine | Hydralazine 50mg/ 5ml oral solution |
| 61116 | 8528811000001109 | Hydralazine | Hydralazine 50mg/ 5ml oral suspension |
| 63652 | 245915001000027107 | Hydralazine | Hydralazine Tablet |
| 64253 | 12537811000001103 | Hydralazine | Hydralazine 5mg/ 5ml oral suspension |
| 70655 | 8581011000001103 | Hydralazine | Hydralazine 25mg/ 5ml oral suspension |
| 71097 | 19819011000001104 | Hydralazine | Hydralazine 50mg tablets (Almus Pharmaceuticals Ltd) |
| 71256 | 30172911000001106 | Hydralazine | Hydralazine 20mg powder for concentrate for solution for injection ampoules (AMCo) |
| 74749 | 414426001 | Hydralazine | Hydralazine 10mg tablets |
| 75259 | 12536611000001104 | Hydralazine | Hydralazine 1mg/ 5ml oral suspension |
| 7922 | 15305001000027108 | Guanethidine Monosulphate | Ismelin 10mg Tablet (Sovereign Medical Ltd) |
| 7923 | 189785001000027102 | Guanethidine Monosulphate | Guanethidine 10mg Tablet |
| 13379 | 15315001000027105 | Guanethidine Monosulphate | Ismelin 25mg Tablet (Sovereign Medical Ltd) |
| 17291 | 189795001000027103 | Guanethidine Monosulphate | Guanethidine 25mg Tablet |
| 30127 | 15325001000027101 | Guanethidine monosulfate | Ismelin 10mg/ ml Injection (Sovereign Medical Ltd) |
| 31080 | 36053311000001103 | Guanethidine monosulfate | Guanethidine 10mg/ 1ml solution for injection ampoules |
| 48189 | 4369611000001106 | Guanethidine monosulfate | Ismelin 10mg/ 1ml solution for injection ampoules (Amdipharm Plc) |
| 119 | 318781001 | Doxazosin mesilate | Doxazosin 1mg tablets |
| 493 | 318782008 | Doxazosin mesilate | Doxazosin 2mg tablets |
| 582 | 134456001 | Doxazosin mesilate | Doxazosin 4mg modified-release tablets |
| 755 | 123911000001106 | Doxazosin mesilate | Cardura XL 4mg tablets (Pfizer Ltd) |
| 1294 | 318783003 | Doxazosin mesilate | Doxazosin 4mg tablets |
| 4449 | 907711000001109 | Doxazosin mesilate | Cardura 1mg tablets (Pfizer Ltd) |
| 4802 | 41811000001109 | Doxazosin mesilate | Cardura 2mg tablets (Pfizer Ltd) |
| 5496 | 135921004 | Doxazosin mesilate | Doxazosin 8mg modified-release tablets |
| 5618 | 873411000001109 | Doxazosin mesilate | Cardura XL 8mg tablets (Pfizer Ltd) |
| 7547 | 4857711000001103 | Doxazosin mesilate | Doxadura 2mg tablets (Discovery Pharmaceuticals) |
| 7549 | 4857511000001108 | Doxazosin mesilate | Doxadura 1mg tablets (Discovery Pharmaceuticals) |
| 8086 | 159565001000027104 | Doxazosin mesilate | Cardura 4mg Tablet (Pfizer Ltd) |
| 10088 | 4858111000001103 | Doxazosin mesilate | Doxadura 4mg tablets (Discovery Pharmaceuticals) |
| 19193 | 481211000001106 | Doxazosin mesilate | Doxazosin 2mg tablets (Teva UK Ltd) |

| | | | |
|---|---|---|---|
| 19216 | 525011000001104 | Doxazosin mesilate | Doxazosin 4mg tablets (IVAX Pharmaceuticals UK Ltd) |
| 20369 | 8483411000001101 | Doxazosin mesilate | Doxazosin 1mg/ 5ml oral suspension |
| 25487 | 904511000001107 | Doxazosin mesilate | Cascor 2mg tablets (Ranbaxy (UK) Ltd) |
| 25551 | 179311000001107 | Doxazosin mesilate | Cascor 4mg tablets (Ranbaxy (UK) Ltd) |
| 33094 | 674011000001107 | Doxazosin mesilate | Doxazosin 2mg tablets (Mylan) |
| 34342 | 647711000001101 | Doxazosin mesilate | Doxazosin 1mg tablets (Teva UK Ltd) |
| 34553 | 221311000001106 | Doxazosin mesilate | Doxazosin 4mg tablets (Mylan) |
| 34601 | 565111000001107 | Doxazosin mesilate | Doxazosin 1mg tablets (Mylan) |
| 34625 | 554711000001100 | Doxazosin mesilate | Doxazosin 2mg tablets (A A H Pharmaceuticals Ltd) |
| 34715 | 228711000001100 | Doxazosin mesilate | Doxazosin 1mg tablets (A A H Pharmaceuticals Ltd) |
| 35272 | 11269911000001101 | Doxazosin mesilate | Doxadura XL 4mg tablets (Discovery Pharmaceuticals) |
| 35603 | 8483511000001102 | Doxazosin mesilate | Doxazosin 4mg/ 5ml oral suspension |
| 36023 | 24461500100002710З | Doxazosin mesilate | Cardozin xl 4mg Tablet (Hillcross Pharmaceuticals Ltd) |
| 36740 | 11098311000001104 | Doxazosin mesilate | Slocinx XL 4mg tablets (Zentiva) |
| 37243 | 248505001000027109 | Doxazosin mesilate | Cardozin xl 4mg Tablet (Teva UK Ltd) |
| 38461 | 11752411000001104 | Doxazosin mesilate | Cardozin XL 4mg tablets (Arrow Generics Ltd) |
| 40678 | 653011000001107 | Doxazosin mesilate | Doxazosin 4mg tablets (Teva UK Ltd) |
| 40891 | 840911000001105 | Doxazosin mesilate | Doxazosin 2mg tablets (IVAX Pharmaceuticals UK Ltd) |
| 41543 | 854611000001108 | Doxazosin mesilate | Doxazosin 1mg tablets (IVAX Pharmaceuticals UK Ltd) |
| 43695 | 11757711000001107 | Doxazosin mesilate | Colixil XL 4mg tablets (Sandoz Ltd) |
| 45040 | 17338211000001104 | Doxazosin mesilate | Larbex XL 4mg tablets (Teva UK Ltd) |
| 45265 | 198435001000027102 | Doxazosin mesilate | Doxazosin sr 4mg Tablet (Generics (UK) Ltd) |
| 45328 | 462611000001100 | Doxazosin mesilate | Doxazosin 1mg tablets (Sandoz Ltd) |
| 45342 | 761311000001108 | Doxazosin mesilate | Doxazosin 4mg tablets (Sandoz Ltd) |
| 45583 | 11554711000001104 | Doxazosin mesilate | Doxazosin 2mg tablets (Dexcel-Pharma Ltd) |
| 46066 | 18197411000001107 | Doxazosin mesilate | Cardozin XL 4mg tablets (Almus Pharmaceuticals Ltd) |
| 46526 | 18164911000001108 | Doxazosin mesilate | Raporsin XL 4mg tablets (Actavis UK Ltd) |
| 47807 | 203155001000027108 | Doxazosin mesilate | Doxazosin xl 4mg Tablet (Hillcross Pharmaceuticals Ltd) |
| 48150 | 4250111000001105 | Doxazosin mesilate | Doxazosin 1mg tablets (Actavis UK Ltd) |
| 50467 | 430311000001102 | Doxazosin mesilate | Doxazosin 2mg tablets (Alliance Healthcare (Distribution) Ltd) |
| 51685 | 4250511000001101 | Doxazosin mesilate | Doxazosin 4mg tablets (Actavis UK Ltd) |
| 53033 | 13811611000001108 | Doxazosin mesilate | Doxzogen XL 4mg tablets (Mylan) |
| 53322 | 16050211000001103 | Doxazosin mesilate | Doxazosin 4mg tablets (Bristol Laboratories Ltd) |
| 54785 | 19730511000001106 | Doxazosin mesilate | Doxazosin 4mg tablets (Medreich Plc) |
| 55906 | 11554511000001109 | Doxazosin mesilate | Doxazosin 1mg tablets (Dexcel-Pharma Ltd) |
| 55916 | 870611000001103 | Doxazosin mesilate | Doxazosin 1mg tablets (Alliance Healthcare (Distribution) Ltd) |
| 56145 | 4250311000001107 | Doxazosin mesilate | Doxazosin 2mg tablets (Actavis UK Ltd) |
| 57074 | 15083011000001109 | Doxazosin mesilate | Doxazosin 2mg tablets (Sigma Pharmaceuticals Plc) |
| 57448 | 588211000001106 | Doxazosin mesilate | Doxazosin 4mg tablets (A A H Pharmaceuticals Ltd) |
| 57784 | 12083611000001104 | Doxazosin mesilate | Doxazosin 2mg/ 5ml oral suspension |
| 58276 | 19730311000001100 | Doxazosin mesilate | Doxazosin 2mg tablets (Medreich Plc) |
| 58325 | 17894411000001101 | Doxazosin mesilate | Doxazosin 4mg tablets (Phoenix Healthcare Distribution Ltd) |
| 59209 | 252111000001100 | Doxazosin mesilate | Doxazosin 1mg tablets (Kent Pharmaceuticals Ltd) |

| | | | |
|---|---|---|---|
| 59862 | 11554911000001102 | Doxazosin mesilate | Doxazosin 4mg tablets (Dexcel-Pharma Ltd) |
| 60200 | 23970811000001104 | Doxazosin mesilate | Doxazosin 4mg tablets (DE Pharmaceuticals) |
| 60319 | 16049611000001105 | Doxazosin mesilate | Doxazosin 1mg tablets (Bristol Laboratories Ltd) |
| 61066 | 16049911000001104 | Doxazosin mesilate | Doxazosin 2mg tablets (Bristol Laboratories Ltd) |
| 61123 | 23466611000001105 | Doxazosin mesilate | Doxazosin 4mg/ 5ml oral solution |
| 61283 | 578111000001109 | Doxazosin mesilate | Doxazosin 4mg tablets (Alliance Healthcare (Distribution) Ltd) |
| 62019 | 9792611000001102 | Doxazosin mesilate | Doxazosin 1mg tablets (Almus Pharmaceuticals Ltd) |
| 62158 | 979321000001105 | Doxazosin mesilate | Doxazosin 4mg tablets (Almus Pharmaceuticals Ltd) |
| 62351 | 17894211000001100 | Doxazosin mesilate | Doxazosin 2mg tablets (Phoenix Healthcare Distribution Ltd) |
| 63158 | 9792911000001108 | Doxazosin mesilate | Doxazosin 2mg tablets (Almus Pharmaceuticals Ltd) |
| 63314 | 17198011000001109 | Doxazosin mesilate | Doxazosin 1mg tablets (Sovereign Medical Ltd) |
| 64233 | 23591811000001109 | Doxazosin mesilate | Doxazosin 1mg tablets (Waymade Healthcare Plc) |
| 65159 | 23592211000001101 | Doxazosin mesilate | Doxazosin 4mg tablets (Waymade Healthcare Plc) |
| 65853 | 30122411000001102 | Doxazosin mesilate | Doxazosin 1mg tablets (Mawdsley-Brooks & Company Ltd) |
| 66065 | 225411000001101 | Doxazosin mesilate | Doxazosin 2mg tablets (Kent Pharmaceuticals Ltd) |
| 68022 | 17198511000001101 | Doxazosin mesilate | Doxazosin 4mg tablets (Sovereign Medical Ltd) |
| 68161 | 30123811000001108 | Doxazosin mesilate | Doxazosin 4mg tablets (Mawdsley-Brooks & Company Ltd) |
| 69319 | 23970611000001103 | Doxazosin mesilate | Doxazosin 2mg tablets (DE Pharmaceuticals) |
| 69757 | 12084811000001102 | Doxazosin mesilate | Doxazosin 8mg/ 5ml oral suspension |
| 71405 | 13846611000001108 | Doxazosin mesilate | Cardura XL 4mg tablets (DE Pharmaceuticals) |
| 72346 | 12083511000001103 | Doxazosin mesilate | Doxazosin 2mg/ 5ml oral solution |
| 72348 | 24509711000001103 | Doxazosin mesilate | Doxazosin 1mg/ 5ml oral solution |
| 73116 | 17198311000001107 | Doxazosin mesilate | Doxazosin 2mg tablets (Sovereign Medical Ltd) |
| 73637 | 4469911000001108 | Doxazosin mesilate | Doxazosin 1mg tablets (Sterwin Medicines) |
| 74000 | 5346711000001108 | Doxazosin mesilate | Cardura XL 4mg tablets (Waymade Healthcare Plc) |
| 74567 | 421069003 | Doxazosin mesilate | Doxazosin 8mg tablets |
| 74831 | 15083211000001104 | Doxazosin mesilate | Doxazosin 4mg tablets (Sigma Pharmaceuticals Plc) |
| 76149 | | Doxazosin mesilate | Doxazosin 5mg/ 5ml oral suspension |
| 77362 | | Doxazosin mesilate | Cardura 2mg tablets (Sigma Pharmaceuticals Plc) |
| 77436 | | Doxazosin mesilate | Cardura XL 4mg tablets (Mawdsley-Brooks & Company Ltd) |
| 4374 | 108525001000027101 | Debrisoquine Sulphate | Debrisoquine 20mg tablets |
| 8342 | 6915001000027102 | Debrisoquine Sulphate | Declinax 20mg Tablet (Roche Products Ltd) |
| 4375 | 318724006 | Debrisoquine sulfate | Debrisoquine 10mg tablets |
| 7911 | 6905001000027104 | Debrisoquine sulfate | Declinax 10mg Tablet (Roche Products Ltd) |
| 338 | 322840006 | Clonidine | Clonidine 25microgram tablets |
| 2630 | 344511000001105 | Clonidine | Dixarit 25microgram tablets (Boehringer Ingelheim Ltd) |
| 2878 | 318667005 | Clonidine | Clonidine 100microgram tablets |
| 4215 | 215111000001101 | Clonidine | Catapres 100microgram tablets (Boehringer Ingelheim Ltd) |
| 5289 | 36089511000001100 | Clonidine | Clonidine 250microgram modified-release capsules |
| 6694 | 318668000 | Clonidine | Clonidine 300microgram tablets |
| 8296 | 4536811000001109 | Clonidine | Catapres PL Perlongets 250microgram capsules (Boehringer Ingelheim Ltd) |
| 16248 | 36089211000001103 | Clonidine | Clonidine 150micrograms/ 1ml solution for injection ampoules |
| 23380 | 368711000001103 | Clonidine | Catapres 300microgram tablets (Boehringer Ingelheim Ltd) |

| | | | |
|---|---|---|---|
| 30293 | 364911000001108 | Clonidine | Catapres 150micrograms/ 1ml solution for injection ampoules (Boehringer Ingelheim Ltd) |
| 33093 | 3963611000001104 | Clonidine | Clonidine 25microgram tablets (Sandoz Ltd) |
| 45578 | 4920211000001104 | Clonidine | Clonidine 25microgram tablets (A A H Pharmaceuticals Ltd) |
| 49684 | 10448511000001104 | Clonidine | Clonidine 100micrograms/ 24hours transdermal patches |
| 52555 | 8398511000001100 | Clonidine | Clonidine 50micrograms/ 5ml oral solution |
| 53142 | 8398611000001101 | Clonidine | Clonidine 50micrograms/ 5ml oral suspension |
| 54467 | 10449111000001101 | Clonidine | Clonidine 300micrograms/ 24hours transdermal patches |
| 55797 | 8398311000001106 | Clonidine | Clonidine 25micrograms/ 5ml oral solution |
| 58090 | 11814111000001107 | Clonidine | Clonidine 75micrograms/ 5ml oral solution |
| 58529 | 7660211000001104 | Clonidine | Clonidine 200micrograms/ 24hours transdermal patches |
| 60089 | 21922811000001105 | Clonidine | Clonidine 25microgram tablets (Waymade Healthcare Plc) |
| 60136 | 11812911000001101 | Clonidine | Clonidine 100micrograms/ 5ml oral solution |
| 61256 | 8426311000001108 | Clonidine | Clonidine 5micrograms/ 5ml oral suspension |
| 61710 | 18402811000001107 | Clonidine | Clonidine 25microgram tablets (Teva UK Ltd) |
| 63971 | 15077611000001101 | Clonidine | Clonidine 25microgram tablets (Sigma Pharmaceuticals Plc) |
| 64284 | 16159511000001104 | Clonidine | Dixarit 25microgram tablets (Lexon (UK) Ltd) |
| 67200 | 8397911000001106 | Clonidine | Clonidine 10micrograms/ 5ml oral solution |
| 72130 | 11813011000001109 | Clonidine | Clonidine 100micrograms/ 5ml oral suspension |
| 72227 | 13867511000001104 | Clonidine | Dixarit 25microgram tablets (DE Pharmaceuticals) |
| 72807 | 11813311000001107 | Clonidine | Clonidine 15micrograms/ 5ml oral solution |
| 74534 | 32392011000001103 | Clonidine | Clonidine 250micrograms/ 5ml oral suspension |
| 75207 | 36392711000001102 | Clonidine | Clonidine 50micrograms/ 5ml oral solution sugar free |
| 76766 | | Clonidine | Catapres TTS 3 patches (Imported (Germany)) |
| 77591 | | Clonidine | Clonidine 12.5micrograms/ 5ml oral suspension |
| 29560 | 407815004 | Bosentan monohydrate | Bosentan 62.5mg tablets |
| 29561 | 407816003 | Bosentan monohydrate | Bosentan 125mg tablets |
| 47654 | 4765211000001107 | Bosentan monohydrate | Tracleer 62.5mg tablets (Actelion Pharmaceuticals UK Ltd) |
| 58632 | 4765711000001100 | Bosentan monohydrate | Tracleer 125mg tablets (Actelion Pharmaceuticals UK Ltd) |
| 10879 | 122435001000027102 | Betanidine Sulphate | Bethanidine sulphate 10mg tablets |
| 14442 | 10505001000027103 | Betanidine Sulphate | Esbatal 50mg Tablet (Wellcome Medical Division) |
| 18247 | 10495001000027109 | Betanidine Sulphate | Esbatal 10mg Tablet (Wellcome Medical Division) |
| 28676 | 122445001000027101 | Betanidine Sulphate | Bethanidine sulphate 50mg tablets |
| 29443 | 122475001000027100 | Betanidine Sulphate | Bendogen 10mg Tablet (Lagap) |
| 40421 | 122485001000027105 | Betanidine Sulphate | Bendogen 50mg Tablet (Lagap) |
| 40527 | 428480001 | Ambrisentan | Ambrisentan 10mg tablets |
| 40528 | 429662009 | Ambrisentan | Ambrisentan 5mg tablets |
| 214 | 168745001000027105 | | HYDRALAZINE 1 MG SYR |
| 445 | 4078311000001100 | | Prazosin 1mg tablets and Prazosin 500microgram tablets |
| 2649 | 70895001000027105 | | METHYLDOPA 250 MG CAP |
| 4507 | 145525001000027109 | | HYDRALAZINE 12.5 MG TAB |
| 19635 | | | DIXARIT |
| 20808 | | | CATAPRES PERLONGETS |
| 21749 | 168685001000027105 | | HYDRALAZINE 100 MG TAB |

| | | | |
|---|---|---|---|
| 23746 | 179545001000027108 | | HYDRALAZINE 10 MG TAB |
| 25088 | | | CATAPRES |
| 25836 | 141085001000027104 | | METHYLDOPA 200 MG TAB |
| 27894 | | | CLONIDINE |
| 28790 | | | CATAPRES |
| 31971 | 145575001000027108 | | HYDRALAZINE 6.25 MG SYR |
| **Beta-blockers** | | | |
| 7852 | 51195001000027109 | Timolol maleate | Blocadren 10mg Tablet (Merck Sharp & Dohme Ltd) |
| 7853 | 318534000 | Timolol maleate | Timolol 10mg tablets |
| 12037 | 3315001000027104 | Timolol maleate | Betim 10mg Tablet (ICN Pharmaceuticals France S.A.) |
| 29610 | 72811000001104 | Timolol maleate | Betim 10mg tablets (Meda Pharmaceuticals Ltd) |
| 786 | 318525005 | Sotalol | Sotalol 40mg tablets |
| 1572 | 318526006 | Sotalol | Sotalol 80mg tablets |
| 4004 | 463811000001109 | Sotalol | Sotacor 80mg tablets (Bristol-Myers Squibb Pharmaceuticals Ltd) |
| 5858 | 900511000001103 | Sotalol | Beta-Cardone 40mg tablets (Focus Pharmaceuticals Ltd) |
| 6751 | 104011000001101 | Sotalol | Beta-Cardone 80mg tablets (Focus Pharmaceuticals Ltd) |
| 9292 | 318528007 | Sotalol | Sotalol 160mg tablets |
| 11380 | 446211000001108 | Sotalol | Sotacor 160mg tablets (Bristol-Myers Squibb Pharmaceuticals Ltd) |
| 13051 | 318527002 | Sotalol | Sotalol 200mg tablets |
| 13487 | 447111000001104 | Sotalol | Beta-Cardone 200mg tablets (Focus Pharmaceuticals Ltd) |
| 17679 | 141245001000027103 | Sotalol | Sotalol 10mg/ ml injection |
| 24635 | 141365001000027103 | Sotalol | Sotacor 10mg/ ml Injection (Bristol-Myers Squibb Pharmaceuticals Ltd) |
| 27727 | 141255001000027100 | Sotalol | Sotalol 2mg/ ml injection |
| 33578 | 694711000001100 | Sotalol | Sotacor 40mg/ 4ml solution for injection ampoules (Bristol-Myers Squibb Pharmaceuticals Ltd) |
| 34371 | 936211000001109 | Sotalol | Sotalol 40mg tablets (A A H Pharmaceuticals Ltd) |
| 34520 | 680311000001106 | Sotalol | Sotalol 80mg tablets (Mylan) |
| 34600 | 3659211000001106 | Sotalol | Sotalol 40mg tablets (Teva UK Ltd) |
| 34640 | 89165001000027103 | Sotalol | Sotalol 40mg Tablet (Tillomed Laboratories Ltd) |
| 34690 | 170011000001101 | Sotalol | Sotalol 80mg tablets (Sandoz Ltd) |
| 35710 | 8726911000001102 | Sotalol | Sotalol 25mg/ 5ml oral suspension |
| 38498 | 35930811000001109 | Sotalol | Sotalol 40mg/ 4ml solution for injection ampoules |
| 39423 | 909811000001109 | Sotalol | Sotalol 80mg tablets (A A H Pharmaceuticals Ltd) |
| 43549 | 4230411000001104 | Sotalol | Sotalol 40mg tablets (IVAX Pharmaceuticals UK Ltd) |
| 51492 | 8726811000001107 | Sotalol | Sotalol 25mg/ 5ml oral solution |
| 70161 | 8727111000001102 | Sotalol | Sotalol 40mg/ 5ml oral solution |
| 70162 | 8727211000001108 | Sotalol | Sotalol 40mg/ 5ml oral suspension |
| 70734 | 11427411000001105 | Sotalol | Sotalol 40mg tablets (Almus Pharmaceuticals Ltd) |
| 71479 | 16067611000001105 | Sotalol | Sotalol 40mg tablets (Bristol Laboratories Ltd) |
| 72347 | 8727511000001106 | Sotalol | Sotalol 80mg/ 5ml oral suspension |
| 73481 | 3658411000001102 | Sotalol | Sotalol 160mg tablets (Teva UK Ltd) |
| 74628 | 3659411000001105 | Sotalol | Sotalol 80mg tablets (Teva UK Ltd) |
| 74829 | 17799811000001102 | Sotalol | Sotalol 40mg tablets (Phoenix Healthcare Distribution Ltd) |

| | | | |
|---|---|---|---|
| 75060 | 243155001000027107 | Sotalol | Sotalol oral solution |
| 75821 | | Sotalol | Sotalol 40mg tablets (Alliance Healthcare (Distribution) Ltd) |
| 220 | 89655001000027106 | Propranolol | Propranolol 5mg/ 5ml oral solution |
| 297 | 318352004 | Propranolol | Propranolol 10mg tablets |
| 707 | 318353009 | Propranolol | Propranolol 40mg tablets |
| 769 | 318407001 | Propranolol | Propranolol 80mg modified-release capsules |
| 940 | 318354003 | Propranolol | Propranolol 80mg tablets |
| 1006 | 226211000001106 | Propranolol | Half Inderal LA 80mg capsules (AstraZeneca UK Ltd) |
| 1048 | 821011000001100 | Propranolol | Inderal 80mg tablets (AstraZeneca UK Ltd) |
| 1050 | 930211000001107 | Propranolol | Inderal 40mg tablets (AstraZeneca UK Ltd) |
| 1448 | 318406005 | Propranolol | Propranolol 160mg modified-release capsules |
| 2414 | 567911000001100 | Propranolol | Inderal 10mg tablets (AstraZeneca UK Ltd) |
| 3005 | 154311000001108 | Propranolol | Inderal LA 160mg capsules (AstraZeneca UK Ltd) |
| 3087 | 15356211000001102 | Propranolol | Propranolol 40mg/ 5ml oral solution sugar free |
| 3167 | 318355002 | Propranolol | Propranolol 160mg tablets |
| 3827 | 121565001000027103 | Propranolol | Propanix 40mg Tablet (Ashbourne Pharmaceuticals Ltd) |
| 5478 | 35932311000001101 | Propranolol | Propranolol 10mg/ 5ml oral solution sugar free |
| 8331 | 51875001000027109 | Propranolol | Inderal 160mg Tablet (AstraZeneca UK Ltd) |
| 8978 | 120695001000027100 | Propranolol | Propanix 160mg Modified-release capsule (Ashbourne Pharmaceuticals Ltd) |
| 9185 | 196405001000027103 | Propranolol | Propranolol 80mg/ 5ml oral solution |
| 10294 | 4371111000001106 | Propranolol | Inderal 1mg/ 1ml solution for injection ampoules (AstraZeneca UK Ltd) |
| 11711 | 196395001000027106 | Propranolol | Propranolol 50mg/ 5ml oral solution |
| 12495 | 54385001000027105 | Propranolol | Berkolol 10mg Tablet (Berk Pharmaceuticals Ltd) |
| 14552 | 121555001000027107 | Propranolol | Propanix 10mg Tablet (Ashbourne Pharmaceuticals Ltd) |
| 14808 | 573711000001104 | Propranolol | Bedranol SR 80mg capsules (Sandoz Ltd) |
| 15619 | 72255001000027102 | Propranolol | Half-betadur cr 80mg Capsule (Monmouth Pharmaceuticals Ltd) |
| 17082 | 295911000001102 | Propranolol | Syprol 5mg/ 5ml oral solution (Rosemont Pharmaceuticals Ltd) |
| 20468 | 724111000001103 | Propranolol | Half Beta-Prograne 80mg modified-release capsules (Tillomed Laboratories Ltd) |
| 21838 | 121575001000027105 | Propranolol | Propanix 80mg Tablet (Ashbourne Pharmaceuticals Ltd) |
| 21839 | 54405001000027101 | Propranolol | Berkolol 80mg Tablet (Berk Pharmaceuticals Ltd) |
| 21866 | 54395001000027109 | Propranolol | Berkolol 40mg Tablet (Berk Pharmaceuticals Ltd) |
| 22208 | 227945001000027104 | Propranolol | Half propanix la 80mg Modified-release capsule (Ashbourne Pharmaceuticals Ltd) |
| 23326 | 175605001000027100 | Propranolol | Betadur cr 160mg Modified-release capsule (Monmouth Pharmaceuticals Ltd) |
| 23587 | 138495001000027100 | Propranolol | Sloprolol 160mg Capsule (C P Pharmaceuticals Ltd) |
| 24218 | 54445001000027106 | Propranolol | Berkolol 160mg Tablet (Berk Pharmaceuticals Ltd) |
| 25359 | 2899211000001100 | Propranolol | Rapranol SR 160mg capsules (Ranbaxy (UK) Ltd) |
| 25367 | 2899011000001105 | Propranolol | Rapranol SR 80mg capsules (Ranbaxy (UK) Ltd) |
| 26228 | 231365001000027104 | Propranolol | Propanix LA 160mg Modified-release capsule (Ashbourne Pharmaceuticals Ltd) |
| 26229 | 510611000001108 | Propranolol | Beta-Prograne 160mg modified-release capsules (Tillomed Laboratories Ltd) |
| 26255 | 207165001000027106 | Propranolol | Lopranol la 160mg Capsule (Opus Pharmaceuticals Ltd) |
| 26895 | 28711000001109 | Propranolol | Syprol 10mg/ 5ml oral solution (Rosemont Pharmaceuticals Ltd) |
| 27486 | 35932611000001106 | Propranolol | Propranolol 1mg/ 1ml solution for injection ampoules |
| 27700 | 92011000001103 | Propranolol | Propranolol 40mg tablets (Actavis UK Ltd) |

| | | | |
|---|---|---|---|
| 27964 | 75515001000027103 | Propranolol | Apsolol 40mg Tablet (Approved Prescription Services Ltd) |
| 28048 | 90305001000027107 | Propranolol | Angilol 10mg Tablet (DDSA Pharmaceuticals Ltd) |
| 28128 | 49655001000027103 | Propranolol | Propranolol 80mg Modified-release capsule (Actavis UK Ltd) |
| 28788 | 232405001000027109 | Propranolol | Half propatard la 80mg Modified-release capsule (Galen Ltd) |
| 28996 | 549611000001106 | Propranolol | Bedranol SR 160mg capsules (Sandoz Ltd) |
| 29763 | 120685001000027104 | Propranolol | Propanix 160mg Tablet (Ashbourne Pharmaceuticals Ltd) |
| 31214 | 694511000001105 | Propranolol | Propranolol 80mg tablets (Mylan) |
| 31776 | 23611000001107 | Propranolol | Propranolol 40mg tablets (Mylan) |
| 31833 | 90325001000027100 | Propranolol | Angilol 80mg Tablet (DDSA Pharmaceuticals Ltd) |
| 32162 | 90295001000027100 | Propranolol | Propranolol 80mg Modified-release capsule (Lagap) |
| 33376 | 208225001000027105 | Propranolol | Probeta LA 160mg Capsule (Trinity Pharmaceuticals Ltd) |
| 33602 | 240711000001100 | Propranolol | Slo-Pro 160mg capsules (Mylan) |
| 33644 | 763811000001100 | Propranolol | Propranolol 80mg tablets (A A H Pharmaceuticals Ltd) |
| 33836 | 75555001000027108 | Propranolol | Apsolol 160mg Tablet (Approved Prescription Services Ltd) |
| 34185 | 65085001000027102 | Propranolol | Propranolol LA 80mg Modified-release capsule (Approved Prescription Services Ltd) |
| 34208 | 30275001000027102 | Propranolol | Propranolol SR 160mg Modified-release capsule (C P Pharmaceuticals Ltd) |
| 34214 | 696011000001107 | Propranolol | Propranolol 160mg tablets (Actavis UK Ltd) |
| 34378 | 695911000001104 | Propranolol | Propranolol 10mg tablets (A A H Pharmaceuticals Ltd) |
| 34783 | 98911000001101 | Propranolol | Propranolol 10mg tablets (Actavis UK Ltd) |
| 34804 | 212711000001109 | Propranolol | Propranolol 10mg tablets (Teva UK Ltd) |
| 34867 | 63965001000027101 | Propranolol | Propranolol 80mg Capsule (IVAX Pharmaceuticals UK Ltd) |
| 34868 | 859211000001105 | Propranolol | Propranolol 40mg tablets (Teva UK Ltd) |
| 34884 | 141395001000027101 | Propranolol | Propranolol 160mg Modified-release capsule (Sandoz Ltd) |
| 34945 | 90285001000027104 | Propranolol | Propranolol 160mg Modified-release capsule (Lagap) |
| 34949 | 49665001000027107 | Propranolol | Propranolol 160mg Modified-release capsule (Actavis UK Ltd) |
| 35938 | 168311000001107 | Propranolol | Propranolol 80mg modified-release capsules (A A H Pharmaceuticals Ltd) |
| 36576 | 397911000001104 | Propranolol | Propranolol 10mg tablets (Mylan) |
| 36603 | 22555001000027104 | Propranolol | Propranolol SR 160mg Modified-release capsule (Hillcross Pharmaceuticals Ltd) |
| 38433 | 60395001000027107 | Propranolol | Propranolol 50mg/ 5ml Oral solution (Rosemont Pharmaceuticals Ltd) |
| 39233 | 8393911000001102 | Propranolol | Propranolol 80mg modified-release capsules (Teva UK Ltd) |
| 40241 | 65075001000027107 | Propranolol | Propranolol LA 160mg Capsule (Approved Prescription Services Ltd) |
| 41555 | 188311000001109 | Propranolol | Propranolol 40mg tablets (A A H Pharmaceuticals Ltd) |
| 42152 | 823311000001101 | Propranolol | Syprol 50mg/ 5ml oral solution (Rosemont Pharmaceuticals Ltd) |
| 43525 | 395011000001105 | Propranolol | Propranolol 10mg tablets (IVAX Pharmaceuticals UK Ltd) |
| 45297 | 502911000001101 | Propranolol | Propranolol 40mg tablets (IVAX Pharmaceuticals UK Ltd) |
| 45343 | 30285001000027107 | Propranolol | Propranolol SR 80mg Modified-release capsule (C P Pharmaceuticals Ltd) |
| 45494 | 9808311000001105 | Propranolol | Propranolol 10mg tablets (Almus Pharmaceuticals Ltd) |
| 45765 | 15307311000001100 | Propranolol | Syprol 40mg/ 5ml oral solution (Rosemont Pharmaceuticals Ltd) |
| 45877 | 15610411000001104 | Propranolol | Beta-Prograne 160mg modified-release capsules (Teva UK Ltd) |
| 46363 | 15610611000001101 | Propranolol | Half Beta-Prograne 80mg modified-release capsules (Teva UK Ltd) |
| 47543 | 18069311000001100 | Propranolol | Half Beta-Prograne 80mg modified-release capsules (Actavis UK Ltd) |
| 47833 | 18694811000001101 | Propranolol | Bedranol SR 80mg capsules (Almus Pharmaceuticals Ltd) |
| 47907 | 18695311000001109 | Propranolol | Bedranol SR 160mg capsules (Almus Pharmaceuticals Ltd) |

| | | | |
|---|---|---|---|
| 48682 | 35932711000001102 | Propranolol | Propranolol 50mg/ 5ml oral solution sugar free |
| 49863 | 35932811000001105 | Propranolol | Propranolol 5mg/ 5ml oral solution sugar free |
| 52136 | 16077500100027108 | Propranolol | Bedranol sr 160mg Capsule (Lagap) |
| 52609 | 14252111000001108 | Propranolol | Inderal LA 160mg capsules (Sigma Pharmaceuticals Plc) |
| 52777 | 587411000001106 | Propranolol | Propranolol 40mg tablets (Kent Pharmaceuticals Ltd) |
| 53177 | 24446500100027101 | Propranolol | Propranolol oral solution |
| 54297 | 13199511000001100 | Propranolol | Propranolol 50mg/ 5ml oral solution |
| 54623 | 18069011000001103 | Propranolol | Beta-Prograne 160mg modified-release capsules (Actavis UK Ltd) |
| 55228 | 14782911000001107 | Propranolol | Propranolol 40mg tablets (Boston Healthcare Ltd) |
| 55416 | 9808611000001100 | Propranolol | Propranolol 40mg tablets (Almus Pharmaceuticals Ltd) |
| 55849 | 582811000001105 | Propranolol | Propranolol 160mg tablets (Mylan) |
| 55949 | 8672711000001106 | Propranolol | Propranolol 40mg/ 5ml oral solution |
| 56173 | 18069311000001100 | Propranolol | Half Beta-Prograne 80mg modified-release capsules (Actavis UK Ltd) |
| 56764 | 21870611000001101 | Propranolol | Propranolol 40mg tablets (Waymade Healthcare Plc) |
| 57063 | 18694811000001101 | Propranolol | Bedranol SR 80mg capsules (Almus Pharmaceuticals Ltd) |
| 57342 | 17919711000001101 | Propranolol | Propranolol 40mg tablets (Phoenix Healthcare Distribution Ltd) |
| 57567 | 13160911000001104 | Propranolol | Propranolol 10mg/ 5ml oral suspension |
| 58297 | 163111000001101 | Propranolol | Propranolol 10mg tablets (Kent Pharmaceuticals Ltd) |
| 58407 | 59811000001107 | Propranolol | Propranolol 80mg tablets (Teva UK Ltd) |
| 58491 | 316911000001101 | Propranolol | Propranolol 40mg tablets (Alliance Healthcare (Distribution) Ltd) |
| 59415 | 23489711000001109 | Propranolol | Propranolol 40mg tablets (Accord Healthcare Ltd) |
| 59597 | 4577511000001105 | Propranolol | Propranolol 160mg modified-release capsules (A A H Pharmaceuticals Ltd) |
| 60565 | 9153911000001101 | Propranolol | Propranolol 40mg tablets (Ranbaxy (UK) Ltd) |
| 60934 | 388811000001109 | Propranolol | Propranolol 80mg modified-release capsules (Kent Pharmaceuticals Ltd) |
| 61727 | 23489111000001108 | Propranolol | Propranolol 10mg tablets (Accord Healthcare Ltd) |
| 62711 | 23601111000001107 | Propranolol | Propranolol 80mg modified-release capsules (Waymade Healthcare Plc) |
| 64160 | 22352811000001103 | Propranolol | Propranolol 5mg/ 5ml oral solution sugar free (AM Distributions (Yorkshire) Ltd) |
| 65435 | 21870211000001103 | Propranolol | Propranolol 10mg tablets (Waymade Healthcare Plc) |
| 65986 | 795211000001104 | Propranolol | Propranolol 10mg tablets (Alliance Healthcare (Distribution) Ltd) |
| 66555 | 14782711000001105 | Propranolol | Propranolol 10mg tablets (Boston Healthcare Ltd) |
| 68400 | 30081311000001101 | Propranolol | Propranolol 160mg tablets (DE Pharmaceuticals) |
| 69661 | 13172311000001106 | Propranolol | Propranolol 3mg/ 5ml oral solution |
| 70680 | 13200011000001100 | Propranolol | Propranolol 6mg/ 5ml oral suspension |
| 70681 | 13171211000001105 | Propranolol | Propranolol 3mg/ 5ml oral suspension |
| 71150 | 13199711000001105 | Propranolol | Propranolol 5mg/ 5ml oral solution |
| 71173 | 30860011000001105 | Propranolol | Propranolol 80mg modified-release capsules (Mawdsley-Brooks & Company Ltd) |
| 72220 | 24412311000001103 | Propranolol | Propranolol 10mg/ 5ml oral solution sugar free (CST Pharma Ltd) |
| 72222 | 30080311000001106 | Propranolol | Propranolol 80mg modified-release capsules (DE Pharmaceuticals) |
| 73653 | 9154111000001102 | Propranolol | Propranolol 80mg tablets (Ranbaxy (UK) Ltd) |
| 73765 | 9153711000001103 | Propranolol | Propranolol 10mg tablets (Ranbaxy (UK) Ltd) |
| 75278 | 13199811000001102 | Propranolol | Propranolol 5mg/ 5ml oral suspension |
| 75289 | 32759511000001108 | Propranolol | Propranolol 5mg/ 5ml oral solution sugar free (Rosemont Pharmaceuticals Ltd) |
| 75312 | 36106711000001107 | Propranolol | Bedranol 80mg tablets (Ennogen Pharma Ltd) |

| | | | |
|---|---|---|---|
| 75507 | | Propranolol | Propranolol 160mg Capsule (IVAX Pharmaceuticals UK Ltd) |
| 76252 | | Propranolol | Propranolol 160mg modified-release capsules (Teva UK Ltd) |
| 76431 | | Propranolol | Cardinol 10mg Tablet (C P Pharmaceuticals Ltd) |
| 76633 | | Propranolol | Propranolol 10mg tablets (DE Pharmaceuticals) |
| 77413 | | Propranolol | Inderal LA 160mg capsules (Waymade Healthcare Plc) |
| 77649 | | Propranolol | Half Inderal LA 80mg capsules (DE Pharmaceuticals) |
| 20169 | 137695001000027105 | Practolol | Practolol 2mg/ ml injection |
| 4588 | 34545001000027108 | Pindolol | Visken 5mg Tablet (Sovereign Medical Ltd) |
| 5284 | 318512002 | Pindolol | Pindolol 5mg tablets |
| 14673 | 318513007 | Pindolol | Pindolol 15mg tablets |
| 20012 | 34555001000027106 | Pindolol | Visken 15mg Tablet (Sovereign Medical Ltd) |
| 32787 | 3887411000001101 | Pindolol | Visken 15mg tablets (AMCo) |
| 35695 | 3706111000001105 | Pindolol | Visken 5mg tablets (AMCo) |
| 55853 | 96905001000027105 | Pindolol | Pindolol 15mg Tablet (Hillcross Pharmaceuticals Ltd) |
| 73413 | 3707011000001107 | Pindolol | Pindolol 5mg tablets (A A H Pharmaceuticals Ltd) |
| 1333 | 318484005 | Oxprenolol | Oxprenolol 40mg tablets |
| 1334 | 36023011000001102 | Oxprenolol | Oxprenolol 160mg modified-release tablets |
| 2361 | 54835001000027104 | Oxprenolol | Trasicor 80mg Tablet (Novartis Pharmaceuticals UK Ltd) |
| 2780 | 318485006 | Oxprenolol | Oxprenolol 80mg tablets |
| 3516 | 318483004 | Oxprenolol | Oxprenolol 20mg tablets |
| 3748 | 89605001000027103 | Oxprenolol | Oxprenolol 160mg Tablet |
| 4025 | 473011000001107 | Oxprenolol | Slow-Trasicor 160mg tablets (AMCo) |
| 7474 | 54815001000027105 | Oxprenolol | Trasicor 20mg Tablet (Novartis Pharmaceuticals UK Ltd) |
| 8290 | 54825001000027101 | Oxprenolol | Trasicor 40mg Tablet (Novartis Pharmaceuticals UK Ltd) |
| 10777 | 54865001000027109 | Oxprenolol | Trasicor 160mg Tablet (Novartis Pharmaceuticals UK Ltd) |
| 21885 | 195255001000027109 | Oxprenolol | Oxyprenix SR 160mg tablets |
| 24094 | 592911000001104 | Oxprenolol | Trasicor 40mg tablets (Amdipharm Plc) |
| 25644 | 75595001000027102 | Oxprenolol | Apsolox 80mg Tablet (Approved Prescription Services Ltd) |
| 27357 | 9205001000027107 | Oxprenolol | Oxprenolol 40mg Tablet (Actavis UK Ltd) |
| 29180 | 568511000001106 | Oxprenolol | Trasicor 80mg tablets (Amdipharm Plc) |
| 29230 | 90575001000027104 | Oxprenolol | Slow-pren 160mg Tablet (IVAX Pharmaceuticals UK Ltd) |
| 33569 | 9405001000027108 | Oxprenolol | Oxprenolol sr 160mg Modified-release tablet (Hillcross Pharmaceuticals Ltd) |
| 35062 | 3379011000001106 | Oxprenolol | Trasicor 20mg tablets (Amdipharm Plc) |
| 74015 | 17595711000001107 | Oxprenolol | Slow-Trasicor 160mg tablets (Sigma Pharmaceuticals Plc) |
| 77445 | | Oxprenolol | Slow-Trasicor 160mg tablets (DE Pharmaceuticals) |
| 77461 | | Oxprenolol | Slow-Trasicor 160mg tablets (Waymade Healthcare Plc) |
| 751 | 318640004 | Nebivolol | Nebivolol 5mg tablets |
| 7528 | 726011000001109 | Nebivolol | Nebilet 5mg tablets (A. Menarini Farmaceutica Internazionale SRL) |
| 40761 | 432118006 | Nebivolol | Nebivolol 2.5mg tablets |
| 44808 | 16551311000001103 | Nebivolol | Nebivolol 2.5mg tablets (A A H Pharmaceuticals Ltd) |
| 47300 | 15638611000001106 | Nebivolol | Nebivolol 2.5mg tablets (Glenmark Pharmaceuticals Europe Ltd) |
| 54487 | 21266011000001105 | Nebivolol | Nebivolol 2.5mg tablets (Sigma Pharmaceuticals Plc) |
| 59961 | 432159000 | Nebivolol | Nebivolol 10mg tablets |

| | | | |
|---|---|---|---|
| 64703 | 15638211000001109 | Nebivolol | Nebivolol 5mg tablets (Glenmark Pharmaceuticals Europe Ltd) |
| 66559 | 5393311000001106 | Nebivolol | Nebilet 5mg tablets (Waymade Healthcare Plc) |
| 67595 | 24483711000001109 | Nebivolol | Nebivolol 5mg tablets (Almus Pharmaceuticals Ltd) |
| 68677 | 14404711000001108 | Nebivolol | Nebivolol 5mg tablets (Accord Healthcare Ltd) |
| 69115 | 14033711000001109 | Nebivolol | Nebivolol 5mg tablets (A A H Pharmaceuticals Ltd) |
| 71032 | 20312411000001102 | Nebivolol | Nebivolol 5mg tablets (Sandoz Ltd) |
| 74076 | 13801211000001108 | Nebivolol | Nebivolol 5mg tablets (PLIVA Pharma Ltd) |
| 76484 | | Nebivolol | Nebivolol 5mg tablets (Sigma Pharmaceuticals Plc) |
| 2499 | 318481002 | Nadolol | Nadolol 80mg tablets |
| 8935 | 318480001 | Nadolol | Nadolol 40mg tablets |
| 10716 | 3687411000001106 | Nadolol | Corgard 80mg tablets (Sanofi) |
| 13415 | 3686511000001108 | Nadolol | Corgard 40mg tablets (Sanofi-Synthelabo Ltd) |
| 66464 | 12300711000001104 | Nadolol | Nadolol 40mg/ 5ml oral solution |
| 66779 | 16110011000001107 | Nadolol | Nadolol 30mg/ 5ml oral suspension |
| 67424 | 15227211000001106 | Nadolol | Nadolol 80mg/ 5ml oral suspension |
| 72159 | 16131011000001109 | Nadolol | Nadolol 20mg/ 5ml oral suspension |
| 73219 | 12300811000001107 | Nadolol | Nadolol 40mg/ 5ml oral suspension |
| 74929 | 250335001000027108 | Nadolol | Nadolol Oral solution |
| 74930 | 16104311000001104 | Nadolol | Nadolol 30mg/ 5ml oral suspension (Drug Tariff Special Order) |
| 77002 | | Nadolol | Corgard 80mg tablets (Lexon (UK) Ltd) |
| 77945 | | Nadolol | Nadolol 20mg/ 5ml oral solution |
| 739 | 318475005 | Metoprolol tartrate | Metoprolol 50mg tablets |
| 753 | 318474009 | Metoprolol tartrate | Metoprolol 100mg tablets |
| 3344 | 193911000001101 | Metoprolol tartrate | Betaloc 100mg tablets (AstraZeneca UK Ltd) |
| 3474 | 185111000001102 | Metoprolol tartrate | Betaloc-SA 200mg tablets (AstraZeneca UK Ltd) |
| 8068 | 36035411000001108 | Metoprolol tartrate | Metoprolol 200mg modified-release tablets |
| 8071 | 31811000001105 | Metoprolol tartrate | Betaloc 50mg tablets (AstraZeneca UK Ltd) |
| 10429 | 17735001000027102 | Metoprolol tartrate | Lopresor 50mg Tablet (Novartis Pharmaceuticals UK Ltd) |
| 11793 | 8669111000001108 | Metoprolol tartrate | Metoprolol 50mg/ 5ml oral suspension |
| 13499 | 17745001000027101 | Metoprolol tartrate | Lopresor 100mg Tablet (Novartis Pharmaceuticals UK Ltd) |
| 14502 | 3631211000001109 | Metoprolol tartrate | Metoprolol 5mg/ 5ml solution for injection ampoules |
| 20082 | 916111000001108 | Metoprolol tartrate | Lopresor SR 200mg tablets (Recordati Pharmaceuticals Ltd) |
| 24461 | 3615711000001109 | Metoprolol tartrate | Betaloc I.V. 5mg/ 5ml solution for injection ampoules (AstraZeneca UK Ltd) |
| 29762 | 120275001000027107 | Metoprolol tartrate | Mepranix 50mg Tablet (Ashbourne Pharmaceuticals Ltd) |
| 30400 | 120285001000027102 | Metoprolol tartrate | Mepranix 100mg Tablet (Ashbourne Pharmaceuticals Ltd) |
| 32836 | 683311000001100 | Metoprolol tartrate | Metoprolol 50mg tablets (Mylan) |
| 34092 | 89911000001105 | Metoprolol tartrate | Metoprolol 100mg tablets (Teva UK Ltd) |
| 34094 | 393511000001101 | Metoprolol tartrate | Metoprolol 50mg tablets (A A H Pharmaceuticals Ltd) |
| 34125 | 121711000001108 | Metoprolol tartrate | Metoprolol 100mg tablets (A A H Pharmaceuticals Ltd) |
| 34407 | 126811000001105 | Metoprolol tartrate | Metoprolol 50mg tablets (Teva UK Ltd) |
| 34430 | 499011000001103 | Metoprolol tartrate | Metoprolol 50mg tablets (Actavis UK Ltd) |
| 34509 | 497611000001100 | Metoprolol tartrate | Metoprolol 100mg tablets (Mylan) |
| 34584 | 407511000001102 | Metoprolol tartrate | Metoprolol 50mg tablets (IVAX Pharmaceuticals UK Ltd) |

| | | | |
|---|---|---|---|
| 34854 | 834111000001104 | Metoprolol tartrate | Metoprolol 100mg tablets (Actavis UK Ltd) |
| 34890 | 43455001000027104 | Metoprolol tartrate | Metoprolol 50mg Tablet (Berk Pharmaceuticals Ltd) |
| 34925 | 519211000001104 | Metoprolol tartrate | Metoprolol 50mg tablets (Sandoz Ltd) |
| 40167 | 203711000001103 | Metoprolol tartrate | Metoprolol 100mg tablets (IVAX Pharmaceuticals UK Ltd) |
| 45289 | 243965001000027101 | Metoprolol Tartrate | Metoprolol tartrate Oral solution |
| 46614 | 113511000001109 | Metoprolol tartrate | Lopresor 50mg tablets (Recordati Pharmaceuticals Ltd) |
| 46740 | 756911000001107 | Metoprolol tartrate | Lopresor 100mg tablets (Recordati Pharmaceuticals Ltd) |
| 47536 | 304045001000027103 | Metoprolol Tartrate | Metoprolol tartrate 12.5mg/ 5ml Oral suspension |
| 51447 | 8668411000001106 | Metoprolol tartrate | Metoprolol 12.5mg/ 5ml oral suspension |
| 55979 | 8668811000001108 | Metoprolol tartrate | Metoprolol 25mg/ 5ml oral suspension |
| 57240 | 8635811000001107 | Metoprolol tartrate | Metoprolol 50mg/ 5ml oral suspension (Special Order) |
| 63724 | 21799011000001109 | Metoprolol tartrate | Metoprolol 100mg tablets (Waymade Healthcare Plc) |
| 65227 | 8668211000001107 | Metoprolol tartrate | Metoprolol 12.5mg/ 5ml oral solution |
| 66670 | 610811000001104 | Metoprolol tartrate | Metoprolol 100mg tablets (Alliance Healthcare (Distribution) Ltd) |
| 68881 | 16632411000001109 | Metoprolol tartrate | Metoprolol 12.5mg capsules |
| 70116 | 8668911000001103 | Metoprolol tartrate | Metoprolol 50mg/ 5ml oral solution |
| 71098 | 21286311000001105 | Metoprolol tartrate | Metoprolol 50mg tablets (Accord Healthcare Ltd) |
| 74854 | 8668711000001100 | Metoprolol tartrate | Metoprolol 25mg/ 5ml oral solution |
| 75010 | 16235001000027106 | Metoprolol tartrate | Metoprolol tartrate 50mg Tablet (C P Pharmaceuticals Ltd) |
| 76180 | | Metoprolol tartrate | Metoprolol 5mg/ 5ml oral solution |
| 76531 | | Metoprolol tartrate | Metoprolol 10mg/ 5ml oral suspension |
| 27719 | 160665001000027109 | Metoprolol | Metoros ls 95mg Tablet (Geigy Pharmaceuticals) |
| 29998 | 160625001000027101 | Metoprolol | Metoros 190mg Tablet (Novartis Pharmaceuticals UK Ltd) |
| 77282 | | Metoprolol | Metoprolol 190mg Modified-release tablet |
| 1295 | 318447008 | Labetalol | Labetalol 400mg tablets |
| 1597 | 318445000 | Labetalol | Labetalol 100mg tablets |
| 2775 | 318446004 | Labetalol | Labetalol 200mg tablets |
| 4725 | 318458004 | Labetalol | Labetalol 50mg tablets |
| 8707 | 77311000001105 | Labetalol | Trandate 200mg tablets (RPH Pharmaceuticals AB) |
| 8807 | 674711000001109 | Labetalol | Trandate 400mg tablets (RPH Pharmaceuticals AB) |
| 9016 | 639011000001106 | Labetalol | Trandate 100mg tablets (RPH Pharmaceuticals AB) |
| 9273 | 42611000001104 | Labetalol | Trandate 50mg tablets (RPH Pharmaceuticals AB) |
| 16645 | 82695001000027107 | Labetalol | Labrocol 400mg Tablet (Lagap) |
| 19068 | 7978711000001103 | Labetalol | Labetalol 50mg/ 10ml solution for injection pre-filled syringes |
| 19998 | 4413911000001106 | Labetalol | Trandate 100mg/ 20ml solution for injection ampoules (Focus Pharmaceuticals Ltd) |
| 22793 | 82685001000027108 | Labetalol | Labrocol 200mg Tablet (Lagap) |
| 30770 | 22311000001107 | Labetalol | Labetalol 200mg tablets (A A H Pharmaceuticals Ltd) |
| 34171 | 14865001000027100 | Labetalol | Labetalol 100mg Tablet (C P Pharmaceuticals Ltd) |
| 34177 | 50911000001108 | Labetalol | Labetalol 100mg tablets (A A H Pharmaceuticals Ltd) |
| 34188 | 27395001000027109 | Labetalol | Labetalol 200mg Tablet (Celltech Pharma Europe Ltd) |
| 35778 | 82675001000027103 | Labetalol | Labrocol 100mg Tablet (Lagap) |
| 38370 | 36037611000001104 | Labetalol | Labetalol 100mg/ 20ml solution for injection ampoules |
| 40240 | 246511000001107 | Labetalol | Labetalol 400mg tablets (A A H Pharmaceuticals Ltd) |

| | | | |
|---|---|---|---|
| 41827 | 2211000001109 | Labetalol | Labetalol 100mg tablets (Mylan) |
| 44083 | 265911000001104 | Labetalol | Labetalol 200mg tablets (Actavis UK Ltd) |
| 45250 | 929111000001109 | Labetalol | Labetalol 400mg tablets (Sandoz Ltd) |
| 47673 | 52485001000027108 | Labetalol | Labetalol 400mg Tablet (Approved Prescription Services Ltd) |
| 47674 | 14875001000027107 | Labetalol | Labetalol 200mg Tablet (C P Pharmaceuticals Ltd) |
| 59222 | 21024711000001102 | Labetalol | Labetalol 100mg/ 20ml solution for injection ampoules (RPH Pharmaceuticals AB) |
| 62638 | 281111000001101 | Labetalol | Labetalol 100mg tablets (Actavis UK Ltd) |
| 63736 | 22055311000001102 | Labetalol | Labetalol 100mg tablets (Waymade Healthcare Plc) |
| 72514 | 12643411000001100 | Labetalol | Labetalol 1.5mg/ 5ml oral solution |
| 75179 | 99411000001101 | Labetalol | Labetalol 200mg tablets (Mylan) |
| 77958 | | Labetalol | Labetalol 50mg/ 5ml oral suspension |
| 26922 | 10070311000001104 | Esmolol | Brevibloc Premixed 100mg/ 10ml solution for injection vials (Baxter Healthcare Ltd) |
| 30541 | 71225001000027104 | Esmolol | Esmolol 250mg/ ml concentrate solution for infusion |
| 32135 | 10070411000001106 | Esmolol | Brevibloc Concentrate 2.5g/ 10ml solution for infusion ampoules (Baxter Healthcare Ltd) |
| 39819 | 17033711000001101 | Esmolol | Esmolol 2.5g/ 250ml infusion bags |
| 4265 | 157565001000027107 | Celiprolol | Celectol 200mg Tablet (Pantheon Healthcare Ltd) |
| 7974 | 318622003 | Celiprolol | Celiprolol 400mg tablets |
| 8262 | 318619000 | Celiprolol | Celiprolol 200mg tablets |
| 16776 | 157575001000027100 | Celiprolol | Celectol 400mg Tablet (Pantheon Healthcare Ltd) |
| 35054 | 864811000001100 | Celiprolol | Celectol 200mg tablets (Zentiva) |
| 35940 | 479211000001100 | Celiprolol | Celectol 400mg tablets (Zentiva) |
| 41740 | 749111000001104 | Celiprolol | Celiprolol 200mg tablets (Teva UK Ltd) |
| 42795 | 354811000001105 | Celiprolol | Celiprolol 200mg tablets (Mylan) |
| 56485 | 11028811000001103 | Celiprolol | Celectol 200mg tablets (Waymade Healthcare Plc) |
| 57573 | 5530311000001105 | Celiprolol | Celectol 200mg tablets (Dowelhurst Ltd) |
| 67292 | 14493011000001107 | Celiprolol | Celectol 200mg tablets (Sigma Pharmaceuticals Plc) |
| 74062 | 14493611000001100 | Celiprolol | Celectol 400mg tablets (Sigma Pharmaceuticals Plc) |
| 74623 | 684111000001100 | Celiprolol | Celiprolol 400mg tablets (Mylan) |
| 817 | 318633000 | Carvedilol | Carvedilol 3.125mg tablets |
| 2629 | 318631003 | Carvedilol | Carvedilol 12.5mg tablets |
| 4410 | 318635007 | Carvedilol | Carvedilol 6.25mg tablets |
| 7049 | 318632005 | Carvedilol | Carvedilol 25mg tablets |
| 14117 | 873111000001104 | Carvedilol | Eucardic 3.125mg tablets (Roche Products Ltd) |
| 14146 | 690511000001101 | Carvedilol | Eucardic 6.25mg tablets (Roche Products Ltd) |
| 18414 | 334711000001101 | Carvedilol | Eucardic 12.5mg tablets (Roche Products Ltd) |
| 19202 | 7333911000001104 | Carvedilol | Carvedilol 6.25mg tablets (Teva UK Ltd) |
| 19437 | 709911000001107 | Carvedilol | Eucardic 25mg tablets (Roche Products Ltd) |
| 33374 | 9034211000001104 | Carvedilol | Carvedilol 12.5mg tablets (Genus Pharmaceuticals Ltd) |
| 34501 | 7332611000001100 | Carvedilol | Carvedilol 12.5mg tablets (Actavis UK Ltd) |
| 34740 | 7332411000001103 | Carvedilol | Carvedilol 6.25mg tablets (Actavis UK Ltd) |
| 34741 | 7402011000001106 | Carvedilol | Carvedilol 3.125mg tablets (IVAX Pharmaceuticals UK Ltd) |
| 46935 | 7398911000001101 | Carvedilol | Carvedilol 3.125mg tablets (Actavis UK Ltd) |
| 46936 | 7493711000001102 | Carvedilol | Carvedilol 3.125mg tablets (A A H Pharmaceuticals Ltd) |

| | | | |
|---|---|---|---|
| 47107 | 8356811000001107 | Carvedilol | Carvedilol 5mg/ 5ml oral suspension |
| 49142 | 12422611000001106 | Carvedilol | Carvedilol 3.125mg/ 5ml oral suspension |
| 54106 | 14012311000001106 | Carvedilol | Carvedilol 1.5mg/ 5ml oral suspension |
| 59549 | 8356811000001107 | Carvedilol | Carvedilol 5mg/ 5ml oral suspension |
| 61663 | 7333711000001101 | Carvedilol | Carvedilol 3.125mg tablets (Teva UK Ltd) |
| 63422 | 21820811000001101 | Carvedilol | Carvedilol 12.5mg tablets (Waymade Healthcare Plc) |
| 67661 | 15072711000001108 | Carvedilol | Carvedilol 6.25mg tablets (Sigma Pharmaceuticals Plc) |
| 72507 | 7334311000001103 | Carvedilol | Carvedilol 25mg tablets (Teva UK Ltd) |
| 73451 | 11401811000001105 | Carvedilol | Carvedilol 6.25mg tablets (Almus Pharmaceuticals Ltd) |
| 74619 | 7334111000001100 | Carvedilol | Carvedilol 12.5mg tablets (Teva UK Ltd) |
| 77648 | | Carvedilol | Carvedilol 12.5mg tablets (Almus Pharmaceuticals Ltd) |
| 472 | 318590006 | Bisoprolol fumarate | Bisoprolol 5mg tablets |
| 594 | 318605000 | Bisoprolol fumarate | Bisoprolol 2.5mg tablets |
| 599 | 318604001 | Bisoprolol fumarate | Bisoprolol 1.25mg tablets |
| 822 | 243085001000027105 | Bisoprolol Fumarate | Bisoprolol 1.5mg/ 5ml oral suspension |
| 1290 | 318591005 | Bisoprolol fumarate | Bisoprolol 10mg tablets |
| 3588 | 349611000001108 | Bisoprolol fumarate | Monocor 5mg tablets (Wyeth Pharmaceuticals) |
| 4771 | 483911000001101 | Bisoprolol fumarate | Emcor LS 5mg tablets (Merck Serono Ltd) |
| 5713 | 318607008 | Bisoprolol fumarate | Bisoprolol 7.5mg tablets |
| 5968 | 329611000001106 | Bisoprolol fumarate | Monocor 10mg tablets (Wyeth Pharmaceuticals) |
| 7091 | 318606004 | Bisoprolol fumarate | Bisoprolol 3.75mg tablets |
| 7553 | 8304811000001108 | Bisoprolol fumarate | Bisoprolol 5mg/ 5ml oral suspension |
| 10892 | 106611000001104 | Bisoprolol fumarate | Emcor 10mg tablets (Merck Serono Ltd) |
| 14030 | 628711000001102 | Bisoprolol fumarate | Cardicor 2.5mg tablets (Merck Serono Ltd) |
| 14058 | 712511000001106 | Bisoprolol fumarate | Cardicor 1.25mg tablets (Merck Serono Ltd) |
| 17615 | 19711000001106 | Bisoprolol fumarate | Cardicor 5mg tablets (Merck Serono Ltd) |
| 18185 | 404511000001105 | Bisoprolol fumarate | Cardicor 7.5mg tablets (Merck Serono Ltd) |
| 19178 | 78411000001104 | Bisoprolol fumarate | Bisoprolol 10mg tablets (Ranbaxy (UK) Ltd) |
| 19200 | 433811000001104 | Bisoprolol fumarate | Bisoprolol 5mg tablets (IVAX Pharmaceuticals UK Ltd) |
| 19853 | 823111000001103 | Bisoprolol fumarate | Cardicor 3.75mg tablets (Merck Serono Ltd) |
| 19858 | 890111000001100 | Bisoprolol fumarate | Cardicor 10mg tablets (Merck Serono Ltd) |
| 21905 | 405411000001107 | Bisoprolol fumarate | Bipranix 10mg tablets (Ashbourne Pharmaceuticals Ltd) |
| 21966 | 501211000001103 | Bisoprolol fumarate | Bipranix 5mg tablets (Ashbourne Pharmaceuticals Ltd) |
| 24083 | 853911000001104 | Bisoprolol fumarate | Bisoprolol 5mg tablets (Teva UK Ltd) |
| 32114 | 573811000001107 | Bisoprolol fumarate | Bisoprolol 5mg tablets (Mylan) |
| 32552 | 10241511000001108 | Bisoprolol fumarate | Congescor 2.5mg tablets (Tillomed Laboratories Ltd) |
| 32630 | 9739311000001105 | Bisoprolol fumarate | Vivacor 10mg tablets (Lexon (UK) Ltd) |
| 33839 | 13211000001101 | Bisoprolol fumarate | Bisoprolol 10mg tablets (Actavis UK Ltd) |
| 33909 | 10241311000001102 | Bisoprolol fumarate | Congescor 1.25mg tablets (Tillomed Laboratories Ltd) |
| 34821 | 502311000001102 | Bisoprolol fumarate | Bisoprolol 10mg tablets (Mylan) |
| 34963 | 706111000001101 | Bisoprolol fumarate | Bisoprolol 5mg tablets (Actavis UK Ltd) |
| 37118 | 10743711000001105 | Bisoprolol fumarate | Bisoprolol 2.5mg tablets (A A H Pharmaceuticals Ltd) |
| 37837 | 187815001000027109 | Bisoprolol fumarate | Bisoprolol 2.5mg Tablet (Teva UK Ltd) |

| | | | |
|---|---|---|---|
| 38991 | 13919811000001103 | Bisoprolol fumarate | Bisoprolol 7.5mg tablets (A A H Pharmaceuticals Ltd) |
| 39646 | 256485001000027100 | Bisoprolol Fumarate | Bisoprolol 0.625mg/ 5ml oral solution |
| 39846 | 9105811000001100 | Bisoprolol fumarate | Vivacor 5mg tablets (Lexon (UK) Ltd) |
| 41591 | 888911000001101 | Bisoprolol fumarate | Bisoprolol 10mg tablets (Teva UK Ltd) |
| 43251 | 13436011000001108 | Bisoprolol fumarate | Bisoprolol 1.25mg tablets (Mylan) |
| 43564 | 112235001000027100 | Bisoprolol fumarate | Bisoprolol 5mg Tablet (PLIVA Pharma Ltd) |
| 44000 | 12287611000001100 | Bisoprolol fumarate | Bisoprolol 2.5mg/ 5ml oral suspension |
| 47041 | 13436211000001103 | Bisoprolol fumarate | Bisoprolol 2.5mg tablets (Mylan) |
| 50224 | 20475811000001108 | Bisoprolol fumarate | Congescor 2.5mg tablets (Teva UK Ltd) |
| 50300 | 20475611000001109 | Bisoprolol fumarate | Congescor 1.25mg tablets (Teva UK Ltd) |
| 50403 | 187775001000027103 | Bisoprolol fumarate | Bisoprolol 1.25mg Tablet (Teva UK Ltd) |
| 50514 | 17012411000001106 | Bisoprolol fumarate | Bisoprolol 2.5mg tablets (Chanelle Medical UK Ltd) |
| 51528 | 20358011000001102 | Bisoprolol fumarate | Bisoprolol 1.25mg tablets (Actavis UK Ltd) |
| 52548 | 18506211000001103 | Bisoprolol fumarate | Bisoprolol 1.25mg tablets (Almus Pharmaceuticals Ltd) |
| 52611 | 12287311000001105 | Bisoprolol fumarate | Bisoprolol 10mg/ 5ml oral solution |
| 52635 | 557311000001108 | Bisoprolol fumarate | Bisoprolol 5mg tablets (Alliance Healthcare (Distribution) Ltd) |
| 52686 | 12287511000001104 | Bisoprolol fumarate | Bisoprolol 2.5mg/ 5ml oral solution |
| 53334 | 632011000001104 | Bisoprolol fumarate | Bisoprolol 10mg tablets (A A H Pharmaceuticals Ltd) |
| 53664 | 20283611000001102 | Bisoprolol fumarate | Bisoprolol 2.5mg tablets (Sandoz Ltd) |
| 53885 | 11252511000001108 | Bisoprolol fumarate | Bisoprolol 1.25mg tablets (A A H Pharmaceuticals Ltd) |
| 53916 | 18506411000001104 | Bisoprolol fumarate | Bisoprolol 2.5mg tablets (Almus Pharmaceuticals Ltd) |
| 54479 | 10740611000001105 | Bisoprolol fumarate | Bisoprolol 1.25mg tablets (Alliance Healthcare (Distribution) Ltd) |
| 55298 | 15058011000001107 | Bisoprolol fumarate | Bisoprolol 10mg tablets (Sigma Pharmaceuticals Plc) |
| 55791 | 20357211000001108 | Bisoprolol fumarate | Bisoprolol 3.75mg tablets (Actavis UK Ltd) |
| 55929 | 21281511000001108 | Bisoprolol fumarate | Bisoprolol 5mg tablets (Accord Healthcare Ltd) |
| 56240 | 20283811000001103 | Bisoprolol fumarate | Bisoprolol 3.75mg tablets (Sandoz Ltd) |
| 56459 | 21281311000001102 | Bisoprolol fumarate | Bisoprolol 2.5mg tablets (Accord Healthcare Ltd) |
| 56486 | 5582411000001107 | Bisoprolol fumarate | Monocor 10mg tablets (Dowelhurst Ltd) |
| 56768 | 18553111000001106 | Bisoprolol fumarate | Bisoprolol 2.5mg tablets (Niche Generics Ltd) |
| 57023 | 18506411000001104 | Bisoprolol fumarate | Bisoprolol 2.5mg tablets (Almus Pharmaceuticals Ltd) |
| 57176 | 21281711000001103 | Bisoprolol fumarate | Bisoprolol 10mg tablets (Accord Healthcare Ltd) |
| 57578 | 17619411000001108 | Bisoprolol fumarate | Cardicor 2.5mg tablets (Necessity Supplies Ltd) |
| 57626 | 8305311000001100 | Bisoprolol fumarate | Bisoprolol 1.25mg/ 5ml oral solution |
| 57934 | 7376711000001106 | Bisoprolol fumarate | Bisoprolol 5mg tablets (Sandoz Ltd) |
| 58109 | 8305211000001108 | Bisoprolol fumarate | Bisoprolol 1.25mg/ 5ml oral suspension |
| 58455 | 20284011000001106 | Bisoprolol fumarate | Bisoprolol 7.5mg tablets (Sandoz Ltd) |
| 58498 | 22496711000001105 | Bisoprolol fumarate | Bisoprolol 2.5mg tablets (Medreich Plc) |
| 58511 | 20283411000001100 | Bisoprolol fumarate | Bisoprolol 1.25mg tablets (Sandoz Ltd) |
| 58763 | 21802011000001102 | Bisoprolol fumarate | Bisoprolol 2.5mg tablets (Waymade Healthcare Plc) |
| 58973 | 10437911000001106 | Bisoprolol fumarate | Bisoprolol 10mg tablets (Niche Generics Ltd) |
| 58974 | 10740811000001109 | Bisoprolol fumarate | Bisoprolol 2.5mg tablets (Alliance Healthcare (Distribution) Ltd) |
| 58982 | 19734511000001102 | Bisoprolol fumarate | Bisoprolol 10mg tablets (Medreich Plc) |
| 59037 | 249811000001104 | Bisoprolol fumarate | Bisoprolol 5mg tablets (A A H Pharmaceuticals Ltd) |

| | | | |
|---|---|---|---|
| 59148 | 20577811000001109 | Bisoprolol fumarate | Bisoprolol 2.5mg tablets (Zentiva) |
| 59495 | 21105111000001104 | Bisoprolol fumarate | Bisoprolol 1.25mg tablets (Teva UK Ltd) |
| 59969 | 18497711000001100 | Bisoprolol fumarate | Bisoprolol 5mg tablets (Almus Pharmaceuticals Ltd) |
| 60502 | 19708211000001104 | Bisoprolol fumarate | Bisoprolol 3.75mg tablets (DE Pharmaceuticals) |
| 60761 | 22495911000001100 | Bisoprolol fumarate | Bisoprolol 1.25mg tablets (Medreich Plc) |
| 60896 | 19734211000001100 | Bisoprolol fumarate | Bisoprolol 5mg tablets (Medreich Plc) |
| 61115 | 8304911000001103 | Bisoprolol fumarate | Bisoprolol 5mg/ 5ml oral solution |
| 61340 | 19708411000001100 | Bisoprolol fumarate | Bisoprolol 5mg tablets (DE Pharmaceuticals) |
| 61564 | 21802311000001104 | Bisoprolol fumarate | Bisoprolol 3.75mg tablets (Waymade Healthcare Plc) |
| 61651 | 18506811000001102 | Bisoprolol fumarate | Bisoprolol 7.5mg tablets (Almus Pharmaceuticals Ltd) |
| 62361 | 17012211000001107 | Bisoprolol fumarate | Bisoprolol 1.25mg tablets (Chanelle Medical UK Ltd) |
| 62407 | 250065001000027102 | Bisoprolol Fumarate | Bisoprolol oral solution |
| 63493 | 17328511000001103 | Bisoprolol fumarate | Bisoprolol 2.5mg tablets (Actavis UK Ltd) |
| 63535 | 10285811000001100 | Bisoprolol fumarate | Bisoprolol 5mg tablets (Relonchem Ltd) |
| 63850 | 21108211000001103 | Bisoprolol fumarate | Bisoprolol 2.5mg tablets (Teva UK Ltd) |
| 64538 | 18743711000001108 | Bisoprolol fumarate | Bisoprolol 3.75mg tablets (Teva UK Ltd) |
| 64784 | 10438411000001104 | Bisoprolol fumarate | Bisoprolol 5mg tablets (Niche Generics Ltd) |
| 64850 | 19708011000001109 | Bisoprolol fumarate | Bisoprolol 2.5mg tablets (DE Pharmaceuticals) |
| 65805 | 21800411000001105 | Bisoprolol fumarate | Bisoprolol 1.25mg tablets (Waymade Healthcare Plc) |
| 65821 | 14159111000001101 | Bisoprolol fumarate | Bisoprolol 7.5mg/ 5ml oral suspension |
| 69156 | 22495711000001102 | Bisoprolol fumarate | Bisoprolol 3.75mg tablets (Medreich Plc) |
| 71472 | 21801411000001101 | Bisoprolol fumarate | Bisoprolol 5mg tablets (Waymade Healthcare Plc) |
| 72285 | 30001311000001100 | Bisoprolol fumarate | Bisoprolol 1.25mg tablets (Mawdsley-Brooks & Company Ltd) |
| 72540 | 12287411000001103 | Bisoprolol fumarate | Bisoprolol 10mg/ 5ml oral suspension |
| 73638 | 18497711000001100 | Bisoprolol fumarate | Bisoprolol 5mg tablets (Almus Pharmaceuticals Ltd) |
| 73641 | 18497911000001103 | Bisoprolol fumarate | Bisoprolol 10mg tablets (Almus Pharmaceuticals Ltd) |
| 73809 | 18506211000001103 | Bisoprolol fumarate | Bisoprolol 1.25mg tablets (Almus Pharmaceuticals Ltd) |
| 74742 | 15160311000001108 | Bisoprolol fumarate | Bisoprolol 625micrograms/ 5ml oral suspension |
| 75463 | | Bisoprolol fumarate | Bisoprolol 625micrograms/ 5ml oral solution |
| 76299 | | Bisoprolol fumarate | Bisoprolol 10mg tablets (Kent Pharmaceuticals Ltd) |
| 76322 | | Bisoprolol fumarate | Bisoprolol 10mg tablets (Almus Pharmaceuticals Ltd) |
| 76483 | | Bisoprolol fumarate | Bisoprolol 1.25mg tablets (Sigma Pharmaceuticals Plc) |
| 77219 | | Bisoprolol fumarate | Bisoprolol 7.5mg tablets (Waymade Healthcare Plc) |
| 5 | 318420003 | Atenolol | Atenolol 50mg tablets |
| 24 | 318421004 | Atenolol | Atenolol 100mg tablets |
| 26 | 318434003 | Atenolol | Atenolol 25mg tablets |
| 197 | 35903411000001106 | Atenolol | Atenolol 5mg/ 10ml solution for injection ampoules |
| 2432 | 423911000001107 | Atenolol | Tenormin LS 50mg tablets (AstraZeneca UK Ltd) |
| 2587 | 162411000001102 | Atenolol | Tenormin 100mg tablets (AstraZeneca UK Ltd) |
| 2590 | 317111000001101 | Atenolol | Tenormin 25mg tablets (AstraZeneca UK Ltd) |
| 6066 | 35903211000001107 | Atenolol | Atenolol 25mg/ 5ml oral solution sugar free |
| 7429 | 9111000001107 | Atenolol | Tenormin 5mg/ 10ml solution for injection ampoules (AstraZeneca UK Ltd) |
| 10191 | 271911000001106 | Atenolol | Atenix 50 tablets (Ashbourne Pharmaceuticals Ltd) |

| | | | |
|---|---|---|---|
| 13394 | 373311000001100 | Atenolol | Tenormin 25mg/ 5ml syrup (AstraZeneca UK Ltd) |
| 15176 | 18217500100002710 8 | Atenolol | Totamol 50mg Tablet (C P Pharmaceuticals Ltd) |
| 15730 | 18218500100002710 3 | Atenolol | Totamol 100mg Tablet (C P Pharmaceuticals Ltd) |
| 17322 | 482511000001101 | Atenolol | Atenix 25 tablets (Ashbourne Pharmaceuticals Ltd) |
| 18950 | 18219500100002710 4 | Atenolol | Totamol 25mg Tablet (C P Pharmaceuticals Ltd) |
| 19172 | 600811000001101 | Atenolol | Atenolol 25mg tablets (IVAX Pharmaceuticals UK Ltd) |
| 19182 | 852111000001108 | Atenolol | Atenolol 50mg tablets (IVAX Pharmaceuticals UK Ltd) |
| 19191 | 857511000001101 | Atenolol | Atenolol 100mg tablets (Teva UK Ltd) |
| 20502 | 181611000001107 | Atenolol | Atenix 100 tablets (Ashbourne Pharmaceuticals Ltd) |
| 20728 | 23489500100002710 6 | Atenolol | Atenamin 25mg Tablet (OPD Pharm) |
| 21133 | 23490500100002710 3 | Atenolol | Atenamin 50mg Tablet (OPD Pharm) |
| 24191 | 874711000001101 | Atenolol | Antipressan 50mg tablets (Teva UK Ltd) |
| 24195 | 734011000001102 | Atenolol | Antipressan 100mg tablets (Teva UK Ltd) |
| 26211 | 877011000001106 | Atenolol | Antipressan 25mg tablets (Teva UK Ltd) |
| 29368 | 623811000001100 | Atenolol | Atenolol 25mg tablets (Teva UK Ltd) |
| 29398 | 23491500100002710 1 | Atenolol | Atenamin 100mg Tablet (OPD Pharm) |
| 30636 | 18214500100002710 7 | Atenolol | Vasaten 50mg Tablet (Shire Pharmaceuticals Ltd) |
| 31536 | 907211000001102 | Atenolol | Atenolol 25mg tablets (Kent Pharmaceuticals Ltd) |
| 31934 | 646611000001108 | Atenolol | Atenolol 100mg tablets (IVAX Pharmaceuticals UK Ltd) |
| 33079 | 35311000001104 | Atenolol | Atenolol 100mg tablets (Mylan) |
| 33085 | 422011000001104 | Atenolol | Atenolol 100mg tablets (A A H Pharmaceuticals Ltd) |
| 33092 | 58311000001109 | Atenolol | Atenolol 50mg tablets (A A H Pharmaceuticals Ltd) |
| 33184 | 417811000001102 | Atenolol | Atenolol 100mg tablets (Wockhardt UK Ltd) |
| 33650 | 225111000001106 | Atenolol | Atenolol 50mg tablets (Mylan) |
| 33657 | 10811000001107 | Atenolol | Atenolol 25mg tablets (A A H Pharmaceuticals Ltd) |
| 33850 | 401411000001109 | Atenolol | Atenolol 50mg tablets (Actavis UK Ltd) |
| 34265 | 884011000001109 | Atenolol | Atenolol 50mg tablets (Sandoz Ltd) |
| 34365 | 674511000001104 | Atenolol | Atenolol 50mg tablets (Teva UK Ltd) |
| 34443 | 335611000001106 | Atenolol | Atenolol 50mg tablets (Wockhardt UK Ltd) |
| 34492 | 774211000001105 | Atenolol | Atenolol 25mg tablets (Mylan) |
| 34575 | 375411000001106 | Atenolol | Atenolol 25mg tablets (Wockhardt UK Ltd) |
| 34585 | 393011000001109 | Atenolol | Atenolol 25mg tablets (Sandoz Ltd) |
| 34695 | 790911000001104 | Atenolol | Atenolol 50mg tablets (Kent Pharmaceuticals Ltd) |
| 34754 | 282111000001106 | Atenolol | Atenolol 100mg tablets (Sandoz Ltd) |
| 34882 | 3898500100002710 9 | Atenolol | Atenolol 50mg Tablet (Berk Pharmaceuticals Ltd) |
| 34976 | 13740111000001101 | Atenolol | Atenolol 25mg tablets (Tillomed Laboratories Ltd) |
| 36261 | 13740311000001104 | Atenolol | Atenolol 50mg tablets (Tillomed Laboratories Ltd) |
| 44858 | 244111000001105 | Atenolol | Atenolol 25mg tablets (Actavis UK Ltd) |
| 46908 | 721711000001100 | Atenolol | Atenolol 100mg tablets (Kent Pharmaceuticals Ltd) |
| 46931 | 275811000001103 | Atenolol | Atenolol 100mg tablets (Actavis UK Ltd) |
| 47870 | 9791011000001101 | Atenolol | Atenolol 25mg tablets (Almus Pharmaceuticals Ltd) |
| 49953 | 15986211000001103 | Atenolol | Atenolol 25mg tablets (Bristol Laboratories Ltd) |
| 50702 | 47711000001105 | Atenolol | Atenolol 25mg tablets (Alliance Healthcare (Distribution) Ltd) |

| | | | |
|---|---|---|---|
| 51643 | 11560811000001105 | Atenolol | Atenolol 25mg/ 5ml oral solution sugar free (Alliance Healthcare (Distribution) Ltd) |
| 51998 | 18280111000001102 | Atenolol | Atenolol 25mg tablets (Strides Shasun (UK) Ltd) |
| 52310 | 20137611000001100 | Atenolol | Atenolol 25mg tablets (Crescent Pharma Ltd) |
| 52500 | 9791711000001104 | Atenolol | Atenolol 50mg tablets (Almus Pharmaceuticals Ltd) |
| 53204 | 263511000001107 | Atenolol | Atenolol 50mg tablets (Alliance Healthcare (Distribution) Ltd) |
| 53215 | 15986411000001104 | Atenolol | Atenolol 50mg tablets (Bristol Laboratories Ltd) |
| 53414 | 18458611000001100 | Atenolol | Atenolol 50mg tablets (Accord Healthcare Ltd) |
| 53802 | 15070211000001102 | Atenolol | Atenolol 25mg tablets (Sigma Pharmaceuticals Plc) |
| 53826 | 14801811000001107 | Atenolol | Atenolol 25mg tablets (Boston Healthcare Ltd) |
| 54542 | 15968011000001105 | Atenolol | Atenolol 25mg tablets (Zanza Laboratories Ltd) |
| 54752 | 18279911000001104 | Atenolol | Atenolol 50mg tablets (Strides Shasun (UK) Ltd) |
| 55778 | 17788311000001101 | Atenolol | Atenolol 50mg tablets (Phoenix Healthcare Distribution Ltd) |
| 56445 | 16183211000001105 | Atenolol | Atenolol 25mg/ 5ml oral solution sugar free (A A H Pharmaceuticals Ltd) |
| 57817 | 11177311000001101 | Atenolol | Atenolol 50mg tablets (Zentiva) |
| 59695 | 14802011000001109 | Atenolol | Atenolol 50mg tablets (Boston Healthcare Ltd) |
| 59982 | 18458411000001103 | Atenolol | Atenolol 25mg tablets (Accord Healthcare Ltd) |
| 61573 | 12015011000001105 | Atenolol | Atenolol 25mg/ 5ml oral solution |
| 62325 | 21778611000001100 | Atenolol | Atenolol 25mg tablets (Waymade Healthcare Plc) |
| 64973 | 29771711000001105 | Atenolol | Atenolol 50mg tablets (Sigma Pharmaceuticals Plc) |
| 66548 | 22612611000001101 | Atenolol | Atenolol 50mg tablets (DE Pharmaceuticals) |
| 69526 | 12079211000001106 | Atenolol | Atenolol 50mg/ 5ml oral solution |
| 70135 | 22612411000001104 | Atenolol | Atenolol 25mg tablets (DE Pharmaceuticals) |
| 71026 | 11399511000001103 | Atenolol | Atenolol 100mg tablets (Almus Pharmaceuticals Ltd) |
| 72043 | 21778811000001101 | Atenolol | Atenolol 50mg tablets (Waymade Healthcare Plc) |
| 72810 | 12014311000001109 | Atenolol | Atenolol 10mg/ 5ml oral solution |
| 73151 | 28945211000001109 | Atenolol | Atenolol 50mg tablets (Crescent Pharma Ltd) |
| 76593 | | Atenolol | Atenolol 25mg/ 5ml oral solution sugar free (DE Pharmaceuticals) |
| 77198 | | Atenolol | Atenolol 100mg tablets (Phoenix Healthcare Distribution Ltd) |
| 77613 | | Atenolol | Atenolol 25mg/ 5ml oral suspension |
| 7620 | 318414004 | Acebutolol | Acebutolol 400mg tablets |
| 8023 | 298111000001105 | Acebutolol | Sectral 400mg tablets (Sanofi) |
| 8113 | 318413005 | Acebutolol | Acebutolol 200mg capsules |
| 8172 | 318412000 | Acebutolol | Acebutolol 100mg capsules |
| 8555 | 925711000001108 | Acebutolol | Sectral 200mg capsules (Sanofi) |
| 12296 | 632811000001105 | Acebutolol | Sectral 100mg capsules (Sanofi) |
| 45309 | 9703611000001105 | Acebutolol | Acebutolol 400mg tablets (A A H Pharmaceuticals Ltd) |
| 65438 | 9703211000001108 | Acebutolol | Acebutolol 100mg capsules (A A H Pharmaceuticals Ltd) |
| 72545 | 9703411000001107 | Acebutolol | Acebutolol 200mg capsules (A A H Pharmaceuticals Ltd) |
| 77435 | | Acebutolol | Sectral 400mg tablets (Mawdsley-Brooks & Company Ltd) |
| 3041 | 108205001000027101 |  | SOTALOL 40 MG INJ |
| 4021 | 177775001000027104 | | PROPRANOLOL 20 MG TAB |
| 7491 | 177255001000027105 | | LABETALOL TAB |
| 12119 | 132615001000027106 |  | SOTALOL S/ R 80 MG TAB |

| | | | |
|---|---|---|---|
| 12497 | 166645001000027109 | | OXPRENOLOL 10 MG TAB |
| 16669 | 72615001000027105 | | LOPRESOR SR 200 MG TAB |
| 17876 | 112415001000027107 | | METOPROLOL FUMARATE 190 MG TAB |
| 18722 | | | CARVEDILOL |
| 19810 | 181045001000027103 | | BEDRANOL SR 80 MG CAP |
| 20813 | | | BETALOC S.A. |
| 22634 | 135155001000027102 | | PROPRANOLOL 10 MG SUS |
| 22796 | | | CARVEDILOL 3.125 MG |
| 23598 | | | SOTALOL S/ R |
| 23604 | | | PROPRANOLOL S/ R |
| 24378 | | | BETALOC S.A. (CALENDAR PACK) |
| 24677 | | | ATENOLOL |
| 25037 | | | ATENOLOL |
| 25052 | | | HALF-INDERAL LA |
| 25818 | 164975001000027100 | | PROPRANOLOL 30 MG SUS |
| 26105 | 165015001000027106 | | PROPRANOLOL paed 4 MG TAB |
| 26290 | 99325001000027104 | | SOTACOR 40 MG INJ |
| 26788 | 135175001000027104 | | PROPRANOLOL 2.5 MG ELI |
| 27036 | 164995001000027109 | | PROPRANOLOL POWDERS 5 MG POW |
| 28493 | 100955001000027108 | | METOPROLOL FUMARATE 95 MG TAB |
| 29803 | 135135001000027101 | | PROPRANOLOL 3 MG ELI |
| 32470 | 135035001000027107 | | PROPRANOLOL 1 MG LIQ |
| 44310 | | | INDERAL |
| 46493 | 135125001000027103 | | PROPRANOLOL 15 MG SYR |
| **Betablockers in combination with calcium channel blockers** | | | |
| 15117 | 156825001000027102 | Nifedipine/ Atenolol | Nifedipine with atenolol 20mg + 50mg Capsule |
| 1684 | 3142711000001107 | Atenolol/ Nifedipine | Beta-Adalat modified-release capsules (Bayer Plc) |
| 4542 | 35903311000001104 | Atenolol/ Nifedipine | Atenolol 50mg / Nifedipine 20mg modified-release capsules |
| 8642 | 3142511000001102 | Atenolol/ Nifedipine | Tenif 50mg/ 20mg modified-release capsules (AstraZeneca UK Ltd) |
| 52728 | 16141111000001106 | Atenolol/ Nifedipine | Beta-Adalat modified-release capsules (Lexon (UK) Ltd) |
| 61719 | 10490311000001102 | Atenolol/ Nifedipine | Beta-Adalat modified-release capsules (Waymade Healthcare Plc) |
| 68020 | 14208611000001108 | Atenolol/ Nifedipine | Beta-Adalat modified-release capsules (Sigma Pharmaceuticals Plc) |
| 74039 | 14625611000001108 | Atenolol/ Nifedipine | Tenif 50mg/ 20mg modified-release capsules (Sigma Pharmaceuticals Plc) |
| 76440 | | Atenolol/ Nifedipine | Beta-Adalat modified-release capsules (Dowelhurst Ltd) |
| **Betablockers and Diuretics** | | | |
| 8623 | 38405001000027107 | Timolol maleate/ Bendroflumethiazide | Prestim Tablet (ICN Pharmaceuticals France S.A.) |
| 12517 | 144255001000027109 | Timolol Maleate/ Bendroflumethiazide | Timolol maleate with bendroflumethiazide 20mg + 5mg Tablet |
| 12651 | 318556002 | Timolol maleate/ Bendroflumethiazide | Timolol 10mg / Bendroflumethiazide 2.5mg tablets |

| | | | |
|---|---|---|---|
| 19142 | 14885501000027103 | Timolol Maleate/ Bendroflumethiazide | Bendroflumethiazide 2.5mg with Timolol maleate 10mg tablets |
| 21025 | 14432501000027103 | Timolol Maleate/ Bendroflumethiazide | Prestim forte Tablet (LEO Pharma) |
| 25363 | 98411000001109 | Timolol maleate/ Bendroflumethiazide | Prestim tablets (Meda Pharmaceuticals Ltd) |
| 3691 | 14129501000027106 | Sotalol / Hydrochlorothiazide | Sotalol 160mg with hydrochlorothiazide 25mg tablet |
| 8061 | 14128501000027107 | Sotalol / Hydrochlorothiazide | Sotalol 80mg with hydrochlorothiazide 12.5mg tablet |
| 12456 | 29205001000027105 | Sotalol / Hydrochlorothiazide | Sotazide Tablet (Bristol-Myers Squibb Pharmaceuticals Ltd) |
| 15042 | 14140501000027109 | Sotalol / Hydrochlorothiazide | Tolerzide Tablet (Bristol-Myers Squibb Pharmaceuticals Ltd) |
| 17783 | 75195001000027107 | Propranolol / Spironolactone | Spiroprop Tablet (Pharmacia Ltd) |
| 4796 | 333111000001104 | Propranolol / Bendroflumethiazide | Inderetic 80mg/ 2.5mg capsules (AstraZeneca UK Ltd) |
| 8369 | 350811000001100 | Propranolol / Bendroflumethiazide | Inderex 160mg/ 5mg modified-release capsules (AstraZeneca UK Ltd) |
| 8987 | 35932411000001108 | Propranolol / Bendroflumethiazide | Propranolol 160mg modified-release / Bendroflumethiazide 5mg capsules |
| 12054 | 318584007 | Propranolol / Bendroflumethiazide | Propranolol 80mg / Bendroflumethiazide 2.5mg capsules |
| 22912 | 14873501000027104 | Propranolol / Bendroflumethiazide | Bendroflumethiazide 2.5mg with Propanolol 80mg capsules |
| 23131 | 14874501000027103 | Propranolol / Bendroflumethiazide | Bendroflumethiazide 5mg with Propanolol 160mg modified-release capsules |
| 25462 | 10333501000027100 | Pindolol/ Clopamide | Clopamide 5mg with Pindolol 10mg tablets |
| 77387 | | Pindolol/ Clopamide | Viskaldix tablets (Waymade Healthcare Plc) |
| 24832 | 39675001000027101 | Penbutolol/ Furosemide | Lasipressin Tablet (Hoechst UK Ltd) |
| 26529 | 10801501000027108 | Penbutolol/ Furosemide | Furosemide with penbutolol Tablet |
| 4429 | 3444411000001107 | Oxprenolol / Cyclopenthiazide | Trasidrex modified-release tablets (Mercury Pharma Group Ltd) |
| 8673 | 18958501000027103 | Oxprenolol / Cyclopenthiazide | Oxprenolol with cyclopenthiazide 160mg+0.25mg Modified-release tablet |
| 13871 | 36091611000001101 | Oxprenolol / Cyclopenthiazide | Co-prenozide 160mg/ 0.25mg modified-release tablets |
| 52145 | 10460501000027104 | Oxprenolol / Cyclopenthiazide | Cyclopenthiazide 0.25mg with oxprenolol 160mg modified-release tablets |
| 5330 | 3886211000001102 | Nadolol/ Bendroflumethiazide | Corgaretic 40mg tablets (Sanofi-Synthelabo Ltd) |
| 11338 | 14880501000027106 | Nadolol/ Bendroflumethiazide | Bendroflumethiazide 5mg with Nadolol 40mg tablets |
| 14438 | 4057911000001102 | Nadolol/ Bendroflumethiazide | Corgaretic 80mg tablets (Sanofi-Synthelabo Ltd) |
| 23134 | 318549008 | Nadolol/ Bendroflumethiazide | Nadolol 40mg / Bendroflumethiazide 5mg tablets |
| 27946 | 318550008 | Nadolol/ Bendroflumethiazide | Nadolol 80mg / Bendroflumethiazide 5mg tablets |
| 69334 | 14881501000027108 | Nadolol/ Bendroflumethiazide | Bendroflumethiazide 5mg with Nadolol 80mg tablets |
| 7066 | 318546001 | Metoprolol tartrate/ Hydrochlorothiazide | Metoprolol 100mg / Hydrochlorothiazide 12.5mg tablets |
| 10627 | 2977611000001106 | Metoprolol tartrate/ Hydrochlorothiazide | Co-Betaloc tablets (Pfizer Ltd) |
| 18287 | 3853411000001104 | Metoprolol tartrate/ Hydrochlorothiazide | Co-Betaloc SA tablets (Pfizer Ltd) |
| 20093 | 36035311000001101 | Metoprolol tartrate/ Hydrochlorothiazide | Metoprolol 200mg modified-release / Hydrochlorothiazide 25mg tablets |
| 29427 | 15764501000027106 | Metoprolol Tartrate/ Hydrochlorothiazide | Hydrochlorothiazide with metoprolol tartrate 12.5mg with 100mg tablet |
| 33659 | 15763501000027105 | Metoprolol Tartrate/ Hydrochlorothiazide | Hydrochlorothiazide with metoprolol tartrate 25mg with 200mg Modified-release tablet |

| | | | |
|---|---|---|---|
| 8147 | 17805001000027109 | Metoprolol Tartrate/ Chlortalidone | Lopresoretic Tablet (Novartis Pharmaceuticals UK Ltd) |
| 15488 | 157605001000027101 | Metoprolol Tartrate/ Chlortalidone | Metoprolol tartrate with chlortalidone Tablet |
| 9143 | 3638411000001101 | Clopamide/ Pindolol | Viskaldix tablets (AMCo) |
| 14057 | 318552000 | Clopamide/ Pindolol | Pindolol 10mg / Clopamide 5mg tablets |
| 581 | 148505001000027107 | Chlortalidone/ Atenolol | Atenolol 50mg with Chlortalidone 12.5mg tablets |
| 1788 | 148515001000027109 | Chlortalidone/ Atenolol | Atenolol 100mg with Chlortalidone 25mg tablets |
| 16786 | 101705001000027101 | Chlortalidone/ Atenolol | Chlortalidone 25mg with Atenolol 100mg tablets |
| 19055 | 101695001000027105 | Chlortalidone/ Atenolol | Chlortalidone 12.5mg with Atenolol 50mg tablets |
| 17149 | 4542911000001104 | Bisoprolol fumarate/ Hydrochlorothiazide | Monozide 10 tablets (Wyeth Pharmaceuticals) |
| 17462 | 318597009 | Bisoprolol fumarate/ Hydrochlorothiazide | Bisoprolol 10mg / Hydrochlorothiazide 6.25mg tablets |
| 1124 | 288711000001105 | Atenolol/ Chlortalidone | Tenoretic 100mg/ 25mg tablets (AstraZeneca UK Ltd) |
| 1288 | 825011000001103 | Atenolol/ Chlortalidone | Tenoret 50mg/ 12.5mg tablets (AstraZeneca UK Ltd) |
| 5721 | 377211005 | Atenolol/ Chlortalidone | Co-tenidone 100mg/ 25mg tablets |
| 9783 | 318575007 | Atenolol/ Chlortalidone | Co-tenidone 50mg/ 12.5mg tablets |
| 13526 | 101411000001108 | Atenolol/ Chlortalidone | Atenix Co 100 tablets (Ashbourne Pharmaceuticals Ltd) |
| 21873 | 629011000001109 | Atenolol/ Chlortalidone | Atenix Co 50 tablets (Ashbourne Pharmaceuticals Ltd) |
| 24280 | 212825001000027107 | Atenolol/ Chlortalidone | Totaretic 100mg+25mg Tablet (C P Pharmaceuticals Ltd) |
| 26248 | 298311000001107 | Atenolol/ Chlortalidone | Tenchlor 100mg/ 25mg tablets (Teva UK Ltd) |
| 26741 | 212815001000027103 | Atenolol/ Chlortalidone | Totaretic 50mg+12.5mg Tablet (C P Pharmaceuticals Ltd) |
| 31470 | 439411000001104 | Atenolol/ Chlortalidone | Tenchlor 50mg/ 12.5mg tablets (Teva UK Ltd) |
| 31708 | 653811000001101 | Atenolol/ Chlortalidone | Co-tenidone 50mg/ 12.5mg tablets (Actavis UK Ltd) |
| 32094 | 884711000001106 | Atenolol/ Chlortalidone | Co-tenidone 50mg/ 12.5mg tablets (A A H Pharmaceuticals Ltd) |
| 34012 | 134211000001101 | Atenolol/ Chlortalidone | Co-tenidone 100mg/ 25mg tablets (IVAX Pharmaceuticals UK Ltd) |
| 34034 | 576211000001104 | Atenolol/ Chlortalidone | Co-tenidone 50mg/ 12.5mg tablets (IVAX Pharmaceuticals UK Ltd) |
| 34449 | 587011000001102 | Atenolol/ Chlortalidone | Co-tenidone 50mg/ 12.5mg tablets (Mylan) |
| 34825 | 143311000001108 | Atenolol/ Chlortalidone | Co-tenidone 50mg/ 12.5mg tablets (Teva UK Ltd) |
| 34899 | 793911000001107 | Atenolol/ Chlortalidone | Co-tenidone 100mg/ 25mg tablets (A A H Pharmaceuticals Ltd) |
| 37725 | 594311000001107 | Atenolol/ Chlortalidone | Co-tenidone 100mg/ 25mg tablets (Mylan) |
| 41572 | 485811000001105 | Atenolol/ Chlortalidone | Co-tenidone 100mg/ 25mg tablets (Teva UK Ltd) |
| 46952 | 588911000001102 | Atenolol/ Chlortalidone | Co-tenidone 100mg/ 25mg tablets (Actavis UK Ltd) |
| 62537 | 23858011000001104 | Atenolol/ Chlortalidone | Co-tenidone 100mg/ 25mg tablets (DE Pharmaceuticals) |
| 77414 | | Atenolol/ Chlortalidone | Tenoretic 100mg/ 25mg tablets (Dowelhurst Ltd) |
| 77466 | | Atenolol/ Chlortalidone | Co-tenidone 50mg/ 12.5mg tablets (Kent Pharmaceuticals Ltd) |
| 9178 | 318560004 | Atenolol/ Bendroflumethiazide | Atenolol 25mg / Bendroflumethiazide 1.25mg capsules |
| 18743 | 721411000001106 | Atenolol/ Bendroflumethiazide | Tenben 25mg/ 1.25mg capsules (Galen Ltd) |
| 3526 | 159515001000027103 | Atenolol/ Amiloride / Hydrochlorothiazide | Amiloride with atenolol with hydrochlorothiazide capsules |
| 4983 | 156785001000027105 | Atenolol/ Amiloride / Hydrochlorothiazide | Atenolol with amiloride and hydrochlorothiazide capsules |
| 7543 | 237011000001100 | Atenolol/ Amiloride / Hydrochlorothiazide | Kalten capsules (M & A Pharmachem Ltd) |

| | | | |
|---|---|---|---|
| 28177 | 15675500100002710З | Atenolol/ Amiloride / Hydrochlorothiazide | Hydrochlorothiazide with atenolol and amiloride Capsule |
| 8189 | 878911000001106 | Acebutolol / Hydrochlorothiazide | Secadrex 200mg/ 12.5mg tablets (Sanofi) |
| 14126 | 318586009 | Acebutolol / Hydrochlorothiazide | Acebutolol 200mg / Hydrochlorothiazide 12.5mg tablets |
| 8765 | 156575001000027104 | | ATENOLOL/ CHLORTHALIDONE 50 MG TAB |
| 19003 | 132525001000027106 | | SPIRONOLACTONE/ PROPRANOLOL 50 MG TAB |
| 22151 | | | METOPROLOL 100MG/ CHLORTHALIDONE 12.5MG |
| 24520 | 145355001000027108 | | HYDROCHLOROTHIAZIDE / METOPROLOL TARTRATE 25 MG TAB |
| 25764 | | | PINDOLOL 10MG/ CLOPAMIDE 5MG |
| 27086 | 188055001000027109 | | NADOLOL 80MG/ BENDROFLUAZIDE 5MG MG TAB |
| 41892 | | | METOPROLOL 100MG/ HYDROCHLOROTHIAZ.12.5MG |
| **Calcium channel blockers** | | | |
| 700 | 7418511000001104 | Verapamil | Vera-Til SR 120mg tablets (Tillomed Laboratories Ltd) |
| 1118 | 318204006 | Verapamil | Verapamil 40mg tablets |
| 1120 | 318205007 | Verapamil | Verapamil 80mg tablets |
| 1298 | 318213008 | Verapamil | Verapamil 240mg modified-release tablets |
| 1574 | 36565011000001105 | Verapamil | Verapamil 120mg modified-release capsules |
| 1747 | 318206008 | Verapamil | Verapamil 120mg tablets |
| 1748 | 867011000001103 | Verapamil | Cordilox 120mg tablets (IVAX Pharmaceuticals UK Ltd) |
| 3057 | 140411000001109 | Verapamil | Securon 120mg tablets (Abbott Laboratories Ltd) |
| 3342 | 540111000001108 | Verapamil | Securon SR 240mg tablets (Mylan) |
| 3343 | 846611000001102 | Verapamil | Half Securon SR 120mg tablets (Mylan) |
| 3943 | 318250009 | Verapamil | Verapamil 240mg modified-release capsules |
| 6510 | 219711000001103 | Verapamil | Univer 120mg modified-release capsules (Teva UK Ltd) |
| 8524 | 81585001000027101 | Verapamil | Securon 40mg Tablet (Abbott Laboratories Ltd) |
| 8759 | 68645001000027109 | Verapamil | Verapamil 120mg modified release tablets |
| 8884 | 557111000001106 | Verapamil | Cordilox 40mg tablets (IVAX Pharmaceuticals UK Ltd) |
| 8945 | 65411000001102 | Verapamil | Univer 240mg modified-release capsules (Teva UK Ltd) |
| 8975 | 36149411000001103 | Verapamil | Verapamil 180mg modified-release capsules |
| 9569 | 35367911000001103 | Verapamil | Verapamil 120mg modified-release tablets |
| 10688 | 318248001 | Verapamil | Verapamil 160mg tablets |
| 10832 | 81595001000027102 | Verapamil | Securon 80mg Tablet (Abbott Laboratories Ltd) |
| 11777 | 4139411000001106 | Verapamil | Verapamil 40mg/ 5ml oral solution sugar free |
| 11972 | 675611000001104 | Verapamil | Vertab SR 240 tablets (Chiesi Ltd) |
| 12104 | 783511000001101 | Verapamil | Cordilox 160mg tablets (IVAX Pharmaceuticals UK Ltd) |
| 12392 | 391811000001102 | Verapamil | Univer 180mg modified-release capsules (Teva UK Ltd) |
| 13251 | 7418711000001109 | Verapamil | Vera-Til SR 240mg tablets (Tillomed Laboratories Ltd) |
| 13856 | 7855011000001100 | Verapamil | Verapress MR 240mg tablets (Actavis UK Ltd) |
| 13965 | 740811000001102 | Verapamil | Cordilox MR 240mg tablets (Teva UK Ltd) |
| 16328 | 525311000001101 | Verapamil | Verapress MR 240mg tablets (Dexcel-Pharma Ltd) |
| 16677 | 258911000001104 | Verapamil | Cordilox 80mg tablets (IVAX Pharmaceuticals UK Ltd) |
| 17599 | 7852611000001103 | Verapamil | Verapress MR 240mg tablets (Sandoz Ltd) |

| | | | |
|---|---|---|---|
| 19175 | 422111000001103 | Verapamil | Verapamil 40mg tablets (IVAX Pharmaceuticals UK Ltd) |
| 19325 | 6105001000027101 | Verapamil | Cordilox 2.5mg/ ml Injection (IVAX Pharmaceuticals UK Ltd) |
| 19457 | 3634111000001100 | Verapamil | Ranvera MR 240mg tablets (Ranbaxy (UK) Ltd) |
| 19459 | 4579511000001103 | Verapamil | Verapamil 240mg modified-release tablets (A A H Pharmaceuticals Ltd) |
| 22826 | 69645001000027106 | Verapamil | Securon 160mg Tablet (Abbott Laboratories Ltd) |
| 23872 | 44155001000027103 | Verapamil | Berkatens 40mg Tablet (Berk Pharmaceuticals Ltd) |
| 25059 | 44165001000027107 | Verapamil | Berkatens 80mg Tablet (Berk Pharmaceuticals Ltd) |
| 26252 | 44225001000027102 | Verapamil | Berkatens 160mg Tablet (Berk Pharmaceuticals Ltd) |
| 26674 | 36149611000001100 | Verapamil | Verapamil 5mg/ 2ml solution for injection ampoules |
| 27295 | 4399811000001108 | Verapamil | Securon IV 5mg/ 2ml solution for injection ampoules (Mylan) |
| 28843 | 22045001000027100 | Verapamil | Verapamil hc 80mg Tablet (Celltech Pharma Europe Ltd) |
| 28844 | 44175001000027100 | Verapamil | Berkatens 120mg Tablet (Berk Pharmaceuticals Ltd) |
| 29637 | 10910611000001109 | Verapamil | Verapress MR 240mg tablets (Teva UK Ltd) |
| 30462 | 782111000001104 | Verapamil | Ethimil MR 240mg tablets (Genus Pharmaceuticals Ltd) |
| 31490 | 4129511000001107 | Verapamil | Zolvera 40mg/ 5ml oral solution (Rosemont Pharmaceuticals Ltd) |
| 31711 | 100911000001107 | Verapamil | Verapamil 80mg tablets (A A H Pharmaceuticals Ltd) |
| 32590 | 90311000001104 | Verapamil | Verapamil 40mg tablets (Mylan) |
| 33471 | 797711000001100 | Verapamil | Verapamil 40mg tablets (Actavis UK Ltd) |
| 34959 | 413611000001109 | Verapamil | Verapamil 40mg tablets (A A H Pharmaceuticals Ltd) |
| 35729 | 279811000001100 | Verapamil | Verapamil 80mg tablets (Teva UK Ltd) |
| 39009 | 454211000001101 | Verapamil | Verapamil 40mg tablets (Teva UK Ltd) |
| 40405 | 590011000001102 | Verapamil | Verapamil 120mg tablets (Teva UK Ltd) |
| 41586 | 7911000001103 | Verapamil | Verapamil 80mg tablets (Actavis UK Ltd) |
| 41679 | 534011000001101 | Verapamil | Verapamil 80mg tablets (IVAX Pharmaceuticals UK Ltd) |
| 41693 | 485611000001106 | Verapamil | Verapamil 120mg tablets (Mylan) |
| 42625 | 16449211000001100 | Verapamil | Vera-Til SR 120mg tablets (Actavis UK Ltd) |
| 43879 | 16438111000001103 | Verapamil | Vera-Til SR 240mg tablets (Actavis UK Ltd) |
| 45051 | 66545001000027106 | Verapamil | Verapamil hc 240mg Modified-release tablet (Actavis UK Ltd) |
| 45308 | 9208311000001103 | Verapamil | Verapamil 240mg modified-release tablets (Mylan) |
| 46009 | 887611000001108 | Verapamil | Verapamil 120mg tablets (Kent Pharmaceuticals Ltd) |
| 46884 | 136715001000027100 | Verapamil | Verapamil hc 240mg Modified-release tablet (Sandoz Ltd) |
| 46955 | 803411000001104 | Verapamil | Verapamil 80mg tablets (Mylan) |
| 47222 | 13924111000001102 | Verapamil | Verapamil 120mg modified-release tablets (A A H Pharmaceuticals Ltd) |
| 47230 | 9527111000001101 | Verapamil | Verapamil 240mg modified-release tablets (Teva UK Ltd) |
| 51461 | 5407511000001101 | Verapamil | Securon SR 240mg tablets (Waymade Healthcare Plc) |
| 59264 | 18611011000001108 | Verapamil | Securon SR 240mg tablets (DE Pharmaceuticals) |
| 62552 | 223411000001100 | Verapamil | Verapamil 80mg tablets (Alliance Healthcare (Distribution) Ltd) |
| 67293 | 18633611000001108 | Verapamil | Half Securon SR 120mg tablets (Mawdsley-Brooks & Company Ltd) |
| 68531 | 13015411000001109 | Verapamil | Verapamil 40mg/ 5ml oral suspension |
| 71009 | 81311000001105 | Verapamil | Verapamil 40mg tablets (Kent Pharmaceuticals Ltd) |
| 73467 | 87665001000027108 | Verapamil | Verapamil hc 80mg Tablet (Ranbaxy (UK) Ltd) |
| 76547 | | Verapamil | Verapamil 80mg tablets (Waymade Healthcare Plc) |
| 76554 | | Verapamil | Verapamil 120mg modified-release tablets (DE Pharmaceuticals) |

| | | | |
|---|---|---|---|
| 77506 | | Verapamil | Verapamil 240mg modified-release tablets (Kent Pharmaceuticals Ltd) |
| 77557 | | Verapamil | Securon SR 240mg tablets (Mawdsley-Brooks & Company Ltd) |
| 29116 | 237645001000027104 | Perhexiline Maleate | Perhexiline maleate 100mg tablet |
| 61230 | 504305001000027109 | Perhexiline Maleate | Perhexiline maleate 100mg tablets |
| 75641 | | Perhexiline Maleate | Pexsig 100mg tablets (Imported (New Zealand)) |
| 18038 | 36031911000001102 | Nisoldipine | Nisoldipine 20mg modified-release tablets |
| 19129 | 3877011000001100 | Nisoldipine | Syscor MR 10 tablets (Forest Laboratories UK Ltd) |
| 23805 | 36031811000001107 | Nisoldipine | Nisoldipine 10mg modified-release tablets |
| 23823 | 36032011000001109 | Nisoldipine | Nisoldipine 30mg modified-release tablets |
| 31336 | 4069211000001108 | Nisoldipine | Syscor MR 30 tablets (Forest Laboratories UK Ltd) |
| 31337 | 4070011000001100 | Nisoldipine | Syscor MR 20 tablets (Forest Laboratories UK Ltd) |
| 10595 | 3879211000001104 | Nimodipine | Nimotop 30mg tablets (Bayer Plc) |
| 11547 | 323273000 | Nimodipine | Nimodipine 30mg tablets |
| 269 | 319222004 | Nifedipine | Nifedipine 5mg capsules |
| 410 | 319274001 | Nifedipine | Nifedipine 10mg modified-release tablets |
| 452 | 319223009 | Nifedipine | Nifedipine 10mg capsules |
| 541 | 881811000001100 | Nifedipine | Adalat LA 20mg tablets (Bayer Plc) |
| 662 | 27111000001107 | Nifedipine | Adalat 5mg capsules (Bayer Plc) |
| 737 | 319273007 | Nifedipine | Nifedipine 20mg modified-release capsules |
| 1262 | 129175001000027109 | Nifedipine | Nifedipine 12 20mg Modified-release tablet |
| 1300 | 55365001000027100 | Nifedipine | Nifensar xl 20mg Modified-release tablet (Rhone-Poulenc Rorer Ltd) |
| 1449 | 214905001000027101 | Nifedipine | Nifedipine 24 30mg Modified-release tablet |
| 1854 | 125735001000027108 | Nifedipine | Adalat la 30mg Tablet (Bayer Plc) |
| 2280 | 569011000001108 | Nifedipine | Adalat retard 10mg tablets (Bayer Plc) |
| 2343 | 5011000001109 | Nifedipine | Adalat retard 20mg tablets (Bayer Plc) |
| 2521 | 782511000001108 | Nifedipine | Adalat 10mg capsules (Bayer Plc) |
| 2605 | 319272002 | Nifedipine | Nifedipine 10mg modified-release capsules |
| 2746 | 126411000001108 | Nifedipine | Coracten SR 10mg capsules (UCB Pharma Ltd) |
| 3711 | 74111000001103 | Nifedipine | Adipine MR 20 tablets (Chiesi Ltd) |
| 3712 | 162811000001100 | Nifedipine | Coracten XL 30mg capsules (UCB Pharma Ltd) |
| 3930 | 319276004 | Nifedipine | Nifedipine 60mg modified-release tablets |
| 4227 | 125745001000027109 | Nifedipine | Adalat la 60mg Tablet (Bayer Plc) |
| 4239 | 833611000001109 | Nifedipine | Adipine MR 10 tablets (Chiesi Ltd) |
| 4856 | 389611000001101 | Nifedipine | Coracten SR 20mg capsules (UCB Pharma Ltd) |
| 4939 | 3381511000001105 | Nifedipine | Coracten XL 60mg capsules (UCB Pharma Ltd) |
| 5162 | 319247002 | Nifedipine | Nifedipine 30mg modified-release capsules |
| 5181 | 865011000001105 | Nifedipine | Angiopine MR 20mg tablets (Ashbourne Pharmaceuticals Ltd) |
| 5277 | 188711000001108 | Nifedipine | Fortipine LA 40 tablets (AMCo) |
| 5806 | 385611000001104 | Nifedipine | Tensipine MR 20 tablets (Thornton & Ross Ltd) |
| 7541 | 280811000001100 | Nifedipine | Nifopress Retard 20mg tablets (AMCo) |
| 8213 | 197535001000027103 | Nifedipine | Nifedipine 24 20mg Modified-release tablet |
| 9269 | 319233001 | Nifedipine | Nifedipine 40mg modified-release tablets |
| 9485 | 677411000001108 | Nifedipine | Hypolar Retard 20 tablets (Sandoz Ltd) |

| | | | |
|---|---|---|---|
| 9553 | 630411000001107 | Nifedipine | Slofedipine XL 60 tablets (Zentiva) |
| 9573 | 2882011000001104 | Nifedipine | Slofedipine XL 30mg tablets (Zentiva) |
| 9750 | 319248007 | Nifedipine | Nifedipine 60mg modified-release capsules |
| 10135 | 235325001000027108 | Nifedipine | Nifedipress mr 10mg Modified-release tablet (Sandoz Ltd) |
| 10136 | 619111000001101 | Nifedipine | Nifedipress MR 20 tablets (Dexcel-Pharma Ltd) |
| 10246 | 9049911000001105 | Nifedipine | Adipine XL 60mg tablets (Chiesi Ltd) |
| 11512 | 904011000001104 | Nifedipine | Nifedipress MR 10 tablets (Dexcel-Pharma Ltd) |
| 11769 | 17011000001100 | Nifedipine | Calchan MR 20 tablets (Ranbaxy (UK) Ltd) |
| 12606 | 199885001000027103 | Nifedipine | Nifelease 20mg Modified-release tablet (Eastern Pharmaceuticals Ltd) |
| 12613 | 215915001000027109 | Nifedipine | Unipine xl 30mg Modified-release tablet (Genus Pharmaceuticals Ltd) |
| 13139 | 9049711000001108 | Nifedipine | Adipine XL 30mg tablets (Chiesi Ltd) |
| 13672 | 568911000001104 | Nifedipine | Angiopine MR 10mg tablets (Ashbourne Pharmaceuticals Ltd) |
| 13699 | 222575001000027106 | Nifedipine | Angiopine la 40mg Tablet (Ashbourne Pharmaceuticals Ltd) |
| 14861 | 627111000001104 | Nifedipine | Calchan MR 10 tablets (Ranbaxy (UK) Ltd) |
| 15715 | 230875001000027104 | Nifedipine | Genalat retard 20mg Modified-release tablet (Wyeth Pharmaceuticals) |
| 16073 | 7856211000001104 | Nifedipine | Nifedipress MR 10 tablets (Teva UK Ltd) |
| 17325 | 25911000001100 | Nifedipine | Cardilate MR 10mg tablets (Teva UK Ltd) |
| 17338 | 227175001000027101 | Nifedipine | Nifedotard 20 mr 20mg Modified-release tablet (Galen Ltd) |
| 17342 | 234015001000027109 | Nifedipine | Nivaten retard 10mg Modified-release tablet (Actavis UK Ltd) |
| 17448 | 226035001000027105 | Nifedipine | Nifedipress mr 10mg Modified-release tablet (Sterwin Medicines) |
| 19170 | 413111000001101 | Nifedipine | Tensipine MR 10 tablets (Thornton & Ross Ltd) |
| 20257 | 905711000001103 | Nifedipine | Cardilate MR 20mg tablets (IVAX Pharmaceuticals UK Ltd) |
| 20311 | 77775001000027106 | Nifedipine | Nifedipress mr 20mg Modified-release tablet (Generics (UK) Ltd) |
| 20591 | 7856611000001102 | Nifedipine | Nifedipress MR 20 tablets (Teva UK Ltd) |
| 20878 | 811311000001107 | Nifedipine | Angiopine 10 capsules (Ashbourne Pharmaceuticals Ltd) |
| 21216 | 4878111000001101 | Nifedipine | Hypolar Retard 10mg tablets (Sandoz Ltd) |
| 21245 | 232035001000027107 | Nifedipine | Nifedipress mr 10mg Modified-release tablet (Actavis UK Ltd) |
| 21872 | 169635001000027100 | Nifedipine | Angiopine 5mg Capsule (Ashbourne Pharmaceuticals Ltd) |
| 21886 | 7855311000001102 | Nifedipine | Nifedipress MR 20 tablets (Actavis UK Ltd) |
| 22019 | 210265001000027101 | Nifedipine | Calanif 10mg Capsule (Berk Pharmaceuticals Ltd) |
| 22142 | 170255001000027101 | Nifedipine | Calcilat 10mg Capsule (Eastern Pharmaceuticals Ltd) |
| 22217 | 222315001000027101 | Nifedipine | Nimodrel 10mg modified-release tablet (Opus Pharmaceuticals Ltd) |
| 22696 | 843411000001105 | Nifedipine | Slofedipine 20mg tablets (Sterwin Medicines) |
| 23736 | 2881811000001101 | Nifedipine | Hypolar XL 30 tablets (Sandoz Ltd) |
| 24228 | 222325001000027105 | Nifedipine | Nimodrel 20mg modified-release tablet (Opus Pharmaceuticals Ltd) |
| 25132 | 9485111000001105 | Nifedipine | Nifopress MR 20mg tablets (Teva UK Ltd) |
| 25646 | 234025001000027100 | Nifedipine | Nivaten retard 20mg Modified-release tablet (Actavis UK Ltd) |
| 25919 | 10054311000001109 | Nifedipine | Nifedipine 20mg modified-release tablets (A A H Pharmaceuticals Ltd) |
| 26265 | 210275001000027108 | Nifedipine | Calanif 5mg Capsule (Berk Pharmaceuticals Ltd) |
| 26774 | 242975001000027102 | Nifedipine | Nifedipine 10mg/ 5ml Oral suspension |
| 28688 | 10054111000001107 | Nifedipine | Nifedipine 10mg modified-release tablets (A A H Pharmaceuticals Ltd) |
| 30199 | 319275000 | Nifedipine | Nifedipine 30mg modified-release tablets |
| 30473 | 309611000001102 | Nifedipine | Coroday MR 20mg tablets (Mylan) |

| | | | |
|---|---|---|---|
| 33025 | 10189111000001106 | Nifedipine | Nimodrel XL 30mg tablets (Zurich Pharmaceuticals) |
| 34101 | 103085001000027109 | Nifedipine | Nifedipine mr 20mg Modified-release tablet (IVAX Pharmaceuticals UK Ltd) |
| 34115 | 155025001000027103 | Nifedipine | Nifedipine 60mg Modified-release tablet |
| 34146 | 103075001000027104 | Nifedipine | Nifedipine mr 10mg Modified-release tablet (IVAX Pharmaceuticals UK Ltd) |
| 34187 | 93635001000027105 | Nifedipine | Nifedipine 10mg Modified-release tablet (Generics (UK) Ltd) |
| 34247 | 8795001000027103 | Nifedipine | Nifedipine 10mg Capsule (Berk Pharmaceuticals Ltd) |
| 34522 | 720511000001103 | Nifedipine | Nifedipine 5mg capsules (A A H Pharmaceuticals Ltd) |
| 34607 | 814611000001100 | Nifedipine | Nifedipine 5mg capsules (IVAX Pharmaceuticals UK Ltd) |
| 34975 | 168411000001100 | Nifedipine | Nifedipine 5mg capsules (Teva UK Ltd) |
| 35646 | 10751411000001101 | Nifedipine | Neozipine XL 60mg tablets (Kent Pharmaceuticals Ltd) |
| 37025 | 319277008 | Nifedipine | Nifedipine 20mg modified-release tablets |
| 37184 | 13401911000001109 | Nifedipine | Valni XL 30mg tablets (Zentiva) |
| 37530 | 10751211000001100 | Nifedipine | Neozipine XL 30mg tablets (Kent Pharmaceuticals Ltd) |
| 37726 | 8670011000001105 | Nifedipine | Nifedipine 100mg/ 5ml oral suspension |
| 38107 | 185705001000027105 | Nifedipine | Nifedipine sr 30mg Tablet (Hillcross Pharmaceuticals Ltd) |
| 39800 | 13402111000001101 | Nifedipine | Valni XL 60mg tablets (Zentiva) |
| 40074 | 129135001000027107 | Nifedipine | Nifedipine 20mg Capsule |
| 41979 | 232235001000027104 | Nifedipine | Adipine la 30mg Modified-release tablet (Chiesi Ltd) |
| 42912 | 295511000001109 | Nifedipine | Nifedipine 10mg capsules (Teva UK Ltd) |
| 43222 | 693311000001101 | Nifedipine | Valni 20 Retard tablets (Tillomed Laboratories Ltd) |
| 43410 | 197555001000027104 | Nifedipine | Nifedipine extra 60mg Modified-release tablet |
| 43511 | 498311000001106 | Nifedipine | Nifedipine 10mg capsules (A A H Pharmaceuticals Ltd) |
| 43515 | 639811000001100 | Nifedipine | Nifedipine 10mg capsules (Actavis UK Ltd) |
| 43753 | 2881311000001105 | Nifedipine | Adalat LA 30mg tablets (Bayer Plc) |
| 43818 | 235511000001104 | Nifedipine | Adalat LA 60mg tablets (Bayer Plc) |
| 45685 | 17666011000001106 | Nifedipine | Adanif XL 30mg tablets (Focus Pharmaceuticals Ltd) |
| 46445 | 652811000001109 | Nifedipine | Nifedipine 10mg capsules (IVAX Pharmaceuticals UK Ltd) |
| 46887 | 17666211000001101 | Nifedipine | Adanif XL 60mg tablets (Focus Pharmaceuticals Ltd) |
| 47027 | 116995001000027105 | Nifedipine | Nifedipine 10mg Modified-release tablet (Kent Pharmaceuticals Ltd) |
| 47217 | 232245001000027103 | Nifedipine | Adipine la 60mg Modified-release tablet (Chiesi Ltd) |
| 47285 | 185845001000027103 | Nifedipine | Nifedipine xl 60mg Tablet (Hillcross Pharmaceuticals Ltd) |
| 47529 | 9096811000001100 | Nifedipine | Nifedipine 20mg/ ml oral drops |
| 47614 | 10639711000001103 | Nifedipine | Nifedipine 30mg modified-release tablets (A A H Pharmaceuticals Ltd) |
| 47707 | 247555001000027108 | Nifedipine | Nifedipine Oral solution |
| 47887 | 10189311000001108 | Nifedipine | Nimodrel XL 60mg tablets (Zurich Pharmaceuticals) |
| 49338 | 10056111000001100 | Nifedipine | Nifedipine 20mg modified-release tablets (Alliance Healthcare (Distribution) Ltd) |
| 49762 | 10055911000001109 | Nifedipine | Nifedipine 10mg modified-release tablets (Alliance Healthcare (Distribution) Ltd) |
| 51917 | 14043411000001104 | Nifedipine | Adalat LA 60 tablets (Sigma Pharmaceuticals Plc) |
| 52017 | 17489111000001103 | Nifedipine | Adalat LA 30 tablets (Mawdsley-Brooks & Company Ltd) |
| 53278 | 17574611000001108 | Nifedipine | Adalat LA 30 tablets (Necessity Supplies Ltd) |
| 53357 | 8670111000001106 | Nifedipine | Nifedipine 10mg/ 5ml oral suspension |
| 53500 | 13824711000001105 | Nifedipine | Adalat LA 30 tablets (DE Pharmaceuticals) |
| 53629 | 16135011000001106 | Nifedipine | Adalat retard 20mg tablets (Lexon (UK) Ltd) |

| | | | |
|---|---|---|---|
| 53990 | 8670311000001108 | Nifedipine | Nifedipine 5mg/ 5ml oral suspension |
| 55455 | 18283911000001107 | Nifedipine | Nifedipine 10mg capsules (Strides Shasun (UK) Ltd) |
| 55824 | 88575001000027100 | Nifedipine | Nifedipine 20mg Modified-release tablet (Berk Pharmaceuticals Ltd) |
| 56469 | 17575011000001102 | Nifedipine | Adalat LA 60 tablets (Necessity Supplies Ltd) |
| 57531 | 5335911000001104 | Nifedipine | Adalat LA 60 tablets (Waymade Healthcare Plc) |
| 57653 | 14043011000001108 | Nifedipine | Adalat LA 20 tablets (Sigma Pharmaceuticals Plc) |
| 58557 | 17574211000001106 | Nifedipine | Adalat LA 20 tablets (Necessity Supplies Ltd) |
| 58990 | 21868911000001101 | Nifedipine | Nifedipine 10mg modified-release tablets (Cubic Pharmaceuticals Ltd) |
| 59163 | 21868511000001108 | Nifedipine | Nifedipine 20mg modified-release tablets (Cubic Pharmaceuticals Ltd) |
| 60856 | 24102511000001108 | Nifedipine | Nifedipine 10mg modified-release tablets (Sigma Pharmaceuticals Plc) |
| 63041 | 868811000001108 | Nifedipine | Nifedipine 10mg capsules (Mylan) |
| 63246 | 28407711000001101 | Nifedipine | Nifedipine 10mg modified-release tablets (AM Distributions (Yorkshire) Ltd) |
| 66191 | 13825311000001105 | Nifedipine | Adalat retard 20mg tablets (DE Pharmaceuticals) |
| 66236 | 117035001000027106 | Nifedipine | Nifedipine 20mg Modified-release tablet (Kent Pharmaceuticals Ltd) |
| 67074 | 13824311000001106 | Nifedipine | Adalat 10mg capsules (DE Pharmaceuticals) |
| 69202 | 12862411000001109 | Nifedipine | Nifedipine 2mg/ 5ml oral suspension |
| 69668 | 12862511000001108 | Nifedipine | Nifedipine 30mg/ 5ml oral suspension |
| 71339 | 5439111000001108 | Nifedipine | Adalat LA 30 tablets (Dowelhurst Ltd) |
| 71601 | 12303311000001109 | Nifedipine | Nifedipine 2.5mg/ 5ml oral suspension |
| 71653 | 34685811000001103 | Nifedipine | Nidef 60mg modified-release tablets (Morningside Healthcare Ltd) |
| 71760 | 24615511000001100 | Nifedipine | Nifedipine 10mg modified-release tablets (Ethigen Ltd) |
| 72253 | 24102911000001101 | Nifedipine | Nifedipine 30mg modified-release tablets (Sigma Pharmaceuticals Plc) |
| 72321 | 34685211000001104 | Nifedipine | Nidef 30mg modified-release tablets (Morningside Healthcare Ltd) |
| 73992 | 5440311000001102 | Nifedipine | Adalat LA 60 tablets (Dowelhurst Ltd) |
| 74001 | 10459411000001108 | Nifedipine | Adalat LA 20 tablets (Dowelhurst Ltd) |
| 74061 | 5335411000001107 | Nifedipine | Adalat LA 30 tablets (Waymade Healthcare Plc) |
| 74330 | 26854411000001105 | Nifedipine | Nifedipine 20mg modified-release tablets (Ennogen Healthcare Ltd) |
| 74648 | 8835001000027107 | Nifedipine | Nifedipine 10mg Capsule (C P Pharmaceuticals Ltd) |
| 75109 | 88585001000027105 | Nifedipine | Nifedipine 10mg Modified-release tablet (Berk Pharmaceuticals Ltd) |
| 76488 | | Nifedipine | Nifedipine 5mg capsules (Strides Pharma UK Ltd) |
| 76539 | | Nifedipine | Nifedipin-ratiopharm 20mg/ ml oral drops (Imported (Germany)) |
| 77134 | | Nifedipine | Adalat 10mg capsules (Lexon (UK) Ltd) |
| 77418 | | Nifedipine | Adalat retard 20mg tablets (Waymade Healthcare Plc) |
| 77447 | | Nifedipine | Adalat 10mg capsules (Necessity Supplies Ltd) |
| 77555 | | Nifedipine | Adalat LA 30 tablets (Sigma Pharmaceuticals Plc) |
| 77962 | | Nifedipine | Adalat LA 20 tablets (Lexon (UK) Ltd) |
| 2926 | 319217004 | Nicardipine | Nicardipine 20mg capsules |
| 3302 | 540311000001105 | Nicardipine | Cardene SR 30mg capsules (Astellas Pharma Ltd) |
| 5477 | 319216008 | Nicardipine | Nicardipine 30mg modified-release capsules |
| 7562 | 291111000001102 | Nicardipine | Cardene 30mg capsules (Astellas Pharma Ltd) |
| 8201 | 319218009 | Nicardipine | Nicardipine 30mg capsules |
| 9386 | 319215007 | Nicardipine | Nicardipine 45mg modified-release capsules |
| 11943 | 344811000001108 | Nicardipine | Cardene 20mg capsules (Astellas Pharma Ltd) |

| | | | |
|---|---|---|---|
| 12875 | 118811000001102 | Nicardipine | Cardene SR 45mg capsules (Astellas Pharma Ltd) |
| 45292 | 743711000001101 | Nicardipine | Nicardipine 30mg capsules (A A H Pharmaceuticals Ltd) |
| 73646 | 13760311000001107 | Nicardipine | Nicardipine 20mg capsules (Tillomed Laboratories Ltd) |
| 73968 | 3656311000001105 | Nicardipine | Nicardipine 20mg capsules (Teva UK Ltd) |
| 1529 | 226015001000027104 | Mibefradil | Posicor 50mg Tablet (Roche Products Ltd) |
| 3931 | 226025001000027108 | Mibefradil | Posicor 100mg Tablet (Roche Products Ltd) |
| 15652 | 226705001000027103 | Mibefradil | Mibefradil 50mg Tablet |
| 22241 | 226715001000027101 | Mibefradil | Mibefradil 100mg Tablet |
| 19013 | 155925001000027108 | Lidoflazine | Clinium 120mg Tablet (LEO Pharma) |
| 30758 | 155905001000027101 | Lidoflazine | Lidoflazine 120mg Tablet |
| 5570 | 20011000001105 | Lercanidipine | Zanidip 10mg tablets (Recordati Pharmaceuticals Ltd) |
| 5593 | 319316005 | Lercanidipine | Lercanidipine 10mg tablets |
| 13243 | 10225911000001102 | Lercanidipine | Lercanidipine 20mg tablets |
| 14300 | 10198711000001102 | Lercanidipine | Zanidip 20mg tablets (Recordati Pharmaceuticals Ltd) |
| 47331 | 16666611000001103 | Lercanidipine | Lercanidipine 10mg tablets (Mylan) |
| 56767 | 16666411000001101 | Lercanidipine | Lercanidipine 20mg tablets (Mylan) |
| 57444 | 22337311000001108 | Lercanidipine | Lercanidipine 10mg tablets (Aptil Pharma Ltd) |
| 59233 | 16607211000001104 | Lercanidipine | Lercanidipine 20mg tablets (Actavis UK Ltd) |
| 61611 | 24363711000001103 | Lercanidipine | Lercanidipine 10mg tablets (DE Pharmaceuticals) |
| 63917 | 16732211000001104 | Lercanidipine | Lercanidipine 20mg tablets (A A H Pharmaceuticals Ltd) |
| 64227 | 16606811000001100 | Lercanidipine | Lercanidipine 10mg tablets (Actavis UK Ltd) |
| 64424 | 16640011000001100 | Lercanidipine | Lercanidipine 20mg tablets (Zentiva) |
| 65659 | 16726111000001106 | Lercanidipine | Lercanidipine 20mg tablets (Teva UK Ltd) |
| 69239 | 16732011000001109 | Lercanidipine | Lercanidipine 10mg tablets (A A H Pharmaceuticals Ltd) |
| 70732 | 15608611000001100 | Lercanidipine | Lercanidipine 20mg tablets (Alliance Healthcare (Distribution) Ltd) |
| 70827 | 16725911000001102 | Lercanidipine | Lercanidipine 10mg tablets (Teva UK Ltd) |
| 71018 | 20911211000001100 | Lercanidipine | Lercanidipine 10mg tablets (Arrow Generics Ltd) |
| 71030 | 20911411000001101 | Lercanidipine | Lercanidipine 20mg tablets (Arrow Generics Ltd) |
| 76672 | | Lercanidipine | Lercanidipine 20mg tablets (Sigma Pharmaceuticals Plc) |
| 77363 | | Lercanidipine | Zanidip 10mg tablets (Dowelhurst Ltd) |
| 3221 | 319301007 | Lacidipine | Lacidipine 4mg tablets |
| 5158 | 319300008 | Lacidipine | Lacidipine 2mg tablets |
| 9670 | 333011000001100 | Lacidipine | Motens 4mg tablets (GlaxoSmithKline UK Ltd) |
| 11966 | 910911000001108 | Lacidipine | Motens 2mg tablets (GlaxoSmithKline UK Ltd) |
| 56994 | 18565211000001108 | Lacidipine | Lacidipine 4mg tablets (Teva UK Ltd) |
| 57680 | 18642611000001102 | Lacidipine | Lacidipine 4mg tablets (A A H Pharmaceuticals Ltd) |
| 60699 | 14736811000001107 | Lacidipine | Lacidipine 2mg tablets (Sigma Pharmaceuticals Plc) |
| 70990 | 18642411000001100 | Lacidipine | Lacidipine 2mg tablets (A A H Pharmaceuticals Ltd) |
| 77450 | | Lacidipine | Motens 2mg tablets (Lexon (UK) Ltd) |
| 8257 | 3689711000001108 | Isradipine | Prescal 2.5mg tablets (Novartis Pharmaceuticals UK Ltd) |
| 8310 | 319280009 | Isradipine | Isradipine 2.5mg tablets |
| 491 | 319291002 | Felodipine | Felodipine 2.5mg modified-release tablets |
| 501 | 319287007 | Felodipine | Felodipine 5mg modified-release tablets |

| | | | |
|---|---|---|---|
| 568 | 319288002 | Felodipine | Felodipine 10mg modified-release tablets |
| 7280 | 48511000001101 | Felodipine | Plendil 10mg modified-release tablets (AstraZeneca UK Ltd) |
| 9334 | 562711000001104 | Felodipine | Plendil 2.5mg modified-release tablets (AstraZeneca UK Ltd) |
| 9437 | 490211000001101 | Felodipine | Plendil 5mg modified-release tablets (AstraZeneca UK Ltd) |
| 10153 | 235995001000027107 | Felodipine | Felendil xl 5mg Modified-release tablet (Ratiopharm UK Ltd) |
| 14305 | 5638811000001106 | Felodipine | Vascalpha 10mg modified-release tablets (Actavis UK Ltd) |
| 17557 | 4785111000001103 | Felodipine | Felotens XL 5mg tablets (Thornton & Ross Ltd) |
| 17566 | 4785511000001107 | Felodipine | Felotens XL 10mg tablets (Thornton & Ross Ltd) |
| 20459 | 236005001000027102 | Felodipine | Felendil xl 10mg Modified-release tablet (Ratiopharm UK Ltd) |
| 24365 | 7887011000001104 | Felodipine | Cardioplen XL 5mg tablets (Chiesi Ltd) |
| 24366 | 7887511000001107 | Felodipine | Cardioplen XL 10mg tablets (Chiesi Ltd) |
| 25572 | 4972811000001103 | Felodipine | Felogen XL 5mg tablets (Mylan) |
| 26337 | 3800711000001105 | Felodipine | Cabren 10mg modified-release tablets (Teva UK Ltd) |
| 28721 | 8090111000001106 | Felodipine | Neofel XL 5mg tablets (Kent Pharmaceuticals Ltd) |
| 29044 | 8089811000001107 | Felodipine | Neofel XL 10mg tablets (Kent Pharmaceuticals Ltd) |
| 29145 | 243075001000027100 | Felodipine | Felendil xl 2.5mg Modified-release tablet (Ratiopharm UK Ltd) |
| 30557 | 4973011000001100 | Felodipine | Felogen XL 10mg tablets (Mylan) |
| 30915 | 3800311000001106 | Felodipine | Cabren 2.5mg modified-release tablets (Teva UK Ltd) |
| 30991 | 3800511000001100 | Felodipine | Cabren 5mg modified-release tablets (Teva UK Ltd) |
| 32922 | 152805001000027108 | Felodipine | Felodipine 10mg Modified-release tablet (Sandoz Ltd) |
| 33091 | 16183811000001106 | Felodipine | Felodipine 10mg modified-release tablets (A A H Pharmaceuticals Ltd) |
| 33932 | 7387911000001103 | Felodipine | Parmid XL 5mg tablets (Sandoz Ltd) |
| 35084 | 5638311000001102 | Felodipine | Vascalpha 5mg modified-release tablets (Actavis UK Ltd) |
| 35592 | 11506711000001103 | Felodipine | Cardioplen XL 2.5mg tablets (Chiesi Ltd) |
| 36620 | 7388311000001103 | Felodipine | Parmid XL 10mg tablets (Sandoz Ltd) |
| 37897 | 13127311000001107 | Felodipine | Felotens XL 2.5mg tablets (Thornton & Ross Ltd) |
| 38434 | 10284611000001100 | Felodipine | Keloc SR 10mg tablets (Teva UK Ltd) |
| 39357 | 11493211000001105 | Felodipine | Neofel XL 2.5mg tablets (Kent Pharmaceuticals Ltd) |
| 40633 | 18250911000001108 | Felodipine | Vascalpha 5mg modified-release tablets (Almus Pharmaceuticals Ltd) |
| 43394 | 9359611000001103 | Felodipine | Pinefeld XL 10mg tablets (Tillomed Laboratories Ltd) |
| 43512 | 16183611000001107 | Felodipine | Felodipine 5mg modified-release tablets (A A H Pharmaceuticals Ltd) |
| 43790 | 18252511000001100 | Felodipine | Vascalpha 10mg modified-release tablets (Almus Pharmaceuticals Ltd) |
| 44859 | 142895001000027103 | Felodipine | Felodipine sr 5mg Tablet (Approved Prescription Services Ltd) |
| 48009 | 152785001000027109 | Felodipine | Felodipine 5mg Modified-release tablet (Sandoz Ltd) |
| 55306 | 5008511000001107 | Felodipine | Folpik XL 5mg tablets (Teva UK Ltd) |
| 55740 | 18167311000001103 | Felodipine | Neofel XL 2.5mg tablets (Actavis UK Ltd) |
| 58339 | 18681211000001100 | Felodipine | Neofel XL 2.5mg tablets (Almus Pharmaceuticals Ltd) |
| 60569 | 23594111000001104 | Felodipine | Felodipine 2.5mg modified-release tablets (Waymade Healthcare Plc) |
| 60652 | 24221811000001105 | Felodipine | Parmid XL 2.5mg tablets (Sandoz Ltd) |
| 60884 | 23916011000001102 | Felodipine | Felodipine 2.5mg modified-release tablets (Phoenix Healthcare Distribution Ltd) |
| 63331 | 13565311000001101 | Felodipine | Folpik XL 2.5mg tablets (Teva UK Ltd) |
| 64474 | 16183411000001109 | Felodipine | Felodipine 2.5mg modified-release tablets (A A H Pharmaceuticals Ltd) |
| 64504 | 17621211000001102 | Felodipine | Plendil 5mg modified-release tablets (Necessity Supplies Ltd) |

| | | | |
|---|---|---|---|
| 64719 | 29860311000001105 | Felodipine | Felodipine 2.5mg modified-release tablets (Sigma Pharmaceuticals Plc) |
| 64760 | 23915811000001100 | Felodipine | Felodipine 10mg modified-release tablets (Phoenix Healthcare Distribution Ltd) |
| 64917 | 23594511000001108 | Felodipine | Felodipine 10mg modified-release tablets (Waymade Healthcare Plc) |
| 65349 | 5008911000001100 | Felodipine | Folpik XL 10mg tablets (Teva UK Ltd) |
| 66095 | 30127711000001105 | Felodipine | Felodipine 5mg modified-release tablets (Mawdsley-Brooks & Company Ltd) |
| 66910 | 30822411000001105 | Felodipine | Felodipine 2.5mg modified-release tablets (DE Pharmaceuticals) |
| 68181 | 29860511000001104 | Felodipine | Felodipine 5mg modified-release tablets (Sigma Pharmaceuticals Plc) |
| 68499 | 14293501000027102 | Felodipine | Felodipine sr 10mg Tablet (Approved Prescription Services Ltd) |
| 68828 | 23977111000001107 | Felodipine | Felodipine 5mg modified-release tablets (DE Pharmaceuticals) |
| 69206 | 18029311000001107 | Felodipine | Felodipine 2.5mg/ 5ml oral solution |
| 72181 | 18029411000001100 | Felodipine | Felodipine 5mg/ 5ml oral solution |
| 72221 | 23594311000001102 | Felodipine | Felodipine 5mg modified-release tablets (Waymade Healthcare Plc) |
| 73715 | 23915611000001104 | Felodipine | Felodipine 5mg modified-release tablets (Phoenix Healthcare Distribution Ltd) |
| 74004 | 5575011000001104 | Felodipine | Plendil 5mg modified-release tablets (Dowelhurst Ltd) |
| 74012 | 5401711000001102 | Felodipine | Plendil 2.5mg modified-release tablets (Waymade Healthcare Plc) |
| 75345 | 34447411000001102 | Felodipine | Felodipine 2.5mg/ 5ml oral suspension |
| 219 | 418945005 | Diltiazem | Diltiazem 120mg modified-release tablets |
| 517 | 71075001000027107 | Diltiazem | Adizem sr 120mg Modified-release capsule (Napp Pharmaceuticals Ltd) |
| 536 | 111175001000027101 | Diltiazem | Tildiem la 200mg Modified-release capsule (Sanofi) |
| 636 | 319185009 | Diltiazem | Diltiazem 60mg modified-release capsules |
| 793 | 196035001000027100 | Diltiazem | Adizem xl 240mg Capsule (Napp Pharmaceuticals Ltd) |
| 939 | 103611000001105 | Diltiazem | Tildiem Retard 90mg tablets (Sanofi) |
| 1130 | 407011000001105 | Diltiazem | Viazem XL 300mg capsules (Thornton & Ross Ltd) |
| 1289 | 383911000001109 | Diltiazem | Tildiem Retard 120mg tablets (Sanofi) |
| 1538 | 155445001000027109 | Diltiazem | Diltiazem 60mg tablets |
| 1686 | 319182007 | Diltiazem | Diltiazem 90mg modified-release capsules |
| 1836 | 319205001 | Diltiazem | Diltiazem 60mg modified-release tablets |
| 1995 | 71715001000027101 | Diltiazem | Diltiazem 12hr 120mg modified-release capsules |
| 2453 | 155455001000027107 | Diltiazem | Diltiazem 60mg modified-release capsules |
| 2528 | 130211000001109 | Diltiazem | Slozem 120mg capsules (Merck Serono Ltd) |
| 2592 | 32311000001105 | Diltiazem | Viazem XL 120mg capsules (Thornton & Ross Ltd) |
| 2663 | 181565001000027106 | Diltiazem | Diltiazem 240mg modified-release capsules |
| 2686 | 196445001000027108 | Diltiazem | Dilzem xl mr 240mg Modified-release capsule (Elan Pharma) |
| 2811 | 71085001000027102 | Diltiazem | Adizem sr 180mg Modified-release capsule (Napp Pharmaceuticals Ltd) |
| 2888 | 762011000001102 | Diltiazem | Tildiem 60mg modified-release tablets (Sanofi) |
| 3061 | 71725001000027105 | Diltiazem | Diltiazem 12hr 180mg modified-release capsules |
| 3118 | 181545001000027100 | Diltiazem | Adizem sr 90mg Modified-release capsule (Napp Pharmaceuticals Ltd) |
| 3370 | 196425001000027105 | Diltiazem | Dilzem xl mr 120mg Modified-release capsule (Elan Pharma) |
| 3676 | 196435001000027107 | Diltiazem | Dilzem xl mr 180mg Modified-release capsule (Elan Pharma) |
| 4308 | 193225001000027104 | Diltiazem | Dilzem sr 90mg Capsule (Elan Pharma) |
| 4408 | 599811000001104 | Diltiazem | Slozem 240mg capsules (Merck Serono Ltd) |
| 4635 | 319187001 | Diltiazem | Diltiazem 200mg modified-release capsules |
| 4732 | 319180004 | Diltiazem | Diltiazem 90mg modified-release tablets |

| | | | |
|---|---|---|---|
| 4808 | 319186005 | Diltiazem | Diltiazem 240mg modified-release capsules |
| 4852 | 181525001000027103 | Diltiazem | Adizem sr 120mg Modified-release tablet (Napp Pharmaceuticals Ltd) |
| 4923 | 197585001000027106 | Diltiazem | Diltiazem 24hr 180mg modified-release capsules |
| 5054 | 336611000001101 | Diltiazem | Angitil SR 180 capsules (Ethypharm UK Ltd) |
| 5194 | 193235001000027102 | Diltiazem | Dilzem sr 120mg Capsule (Elan Pharma) |
| 5234 | 119211000001108 | Diltiazem | Slozem 180mg capsules (Merck Serono Ltd) |
| 5296 | 111165001000027108 | Diltiazem | Tildiem la 300mg Modified-release capsule (Sanofi) |
| 5326 | 198095001000027102 | Diltiazem | Diltiazem 24hr 300mg modified-release capsules |
| 5348 | 319181000 | Diltiazem | Diltiazem 300mg modified-release capsules |
| 5513 | 193215001000027108 | Diltiazem | Dilzem sr 60mg Capsule (Elan Pharma) |
| 6309 | 71095001000027103 | Diltiazem | Adizem xl 300mg Capsule (Napp Pharmaceuticals Ltd) |
| 7398 | 648211000001107 | Diltiazem | Viazem XL 360mg capsules (Thornton & Ross Ltd) |
| 8558 | 196015001000027106 | Diltiazem | Adizem xl 120mg Capsule (Napp Pharmaceuticals Ltd) |
| 9240 | 196025001000027102 | Diltiazem | Adizem xl 180mg Capsule (Napp Pharmaceuticals Ltd) |
| 9374 | 181535001000027101 | Diltiazem | Adizem 60mg Modified-release tablet (Napp Pharmaceuticals Ltd) |
| 9410 | 298811000001103 | Diltiazem | Angitil SR 120 capsules (Ethypharm UK Ltd) |
| 9708 | 197575001000027101 | Diltiazem | Diltiazem 24hr 120mg modified-release capsules |
| 9723 | 219611000001107 | Diltiazem | Calcicard CR 90mg tablets (Teva UK Ltd) |
| 10267 | 2886511000001108 | Diltiazem | Adizem-XL 200mg capsules (Napp Pharmaceuticals Ltd) |
| 11223 | 857011000001109 | Diltiazem | Angitil SR 90 capsules (Ethypharm UK Ltd) |
| 11770 | 417111000001109 | Diltiazem | Dilzem SR 60 capsules (Teva UK Ltd) |
| 11922 | 8456911000001108 | Diltiazem | Diltiazem 60mg/ 5ml oral suspension |
| 11973 | 104111000001100 | Diltiazem | Calcicard CR 120mg tablets (Teva UK Ltd) |
| 12639 | 70315001000027107 | Diltiazem | Diltiazem 90mg Modified-release tablet (Actavis UK Ltd) |
| 12705 | 440711000001104 | Diltiazem | Angiozem CR 90mg tablets (Ashbourne Pharmaceuticals Ltd) |
| 13027 | 886511000001107 | Diltiazem | Viazem XL 240mg capsules (Thornton & Ross Ltd) |
| 13033 | 75111000001104 | Diltiazem | Angitil XL 240 capsules (Ethypharm UK Ltd) |
| 13075 | 254911000001106 | Diltiazem | Dilzem XL 180 capsules (Teva UK Ltd) |
| 13127 | 733511000001107 | Diltiazem | Dilzem XL 240 capsules (Teva UK Ltd) |
| 13240 | 243111000001108 | Diltiazem | Dilzem XL 120 capsules (Teva UK Ltd) |
| 13302 | 682311000001105 | Diltiazem | Dilzem SR 90 capsules (Teva UK Ltd) |
| 13410 | 672311000001103 | Diltiazem | Angiozem 60mg modified-release tablets (Ashbourne Pharmaceuticals Ltd) |
| 13926 | 319198000 | Diltiazem | Diltiazem 360mg modified-release capsules |
| 15221 | 199355001000027107 | Diltiazem | Dilcardia xl 180mg Modified-release capsule (Generics (UK) Ltd) |
| 15288 | 467111000001105 | Diltiazem | Angitil XL 300 capsules (Ethypharm UK Ltd) |
| 16038 | 5711000001106 | Diltiazem | Dilzem SR 120 capsules (Teva UK Ltd) |
| 16850 | 144811000001101 | Diltiazem | Angiozem CR 120mg tablets (Ashbourne Pharmaceuticals Ltd) |
| 17406 | 813611000001103 | Diltiazem | Zemtard 180 XL capsules (Galen Ltd) |
| 17425 | 105211000001107 | Diltiazem | Zemtard 120 XL capsules (Galen Ltd) |
| 17492 | 866811000001107 | Diltiazem | Zemtard 300 XL capsules (Galen Ltd) |
| 17586 | 550211000001104 | Diltiazem | Slozem 300mg capsules (Merck Serono Ltd) |
| 17666 | 33211000001108 | Diltiazem | Viazem XL 180mg capsules (Thornton & Ross Ltd) |
| 18379 | 353011000001101 | Diltiazem | Dilcardia SR 90mg capsules (Mylan) |

| | | | |
|---|---|---|---|
| 18403 | 95745001000027106 | Diltiazem | Diltiazem 180mg Modified-release capsule (Hillcross Pharmaceuticals Ltd) |
| 18404 | 4772111000001102 | Diltiazem | Diltiazem 60mg modified-release capsules (A A H Pharmaceuticals Ltd) |
| 18830 | 256611000001109 | Diltiazem | Disogram SR 90mg capsules (Ranbaxy (UK) Ltd) |
| 18834 | 469811000001106 | Diltiazem | Disogram SR 60mg capsules (Ranbaxy (UK) Ltd) |
| 18852 | 116711000001106 | Diltiazem | Disogram SR 120mg capsules (Ranbaxy (UK) Ltd) |
| 18874 | 713411000001103 | Diltiazem | Disogram SR 180mg capsules (Ranbaxy (UK) Ltd) |
| 18975 | 152715001000027107 | Diltiazem | Calcicard 60mg Tablet (3M Health Care Ltd) |
| 19426 | 108711000001107 | Diltiazem | Disogram SR 240mg capsules (Ranbaxy (UK) Ltd) |
| 19440 | 34711000001104 | Diltiazem | Disogram SR 300mg capsules (Ranbaxy (UK) Ltd) |
| 20642 | 228225001000027107 | Diltiazem | Bi-carzem sr 60mg Modified-release capsule (Tillomed Laboratories Ltd) |
| 20890 | 345411000001107 | Diltiazem | Zemtard 240 XL capsules (Galen Ltd) |
| 21145 | 937011000001101 | Diltiazem | Dilcardia SR 60mg capsules (Mylan) |
| 21763 | 215411000001106 | Diltiazem | Diltiazem 60mg modified-release tablets (A A H Pharmaceuticals Ltd) |
| 21773 | 96705001000027103 | Diltiazem | Diltiazem 60mg Tablet (Generics (UK) Ltd) |
| 21778 | 391911000001107 | Diltiazem | Diltiazem 60mg modified-release tablets (Teva UK Ltd) |
| 21795 | 865711000001107 | Diltiazem | Retalzem 60 modified-release tablets (Kent Pharmaceuticals Ltd) |
| 21918 | 527711000001108 | Diltiazem | Optil 60mg modified-release tablets (Opus Pharmaceuticals Ltd) |
| 22619 | 155475001000027105 | Diltiazem | Britiazim 60mg Modified-release tablet (Thames Laboratories Ltd) |
| 23233 | 228235001000027109 | Diltiazem | Bi-carzem sr 90mg Modified-release capsule (Tillomed Laboratories Ltd) |
| 23733 | 159965001000027107 | Diltiazem | Optil sr 90mg Modified-release capsule (Opus Pharmaceuticals Ltd) |
| 25777 | 723811000001107 | Diltiazem | Dilcardia SR 120mg capsules (Mylan) |
| 26267 | 159975001000027100 | Diltiazem | Optil sr 120mg Modified-release capsule (Opus Pharmaceuticals Ltd) |
| 26269 | 159985001000027105 | Diltiazem | Optil sr 180mg Modified-release capsule (Opus Pharmaceuticals Ltd) |
| 26270 | 203585001000027108 | Diltiazem | Optil xl 300mg Modified-release capsule (Opus Pharmaceuticals Ltd) |
| 26309 | 203575001000027103 | Diltiazem | Optil xl 240mg Modified-release capsule (Opus Pharmaceuticals Ltd) |
| 26460 | 199365001000027103 | Diltiazem | Dilcardia xl 240mg Modified-release capsule (Generics (UK) Ltd) |
| 26463 | 215655001000027103 | Diltiazem | Zemret xl 240mg Capsule (Neo Laboratories Ltd) |
| 26759 | 721811000001108 | Diltiazem | Zildil SR 60mg capsules (Chanelle Medical UK Ltd) |
| 27135 | 155305001000027109 | Diltiazem | Diltiazem sr 90mg Capsule (Hillcross Pharmaceuticals Ltd) |
| 27136 | 10065211000001107 | Diltiazem | Diltiazem 90mg modified-release tablets (A A H Pharmaceuticals Ltd) |
| 27401 | 8886211000001101 | Diltiazem | Kenzem SR 90mg capsules (Kent Pharmaceuticals Ltd) |
| 27685 | 129475001000027105 | Diltiazem | Diltiazem 300mg Capsule (PLIVA Pharma Ltd) |
| 28949 | 228245001000027105 | Diltiazem | Bi-carzem sr 120mg Modified-release capsule (Tillomed Laboratories Ltd) |
| 29676 | 214485001000027100 | Diltiazem | Calazem 60mg Modified-release tablet (Berk Pharmaceuticals Ltd) |
| 30197 | 319183002 | Diltiazem | Diltiazem 120mg modified-release capsules |
| 30242 | 319184008 | Diltiazem | Diltiazem 180mg modified-release capsules |
| 31489 | 233535001000027103 | Diltiazem | Bi-carzem xl 240mg Capsule (Tillomed Laboratories Ltd) |
| 31676 | 70325001000027103 | Diltiazem | Diltiazem 120mg Modified-release tablet (Actavis UK Ltd) |
| 31737 | 655411000001104 | Diltiazem | Zildil SR 120mg capsules (Chanelle Medical UK Ltd) |
| 32089 | 12155001000027107 | Diltiazem | Diltiazem 120mg Modified-release capsule (Hillcross Pharmaceuticals Ltd) |
| 32262 | 12115001000027102 | Diltiazem | Diltiazem 60mg Tablet (C P Pharmaceuticals Ltd) |
| 32658 | 199345001000027109 | Diltiazem | Dilcardia xl 120mg Modified-release capsule (Generics (UK) Ltd) |
| 32870 | 673611000001103 | Diltiazem | Diltiazem 60mg modified-release tablets (Sterwin Medicines) |

| | | | |
|---|---|---|---|
| 34377 | 95755001000027109 | Diltiazem | Diltiazem 90mg Modified-release capsule (Hillcross Pharmaceuticals Ltd) |
| 34475 | 45985001000027103 | Diltiazem | Diltiazem 90mg Modified-release tablet (IVAX Pharmaceuticals UK Ltd) |
| 34581 | 152545001000027104 | Diltiazem | Diltiazem 60mg Modified-release tablet (Kent Pharmaceuticals Ltd) |
| 34824 | 45995001000027104 | Diltiazem | Diltiazem 120mg Modified-release tablet (IVAX Pharmaceuticals UK Ltd) |
| 35696 | 8886511000001103 | Diltiazem | Kenzem SR 120mg capsules (Kent Pharmaceuticals Ltd) |
| 36583 | 215645001000027101 | Diltiazem | Zemret xl 180mg Capsule (Neo Laboratories Ltd) |
| 36664 | 215665001000027107 | Diltiazem | Zemret xl 300mg Capsule (Neo Laboratories Ltd) |
| 37774 | 8885711000001100 | Diltiazem | Kenzem SR 60mg capsules (Kent Pharmaceuticals Ltd) |
| 38066 | 89675001000027109 | Diltiazem | Diltiazem 60mg Modified-release tablet (Lagap) |
| 38545 | 261611000001107 | Diltiazem | Tildiem LA 200 capsules (Sanofi) |
| 38632 | 2887011000001102 | Diltiazem | Adizem-SR 90mg capsules (Napp Pharmaceuticals Ltd) |
| 38634 | 2886111000001104 | Diltiazem | Adizem-XL 300mg capsules (Napp Pharmaceuticals Ltd) |
| 38818 | 2887311000001104 | Diltiazem | Adizem-SR 120mg capsules (Napp Pharmaceuticals Ltd) |
| 38831 | 2886711000001103 | Diltiazem | Adizem-SR 180mg capsules (Napp Pharmaceuticals Ltd) |
| 38855 | 2938011000001101 | Diltiazem | Adizem-XL 180mg capsules (Napp Pharmaceuticals Ltd) |
| 38865 | 2937811000001108 | Diltiazem | Adizem-XL 120mg capsules (Napp Pharmaceuticals Ltd) |
| 38876 | 893111000001107 | Diltiazem | Tildiem LA 300 capsules (Sanofi) |
| 38882 | 2886311000001102 | Diltiazem | Adizem-XL 240mg capsules (Napp Pharmaceuticals Ltd) |
| 38964 | 2885611000001102 | Diltiazem | Adizem-SR 120mg tablets (Napp Pharmaceuticals Ltd) |
| 39171 | 540011000001107 | Diltiazem | Bi-Carzem SR 60mg capsules (Tillomed Laboratories Ltd) |
| 39298 | 764511000001100 | Diltiazem | Bi-Carzem SR 90mg capsules (Tillomed Laboratories Ltd) |
| 41489 | 580711000001101 | Diltiazem | Bi-Carzem SR 120mg capsules (Tillomed Laboratories Ltd) |
| 41635 | 544111000001104 | Diltiazem | Diltiazem 60mg modified-release tablets (IVAX Pharmaceuticals UK Ltd) |
| 42731 | 155325001000027102 | Diltiazem | Diltiazem sr 120mg Capsule (Hillcross Pharmaceuticals Ltd) |
| 42804 | 129425001000027106 | Diltiazem | Diltiazem 180mg Capsule (PLIVA Pharma Ltd) |
| 42819 | 163525001000027109 | Diltiazem | Diltiazem xl 240mg Capsule (Hillcross Pharmaceuticals Ltd) |
| 43430 | 10065011000001102 | Diltiazem | Diltiazem 120mg modified-release tablets (A A H Pharmaceuticals Ltd) |
| 44192 | 823511000001107 | Diltiazem | Zemret 240 XL capsules (Tillomed Laboratories Ltd) |
| 44887 | 233545001000027102 | Diltiazem | Bi-carzem xl 300mg Capsule (Tillomed Laboratories Ltd) |
| 45759 | 129455001000027107 | Diltiazem | Diltiazem 240mg Capsule (PLIVA Pharma Ltd) |
| 46937 | 896111000001102 | Diltiazem | Diltiazem 60mg modified-release tablets (Actavis UK Ltd) |
| 47415 | 203125001000027107 | Diltiazem | Diltiazem sr 60mg Capsule (Hillcross Pharmaceuticals Ltd) |
| 47530 | 591411000001102 | Diltiazem | Horizem SR 60mg capsules (Horizon lifecare) |
| 47608 | 640311000001101 | Diltiazem | Zemret 300 XL capsules (Tillomed Laboratories Ltd) |
| 47724 | 884811000001103 | Diltiazem | Bi-Carzem XL 240mg capsules (Tillomed Laboratories Ltd) |
| 47732 | 924711000001109 | Diltiazem | Zemret 180 XL capsules (Tillomed Laboratories Ltd) |
| 48272 | 10054911000001105 | Diltiazem | Diltiazem 60mg modified-release capsules (Alliance Healthcare (Distribution) Ltd) |
| 48282 | 18311411000001106 | Diltiazem | Diltiazem 90mg modified-release capsules (A A H Pharmaceuticals Ltd) |
| 48288 | 18311611000001109 | Diltiazem | Diltiazem 120mg modified-release capsules (A A H Pharmaceuticals Ltd) |
| 48457 | 10055111000001106 | Diltiazem | Diltiazem 90mg modified-release capsules (Alliance Healthcare (Distribution) Ltd) |
| 48870 | 19871811000001100 | Diltiazem | Adizem-SR 90mg capsules (DE Pharmaceuticals) |
| 49001 | 10055311000001108 | Diltiazem | Diltiazem 120mg modified-release tablets (Alliance Healthcare (Distribution) Ltd) |
| 49289 | 10054711000001108 | Diltiazem | Diltiazem 120mg modified-release capsules (Alliance Healthcare (Distribution) Ltd) |

| | | | |
|---|---|---|---|
| 49390 | 10055511000001102 | Diltiazem | Diltiazem 90mg modified-release tablets (Alliance Healthcare (Distribution) Ltd) |
| 51261 | 18481611000001101 | Diltiazem | Tildiem Retard 120mg tablets (Mawdsley-Brooks & Company Ltd) |
| 52276 | 19872011000001103 | Diltiazem | Adizem-XL 180mg capsules (DE Pharmaceuticals) |
| 52701 | 18481911000001107 | Diltiazem | Tildiem LA 200 capsules (Mawdsley-Brooks & Company Ltd) |
| 54799 | 17465311000001101 | Diltiazem | Tildiem LA 300 capsules (Mawdsley-Brooks & Company Ltd) |
| 55257 | 8457011000001107 | Diltiazem | Diltiazem 60mg/ 5ml oral solution |
| 56467 | 14005111000001104 | Diltiazem | Tildiem 60mg modified-release tablets (DE Pharmaceuticals) |
| 56758 | 21864611000001102 | Diltiazem | Diltiazem 90mg modified-release capsules (Cubic Pharmaceuticals Ltd) |
| 57208 | 21864811000001103 | Diltiazem | Diltiazem 120mg modified-release capsules (Cubic Pharmaceuticals Ltd) |
| 57594 | 5424211000001104 | Diltiazem | Tildiem 60mg modified-release tablets (Waymade Healthcare Plc) |
| 57859 | 21865011000001108 | Diltiazem | Diltiazem 90mg modified-release tablets (Cubic Pharmaceuticals Ltd) |
| 59098 | 16158611000001109 | Diltiazem | Dilzem XL 180 capsules (Lexon (UK) Ltd) |
| 59585 | 21965711000001100 | Diltiazem | Uard 120XL capsules (Ennogen Healthcare Ltd) |
| 59863 | 16158811000001108 | Diltiazem | Dilzem XL 240 capsules (Lexon (UK) Ltd) |
| 60415 | 17537211000001101 | Diltiazem | Dilzem XL 180 capsules (Sigma Pharmaceuticals Plc) |
| 60620 | 23929811000001107 | Diltiazem | Adizem-XL 240mg capsules (Waymade Healthcare Plc) |
| 61010 | 23586211000001104 | Diltiazem | Diltiazem 120mg modified-release tablets (Cubic Pharmaceuticals Ltd) |
| 61245 | 24597111000001107 | Diltiazem | Diltiazem 60mg modified-release capsules (Sigma Pharmaceuticals Plc) |
| 61532 | 24596711000001105 | Diltiazem | Diltiazem 120mg modified-release capsules (Sigma Pharmaceuticals Plc) |
| 62064 | 24523511000001104 | Diltiazem | Diltiazem 120mg modified-release tablets (Mawdsley-Brooks & Company Ltd) |
| 62065 | 24117911000001101 | Diltiazem | Diltiazem 90mg modified-release tablets (Colorama Pharmaceuticals Ltd) |
| 62207 | 23928711000001108 | Diltiazem | Adizem-SR 120mg capsules (Waymade Healthcare Plc) |
| 62912 | 28405011000001105 | Diltiazem | Diltiazem 120mg modified-release capsules (AM Distributions (Yorkshire) Ltd) |
| 65504 | 29955011000001104 | Diltiazem | Adizem-SR 180mg capsules (Lexon (UK) Ltd) |
| 65602 | 23929211000001106 | Diltiazem | Adizem-XL 120mg capsules (Waymade Healthcare Plc) |
| 65636 | 29955211000001109 | Diltiazem | Adizem-XL 120mg capsules (Lexon (UK) Ltd) |
| 66048 | 14005611000001107 | Diltiazem | Tildiem Retard 120mg tablets (DE Pharmaceuticals) |
| 66172 | 21965911000001103 | Diltiazem | Uard 180XL capsules (Ennogen Healthcare Ltd) |
| 66635 | 18574511000001102 | Diltiazem | Dilzem XL 180 capsules (DE Pharmaceuticals) |
| 66701 | 24101311000001106 | Diltiazem | Diltiazem 240mg modified-release capsules (DE Pharmaceuticals) |
| 66834 | 21966111000001107 | Diltiazem | Uard 240XL capsules (Ennogen Healthcare Ltd) |
| 66850 | 25711011000001108 | Diltiazem | Diltiazem 240mg modified-release capsules (Icarus Pharmaceuticals Ltd) |
| 67317 | 18189911000001107 | Diltiazem | Dilzem XL 120 capsules (Mawdsley-Brooks & Company Ltd) |
| 67344 | 24343511000001100 | Diltiazem | Diltiazem 300mg modified-release capsules (Ennogen Pharma Ltd) |
| 67890 | 21965511000001105 | Diltiazem | Uard 300XL capsules (Ennogen Healthcare Ltd) |
| 68054 | 526711000001105 | Diltiazem | Diltiazem 60mg modified-release tablets (Alliance Healthcare (Distribution) Ltd) |
| 68429 | 24615311000001106 | Diltiazem | Diltiazem 120mg modified-release capsules (Ethigen Ltd) |
| 69028 | 23969211000001102 | Diltiazem | Diltiazem 60mg modified-release capsules (DE Pharmaceuticals) |
| 69108 | 29956011000001108 | Diltiazem | Adizem-XL 300mg capsules (Lexon (UK) Ltd) |
| 69116 | 30139111000001101 | Diltiazem | Diltiazem 60mg modified-release tablets (DE Pharmaceuticals) |
| 69277 | 29955811000001105 | Diltiazem | Adizem-XL 240mg capsules (Lexon (UK) Ltd) |
| 70306 | 24524011000001109 | Diltiazem | Diltiazem 180mg modified-release capsules (Mawdsley-Brooks & Company Ltd) |
| 70961 | 14005411000001109 | Diltiazem | Tildiem Retard 90mg tablets (DE Pharmaceuticals) |

| | | | |
|---|---|---|---|
| 71342 | 10530911000001104 | Diltiazem | Dilzem XL 240 capsules (Waymade Healthcare Plc) |
| 71413 | 4773111000001108 | Diltiazem | Diltiazem 120mg modified-release capsules (A A H Pharmaceuticals Ltd) |
| 71702 | 8443011000001104 | Diltiazem | Diltiazem 60mg/ 5ml oral solution (Special Order) |
| 71969 | 24597911000001105 | Diltiazem | Diltiazem 300mg modified-release capsules (Sigma Pharmaceuticals Plc) |
| 72091 | 24102211000001105 | Diltiazem | Diltiazem 180mg modified-release capsules (DE Pharmaceuticals) |
| 72488 | 33424611000001105 | Diltiazem | Diltiazem 300mg modified-release capsules (A A H Pharmaceuticals Ltd) |
| 72839 | 24410111000001104 | Diltiazem | Diltiazem 300mg modified-release capsules (DE Pharmaceuticals) |
| 74022 | 5426711000001102 | Diltiazem | Tildiem LA 200 capsules (Waymade Healthcare Plc) |
| 74029 | 5619211000001106 | Diltiazem | Tildiem Retard 90mg tablets (Dowelhurst Ltd) |
| 74053 | 17537411000001102 | Diltiazem | Dilzem XL 240 capsules (Sigma Pharmaceuticals Plc) |
| 74689 | 24342811000001105 | Diltiazem | Diltiazem 240mg modified-release capsules (Ennogen Pharma Ltd) |
| 74915 | 22409111000001104 | Diltiazem | Diltiazem 120mg modified-release capsules (DE Pharmaceuticals) |
| 77077 | | Diltiazem | Diltiazem 10mg/ 5ml oral solution |
| 77442 | | Diltiazem | Tildiem 60mg modified-release tablets (Dowelhurst Ltd) |
| 77947 | | Diltiazem | Diltiazem 120mg/ 5ml oral solution |
| 29 | 182975001000027106 | Amlodipine | Amlodipine besilate 5mg tablets |
| 71 | 182985001000027101 | Amlodipine | Amlodipine besilate 10mg tablets |
| 729 | 237575001000027105 | Amlodipine | Amlodipine maleate 5mg tablets |
| 749 | 319283006 | Amlodipine | Amlodipine 5mg tablets |
| 3917 | 172711000001100 | Amlodipine | Istin 5mg tablets (Pfizer Ltd) |
| 5914 | 408111000001107 | Amlodipine | Istin 10mg tablets (Pfizer Ltd) |
| 6477 | 237585001000027100 | Amlodipine | Amlodipine maleate 10mg tablets |
| 6856 | 319284000 | Amlodipine | Amlodipine 10mg tablets |
| 16162 | 8278311000001107 | Amlodipine | Amlodipine 5mg/ 5ml oral suspension |
| 17640 | 8046211000001107 | Amlodipine | Amlostin 5mg tablets (Discovery Pharmaceuticals) |
| 31761 | 8046411000001106 | Amlodipine | Amlostin 10mg tablets (Discovery Pharmaceuticals) |
| 32595 | 7305311000001108 | Amlodipine | Amlodipine 5mg tablets (A A H Pharmaceuticals Ltd) |
| 32917 | 7305711000001107 | Amlodipine | Amlodipine 5mg tablets (IVAX Pharmaceuticals UK Ltd) |
| 34093 | 7305011000001105 | Amlodipine | Amlodipine 10mg tablets (A A H Pharmaceuticals Ltd) |
| 36202 | 8038411000001106 | Amlodipine | Amlodipine 10mg tablets (Actavis UK Ltd) |
| 39804 | 11008811000001105 | Amlodipine | Amlodipine 5mg tablets (Dr Reddy's Laboratories (UK) Ltd) |
| 39914 | 7333311000001100 | Amlodipine | Amlodipine 5mg tablets (Teva UK Ltd) |
| 42210 | 9557311000001100 | Amlodipine | Amlodipine 10mg tablets (Zentiva) |
| 43470 | 14768911000001108 | Amlodipine | Amlodipine 5mg tablets (Wockhardt UK Ltd) |
| 43880 | 11398511000001109 | Amlodipine | Amlodipine 5mg tablets (Almus Pharmaceuticals Ltd) |
| 45070 | 8278111000001105 | Amlodipine | Amlodipine 10mg/ 5ml oral suspension |
| 45279 | 7376311000001107 | Amlodipine | Amlodipine 5mg tablets (Sandoz Ltd) |
| 46233 | 243185001000027100 | Amlodipine | Amlodipine Oral solution |
| 46724 | 13892511000001100 | Amlodipine | Amlodipine 5mg/ 5ml oral solution |
| 47002 | 266105001000027102 | Amlodipine | Amlodipine 10mg/ 5ml sugar free Oral suspension |
| 49636 | 19704911000001101 | Amlodipine | Amlodipine 10mg tablets (DE Pharmaceuticals) |
| 52440 | 20478011000001105 | Amlodipine | Amlodipine 10mg/ 5ml oral solution |
| 53868 | 8038211000001107 | Amlodipine | Amlodipine 5mg tablets (Actavis UK Ltd) |

| | | | |
|---|---|---|---|
| 54515 | 7304811000001100 | Amlodipine | Amlodipine 10mg tablets (Alliance Healthcare (Distribution) Ltd) |
| 54633 | 15981411000001105 | Amlodipine | Amlodipine 5mg tablets (Bristol Laboratories Ltd) |
| 54654 | 11399011000001106 | Amlodipine | Amlodipine 10mg tablets (Almus Pharmaceuticals Ltd) |
| 54696 | 7376511000001101 | Amlodipine | Amlodipine 10mg tablets (Sandoz Ltd) |
| 54983 | 8278211000001104 | Amlodipine | Amlodipine 2.5mg/ 5ml oral suspension |
| 56147 | 18457811000001100 | Amlodipine | Amlodipine 10mg tablets (Accord Healthcare Ltd) |
| 56334 | 15981211000001106 | Amlodipine | Amlodipine 10mg tablets (Bristol Laboratories Ltd) |
| 58580 | 19833311000001108 | Amlodipine | Amlodipine 10mg tablets (APC Pharmaceuticals & Chemicals (Europe) Ltd) |
| 59001 | 7391311000001109 | Amlodipine | Amlodipine 10mg tablets (Mylan) |
| 59762 | 7333511000001106 | Amlodipine | Amlodipine 10mg tablets (Teva UK Ltd) |
| 60244 | 17779511000001109 | Amlodipine | Amlodipine 10mg tablets (Phoenix Healthcare Distribution Ltd) |
| 61374 | 11712011000001102 | Amlodipine | Amlodipine 4mg/ 5ml oral suspension |
| 61422 | 18458011000001107 | Amlodipine | Amlodipine 5mg tablets (Accord Healthcare Ltd) |
| 63515 | 11009211000001104 | Amlodipine | Amlodipine 10mg tablets (Dr Reddy's Laboratories (UK) Ltd) |
| 64166 | 29826311000001101 | Amlodipine | Amlodipine 5mg/ 5ml oral solution sugar free |
| 64327 | 7391211000001101 | Amlodipine | Amlodipine 5mg tablets (Mylan) |
| 64418 | 22080611000001107 | Amlodipine | Amlodipine 5mg tablets (Waymade Healthcare Plc) |
| 64441 | 7304611000001104 | Amlodipine | Amlodipine 5mg tablets (Alliance Healthcare (Distribution) Ltd) |
| 64447 | 19164711000001109 | Amlodipine | Amlodipine 5mg tablets (Somex Pharma) |
| 64606 | 10287111000001103 | Amlodipine | Amlodipine 5mg tablets (Focus Pharmaceuticals Ltd) |
| 64623 | 29826211000001109 | Amlodipine | Amlodipine 10mg/ 5ml oral solution sugar free |
| 65745 | 19705311000001103 | Amlodipine | Amlodipine 5mg tablets (DE Pharmaceuticals) |
| 66430 | 7378711000001105 | Amlodipine | Amlodipine 5mg tablets (Kent Pharmaceuticals Ltd) |
| 66574 | 17779111000001100 | Amlodipine | Amlodipine 5mg tablets (Phoenix Healthcare Distribution Ltd) |
| 66817 | 14769111000001103 | Amlodipine | Amlodipine 10mg tablets (Wockhardt UK Ltd) |
| 67662 | 13917011000001100 | Amlodipine | Istin 10mg tablets (DE Pharmaceuticals) |
| 68221 | 7378911000001107 | Amlodipine | Amlodipine 10mg tablets (Kent Pharmaceuticals Ltd) |
| 68311 | 29932511000001100 | Amlodipine | Amlodipine 5mg/ 5ml oral solution sugar free (A A H Pharmaceuticals Ltd) |
| 70999 | 19833111000001106 | Amlodipine | Amlodipine 5mg tablets (APC Pharmaceuticals & Chemicals (Europe) Ltd) |
| 71344 | 5332311000001106 | Amlodipine | Amlodipine 5mg tablets (Waymade Healthcare Plc) |
| 71353 | 5450111000001102 | Amlodipine | Amlodipine 5mg tablets (Dowelhurst Ltd) |
| 71939 | 10286711000001100 | Amlodipine | Amlodipine 10mg tablets (Focus Pharmaceuticals Ltd) |
| 72049 | 29992111000001103 | Amlodipine | Amlodipine 5mg tablets (Mawdsley-Brooks & Company Ltd) |
| 72199 | 35134611000001100 | Amlodipine | Amlodipine 5mg tablets (Aurobindo Pharma Ltd) |
| 72430 | 29781411000001107 | Amlodipine | Amlodipine 10mg/ 5ml oral solution sugar free (Alliance Healthcare (Distribution) Ltd) |
| 73612 | 30821811000001103 | Amlodipine | Amlodipine 5mg/ 5ml oral solution sugar free (DE Pharmaceuticals) |
| 74108 | 15064611000001101 | Amlodipine | Amlodipine 5mg tablets (Sigma Pharmaceuticals Plc) |
| 74121 | 30216911000001104 | Amlodipine | Amlodipine 5mg/ 5ml oral solution sugar free (Thame Laboratories Ltd) |
| 74795 | 19164911000001106 | Amlodipine | Amlodipine 10mg tablets (Somex Pharma) |
| 74820 | 15064211000001103 | Amlodipine | Amlodipine 10mg tablets (Sigma Pharmaceuticals Plc) |
| 74821 | 13764411000001106 | Amlodipine | Amlodipine 5mg tablets (Apotex UK Ltd) |
| 75387 | 10378211000001106 | Amlodipine | Amlodipine 5mg tablets (Arrow Generics Ltd) |
| 75609 | | Amlodipine | Amlodipine 5mg/ 5ml oral solution (Special Order) |

| | | | |
|---|---|---|---|
| 75862 | | Amlodipine | Amlodipine 5mg/ 5ml oral suspension sugar free |
| 76714 | | Amlodipine | Amlodipine 10mg/ 5ml oral solution sugar free (Thame Laboratories Ltd) |
| 77416 | | Amlodipine | Istin 10mg tablets (Dowelhurst Ltd) |
| 77420 | | Amlodipine | Istin 5mg tablets (DE Pharmaceuticals) |
| 77446 | | Amlodipine | Istin 10mg tablets (Waymade Healthcare Plc) |
| 7823 | 18899501000027108 | | NIFEDIPINE TAB 5 mg |
| 8024 | 9699501000027108 | | DILTIAZEM XL 300 MG CAP |
| 9094 | 11892501000027105 | | DILTIAZEM SR 300 MG CAP |
| 9211 | 9519501000027109 | | ADIZEM-XL 180 MG CAP |
| 10897 | 9623501000027102 | | VERAPAMIL S/ F 40 MG/ 5ML SOL |
| 15659 | 15148501000027101 | | DILTIAZEM S/ R 180 CAP |
| 18631 | | | VERAPAMIL MR |
| 18690 | 13344501000027107 | | SECURON (CALENDAR PACK) 120 MG TAB |
| 19015 | 17184501000027100 | | ADIZEM CONTINUS 120 MG TAB |
| 21496 | 16610501000027100 | | PERHEXILINE MALEATE 100 MG TAB |
| 21665 | | | CORDILOX |
| 23458 | | | VERAPAMIL SR |
| 23730 | 16327501000027103 | | VERAPAMIL 100 MG TAB |
| 25026 | | | NIFEDIPINE RETARD |
| 25027 | | | ADALAT RETARD 10 |
| 25044 | | | NIFEDIPINE RETARD |
| 25055 | | | NIFEDIPINE |
| 27910 | | | ADALAT 5 |
| 30491 | | | DILTIAZEM |
| 36684 | 19328501000027105 | | SLOFEDIPINE 20 MG TAB |
| **Diuretics** | | | |
| 4044 | 348911000001105 | Xipamide | Diurexan 20mg tablets (Meda Pharmaceuticals Ltd) |
| 7618 | 317970008 | Xipamide | Xipamide 20mg tablets |
| 9223 | 14514501000027101 | Triamterene/ Hydrochlorothiazide | Triamterene with hydrochlorothiazide 50mg + 25mg Tablet |
| 15127 | 11608501000027104 | Triamterene/ Hydrochlorothiazide | Hydrochlorothiazide with triamterene 25mgwith50mg Tablet |
| 2961 | 25411000001108 | Triamterene/ Furosemide | Frusene 50mg/ 40mg tablets (Orion Pharma (UK) Ltd) |
| 3050 | 10798501000027109 | Triamterene/ Furosemide | Furosemide with triamterene 40mgwith50mg Tablet |
| 11265 | 318101005 | Triamterene/ Furosemide | Triamterene 50mg / Furosemide 40mg tablets |
| 28157 | 4098501000027105 | Triamterene/ Chlortalidone | Kalspare ls Tablet (Dominion Pharma) |
| 37294 | 14526501000027105 | Triamterene/ Chlortalidone | Triamterene with chlortalidone 50mg + 25mg Tablet |
| 7136 | 714911000001108 | Triamterene/ Benzthiazide | Dytide capsules (Mercury Pharma Group Ltd) |
| 7740 | 318098006 | Triamterene/ Benzthiazide | Triamterene 50mg / Benzthiazide 25mg capsules |
| 2179 | 318082004 | Triamterene | Triamterene 50mg capsules |
| 4068 | 3907911000001100 | Triamterene | Dytac 50mg capsules (AMCo) |
| 8052 | 318041004 | Torasemide | Torasemide 5mg tablets |
| 10066 | 3700311000001109 | Torasemide | Torem 5mg tablets (Meda Pharmaceuticals Ltd) |
| 11268 | 3699711000001107 | Torasemide | Torem 2.5mg tablets (Meda Pharmaceuticals Ltd) |
| 11487 | 318040003 | Torasemide | Torasemide 2.5mg tablets |

| | | | |
|---|---|---|---|
| 18096 | 318042006 | Torasemide | Torasemide 10mg tablets |
| 22658 | 3700811000001100 | Torasemide | Torem 10mg tablets (Meda Pharmaceuticals Ltd) |
| 40190 | 19807500100002710 6 | Torasemide | Torasemide iv 20mg/ 4ml Intravenous injection |
| 40738 | 19802500100002710 7 | Torasemide | Torem iv 10mg/ 2ml Intravenous injection (Boehringer Mannheim UK Ltd) |
| 40898 | 5889911000001108 | Torasemide | Torasemide 5mg tablets (A A H Pharmaceuticals Ltd) |
| 46525 | 5592211000001107 | Torasemide | Torasemide 5mg tablets (Teva UK Ltd) |
| 1297 | 4669111000001107 | Spironolactone/ Hydroflumethiazide | Aldactide 50 tablets (Pfizer Ltd) |
| 2001 | 762511000001105 | Spironolactone/ Hydroflumethiazide | Aldactide 25 tablets (Pfizer Ltd) |
| 7961 | 14151500100002710 4 | Spironolactone/ Hydroflumethiazide | Spironolactone 50mg with hydroflumethiazide 50mg tablet |
| 8521 | 14150500100002710 1 | Spironolactone/ Hydroflumethiazide | Spironolactone 25mg with hydroflumethiazide 25mg tablet |
| 11384 | 318128000 | Spironolactone/ Hydroflumethiazide | Co-flumactone 50mg/ 50mg tablets |
| 15811 | 318127005 | Spironolactone/ Hydroflumethiazide | Co-flumactone 25mg/ 25mg tablets |
| 25505 | 19677500100002710 1 | Spironolactone/ Hydroflumethiazide | Spiro-co 50mg+50mg Tablet (IVAX Pharmaceuticals UK Ltd) |
| 29529 | 11622500100002710 1 | Spironolactone/ Hydroflumethiazide | Hydroflumethiazide with spironolactone 25mg+25mg Tablet |
| 31131 | 19676500100002710 8 | Spironolactone/ Hydroflumethiazide | Spiro-co 25mg+25mg Tablet (IVAX Pharmaceuticals UK Ltd) |
| 45916 | 11623500100002710 4 | Spironolactone/ Hydroflumethiazide | Hydroflumethiazide with spironolactone 50mg+50mg Tablet |
| 4661 | 318102003 | Spironolactone/ Furosemide | Spironolactone 50mg / Furosemide 20mg capsules |
| 7441 | 3645811000001107 | Spironolactone/ Furosemide | Lasilactone 20mg/ 50mg capsules (Sanofi) |
| 53508 | 12424811000001104 | Spironolactone/ Chlorothiazide | Spironolactone 5mg/ 5ml / Chlorothiazide 50mg/ 5ml oral suspension |
| 692 | 318056008 | Spironolactone | Spironolactone 25mg tablets |
| 708 | 318057004 | Spironolactone | Spironolactone 50mg tablets |
| 787 | 14147500100002710 3 | Spironolactone | Spironolactone 100mg capsule |
| 2142 | 318058009 | Spironolactone | Spironolactone 100mg tablets |
| 2389 | 930511000001105 | Spironolactone | Aldactone 25mg tablets (Pfizer Ltd) |
| 4161 | 84985001000027102 | Spironolactone | Spiroctan 25mg Tablet (Roche Products Ltd) |
| 4960 | 921811000001103 | Spironolactone | Aldactone 50mg tablets (Pfizer Ltd) |
| 6815 | 19654500100002710 1 | Spironolactone | Spironolactone 50mg/ 5ml oral suspension sugar free |
| 7952 | 421611000001100 | Spironolactone | Aldactone 100mg tablets (Pfizer Ltd) |
| 7991 | 85005001000027103 | Spironolactone | Spiroctan 100mg Capsule (Roche Products Ltd) |
| 10214 | 19636500100002710 9 | Spironolactone | Spironolactone 5mg/ 5ml oral suspension sugar free |
| 11156 | 40795001000027101 | Spironolactone | Spirolone 25mg Tablet (Berk Pharmaceuticals Ltd) |
| 11519 | 19638500100002710 7 | Spironolactone | Spironolactone 25mg/ 5ml oral suspension sugar free |
| 12946 | 19637500100002710 2 | Spironolactone | Spironolactone 10mg/ 5ml oral suspension sugar free |
| 13264 | 8727011000001103 | Spironolactone | Spironolactone 15mg/ 5ml oral suspension |
| 14109 | 19655500100002710 3 | Spironolactone | Spironolactone 100mg/ 5ml oral solution sugar free |
| 15052 | 84995001000027103 | Spironolactone | Spiroctan 50mg Tablet (Roche Products Ltd) |
| 17902 | 40815001000027104 | Spironolactone | Spirolone 100mg Tablet (Berk Pharmaceuticals Ltd) |
| 17950 | 40805001000027101 | Spironolactone | Spirolone 50mg Tablet (Berk Pharmaceuticals Ltd) |
| 19195 | 60755001000027100 | Spironolactone | Spironolactone 50mg Tablet (Wyeth Pharmaceuticals) |
| 21911 | 15505500100002710 2 | Spironolactone | Spirospare 25mg Tablet (Ashbourne Pharmaceuticals Ltd) |
| 23091 | 3411000001104 | Spironolactone | Spirospare 100 tablets (Ashbourne Pharmaceuticals Ltd) |
| 25494 | 75095001000027104 | Spironolactone | Diatensec 50mg Tablet (Pharmacia Ltd) |
| 29397 | 87695001000027105 | Spironolactone | Spiretic 100mg Tablet (DDSA Pharmaceuticals Ltd) |

| | | | |
|---|---|---|---|
| 31219 | 670811000001106 | Spironolactone | Spironolactone 100mg tablets (A A H Pharmaceuticals Ltd) |
| 31529 | 82311000001101 | Spironolactone | Spironolactone 25mg tablets (Teva UK Ltd) |
| 32837 | 330511000001102 | Spironolactone | Spironolactone 50mg tablets (Teva UK Ltd) |
| 34296 | 63111000001106 | Spironolactone | Spironolactone 25mg tablets (A A H Pharmaceuticals Ltd) |
| 34347 | 672411000001105 | Spironolactone | Spironolactone 25mg tablets (Actavis UK Ltd) |
| 34908 | 474611000001102 | Spironolactone | Spironolactone 25mg tablets (IVAX Pharmaceuticals UK Ltd) |
| 35789 | 20575001000027108 | Spironolactone | Spironolactone 25mg Tablet (Celltech Pharma Europe Ltd) |
| 41074 | 980361100001107 | Spironolactone | Spironolactone 25mg tablets (Almus Pharmaceuticals Ltd) |
| 41592 | 38611000001103 | Spironolactone | Spironolactone 100mg tablets (Actavis UK Ltd) |
| 41660 | 610611000001103 | Spironolactone | Spironolactone 100mg tablets (Teva UK Ltd) |
| 41706 | 121211000001101 | Spironolactone | Spironolactone 50mg tablets (IVAX Pharmaceuticals UK Ltd) |
| 43514 | 481811000001107 | Spironolactone | Spironolactone 50mg tablets (A A H Pharmaceuticals Ltd) |
| 45078 | 60285001000027101 | Spironolactone | Spironolactone 25mg/ 5ml Oral solution sugar free (Rosemont Pharmaceuticals Ltd) |
| 46674 | 60315001000027106 | Spironolactone | Spironolactone 50mg/ 5ml Oral suspension sugar free (Rosemont Pharmaceuticals Ltd) |
| 46990 | 8726411000001105 | Spironolactone | Spironolactone 50mg/ 5ml oral suspension |
| 47018 | 8726511000001109 | Spironolactone | Spironolactone 25mg/ 5ml oral suspension |
| 47687 | 87685001000027106 | Spironolactone | Spiretic 25mg Tablet (DDSA Pharmaceuticals Ltd) |
| 49388 | 8727611000001105 | Spironolactone | Spironolactone 100mg/ 5ml oral suspension |
| 50079 | 8727311000001100 | Spironolactone | Spironolactone 10mg/ 5ml oral suspension |
| 50370 | 8726311000001103 | Spironolactone | Spironolactone 5mg/ 5ml oral suspension |
| 51652 | 13583411000001106 | Spironolactone | Spironolactone 25mg tablets (DE Pharmaceuticals) |
| 51720 | 13894811000001108 | Spironolactone | Spironolactone 25mg/ 5ml oral solution |
| 51933 | 13894911000001103 | Spironolactone | Spironolactone 50mg/ 5ml oral solution |
| 52366 | 13895011000001103 | Spironolactone | Spironolactone 5mg/ 5ml oral solution |
| 52970 | 13894711000001100 | Spironolactone | Spironolactone 10mg/ 5ml oral solution |
| 53253 | 8705811000001105 | Spironolactone | Spironolactone 50mg/ 5ml oral suspension (Drug Tariff Special Order) |
| 54120 | 13374211000001104 | Spironolactone | Spironolactone 4mg/ 5ml oral suspension |
| 56067 | 13374111000001105 | Spironolactone | Spironolactone 4mg/ 5ml oral solution |
| 56274 | 13355011000001105 | Spironolactone | Spironolactone 4.5mg/ 5ml oral suspension |
| 56536 | 13894611000001109 | Spironolactone | Spironolactone 100mg/ 5ml oral solution |
| 57104 | 13326311000001102 | Spironolactone | Spironolactone 200mg/ 5ml oral suspension |
| 57556 | 13325611000001102 | Spironolactone | Spironolactone 12mg/ 5ml oral solution |
| 57933 | 13374011000001109 | Spironolactone | Spironolactone 40mg/ 5ml oral suspension |
| 58077 | 13385811000001105 | Spironolactone | Spironolactone 8mg/ 5ml oral suspension |
| 58225 | 13326111000001104 | Spironolactone | Spironolactone 2.5mg/ 5ml oral suspension |
| 58757 | 13326511000001108 | Spironolactone | Spironolactone 20mg/ 5ml oral suspension |
| 60343 | 235211000001102 | Spironolactone | Spironolactone 25mg tablets (Kent Pharmaceuticals Ltd) |
| 60660 | 13354811000001100 | Spironolactone | Spironolactone 3mg/ 5ml oral suspension |
| 61025 | 13326411000001109 | Spironolactone | Spironolactone 20mg/ 5ml oral solution |
| 63309 | 13384811000001109 | Spironolactone | Spironolactone 6mg/ 5ml oral suspension |
| 65582 | 13354711000001108 | Spironolactone | Spironolactone 3mg/ 5ml oral solution |
| 65822 | 13325311000001107 | Spironolactone | Spironolactone 12.5mg/ 5ml oral suspension |
| 67913 | 606411000001101 | Spironolactone | Spironolactone 50mg tablets (Kent Pharmaceuticals Ltd) |

| | | | |
|---|---|---|---|
| 69473 | 13354211000001101 | Spironolactone | Spironolactone 3.5mg/ 5ml oral suspension |
| 71010 | 20326811000001106 | Spironolactone | Spironolactone 25mg tablets (Genesis Pharmaceuticals Ltd) |
| 71398 | 13325211000001104 | Spironolactone | Spironolactone 12.5mg/ 5ml oral solution |
| 73208 | 13385411000001108 | Spironolactone | Spironolactone 7mg/ 5ml oral suspension |
| 73644 | 11010911000001104 | Spironolactone | Spironolactone 25mg tablets (Dr Reddy's Laboratories (UK) Ltd) |
| 74154 | 13353811000001103 | Spironolactone | Spironolactone 2mg/ 5ml oral suspension |
| 74285 | 8707011000001106 | Spironolactone | Spironolactone 25mg/ 5ml oral suspension (Drug Tariff Special Order) |
| 74631 | 46011000001105 | Spironolactone | Spironolactone 50mg tablets (Actavis UK Ltd) |
| 75413 | | Spironolactone | Spironolactone 8mg/ 5ml oral solution (Special Order) |
| 75488 | | Spironolactone | Spironolactone 250mg/ 5ml oral suspension |
| 75593 | | Spironolactone | Spironolactone 10mg/ 5ml Oral solution sugar free (Rosemont Pharmaceuticals Ltd) |
| 76195 | | Spironolactone | Spironolactone 5mg/ 5ml Oral suspension sugar free (Rosemont Pharmaceuticals Ltd) |
| 76321 | | Spironolactone | Spironolactone 100mg tablets (Genesis Pharmaceuticals Ltd) |
| 77604 | | Spironolactone | Spironolactone 7.5mg/ 5ml oral suspension |
| 77610 | | Spironolactone | Spironolactone 6.25mg/ 5ml oral suspension |
| 7582 | 3704811000001105 | Potassium chloride/ Furosemide | Lasikal modified-release tablets (Borg Medicare) |
| 7734 | 4540011000001102 | Potassium chloride/ Furosemide | Diumide-K Continus tablets (Teofarma) |
| 8102 | 4557711000001102 | Potassium chloride/ Furosemide | Furosemide 40mg / Potassium chloride 600mg (potassium 8mmol) modified-release tablets |
| 10781 | 37075001000027100 | Potassium chloride/ Furosemide | Lasix with k Tablet (Hoechst Marion Roussel) |
| 17960 | 36061311000001109 | Potassium chloride/ Furosemide | Furosemide 20mg / Potassium chloride 750mg (potassium 10mmol) modified-release tablets |
| 18983 | 3875001000027105 | Potassium Chloride/ Clopamide | Brinaldix k Effervescent tablet (Berk Pharmaceuticals Ltd) |
| 22839 | 209335001000027100 | Potassium Chloride/ Clopamide | Clopamide with Potassium effervescent tablets |
| 8891 | 14465001000027109 | Potassium Chloride/ Chlortalidone | Hygroton -k Tablet (Novartis Pharmaceuticals UK Ltd) |
| 1776 | 3290211000001109 | Potassium chloride/ Bumetanide | Burinex K modified-release tablets (LEO Pharma) |
| 6160 | 35913011000001100 | Potassium chloride/ Bumetanide | Bumetanide 500microgram / Potassium chloride 573mg (potassium 7.7mmol) modified-release tablets |
| 12360 | 20915001000027105 | Polythiazide | "Nephril 1mg Tablet (Pfizer Ltd)" |
| 12926 | 317967009 | Polythiazide | "Polythiazide 1mg tablets" |
| 7709 | 39735001000027105 | Piretanide | Arelix 6mg Capsule (Hoechst Marion Roussel) |
| 12367 | 135585001000027102 | Piretanide | Piretanide 6mg capsule |
| 4332 | 317965001 | Metolazone | Metolazone 5mg tablets |
| 4334 | 115735001000027107 | Metolazone | Metolazone 500microgram low dose Tablet |
| 8602 | 375611000001109 | Metolazone | Metenix 5mg tablets (Sanofi) |
| 19352 | 176195001000027101 | Metolazone | Xuret 0.5mg Tablet (Galen Ltd) |
| 49752 | 13005911000001103 | Metolazone | Metolazone 2.5mg/ 5ml oral solution |
| 53674 | 374173002 | Metolazone | Metolazone 2.5mg tablets |
| 54329 | 13006411000001102 | Metolazone | Metolazone 5mg/ 5ml oral suspension |
| 54643 | 251255001000027106 | Metolazone | Metolazone Oral solution |
| 55777 | 13006311000001109 | Metolazone | Metolazone 5mg/ 5ml oral solution |
| 61846 | 503445001000027107 | Metolazone | Zaroxolyn 2.5mg tablets (IDIS) |
| 68432 | 13006011000001106 | Metolazone | Metolazone 2.5mg/ 5ml oral suspension |

| | | | |
|---|---|---|---|
| 69009 | 21574611000001109 | Metolazone | Zaroxolyn 5mg tablets (Imported (Canada)) |
| 75260 | 21574911000001103 | Metolazone | Zaroxolyn 2.5mg tablets (Imported (Canada)) |
| 18267 | 124815001000027107 | Methyclothiazide | "Enduron 5mg Tablet (Abbott Laboratories Ltd)" |
| 20057 | 124785001000027106 | Methyclothiazide | "Methyclothiazide 5mg Tablet" |
| 31013 | 124005001000027109 | Mersalyl | Mersalyl 50mg/ ml Injection |
| 15457 | 2515001000027106 | Mefruside | Baycaron 25mg Tablet (Bayer Plc) |
| 17143 | 123565001000027105 | Mefruside | Mefruside 25mg Tablet |
| 2612 | 317956008 | Indapamide hemihydrate | Indapamide 2.5mg tablets |
| 7641 | 321811000001109 | Indapamide hemihydrate | Natrilix 2.5mg tablets (Servier Laboratories Ltd) |
| 26256 | 209045001000027100 | Indapamide hemihydrate | Opumide 2.5mg Tablet (Opus Pharmaceuticals Ltd) |
| 26275 | 424311000001108 | Indapamide hemihydrate | Nindaxa 2.5 tablets (Ashbourne Pharmaceuticals Ltd) |
| 27957 | 207505001000027101 | Indapamide hemihydrate | Natramid 2.5mg Tablet (Trinity Pharmaceuticals Ltd) |
| 33083 | 711711000001108 | Indapamide hemihydrate | Indapamide 2.5mg tablets (Teva UK Ltd) |
| 34551 | 450511000001107 | Indapamide hemihydrate | Indapamide 2.5mg tablets (Mylan) |
| 40907 | 8139411000001107 | Indapamide hemihydrate | Indapamide 2.5mg tablets (Genus Pharmaceuticals Ltd) |
| 42906 | 10437611000001100 | Indapamide hemihydrate | Indapamide 2.5mg tablets (Niche Generics Ltd) |
| 43516 | 397311000001100 | Indapamide hemihydrate | Indapamide 2.5mg tablets (Actavis UK Ltd) |
| 48079 | 214811000001107 | Indapamide hemihydrate | Indapamide 2.5mg tablets (Zentiva) |
| 48099 | 56211000001102 | Indapamide hemihydrate | Indapamide 2.5mg tablets (A A H Pharmaceuticals Ltd) |
| 49529 | 17932311000001102 | Indapamide hemihydrate | Indapamide 2.5mg tablets (Phoenix Healthcare Distribution Ltd) |
| 54316 | 598411000001109 | Indapamide hemihydrate | Indapamide 2.5mg tablets (Alliance Healthcare (Distribution) Ltd) |
| 55259 | 146711000001100 | Indapamide hemihydrate | Indapamide 2.5mg tablets (Kent Pharmaceuticals Ltd) |
| 56296 | 14784911000001103 | Indapamide hemihydrate | Indapamide 2.5mg tablets (Boston Healthcare Ltd) |
| 56760 | 18283111000001109 | Indapamide hemihydrate | Indapamide 2.5mg tablets (Strides Shasun (UK) Ltd) |
| 70509 | 13579511000001104 | Indapamide hemihydrate | Indapamide 2.5mg tablets (DE Pharmaceuticals) |
| 77406 | | Indapamide hemihydrate | Natrilix 2.5mg tablets (Mawdsley-Brooks & Company Ltd) |
| 3056 | 456611000001108 | Indapamide | Natrilix SR 1.5mg tablets (Servier Laboratories Ltd) |
| 5112 | 317954006 | Indapamide | Indapamide 1.5mg modified-release tablets |
| 39447 | 14693611000001102 | Indapamide | Varbim XL 1.5mg tablets (Teva UK Ltd) |
| 41861 | 14242211000001100 | Indapamide | Tensaid XL 1.5mg tablets (Mylan) |
| 41885 | 13824811000001102 | Indapamide | Ethibide XL 1.5mg tablets (Genus Pharmaceuticals Ltd) |
| 43184 | 15600711000001107 | Indapamide | Mapemid XL 1.5mg tablets (Teva UK Ltd) |
| 44168 | 15436111000001104 | Indapamide | Indipam XL 1.5mg tablets (Actavis UK Ltd) |
| 46675 | 14033311000001105 | Indapamide | Indapamide 1.5mg modified-release tablets (A A H Pharmaceuticals Ltd) |
| 59616 | 16737911000001105 | Indapamide | Rawel XL 1.5mg tablets (Consilient Health Ltd) |
| 60020 | 22058211000001103 | Indapamide | Indapamide 1.5mg modified-release tablets (Waymade Healthcare Plc) |
| 62066 | 24331611000001104 | Indapamide | Cardide SR 1.5mg tablets (Teva UK Ltd) |
| 62771 | 24175711000001103 | Indapamide | Indapamide 1.5mg modified-release tablets (DE Pharmaceuticals) |
| 64066 | 21578911000001105 | Indapamide | Indapamide 2.5mg/ 5ml oral suspension |
| 74172 | 15101611000001109 | Indapamide | Indapamide 1.5mg modified-release tablets (Sigma Pharmaceuticals Plc) |
| 77449 | | Indapamide | Natrilix SR 1.5mg tablets (Waymade Healthcare Plc) |
| 12110 | 116195001000027108 | Hydroflumethiazide | "Hydroflumethiazide 50mg Tablet" |
| 13525 | 80235001000027106 | Hydroflumethiazide | "Hydrenox 50mg Tablet (Knoll Ltd)" |

| | | | |
|---|---|---|---|
| 1721 | 132811000001106 | Hydrochlorothiazide/ Triamterene | Dyazide 50mg/ 25mg tablets (AMCo) |
| 5416 | 410896007 | Hydrochlorothiazide/ Triamterene | Co-triamterzide 50mg/ 25mg tablets |
| 8897 | 191211000001106 | Hydrochlorothiazide/ Triamterene | Triam-Co 50mg/ 25mg tablets (IVAX Pharmaceuticals UK Ltd) |
| 18726 | 721311000001104 | Hydrochlorothiazide/ Triamterene | Triamaxco 50mg/ 25mg tablets (Ashbourne Pharmaceuticals Ltd) |
| 47804 | 12779411000001108 | Hydrochlorothiazide/ Triamterene | Co-triamterzide 50mg/ 25mg tablets (A A H Pharmaceuticals Ltd) |
| 67801 | 16160111000001109 | Hydrochlorothiazide/ Triamterene | Dyazide 50mg/ 25mg tablets (Lexon (UK) Ltd) |
| 77386 | | Hydrochlorothiazide/ Triamterene | Dyazide 50mg/ 25mg tablets (Dowelhurst Ltd) |
| 924 | 318121006 | Hydrochlorothiazide/ Amiloride | Co-amilozide 2.5mg/ 25mg tablets |
| 1251 | 314211000001100 | Hydrochlorothiazide/ Amiloride | Moduret 25 tablets (Merck Sharp & Dohme Ltd) |
| 34367 | 925611000001104 | Hydrochlorothiazide/ Amiloride | Co-amilozide 2.5mg/ 25mg tablets (Wockhardt UK Ltd) |
| 60354 | 495611000001107 | Hydrochlorothiazide/ Amiloride | Co-amilozide 2.5mg/ 25mg tablets (Kent Pharmaceuticals Ltd) |
| 542 | 376209006 | Hydrochlorothiazide | "Hydrochlorothiazide 25mg tablets" |
| 3517 | 376508004 | Hydrochlorothiazide | "Hydrochlorothiazide 50mg tablets" |
| 12440 | 4544011000001109 | Hydrochlorothiazide | "Hydrosaluric 25mg tablets (Merck Sharp & Dohme Ltd)" |
| 13363 | 10555001000027106 | Hydrochlorothiazide | "Esidrex 50mg Tablet (Novartis Pharmaceuticals UK Ltd)" |
| 16632 | 4546211000001109 | Hydrochlorothiazide | "Hydrosaluric 50mg tablets (Merck Sharp & Dohme Ltd)" |
| 17252 | 10545001000027108 | Hydrochlorothiazide | "Esidrex 25mg Tablet (Novartis Pharmaceuticals UK Ltd)" |
| 48132 | 250545001000027106 | Hydrochlorothiazide | "Hydrochlorothiazide Capsule" |
| 57488 | 246525001000027108 | Hydrochlorothiazide | "Hydrochlorothiazide Oral solution" |
| 62516 | 24594211000001108 | Hydrochlorothiazide | "Hydrochlorothiazide 12.5mg tablets" |
| 1369 | 107935001000027101 | Furosemide/ Amiloride | Furosemide with amiloride 40mg+5mg Tablet |
| 4211 | 107945001000027100 | Furosemide/ Amiloride | Furosemide with amiloride 20mg+2.5mg Tablet |
| 5220 | 107955001000027102 | Furosemide/ Amiloride | Furosemide with amiloride 80mg+10mg Tablet |
| 9456 | 159445001000027103 | Furosemide/ Amiloride | Amiloride 5mg / furosemide 40mg tablets |
| 15874 | 159435001000027104 | Furosemide/ Amiloride | Amiloride 2.5mg / furosemide 20mg tablets |
| 18497 | 159455001000027100 | Furosemide/ Amiloride | Amiloride 10mg / furosemide 80mg tablets |
| 47647 | 245685001000027108 | Furosemide/ Amiloride | Co-amilofruse oral liquid |
| 6 | 317972000 | Furosemide | Furosemide 40mg tablets |
| 55 | 317971007 | Furosemide | Furosemide 20mg tablets |
| 562 | 107885001000027101 | Furosemide | Furosemide 10mg/ ml Injection |
| 3248 | 317973005 | Furosemide | Furosemide 500mg tablets |
| 3287 | 199505001000027101 | Furosemide | Furosemide 1mg/ ml Oral solution |
| 4182 | 701111000001105 | Furosemide | Lasix 5mg/ 5ml oral solution (Borg Medicare) |
| 4258 | 9611000001104 | Furosemide | Lasix 20mg/ 2ml solution for injection ampoules (Sanofi) |
| 4705 | 145335001000027109 | Furosemide | Furosemide 20mg/ 2ml Injection |
| 5249 | 36564411000001102 | Furosemide | Furosemide 50mg/ 5ml oral solution sugar free |
| 5728 | 318007007 | Furosemide | Furosemide 40mg/ 5ml oral solution sugar free |
| 5868 | 602811000001100 | Furosemide | Frusol 20mg/ 5ml oral solution (Rosemont Pharmaceuticals Ltd) |
| 6118 | 318006003 | Furosemide | Furosemide 20mg/ 5ml oral solution sugar free |
| 7606 | 79411000001107 | Furosemide | Lasix 40mg tablets (Sanofi) |
| 7799 | 701611000001102 | Furosemide | Lasix 20mg tablets (Borg Medicare) |
| 9680 | 494811000001105 | Furosemide | Frusol 40mg/ 5ml oral solution (Rosemont Pharmaceuticals Ltd) |
| 10392 | 829911000001102 | Furosemide | Lasix 500mg tablets (Sanofi) |

| | | | |
|---|---|---|---|
| 10422 | 37095001000027109 | Furosemide | Lasix 50mg/ 5ml Injection (Hoechst UK Ltd) |
| 12318 | 153285001000027109 | Furosemide | Lasix 250mg/ 25ml Injection (Hoechst Marion Roussel) |
| 14761 | 99311000001108 | Furosemide | Frusid 40mg tablets (Dr Reddy's Laboratories (UK) Ltd) |
| 14837 | 855611000001109 | Furosemide | Frusol 50mg/ 5ml oral solution (Rosemont Pharmaceuticals Ltd) |
| 16206 | 714311000001107 | Furosemide | Froop 40mg tablets (Ashbourne Pharmaceuticals Ltd) |
| 18716 | 40555001000027108 | Furosemide | Dryptal 10mg/ ml Injection (Berk Pharmaceuticals Ltd) |
| 19056 | 60235001000027109 | Furosemide | Furosemide 50mg/ 5ml sugar free Oral solution (Rosemont Pharmaceuticals Ltd) |
| 19192 | 118425001000027108 | Furosemide | Furosemide 40mg Tablet (M & A Pharmachem Ltd) |
| 19194 | 17211000001105 | Furosemide | Furosemide 20mg tablets (Teva UK Ltd) |
| 19258 | 36061511000001103 | Furosemide | Furosemide 50mg/ 5ml solution for injection ampoules |
| 20538 | 1795001000027107 | Furosemide | Frumax 40mg Tablet (Ashbourne Pharmaceuticals Ltd) |
| 21849 | 8805001000027103 | Furosemide | Dryptal 40mg Tablet (Berk Pharmaceuticals Ltd) |
| 24835 | 23365001000027103 | Furosemide | Min-i-jet furosemide 10mg/ ml Injection (Celltech Pharma Europe Ltd) |
| 25334 | 119611000001105 | Furosemide | Furosemide 500mg tablets (A A H Pharmaceuticals Ltd) |
| 25717 | 147711000001102 | Furosemide | Furosemide 40mg tablets (Mylan) |
| 26292 | 82155001000027100 | Furosemide | Diuresal 40mg Tablet (Lagap) |
| 27447 | 445111000001108 | Furosemide | Furosemide 40mg tablets (Wockhardt UK Ltd) |
| 27690 | 100611000001101 | Furosemide | Furosemide 40mg tablets (A A H Pharmaceuticals Ltd) |
| 27696 | 765411000001103 | Furosemide | Furosemide 40mg tablets (Kent Pharmaceuticals Ltd) |
| 27926 | 229711000001109 | Furosemide | Furosemide 20mg tablets (Mylan) |
| 29780 | 13295001000027107 | Furosemide | Furosemide 20mg Tablet (C P Pharmaceuticals Ltd) |
| 30625 | 202711000001106 | Furosemide | Furosemide 20mg tablets (A A H Pharmaceuticals Ltd) |
| 30875 | 34193711000001108 | Furosemide | Furosemide 250mg/ 25ml solution for injection ampoules |
| 31548 | 631611000001100 | Furosemide | Furosemide 20mg tablets (Actavis UK Ltd) |
| 32277 | 36061611000001104 | Furosemide | Furosemide 80mg/ 8ml solution for injection pre-filled syringes |
| 32896 | 8751711000001106 | Furosemide | Furosemide 40mg tablets (Ranbaxy (UK) Ltd) |
| 32918 | 662811000001104 | Furosemide | Furosemide 20mg tablets (Sandoz Ltd) |
| 34006 | 351511000001105 | Furosemide | Furosemide 40mg tablets (Actavis UK Ltd) |
| 34374 | 472211000001108 | Furosemide | Furosemide 40mg tablets (Teva UK Ltd) |
| 34557 | 856211000001101 | Furosemide | Furosemide 40mg tablets (IVAX Pharmaceuticals UK Ltd) |
| 35162 | 36061411000001102 | Furosemide | Furosemide 20mg/ 2ml solution for injection ampoules |
| 36190 | 318010000 | Furosemide | Furosemide 5mg/ 5ml oral solution sugar free |
| 40247 | 46575001000027100 | Furosemide | Furosemide 10mg/ ml Injection (Martindale Pharmaceuticals Ltd) |
| 41292 | 539911000001101 | Furosemide | Furosemide 20mg tablets (Wockhardt UK Ltd) |
| 41405 | 752711000001106 | Furosemide | Furosemide 500mg tablets (Teva UK Ltd) |
| 41828 | 822211000001103 | Furosemide | Furosemide 500mg tablets (Actavis UK Ltd) |
| 42388 | 10305811000001105 | Furosemide | Furosemide 40mg/ 5ml oral solution sugar free (Focus Pharmaceuticals Ltd) |
| 42488 | 10214011000001105 | Furosemide | Furosemide 40mg/ 5ml oral solution sugar free (A A H Pharmaceuticals Ltd) |
| 46116 | 58505001000027108 | Furosemide | Furosemide 10mg/ ml Injection (Antigen Pharmaceuticals) |
| 46699 | 9801311000001103 | Furosemide | Furosemide 40mg tablets (Almus Pharmaceuticals Ltd) |
| 46948 | 10429011000001100 | Furosemide | Furosemide 40mg tablets (Arrow Generics Ltd) |
| 47815 | 13335001000027109 | Furosemide | Furosemide 20mg Tablet (Celltech Pharma Europe Ltd) |
| 49268 | 12088211000001109 | Furosemide | Furosemide 50mg/ 5ml oral suspension |

| | | | |
|---|---|---|---|
| 51983 | 13893511000001107 | Furosemide | Furosemide 5mg/ 5ml oral suspension |
| 52045 | 20176911000001106 | Furosemide | Furosemide 250mg/ 5ml solution for injection vials |
| 52887 | 725611000001107 | Furosemide | Furosemide 20mg/ 2ml solution for injection ampoules (A A H Pharmaceuticals Ltd) |
| 52900 | 2898811000001106 | Furosemide | Furosemide 80mg/ 8ml solution for injection Minijet pre-filled syringes (UCB Pharma Ltd) |
| 53967 | 16052611000001100 | Furosemide | Furosemide 20mg tablets (Bristol Laboratories Ltd) |
| 54825 | 15089311000001105 | Furosemide | Furosemide 20mg tablets (Sigma Pharmaceuticals Plc) |
| 55738 | 5088711000001101 | Furosemide | Furosemide 50mg/ 5ml solution for injection ampoules (Hameln Pharmaceuticals Ltd) |
| 56051 | 503711000001106 | Furosemide | Furosemide 20mg tablets (Kent Pharmaceuticals Ltd) |
| 56375 | 18461911000001109 | Furosemide | Furosemide 40mg tablets (Accord Healthcare Ltd) |
| 57600 | 120011000001102 | Furosemide | Furosemide 20mg/ 2ml solution for injection ampoules (Alliance Healthcare (Distribution) Ltd) |
| 57610 | 10305611000001106 | Furosemide | Furosemide 20mg/ 5ml oral solution sugar free (Focus Pharmaceuticals Ltd) |
| 58078 | 12088911000001100 | Furosemide | Furosemide 8mg/ 5ml oral solution |
| 58224 | 12035011000001100 | Furosemide | Furosemide 10mg/ 5ml oral solution |
| 59030 | 12036211000001104 | Furosemide | Furosemide 20mg/ 5ml oral solution |
| 59290 | 121411000001102 | Furosemide | Furosemide 20mg tablets (Alliance Healthcare (Distribution) Ltd) |
| 59884 | 17911411000001106 | Furosemide | Furosemide 20mg tablets (Phoenix Healthcare Distribution Ltd) |
| 59911 | 139011000001107 | Furosemide | Furosemide 40mg tablets (Alliance Healthcare (Distribution) Ltd) |
| 59939 | 12036311000001107 | Furosemide | Furosemide 20mg/ 5ml oral suspension |
| 60291 | 19309711000001101 | Furosemide | Furosemide 40mg tablets (AMCo) |
| 60465 | 13893411000001108 | Furosemide | Furosemide 5mg/ 5ml oral solution |
| 61365 | 12087811000001106 | Furosemide | Furosemide 40mg/ 5ml oral suspension |
| 61475 | 23909911000001105 | Furosemide | Furosemide 20mg tablets (DE Pharmaceuticals) |
| 63237 | 14789111000001100 | Furosemide | Furosemide 20mg tablets (Boston Healthcare Ltd) |
| 64255 | 12036811000001103 | Furosemide | Furosemide 2mg/ 5ml oral solution |
| 64677 | 23910211000001105 | Furosemide | Furosemide 40mg tablets (DE Pharmaceuticals) |
| 64745 | 12089011000001109 | Furosemide | Furosemide 8mg/ 5ml oral suspension |
| 65583 | 12087311000001102 | Furosemide | Furosemide 3mg/ 5ml oral solution |
| 66017 | 9800811000001104 | Furosemide | Furosemide 20mg tablets (Almus Pharmaceuticals Ltd) |
| 66149 | 12087511000001108 | Furosemide | Furosemide 4.5mg/ 5ml oral solution |
| 67910 | 12087711000001103 | Furosemide | Furosemide 40mg/ 5ml oral solution |
| 68068 | 8519911000001102 | Furosemide | Furosemide 4mg/ 5ml oral solution |
| 69338 | 60225001000027107 | Furosemide | Furosemide 40mg/ 5ml sugar free Oral solution (Rosemont Pharmaceuticals Ltd) |
| 69445 | 18706911000001106 | Furosemide | Furosemide 40mg/ 5ml oral solution sugar free (Sigma Pharmaceuticals Plc) |
| 70650 | 34537111000001101 | Furosemide | Furosemide 50mg/ 5ml solution for injection ampoules (Peckforton Pharmaceuticals Ltd) |
| 71406 | 12035811000001106 | Furosemide | Furosemide 1mg/ 5ml oral solution |
| 71950 | 22018711000001104 | Furosemide | Furosemide 20mg tablets (Waymade Healthcare Plc) |
| 73171 | 246455001000027101 | Furosemide | Furosemide Oral solution |
| 73480 | 60215001000027103 | Furosemide | Furosemide 20mg/ 5ml sugar free Oral solution (Rosemont Pharmaceuticals Ltd) |
| 74381 | 12087411000001109 | Furosemide | Furosemide 3mg/ 5ml oral suspension |
| 74677 | 30133911000001101 | Furosemide | Furosemide 40mg tablets (Mawdsley-Brooks & Company Ltd) |
| 74926 | 28986611000001105 | Furosemide | Furosemide 20mg tablets (Crescent Pharma Ltd) |
| 75855 | | Furosemide | Furosemide 10mg/ 5ml oral suspension |

| | | | |
|---|---|---|---|
| 76157 | | Furosemide | Furosemide 50mg/ 5ml oral solution |
| 76323 | | Furosemide | Furosemide 40mg tablets (Crescent Pharma Ltd) |
| 76934 | | Furosemide | Furosemide 6.5mg/ 5ml oral solution |
| 77333 | | Furosemide | Aluzine 20mg Tablet (M A Steinhard Ltd) |
| 77566 | | Furosemide | Furosemide 40mg tablets (Bristol Laboratories Ltd) |
| 12354 | 99155001000027108 | Etacrynic Acid | Etacrynic 50mg tablets |
| 18650 | 9585001000027101 | Etacrynic Acid | Edecrin 50mg Tablet (Merck Sharp & Dohme Ltd) |
| 32002 | 179865001000027103 | Etacrynic Acid | Etacrynic 50mg/ vial injection |
| 1170 | 317940003 | Cyclopenthiazide | "Cyclopenthiazide 500microgram tablets" |
| 2046 | 487811000001100 | Cyclopenthiazide | "Navidrex 500microgram tablets (AMCo)" |
| 12546 | 40975001000027100 | Chlortalidone/ Triamterene | Kalspare Tablet (Dominion Pharma) |
| 12547 | 318100006 | Chlortalidone/ Triamterene | Triamterene 50mg / Chlortalidone 50mg tablets |
| 16498 | 3252011000001105 | Chlortalidone/ Triamterene | Kalspare tablets (DHP Healthcare Ltd) |
| 605 | 317935006 | Chlortalidone | Chlortalidone 50mg tablets |
| 3054 | 14435001000027104 | Chlortalidone | Hygroton 100mg Tablet (Alliance Pharmaceuticals Ltd) |
| 3548 | 101645001000027102 | Chlortalidone | Chlortalidone 100mg tablets |
| 3997 | 285911000001101 | Chlortalidone | Hygroton 50mg tablets (Alliance Pharmaceuticals Ltd) |
| 74942 | 8359911000001106 | Chlortalidone | Chlortalidone 50mg/ 5ml oral suspension |
| 75383 | 376346007 | Chlortalidone | Chlortalidone 25mg tablets |
| 6816 | 408039005 | Chlorothiazide | "Chlorothiazide 250mg/5ml oral suspension" |
| 8836 | 101315001000027103 | Chlorothiazide | "Chlorothiazide 500mg tablets" |
| 13246 | 8358511000001105 | Chlorothiazide | "Chlorothiazide 150mg/5ml oral suspension" |
| 17720 | 27705001000027105 | Chlorothiazide | "Saluric 500mg Tablet (Merck Sharp & Dohme Ltd)" |
| 33724 | 7652811000001107 | Chlorothiazide | "Diuril 250mg/5ml oral suspension (Imported (United States))" |
| 54341 | 12015711000001107 | Chlorothiazide | "Chlorothiazide 5mg/5ml oral suspension" |
| 54679 | 395516007 | Chlorothiazide | "Chlorothiazide 250mg tablets" |
| 55889 | 243885001000027106 | Chlorothiazide | "Chlorothiazide oral solution" |
| 56804 | 8358711000001100 | Chlorothiazide | "Chlorothiazide 25mg/5ml oral suspension" |
| 59834 | 12503011000001109 | Chlorothiazide | "Chlorothiazide 250mg/5ml oral solution" |
| 60603 | 8359111000001108 | Chlorothiazide | "Chlorothiazide 50mg/5ml oral suspension" |
| 63227 | 12016611000001108 | Chlorothiazide | "Chlorothiazide 70mg/5ml oral solution" |
| 64798 | 12431311000001107 | Chlorothiazide | "Chlorothiazide 120mg/5ml oral solution" |
| 71871 | 8359211000001102 | Chlorothiazide | "Chlorothiazide 60mg/5ml oral suspension" |
| 73441 | 12502211000001105 | Chlorothiazide | "Chlorothiazide 200mg/5ml oral solution" |
| 74153 | 12431011000001109 | Chlorothiazide | "Chlorothiazide 10mg/5ml oral solution" |
| 75957 | | Chlorothiazide | "Chlorothiazide 5mg/5ml oral solution" |
| 76031 | | Chlorothiazide | "Chlorothiazide 250mg/5ml oral solution (Special Order)" |
| 77162 | | Chlorothiazide | "Chlorothiazide 12.5mg/5ml oral solution" |
| 77850 | | Chlorothiazide | "Chlorothiazide 500mg/5ml oral solution" |
| 2493 | 33911000001104 | Bumetanide/ Amiloride | Burinex A 5mg/ 1mg tablets (LEO Pharma) |
| 2495 | 189885001000027104 | Bumetanide/ Amiloride | Bumetanide with Amiloride tablets |
| 14587 | 318097001 | Bumetanide/ Amiloride | Amiloride 5mg / Bumetanide 1mg tablets |
| 814 | 318021009 | Bumetanide | Bumetanide 1mg tablets |

| | | | |
|---|---|---|---|
| 2788 | 846411000001100 | Bumetanide | Burinex 1mg tablets (LEO Pharma) |
| 5218 | 318023007 | Bumetanide | Bumetanide 1mg/ 5ml oral solution sugar free |
| 7806 | 318022002 | Bumetanide | Bumetanide 5mg tablets |
| 12226 | 3787500100027101 | Bumetanide | Burinex 1mg/ 5ml Oral solution (LEO Pharma) |
| 12294 | 521811000001107 | Bumetanide | Burinex 5mg tablets (LEO Pharma) |
| 15341 | 3512500100027107 | Bumetanide | Burinex 0.5mg/ ml Injection (LEO Pharma) |
| 19300 | 3591291100001108 | Bumetanide | Bumetanide 2mg/ 4ml solution for injection ampoules |
| 30913 | 232615001000027108 | Bumetanide | Betinex 1mg Tablet (Berk Pharmaceuticals Ltd) |
| 31932 | 689011000001100 | Bumetanide | Bumetanide 1mg tablets (C P Pharmaceuticals Ltd) |
| 32091 | 43411000001106 | Bumetanide | Bumetanide 1mg tablets (A A H Pharmaceuticals Ltd) |
| 34613 | 102011000001107 | Bumetanide | Bumetanide 5mg tablets (Teva UK Ltd) |
| 34934 | 402111000001109 | Bumetanide | Bumetanide 1mg tablets (Mylan) |
| 36767 | 226011000001101 | Bumetanide | Bumetanide 1mg tablets (IVAX Pharmaceuticals UK Ltd) |
| 39602 | 539611000001107 | Bumetanide | Bumetanide 1mg tablets (Actavis UK Ltd) |
| 45305 | 274211000001100 | Bumetanide | Bumetanide 1mg tablets (Teva UK Ltd) |
| 55548 | 731811000001106 | Bumetanide | Bumetanide 1mg tablets (Alliance Healthcare (Distribution) Ltd) |
| 62024 | 376211000001101 | Bumetanide | Bumetanide 5mg tablets (A A H Pharmaceuticals Ltd) |
| 63555 | 17810611000001109 | Bumetanide | Bumetanide 1mg tablets (Phoenix Healthcare Distribution Ltd) |
| 66195 | 9793511000001108 | Bumetanide | Bumetanide 1mg tablets (Almus Pharmaceuticals Ltd) |
| 73152 | 21804811000001102 | Bumetanide | Bumetanide 1mg tablets (Waymade Healthcare Plc) |
| 73195 | 22622811000001107 | Bumetanide | Bumetanide 1mg tablets (DE Pharmaceuticals) |
| 74814 | 14106511000001105 | Bumetanide | Bumetanide 1mg tablets (Niche Generics Ltd) |
| 30272 | 122075001000027106 | Benzthiazide/ Triamterene | Benthiazide with Triamterene capsules |
| 2 | 317919004 | Bendroflumethiazide | "Bendroflumethiazide 2.5mg tablets" |
| 58 | 317920005 | Bendroflumethiazide | "Bendroflumethiazide 5mg tablets" |
| 1209 | 817911000001100 | Bendroflumethiazide | "Neo-Naclex 5mg tablets (Mercury Pharma Group Ltd)" |
| 7351 | 8306811000001101 | Bendroflumethiazide | "Bendroflumethiazide 2.5mg/5ml oral suspension" |
| 7698 | 120911000001103 | Bendroflumethiazide | "Aprinox 5mg tablets (Amdipharm Plc)" |
| 8526 | 672111000001100 | Bendroflumethiazide | "Aprinox 2.5mg tablets (AMCo)" |
| 18973 | 3507500100027101 | Bendroflumethiazide | "Centyl 2.5mg Tablet (Edwin Burgess Ltd)" |
| 21803 | 321500100027107 | Bendroflumethiazide | "Berkozide 2.5mg Tablet (Berk Pharmaceuticals Ltd)" |
| 21867 | 322500100027103 | Bendroflumethiazide | "Berkozide 5mg Tablet (Berk Pharmaceuticals Ltd)" |
| 23427 | 501911000001107 | Bendroflumethiazide | "Bendroflumethiazide 5mg tablets (A A H Pharmaceuticals Ltd)" |
| 24189 | 197885001000027105 | Bendroflumethiazide | "Neo-bendromax 2.5mg Tablet (Ashbourne Pharmaceuticals Ltd)" |
| 24190 | 197895001000027109 | Bendroflumethiazide | "Neo-bendromax 5mg Tablet (Ashbourne Pharmaceuticals Ltd)" |
| 27256 | 454811000001100 | Bendroflumethiazide | "Bendroflumethiazide 2.5mg tablets (Wockhardt UK Ltd)" |
| 27689 | 464311000001103 | Bendroflumethiazide | "Bendroflumethiazide 2.5mg tablets (IVAX Pharmaceuticals UK Ltd)" |
| 29991 | 3508500100027106 | Bendroflumethiazide | "Centyl 5mg Tablet (Edwin Burgess Ltd)" |
| 31670 | 779811000001105 | Bendroflumethiazide | "Bendroflumethiazide 2.5mg tablets (Teva UK Ltd)" |
| 31820 | 159711000001108 | Bendroflumethiazide | "Bendroflumethiazide 5mg tablets (Wockhardt UK Ltd)" |
| 33415 | 914811000001108 | Bendroflumethiazide | "Bendroflumethiazide 2.5mg tablets (Mylan)" |
| 33651 | 772511000001103 | Bendroflumethiazide | "Bendroflumethiazide 2.5mg tablets (A A H Pharmaceuticals Ltd)" |
| 34059 | 1711000001109 | Bendroflumethiazide | "Bendroflumethiazide 2.5mg tablets (Actavis UK Ltd)" |

| | | | |
|---|---|---|---|
| 34124 | 720911000001105 | Bendroflumethiazide | "Bendroflumethiazide 5mg tablets (Actavis UK Ltd)" |
| 34602 | 17195711000001104 | Bendroflumethiazide | "Bendroflumethiazide 2.5mg tablets (Sovereign Medical Ltd)" |
| 34803 | 113045001000027101 | Bendroflumethiazide | "Bendroflumethiazide 2.5mg Tablet (Regent Laboratories Ltd)" |
| 40149 | 320511000001104 | Bendroflumethiazide | "Bendroflumethiazide 5mg tablets (IVAX Pharmaceuticals UK Ltd)" |
| 40886 | 979281100001103 | Bendroflumethiazide | "Bendroflumethiazide 2.5mg tablets (Almus Pharmaceuticals Ltd)" |
| 41517 | 436711000001100 | Bendroflumethiazide | "Bendroflumethiazide 5mg tablets (Teva UK Ltd)" |
| 46302 | 18149011000001101 | Bendroflumethiazide | "Neo-Naclex 2.5mg tablets (AMCo)" |
| 47844 | 750511000001102 | Bendroflumethiazide | "Bendroflumethiazide 2.5mg tablets (Kent Pharmaceuticals Ltd)" |
| 53812 | 244535001000027101 | Bendroflumethiazide | "Bendroflumethiazide oral solution" |
| 64907 | 979311100001104 | Bendroflumethiazide | "Bendroflumethiazide 5mg tablets (Almus Pharmaceuticals Ltd)" |
| 66517 | 830701100001105 | Bendroflumethiazide | "Bendroflumethiazide 1.25mg/5ml oral suspension" |
| 67737 | 11009711000001106 | Bendroflumethiazide | "Bendroflumethiazide 2.5mg tablets (Dr Reddy's Laboratories (UK) Ltd)" |
| 67738 | 11010111000001102 | Bendroflumethiazide | "Bendroflumethiazide 5mg tablets (Dr Reddy's Laboratories (UK) Ltd)" |
| 67780 | 22617011000001105 | Bendroflumethiazide | "Bendroflumethiazide 5mg tablets (DE Pharmaceuticals)" |
| 70989 | 20323211000001103 | Bendroflumethiazide | "Bendroflumethiazide 2.5mg tablets (Genesis Pharmaceuticals Ltd)" |
| 72042 | 288611000001101 | Bendroflumethiazide | "Bendroflumethiazide 2.5mg tablets (Alliance Healthcare (Distribution) Ltd)" |
| 72083 | 21782811000001109 | Bendroflumethiazide | "Bendroflumethiazide 5mg tablets (Waymade Healthcare Plc)" |
| 72914 | 6685001000027101 | Bendroflumethiazide | "Bendroflumethiazide 2.5mg Tablet (Celltech Pharma Europe Ltd)" |
| 77681 | | Bendroflumethiazide | "Bendroflumethiazide 5mg tablets (Mylan)" |
| 348 | 19555001000027107 | Amiloride / Hydrochlorothiazide | Moduretic Tablet (Bristol-Myers Squibb Pharmaceuticals Ltd) |
| 923 | 377566005 | Amiloride / Hydrochlorothiazide | Co-amilozide 5mg/ 50mg tablets |
| 2002 | 159475001000027102 | Amiloride / Hydrochlorothiazide | Amiloride 5mg / hydrochlorothiazide 50mg tablets |
| 3293 | 19565001000027103 | Amiloride / Hydrochlorothiazide | Moduretic Oral solution (Bristol-Myers Squibb Pharmaceuticals Ltd) |
| 3701 | 159485001000027107 | Amiloride / Hydrochlorothiazide | Amiloride 2.5mg / hydrochlorothiazide 25mg tablets |
| 4034 | 159495001000027106 | Amiloride / Hydrochlorothiazide | Amiloride 5mg / hydrochlorothiazide 50mg/ 5ml solution |
| 8058 | 116145001000027100 | Amiloride / Hydrochlorothiazide | Normetic Tablet (Abbott Laboratories Ltd) |
| 18361 | 712211000001108 | Amiloride / Hydrochlorothiazide | Amilmaxco 5mg/ 50mg tablets (Ashbourne Pharmaceuticals Ltd) |
| 18733 | 169595001000027101 | Amiloride / Hydrochlorothiazide | Co-amilozide 5mg with 50mg/ ml oral solution |
| 19890 | 116045001000027108 | Amiloride / Hydrochlorothiazide | Hydrochlorothiazide with amiloride 25mgwith2.5mg Tablet |
| 20066 | 636611000001107 | Amiloride / Hydrochlorothiazide | Amil-Co 5mg/ 50mg tablets (IVAX Pharmaceuticals UK Ltd) |
| 22923 | 116065001000027102 | Amiloride / Hydrochlorothiazide | Hydrochlorothiazide with amiloride 50mg with 5mg Tablet |
| 24008 | 181425001000027109 | Amiloride / Hydrochlorothiazide | Vasetic Tablet (Shire Pharmaceuticals Ltd) |
| 25500 | 152595001000027107 | Amiloride / Hydrochlorothiazide | Hypertane 50 Tablet (Schwarz Pharma Ltd) |
| 26219 | 211855001000027109 | Amiloride / Hydrochlorothiazide | Zida-co 5mg+50mg Tablet (Opus Pharmaceuticals Ltd) |
| 26220 | 151865001000027108 | Amiloride / Hydrochlorothiazide | Delvas Tablet (Berk Pharmaceuticals Ltd) |
| 31150 | 831011000001103 | Amiloride / Hydrochlorothiazide | Co-amilozide 5mg/ 50mg tablets (IVAX Pharmaceuticals UK Ltd) |
| 41556 | 26611000001101 | Amiloride / Hydrochlorothiazide | Co-amilozide 5mg/ 50mg tablets (Teva UK Ltd) |
| 42142 | 453811000001103 | Amiloride / Hydrochlorothiazide | Moduretic 5mg/ 50mg tablets (Merck Sharp & Dohme Ltd) |
| 46916 | 17611000001107 | Amiloride / Hydrochlorothiazide | Co-amilozide 5mg/ 50mg tablets (A A H Pharmaceuticals Ltd) |
| 62249 | 552711000001105 | Amiloride / Hydrochlorothiazide | Co-amilozide 5mg/ 50mg tablets (Alliance Healthcare (Distribution) Ltd) |
| 62700 | 17885211000001100 | Amiloride / Hydrochlorothiazide | Co-amilozide 5mg/ 50mg tablets (Phoenix Healthcare Distribution Ltd) |
| 73337 | 366811000001105 | Amiloride / Hydrochlorothiazide | Co-amilozide 5mg/ 50mg tablets (Wockhardt UK Ltd) |
| 74017 | 539211100001103 | Amiloride / Hydrochlorothiazide | Moduretic 5mg/ 50mg tablets (Waymade Healthcare Plc) |

| | | | |
|---|---|---|---|
| 75069 | 30015911000001100 | Amiloride / Hydrochlorothiazide | Co-amilozide 2.5mg/ 25mg tablets (Mawdsley-Brooks & Company Ltd) |
| 56 | 318136009 | Amiloride / Furosemide | Co-amilofruse 5mg/ 40mg tablets |
| 193 | 318135008 | Amiloride / Furosemide | Co-amilofruse 2.5mg/ 20mg tablets |
| 211 | 40605001000027105 | Amiloride / Furosemide | Frumil 40mg+5mg Tablet (Helios Healthcare Ltd) |
| 1301 | 40615001000027107 | Amiloride / Furosemide | Frumil ls 20mg+2.5mg Tablet (Helios Healthcare Ltd) |
| 2772 | 678511000001106 | Amiloride / Furosemide | Lasoride 5mg/ 40mg tablets (Sanofi) |
| 3793 | 318137000 | Amiloride / Furosemide | Co-amilofruse 10mg/ 80mg tablets |
| 4873 | 818511000001106 | Amiloride / Furosemide | Fru-Co 5mg/ 40mg tablets (Teva UK Ltd) |
| 9431 | 22255001000027108 | Amiloride / Furosemide | Frusemek 40mg+5mg Tablet (Approved Prescription Services Ltd) |
| 13435 | 82611000001106 | Amiloride / Furosemide | Frumil Forte 10mg/ 80mg tablets (Sanofi) |
| 18332 | 19683501000027101 | Amiloride / Furosemide | Aridil 20mg+2.5mg Tablet (C P Pharmaceuticals Ltd) |
| 21938 | 331311000001103 | Amiloride / Furosemide | Froop Co 5mg/ 40mg tablets (Ashbourne Pharmaceuticals Ltd) |
| 25965 | 213911000001104 | Amiloride / Furosemide | Co-amilofruse 2.5mg/ 20mg tablets (Wockhardt UK Ltd) |
| 28129 | 891211000001100 | Amiloride / Furosemide | Co-amilofruse 5mg/ 40mg tablets (Teva UK Ltd) |
| 30773 | 47575001000027101 | Amiloride / Furosemide | Co-amilofruse 5mg+40mg Tablet (Berk Pharmaceuticals Ltd) |
| 31773 | 380811000001106 | Amiloride / Furosemide | Co-amilofruse 5mg/ 40mg tablets (Wockhardt UK Ltd) |
| 33527 | 428711000001104 | Amiloride / Furosemide | Co-amilofruse 5mg/ 40mg tablets (Mylan) |
| 33658 | 11311000001108 | Amiloride / Furosemide | Co-amilofruse 5mg/ 40mg tablets (A A H Pharmaceuticals Ltd) |
| 34280 | 901811000001108 | Amiloride / Furosemide | Co-amilofruse 2.5mg/ 20mg tablets (Sandoz Ltd) |
| 34622 | 457611000001105 | Amiloride / Furosemide | Co-amilofruse 10mg/ 80mg tablets (Wockhardt UK Ltd) |
| 38901 | 550711000001106 | Amiloride / Furosemide | Frumil LS 20mg/ 2.5mg tablets (Sanofi) |
| 39807 | 427411000001106 | Amiloride / Furosemide | Frumil 40mg/ 5mg tablets (Sanofi) |
| 41533 | 242811000001109 | Amiloride / Furosemide | Co-amilofruse 2.5mg/ 20mg tablets (Teva UK Ltd) |
| 41719 | 667111000001108 | Amiloride / Furosemide | Co-amilofruse 5mg/ 40mg tablets (Actavis UK Ltd) |
| 43508 | 295411000001105 | Amiloride / Furosemide | Co-amilofruse 5mg/ 40mg tablets (Sandoz Ltd) |
| 57908 | 571911000001103 | Amiloride / Furosemide | Co-amilofruse 5mg/ 40mg tablets (Kent Pharmaceuticals Ltd) |
| 59412 | 21939011000001108 | Amiloride / Furosemide | Co-amilofruse 5mg/ 40mg tablets (Waymade Healthcare Plc) |
| 60258 | 19189511000001103 | Amiloride / Furosemide | Co-amilofruse 2.5mg/ 20mg tablets (Milpharm Ltd) |
| 71348 | 5372111000001107 | Amiloride / Furosemide | Frumil 40mg/ 5mg tablets (Waymade Healthcare Plc) |
| 71377 | 10855211000001104 | Amiloride / Furosemide | Frumil LS 20mg/ 2.5mg tablets (Waymade Healthcare Plc) |
| 73993 | 14244511000001107 | Amiloride / Furosemide | Frumil 40mg/ 5mg tablets (Sigma Pharmaceuticals Plc) |
| 74800 | 16182411000001100 | Amiloride / Furosemide | Frumil 40mg/ 5mg tablets (Lexon (UK) Ltd) |
| 2255 | 535711000001100 | Amiloride / Cyclopenthiazide | Navispare 2.5mg/ 250microgram tablets (AMCo) |
| 5727 | 318096005 | Amiloride / Cyclopenthiazide | Amiloride 2.5mg / Cyclopenthiazide 250microgram tablets |
| 1060 | 318052005 | Amiloride | Amiloride 5mg tablets |
| 9935 | 35900111000001108 | Amiloride | Amiloride 5mg/ 5ml oral solution sugar free |
| 13352 | 18905001000027107 | Amiloride | Midamor 5mg Tablet (MSD Thomas Morson Pharmaceuticals) |
| 24893 | 18128501000027104 | Amiloride | Amilospare Tablet (Ashbourne Pharmaceuticals Ltd) |
| 26217 | 15667501000027106 | Amiloride | Berkamil 5mg Tablet (Berk Pharmaceuticals Ltd) |
| 31375 | 799711000001108 | Amiloride | Amilamont 5mg/ 5ml oral solution sugar free (Rosemont Pharmaceuticals Ltd) |
| 33837 | 382611000001108 | Amiloride | Amiloride 5mg tablets (A A H Pharmaceuticals Ltd) |
| 34324 | 446011000001103 | Amiloride | Amiloride 5mg tablets (Teva UK Ltd) |
| 34750 | 754911000001104 | Amiloride | Amiloride 5mg tablets (Actavis UK Ltd) |

| | | | |
|---|---|---|---|
| 41630 | 3955001000027101 | Amiloride | Amiloride 5mg Tablet (IVAX Pharmaceuticals UK Ltd) |
| 43523 | 505611000001103 | Amiloride | Amiloride 5mg tablets (Mylan) |
| 44254 | 86595001000027100 | Amiloride | Amiloride 5.67mg tablets |
| 46930 | 533611000001105 | Amiloride | Amiloride 5mg tablets (Wockhardt UK Ltd) |
| 60149 | 8274111000001104 | Amiloride | Amiloride 5mg/ 5ml oral suspension |
| 76165 | | Amiloride | Amiloride 5mg tablets (Accord Healthcare Ltd) |
| 36519 | 10595001000027100 | Potassium Chloride/ Hydrochlorothiazide | "Esidrex -k Tablet (Novartis Pharmaceuticals UK Ltd)" |
| 1125 | 20765001000027100 | Potassium Chloride/ Cyclopenthiazide | "Navidrex -k Tablet (Novartis Pharmaceuticals UK Ltd)" |
| 2833 | 104585001000027108 | Potassium Chloride/ Cyclopenthiazide | "CYCLOPENTHIAZIDE -K tablets" |
| 1211 | 35910111000001106 | Potassium chloride/ Bendroflumethiazide | "Bendroflumethiazide 2.5mg / Potassium chloride 630mg (potassium 8.4mmol) modified-release tablets" |
| 1213 | 3638211000001100 | Potassium chloride/ Bendroflumethiazide | "Neo-Naclex-K modified-release tablets (Mercury Pharma Group Ltd)" |
| 2979 | 4955001000027104 | Potassium chloride/ Bendroflumethiazide | "Centyl k Tablet (Edwin Burgess Ltd)" |
| 17561 | 35910011000001105 | Bendroflumethiazide/ Potassium chloride | "Bendroflumethiazide 2.5mg / Potassium chloride 573mg (potassium 7.7mmol) modified-release tablets" |
| 20426 | 4965001000027108 | Bendroflumethiazide/ Potassium chloride | "Centyl k 2.5mg+7.7mmol Tablet (Edwin Burgess Ltd)" |
| 20431 | 3932711000001105 | Bendroflumethiazide/ Potassium chloride | "Centyl K modified-release tablets (LEO Pharma)" |
| 2681 | 185595001000027107 | | AMILORIDE 10 MG TAB |
| 3285 | 96205001000027106 | | AMILORIDE S/ F 5 MG/ 5ML SOL |
| 3962 | 187005001000027106 | | TRIAMTERENE 50MG HYDROCHLOROTHIAZIDE25MG TAB |
| 10796 | 188965001000027106 | | CHLORTHALIDONE 25MG/ POTASSIUM6.7MMOL S/ R MG TAB |
| 15053 | 96925001000027103 | | SPIRONOLACTONE 10 MG/ 5ML LIQ |
| 15602 | 167105001000027101 | | NATRILIX 5 MG TAB |
| 17721 | 158125001000027104 | | ALDACTIDE 100 MG TAB |
| 19611 | | | AMILORIDE 5MG/ HYDROCHLORTHIAZIDE 50MG |
| 19683 | | | BURINEX K |
| 19695 | | | AMILORIDE |
| 19721 | | | AMILORIDE 5MG/ HYDROCHLORTHIAZIDE 50MG |
| 20160 | 172315001000027106 | | CHLORTHALIDONE 500 MG TAB |
| 20513 | 167925001000027106 | | LASIX 10 MG INJ |
| 20779 | | | MODURETIC |
| 21848 | 4605001000027106 | | AMILOSPARE 5 MG TAB |
| 22539 | | | LASIX (2ML) |
| 23256 | | | LASIX PAED |
| 25630 | | | BRINALDIX K EFFERVESCENT |
| 26328 | | | LASIX (25ML) |

| | | | |
|---|---|---|---|
| 26675 | | | XIPAMIDE |
| 27555 | | | BURINEX |
| 29242 | 179705001000027102 | | CLOREXOLONE 10 MG TAB |
| 30368 | 172785001000027107 | | CHLOROTHIAZIDE/ SPIRONOLACTONE SACHETS 100 MG |
| 41889 | | | TRIAMTERENE 50MG HYDROCHLOROTHIAZIDE25MG |
| 13472 | 149755001000027102 | | "ESIDREX-K TAB" |
| 22242 | | | NAVIDREX |
| 22525 | 154075001000027109 | | "CHLOROTHIAZIDE 250 MG SYR" |
| 23492 | | | "CYCLOPENTHIAZIDE 250MCG/K 8.1MMOL" |
| 26120 | 154245001000027103 | | "CHLOROTHIAZIDE 50 MG SUS" |
| 27489 | 170715001000027104 | | "CHLOROTHIAZIDE 25 MG LIQ" |
| 31235 | 154235001000027104 | | "CHLOROTHIAZIDE SACHETS 60 MG" |
| **Renin inhibitors** | | | |
| 36629 | 425960005 | Aliskiren hemifumarate | Aliskiren 150mg tablets |
| 36878 | 11960911000001108 | Aliskiren hemifumarate | Rasilez 150mg tablets (Noden Pharma DAC) |
| 36879 | 11961711000001103 | Aliskiren hemifumarate | Rasilez 300mg tablets (Noden Pharma DAC) |
| 36909 | 425669009 | Aliskiren hemifumarate | Aliskiren 300mg tablets |
| **ORAL BISPHOSPHONATES** | | | |
| 9208 | 325979009 | Tiludronate disodium | Tiludronic acid 200mg tablets |
| 9525 | 4122511000001103 | Tiludronate disodium | Skelid 200mg tablets (Sanofi) |
| 3680 | 3848211000001108 | Sodium clodronate | Loron 400mg capsules (Roche Products Ltd) |
| 4868 | 325965001 | Sodium clodronate | Sodium clodronate 400mg capsules |
| 4927 | 3847911000001100 | Sodium clodronate | Bonefos 400mg capsules (Bayer Plc) |
| 5629 | 325972000 | Sodium clodronate | Sodium clodronate 800mg tablets |
| 6568 | 325970008 | Sodium clodronate | Sodium clodronate 520mg tablets |
| 9189 | 920611000001107 | Sodium clodronate | Bonefos 800mg tablets (Bayer Plc) |
| 11244 | 3813811000001107 | Sodium clodronate | Loron 520mg tablets (Intrapharm Laboratories Ltd) |
| 39043 | 11550411000001104 | Sodium clodronate | Clasteon 400mg capsules (Kent Pharmaceuticals Ltd) |
| 54989 | 20540511000001106 | Sodium clodronate | Clasteon 800mg tablets (Beacon Pharmaceuticals Ltd) |
| 45280 | 247875001000027107 | Risedronate Sodium/ Calcium Carbonate | Risedronate sodium 35mg & calcium carbonate 1250mg tablet |
| 6058 | 408027002 | Risedronate sodium | Risedronate sodium 35mg tablets |
| 6084 | 215955001000027104 | Risedronate sodium | Actonel once a week 35mg Tablet (Procter & Gamble (Health & Beauty Care) Ltd) |
| 6634 | 325983009 | Risedronate sodium | Risedronate sodium 5mg tablets |
| 7089 | 325984003 | Risedronate sodium | Risedronate sodium 30mg tablets |
| 7527 | 892511000001105 | Risedronate sodium | Actonel 5mg tablets (Warner Chilcott UK Ltd) |
| 7546 | 3778711000001100 | Risedronate sodium | Actonel 30mg tablets (Warner Chilcott UK Ltd) |

| | | | |
|---|---|---|---|
| 44511 | 4028511000001107 | Risedronate sodium | Actonel Once a Week 35mg tablets (Warner Chilcott UK Ltd) |
| 48013 | 18448311000001108 | Risedronate sodium | Risedronate sodium 35mg tablets (A A H Pharmaceuticals Ltd) |
| 52373 | 18626011000001106 | Risedronate sodium | Risedronate sodium 35mg tablets (Phoenix Healthcare Distribution Ltd) |
| 56431 | 18597911000001107 | Risedronate sodium | Risedronate sodium 35mg tablets (Actavis UK Ltd) |
| 56663 | 21887311000001101 | Risedronate sodium | Risedronate sodium 35mg tablets (Waymade Healthcare Plc) |
| 58618 | 16131211000001104 | Risedronate sodium | Risedronate sodium 35mg/ 5ml oral solution |
| 59449 | 19215711000001107 | Risedronate sodium | Risedronate sodium 35mg tablets (Bluefish Pharmaceuticals AB) |
| 59916 | 20889411000001109 | Risedronate sodium | Risedronate sodium 35mg tablets (Sandoz Ltd) |
| 60288 | 16255311000001105 | Risedronate sodium | Actonel Once a Week 35mg tablets (Mawdsley-Brooks & Company Ltd) |
| 61313 | 18448111000001106 | Risedronate sodium | Risedronate sodium 30mg tablets (A A H Pharmaceuticals Ltd) |
| 63802 | 19863711000001101 | Risedronate sodium | Actonel Once a Week 35mg tablets (Lexon (UK) Ltd) |
| 64431 | 18359911000001105 | Risedronate sodium | Risedronate sodium 30mg tablets (Aspire Pharma Ltd) |
| 65971 | 16131311000001107 | Risedronate sodium | Risedronate sodium 35mg/ 5ml oral suspension |
| 66028 | 13211011000001106 | Risedronate sodium | Actonel 35mg tablets (Teva UK Ltd) |
| 67078 | 18308311000001108 | Risedronate sodium | Risedronate sodium 35mg tablets (Teva UK Ltd) |
| 69630 | 20536311000001101 | Risedronate sodium | Risedronate sodium 35mg tablets (Almus Pharmaceuticals Ltd) |
| 69929 | 18344211000001102 | Risedronate sodium | Risedronate sodium 35mg tablets (Alliance Healthcare (Distribution) Ltd) |
| 69958 | 33614811000001104 | Risedronate sodium | Risedronate sodium 35mg tablets (Mylan) |
| 71209 | 30879111000001108 | Risedronate sodium | Risedronate sodium 35mg tablets (Mawdsley-Brooks & Company Ltd) |
| 73454 | 18359411000001102 | Risedronate sodium | Risedronate sodium 35mg tablets (Aspire Pharma Ltd) |
| 73989 | 13098211000001102 | Risedronate sodium | Actonel Once a Week 35mg tablets (Dowelhurst Ltd) |
| 74805 | 13823311000001109 | Risedronate sodium | Actonel Once a Week 35mg tablets (DE Pharmaceuticals) |
| 76178 | | Risedronate sodium | Actonel 5mg tablets (Sigma Pharmaceuticals Plc) |
| 76190 | | Risedronate sodium | Risedronate sodium 5mg tablets (A A H Pharmaceuticals Ltd) |
| 7112 | 9544911000001107 | Ibandronic sodium monohydrate | Bonviva 150mg tablets (Roche Products Ltd) |
| 7146 | 9553111000001105 | Ibandronic sodium monohydrate | Ibandronic acid 150mg tablets |
| 10193 | 410948006 | Ibandronic sodium monohydrate | Ibandronic acid 50mg tablets |
| 26913 | 7540111000001106 | Ibandronic sodium monohydrate | Bondronat 50mg tablets (Roche Products Ltd) |
| 47911 | 19371411000001104 | Ibandronic sodium monohydrate | Iasibon 50mg tablets (Aspire Pharma Ltd) |
| 51342 | 13837111000001101 | Ibandronic sodium monohydrate | Bonviva 150mg tablets (DE Pharmaceuticals) |
| 54453 | 19866411000001102 | Ibandronic sodium monohydrate | Bonviva 150mg tablets (Lexon (UK) Ltd) |
| 56030 | 20641211000001107 | Ibandronic sodium monohydrate | Ibandronic acid 150mg tablets (A A H Pharmaceuticals Ltd) |
| 56369 | 22086511000001105 | Ibandronic sodium monohydrate | Ibandronic acid 150mg tablets (Zentiva) |
| 57980 | 19295711000001108 | Ibandronic sodium monohydrate | Ibandronic acid 50mg tablets (Actavis UK Ltd) |
| 59587 | 23166211000001109 | Ibandronic sodium monohydrate | Ibandronic acid 150mg tablets (Ranbaxy (UK) Ltd) |
| 67159 | 20596011000001106 | Ibandronic sodium monohydrate | Ibandronic acid 50mg tablets (Teva UK Ltd) |
| 71000 | 20971311000001104 | Ibandronic sodium monohydrate | Ibandronic acid 150mg tablets (Alliance Healthcare (Distribution) Ltd) |
| 75425 | | Ibandronic sodium monohydrate | Ibandronic acid 50mg tablets (DE Pharmaceuticals) |
| 75644 | | Ibandronic sodium monohydrate | Quodixor 150mg tablets (Aspire Pharma Ltd) |
| 76545 | | Ibandronic sodium monohydrate | Bonviva 150mg tablets (Mawdsley-Brooks & Company Ltd) |
| 77225 | | Ibandronic sodium monohydrate | Ibandronic acid 150mg tablets (Teva UK Ltd) |
| 78002 | | Ibandronic sodium monohydrate | Ibandronic acid 150mg tablets (Mylan) |
| 766 | 3356811000001108 | Etidronate disodium | Didronel 200mg tablets (Warner Chilcott UK Ltd) |

| | | | |
|---|---|---|---|
| 4680 | 325951003 | Etidronate disodium | Etidronate disodium 200mg tablets |
| 63371 | 5196211000001102 | Etidronate disodium | Etidronate disodium 200mg tablets (Mylan) |
| 77070 | | Etidronate disodium | Etidronate disodium 100mg/ 5ml oral suspension |
| 77996 | | Etidronate disodium | Didronel 400mg tablets (Mawdsley-Brooks & Company Ltd) |
| 53169 | 224555001000027100 | Disodium Etidronate | Etidronate disodium 400mg Tablet |
| 54436 | 247235001000027103 | Disodium Etidronate | Etidronate disodium Oral solution |
| 37575 | 249915001000027103 | Colecalciferol/ Risedronate Sodium/ Calcium Carbonate | Risedronate sodium 35mg with calcium carbonate 2500mg & colecalciferol 22micrograms tablets and granules |
| 7224 | 9526611000001107 | Colecalciferol/ Alendronate sodium | Alendronic acid 70mg / Colecalciferol 70microgram tablets |
| 10227 | 9523811000001102 | Colecalciferol/ Alendronate sodium | Fosavance tablets (Merck Sharp & Dohme Ltd) |
| 66485 | 16182011000001109 | Colecalciferol/ Alendronate sodium | Fosavance tablets (Lexon (UK) Ltd) |
| 70927 | 34741911000001106 | Colecalciferol/ Alendronate sodium | Alendronic acid 70mg / Colecalciferol 70microgram tablets (Creo Pharma Ltd) |
| 76864 | | Colecalciferol/ Alendronate sodium | Alendronic acid 70mg / Colecalciferol 140microgram tablets |
| 45787 | 18683211000001101 | Alendronic acid | Alendronic acid 70mg/ 100ml oral solution unit dose sugar free |
| 52564 | 18680211000001106 | Alendronic acid | Alendronic acid 70mg/ 100ml oral solution unit dose sugar free (Rosemont Pharmaceuticals Ltd) |
| 55295 | 19185711000001106 | Alendronic acid | Alendronic acid 70mg/ 100ml oral solution unit dose sugar free (Alliance Healthcare (Distribution) Ltd) |
| 55998 | 20920711000001107 | Alendronic acid | Alendronic acid 70mg/ 75ml oral solution unit dose |
| 60144 | 21755811000001108 | Alendronic acid | Alendronic acid 70mg/ 100ml oral solution unit dose sugar free (Waymade Healthcare Plc) |
| 72208 | 20005911000001103 | Alendronic acid | Alendronic acid 70mg/ 100ml oral solution unit dose sugar free (A A H Pharmaceuticals Ltd) |
| 544 | 417211000001103 | Alendronate sodium | Fosamax Once Weekly 70mg tablets (Merck Sharp & Dohme Ltd) |
| 663 | 726311000001107 | Alendronate sodium | Fosamax 10mg tablets (Merck Sharp & Dohme Ltd) |
| 688 | 134599008 | Alendronate sodium | Alendronic acid 70mg tablets |
| 782 | 3145911000001101 | Alendronate sodium | Fosamax 5mg tablets (Merck Sharp & Dohme Ltd) |
| 2298 | 325974004 | Alendronate sodium | Alendronic acid 10mg tablets |
| 7530 | 325977006 | Alendronate sodium | Alendronic acid 5mg tablets |
| 35937 | 9221911000001101 | Alendronate sodium | Alendronic acid 70mg tablets (A A H Pharmaceuticals Ltd) |
| 37217 | 9251711000001102 | Alendronate sodium | Alendronic acid 10mg tablets (Teva UK Ltd) |
| 37218 | 9188811000001108 | Alendronate sodium | Alendronic acid 70mg tablets (Teva UK Ltd) |
| 40449 | 9836811000001109 | Alendronate sodium | Alendronic acid 70mg tablets (PLIVA Pharma Ltd) |
| 43958 | 9830711000001104 | Alendronate sodium | Alendronic acid 70mg tablets (Actavis UK Ltd) |
| 46245 | 9554311000001109 | Alendronate sodium | Alendronic acid 70mg tablets (Mylan) |
| 47380 | 10447611000001104 | Alendronate sodium | Alendronic acid 70mg tablets (Arrow Generics Ltd) |
| 50278 | 13441411000001109 | Alendronate sodium | Alendronic acid 70mg tablets (Wockhardt UK Ltd) |
| 50880 | 18264111000001108 | Alendronate sodium | Fosamax 10mg tablets (Necessity Supplies Ltd) |
| 51877 | 9299611000001102 | Alendronate sodium | Alendronic acid 70mg tablets (Alliance Healthcare (Distribution) Ltd) |
| 52284 | 14240211000001101 | Alendronate sodium | Fosamax 10mg tablets (Sigma Pharmaceuticals Plc) |
| 52624 | 17756411000001101 | Alendronate sodium | Alendronic acid 70mg tablets (Phoenix Healthcare Distribution Ltd) |
| 52834 | 18455911000001100 | Alendronate sodium | Alendronic acid 70mg tablets (Accord Healthcare Ltd) |
| 54566 | 9252411000001103 | Alendronate sodium | Alendronic acid 10mg tablets (A A H Pharmaceuticals Ltd) |

| | | | |
|---|---|---|---|
| 55965 | 9990311000001102 | Alendronate sodium | Alendronic acid 70mg tablets (Zentiva) |
| 56061 | 10435111000001104 | Alendronate sodium | Alendronic acid 10mg tablets (Actavis UK Ltd) |
| 56260 | 9208911000001102 | Alendronate sodium | Alendronic acid 70mg tablets (Kent Pharmaceuticals Ltd) |
| 56730 | 17963711000001100 | Alendronate sodium | Alendronic acid 70mg tablets (Almus Pharmaceuticals Ltd) |
| 57875 | 16181811000001107 | Alendronate sodium | Fosamax Once Weekly 70mg tablets (Lexon (UK) Ltd) |
| 58744 | 13880811000001102 | Alendronate sodium | Fosamax Once Weekly 70mg tablets (DE Pharmaceuticals) |
| 59079 | 17961911000001104 | Alendronate sodium | Alendronic acid 10mg tablets (Almus Pharmaceuticals Ltd) |
| 59247 | 18264311000001105 | Alendronate sodium | Fosamax Once Weekly 70mg tablets (Necessity Supplies Ltd) |
| 59485 | 18455711000001102 | Alendronate sodium | Alendronic acid 10mg tablets (Accord Healthcare Ltd) |
| 59555 | 9452511000001100 | Alendronate sodium | Alendronic acid 10mg tablets (Alliance Healthcare (Distribution) Ltd) |
| 61686 | 19701111000001102 | Alendronate sodium | Alendronic acid 70mg tablets (DE Pharmaceuticals) |
| 63008 | 24111211000001100 | Alendronate sodium | Alendronic acid 70mg tablets (Somex Pharma) |
| 63175 | 17756211000001100 | Alendronate sodium | Alendronic acid 10mg tablets (Phoenix Healthcare Distribution Ltd) |
| 64331 | 19700911000001106 | Alendronate sodium | Alendronic acid 10mg tablets (DE Pharmaceuticals) |
| 65008 | 30317811000001101 | Alendronate sodium | Alendronic acid 70mg effervescent tablets sugar free |
| 65905 | 9554111000001107 | Alendronate sodium | Alendronic acid 10mg tablets (Mylan) |
| 66203 | 30316711000001106 | Alendronate sodium | Binosto 70mg effervescent tablets (Internis Pharmaceuticals Ltd) |
| 69995 | 15631411000001101 | Alendronate sodium | Alendronic acid 35mg/ 5ml oral solution |
| 71851 | 15060811000001107 | Alendronate sodium | Alendronic acid 70mg tablets (Sigma Pharmaceuticals Plc) |
| 71963 | 15060411000001105 | Alendronate sodium | Alendronic acid 10mg tablets (Sigma Pharmaceuticals Plc) |
| 72541 | 15166911000001107 | Alendronate sodium | Alendronic acid 70mg/ 5ml oral solution |
| 73560 | 10688611000001108 | Alendronate sodium | Alendronic acid 10mg tablets (PLIVA Pharma Ltd) |
| 74859 | 5366911000001104 | Alendronate sodium | Fosamax Once Weekly 70mg tablets (Waymade Healthcare Plc) |
| 75094 | 21756211000001101 | Alendronate sodium | Alendronic acid 10mg tablets (Waymade Healthcare Plc) |
| 77297 | | Alendronate sodium | Alendronic acid 70mg tablets (Focus Pharmaceuticals Ltd) |
| 110 | 151675001000027109 | | DIDRONEL 100 MG TAB |
| 468 | 3352411000001105 | | Didronel PMO tablets (Warner Chilcott UK Ltd) |
| 3046 | 81085001000027103 | | DISODIUM ETIDRONATE 200 MG TAB |
| 11368 | 3613311000001109 | | Calcium carbonate 1.25g effervescent tablets and Disodium etidronate 400mg tablets |
| 37833 | 13208811000001109 | | Actonel Combi 35mg tablets and 1000mg/ 880unit effervescent granules sachets (Teva UK Ltd) |
| **STATINS** | | | |
| 66505 | 32234311000001109 | Simvastatin/ Fenofibrate | Fenofibrate 145mg / Simvastatin 40mg tablets |
| 66780 | 32234211000001101 | Simvastatin/ Fenofibrate | Fenofibrate 145mg / Simvastatin 20mg tablets |
| 69528 | 32170911000001100 | Simvastatin/ Fenofibrate | Cholib 145mg/ 20mg tablets (Mylan) |
| 70486 | 32169911000001103 | Simvastatin/ Fenofibrate | Cholib 145mg/ 40mg tablets (Mylan) |
| 7552 | 414177002 | Simvastatin/ Ezetimibe | Simvastatin 20mg / Ezetimibe 10mg tablets |
| 10172 | 414178007 | Simvastatin/ Ezetimibe | Simvastatin 40mg / Ezetimibe 10mg tablets |
| 10183 | 240715001000027105 | Simvastatin/ Ezetimibe | Simvastatin 40mg with ezetimibe 10mg tablet |
| 10206 | 240725001000027101 | Simvastatin/ Ezetimibe | Simvastatin 80mg with ezetimibe 10mg tablet |
| 11815 | 240705001000027108 | Simvastatin/ Ezetimibe | Simvastatin 20mg with ezetimibe 10mg tablet |
| 14219 | 414179004 | Simvastatin/ Ezetimibe | Simvastatin 80mg / Ezetimibe 10mg tablets |

| | | | |
|---|---|---|---|
| 16186 | 9310611000001103 | Simvastatin/ Ezetimibe | Inegy 10mg/ 80mg tablets (Merck Sharp & Dohme Ltd) |
| 17059 | 9310311000001108 | Simvastatin/ Ezetimibe | Inegy 10mg/ 40mg tablets (Merck Sharp & Dohme Ltd) |
| 21020 | 9309911000001100 | Simvastatin/ Ezetimibe | Inegy 10mg/ 20mg tablets (Merck Sharp & Dohme Ltd) |
| 25 | 319997009 | Simvastatin | Simvastatin 20mg tablets |
| 42 | 319996000 | Simvastatin | Simvastatin 10mg tablets |
| 51 | 320000009 | Simvastatin | Simvastatin 40mg tablets |
| 802 | 4896711000001108 | Simvastatin | Simvador 40mg tablets (Discovery Pharmaceuticals) |
| 818 | 242705001000027101 | Simvastatin | Simvastatin 20mg/ 5ml oral solution sugar free |
| 2718 | 108111000001106 | Simvastatin | Zocor 10mg tablets (Merck Sharp & Dohme Ltd) |
| 5148 | 320006003 | Simvastatin | Simvastatin 80mg tablets |
| 6168 | 859611000001107 | Simvastatin | Zocor 40mg tablets (Merck Sharp & Dohme Ltd) |
| 7196 | 776811000001104 | Simvastatin | Zocor 20mg tablets (Merck Sharp & Dohme Ltd) |
| 9920 | 4896511000001103 | Simvastatin | Simvador 20mg tablets (Discovery Pharmaceuticals) |
| 13041 | 4896211000001101 | Simvastatin | Simvador 10mg tablets (Discovery Pharmaceuticals) |
| 22579 | 113211000001106 | Simvastatin | Zocor 80mg tablets (Merck Sharp & Dohme Ltd) |
| 31930 | 238335001000027101 | Simvastatin | Zocor heart-pro 10mg Tablet (McNeil Products Ltd) |
| 32909 | 4579211000001101 | Simvastatin | Simvastatin 80mg tablets (A A H Pharmaceuticals Ltd) |
| 33082 | 4578811000001107 | Simvastatin | Simvastatin 20mg tablets (A A H Pharmaceuticals Ltd) |
| 34312 | 4464011000001108 | Simvastatin | Simvastatin 20mg tablets (Mylan) |
| 34316 | 4380911000001108 | Simvastatin | Simvastatin 20mg tablets (Teva UK Ltd) |
| 34353 | 4464211000001103 | Simvastatin | Simvastatin 40mg tablets (Mylan) |
| 34366 | 4466511000001106 | Simvastatin | Simvastatin 20mg tablets (IVAX Pharmaceuticals UK Ltd) |
| 34376 | 4381111000001104 | Simvastatin | Simvastatin 40mg tablets (Teva UK Ltd) |
| 34381 | 4466711000001101 | Simvastatin | Simvastatin 40mg tablets (IVAX Pharmaceuticals UK Ltd) |
| 34476 | 134095001000027108 | Simvastatin | Simvastatin 20mg Tablet (Ratiopharm UK Ltd) |
| 34481 | 4466211000001108 | Simvastatin | Simvastatin 10mg tablets (IVAX Pharmaceuticals UK Ltd) |
| 34502 | 4579011000001106 | Simvastatin | Simvastatin 40mg tablets (A A H Pharmaceuticals Ltd) |
| 34535 | 4463811000001100 | Simvastatin | Simvastatin 10mg tablets (Mylan) |
| 34545 | 134145001000027108 | Simvastatin | Simvastatin 40mg Tablet (Ratiopharm UK Ltd) |
| 34560 | 134055001000027102 | Simvastatin | Simvastatin 10mg Tablet (Ratiopharm UK Ltd) |
| 34746 | 136355001000027109 | Simvastatin | Simvastatin 20mg Tablet (Niche Generics Ltd) |
| 34814 | 4480511000001107 | Simvastatin | Simvastatin 20mg tablets (Wockhardt UK Ltd) |
| 34879 | 136385001000027102 | Simvastatin | Simvastatin 40mg Tablet (Niche Generics Ltd) |
| 34891 | 4467111000001104 | Simvastatin | Simvastatin 20mg tablets (Kent Pharmaceuticals Ltd) |
| 34907 | 4480711000001102 | Simvastatin | Simvastatin 40mg tablets (Wockhardt UK Ltd) |
| 34955 | 4578011000001101 | Simvastatin | Simvastatin 10mg tablets (A A H Pharmaceuticals Ltd) |
| 34969 | 4437211000001100 | Simvastatin | Simvastatin 40mg tablets (Actavis UK Ltd) |
| 37434 | 4580311000001109 | Simvastatin | Simvastatin 40mg tablets (Sandoz Ltd) |
| 39060 | 11551511000001107 | Simvastatin | Simvastatin 20mg tablets (Dexcel-Pharma Ltd) |
| 39652 | 256505001000027107 | Simvastatin | Simvastatin 40mg/ 5ml oral solution sugar free |
| 39675 | 183305001000027103 | Simvastatin | Simvastatin 20mg/ 5ml Oral suspension (Martindale Pharmaceuticals Ltd) |
| 39870 | 15158611000001106 | Simvastatin | Simvador 80mg tablets (Discovery Pharmaceuticals) |
| 40340 | 4380611000001102 | Simvastatin | Simvastatin 10mg tablets (Teva UK Ltd) |

| | | | |
|---|---|---|---|
| 40601 | 4465311000001105 | Simvastatin | Simvastatin 20mg tablets (Ranbaxy (UK) Ltd) |
| 41657 | 5476111000001102 | Simvastatin | Simvastatin 80mg tablets (Teva UK Ltd) |
| 44528 | 17305411000001100 | Simvastatin | Simvastatin 20mg/ 5ml oral suspension sugar free (Rosemont Pharmaceuticals Ltd) |
| 44650 | 11551711000001102 | Simvastatin | Simvastatin 40mg tablets (Dexcel-Pharma Ltd) |
| 44878 | 7630211000001106 | Simvastatin | Ranzolont 10mg tablets (Ranbaxy (UK) Ltd) |
| 45219 | 4467311000001102 | Simvastatin | Simvastatin 40mg tablets (Kent Pharmaceuticals Ltd) |
| 45235 | 4580111000001107 | Simvastatin | Simvastatin 20mg tablets (Sandoz Ltd) |
| 45245 | 4437011000001105 | Simvastatin | Simvastatin 20mg tablets (Actavis UK Ltd) |
| 45346 | 10414211000001101 | Simvastatin | Simvastatin 40mg tablets (Arrow Generics Ltd) |
| 46878 | 9804311000001100 | Simvastatin | Simvastatin 40mg tablets (Almus Pharmaceuticals Ltd) |
| 46956 | 10414411000001102 | Simvastatin | Simvastatin 80mg tablets (Arrow Generics Ltd) |
| 47774 | 10413811000001103 | Simvastatin | Simvastatin 10mg tablets (Arrow Generics Ltd) |
| 47948 | 13762911000001107 | Simvastatin | Simvastatin 10mg tablets (Tillomed Laboratories Ltd) |
| 48018 | 10414011000001106 | Simvastatin | Simvastatin 20mg tablets (Arrow Generics Ltd) |
| 48051 | 4466911000001104 | Simvastatin | Simvastatin 10mg tablets (Kent Pharmaceuticals Ltd) |
| 48058 | 4465011000001107 | Simvastatin | Simvastatin 10mg tablets (Ranbaxy (UK) Ltd) |
| 48078 | 4436811000001101 | Simvastatin | Simvastatin 10mg tablets (Actavis UK Ltd) |
| 48221 | 17369311000001105 | Simvastatin | Simvastatin 20mg/ 5ml oral suspension sugar free |
| 48431 | 17429811000001102 | Simvastatin | Simvastatin 40mg/ 5ml oral suspension sugar free |
| 48867 | 4574811000001104 | Simvastatin | Simvastatin 40mg tablets (Alliance Healthcare (Distribution) Ltd) |
| 49061 | 16067211000001108 | Simvastatin | Simvastatin 40mg tablets (Bristol Laboratories Ltd) |
| 49062 | 4574611000001103 | Simvastatin | Simvastatin 20mg tablets (Alliance Healthcare (Distribution) Ltd) |
| 49587 | 9804011000001103 | Simvastatin | Simvastatin 80mg tablets (Almus Pharmaceuticals Ltd) |
| 50483 | 10297511000001106 | Simvastatin | Simvastatin 40mg tablets (Relonchem Ltd) |
| 50564 | 10297311000001100 | Simvastatin | Simvastatin 20mg tablets (Relonchem Ltd) |
| 50670 | 19197811000001107 | Simvastatin | Simvastatin 40mg tablets (Milpharm Ltd) |
| 50703 | 18468511000001107 | Simvastatin | Simvastatin 40mg tablets (Accord Healthcare Ltd) |
| 50754 | 19733411000001104 | Simvastatin | Simvastatin 20mg tablets (Medreich Plc) |
| 50882 | 10618911000001100 | Simvastatin | Simvastatin 40mg tablets (Somex Pharma) |
| 51085 | 19733211000001103 | Simvastatin | Simvastatin 10mg tablets (Medreich Plc) |
| 51166 | 19733611000001101 | Simvastatin | Simvastatin 40mg tablets (Medreich Plc) |
| 51233 | 4574411000001101 | Simvastatin | Simvastatin 10mg tablets (Alliance Healthcare (Distribution) Ltd) |
| 51483 | 19197611000001108 | Simvastatin | Simvastatin 20mg tablets (Milpharm Ltd) |
| 51715 | 15188311000001107 | Simvastatin | Simvastatin 10mg tablets (Sigma Pharmaceuticals Plc) |
| 52098 | 4465611000001100 | Simvastatin | Simvastatin 40mg tablets (Ranbaxy (UK) Ltd) |
| 52257 | 18468011000001104 | Simvastatin | Simvastatin 20mg tablets (Accord Healthcare Ltd) |
| 52625 | 4480311000001101 | Simvastatin | Simvastatin 10mg tablets (Wockhardt UK Ltd) |
| 52676 | 8722011000001100 | Simvastatin | Simvastatin 10mg/ 5ml oral suspension |
| 52812 | 15171911000001107 | Simvastatin | Simvastatin 20mg tablets (Sigma Pharmaceuticals Plc) |
| 52953 | 16066911000001102 | Simvastatin | Simvastatin 20mg tablets (Bristol Laboratories Ltd) |
| 52962 | 19733811000001102 | Simvastatin | Simvastatin 80mg tablets (Medreich Plc) |
| 53087 | 10618711000001102 | Simvastatin | Simvastatin 20mg tablets (Somex Pharma) |
| 53340 | 16451611000001102 | Simvastatin | Zocor 40mg tablets (Lexon (UK) Ltd) |

| | | | |
|---|---|---|---|
| 53415 | 19197411000001105 | Simvastatin | Simvastatin 10mg tablets (Milpharm Ltd) |
| 53676 | 13763111000001103 | Simvastatin | Simvastatin 20mg tablets (Tillomed Laboratories Ltd) |
| 53822 | 16066711000001104 | Simvastatin | Simvastatin 10mg tablets (Bristol Laboratories Ltd) |
| 53908 | 11551311000001101 | Simvastatin | Simvastatin 10mg tablets (Dexcel-Pharma Ltd) |
| 53966 | 17793911000001109 | Simvastatin | Simvastatin 40mg tablets (Phoenix Healthcare Distribution Ltd) |
| 54240 | 15172411000001109 | Simvastatin | Simvastatin 40mg tablets (Sigma Pharmaceuticals Plc) |
| 54266 | 8722111000001104 | Simvastatin | Simvastatin 20mg/ 5ml oral suspension |
| 54493 | 10296911000001102 | Simvastatin | Simvastatin 10mg tablets (Relonchem Ltd) |
| 54606 | 18757511000001106 | Simvastatin | Simvastatin 20mg/ 5ml oral suspension sugar free (A A H Pharmaceuticals Ltd) |
| 54655 | 18467811000001106 | Simvastatin | Simvastatin 10mg tablets (Accord Healthcare Ltd) |
| 54819 | 17305711000001106 | Simvastatin | Simvastatin 40mg/ 5ml oral suspension sugar free (Rosemont Pharmaceuticals Ltd) |
| 54947 | 9804811000001109 | Simvastatin | Simvastatin 20mg tablets (Almus Pharmaceuticals Ltd) |
| 54976 | 10618511000001107 | Simvastatin | Simvastatin 10mg tablets (Somex Pharma) |
| 54985 | 13894411000001106 | Simvastatin | Simvastatin 40mg/ 5ml oral suspension |
| 55452 | 17785511000001104 | Simvastatin | Simvastatin 20mg tablets (Phoenix Healthcare Distribution Ltd) |
| 56065 | 21898011000001102 | Simvastatin | Simvastatin 20mg/ 5ml oral suspension sugar free (Waymade Healthcare Plc) |
| 56481 | 14732411000001109 | Simvastatin | Zocor 10mg tablets (Sigma Pharmaceuticals Plc) |
| 56494 | 14732811000001106 | Simvastatin | Zocor 20mg tablets (Sigma Pharmaceuticals Plc) |
| 57329 | 16091011000001105 | Simvastatin | Simvastatin 25mg/ 5ml oral suspension |
| 57568 | 16451411000001100 | Simvastatin | Zocor 10mg tablets (Lexon (UK) Ltd) |
| 58315 | 21899311000001105 | Simvastatin | Simvastatin 20mg tablets (Waymade Healthcare Plc) |
| 58755 | 17928611000001107 | Simvastatin | Simvastatin 10mg tablets (Phoenix Healthcare Distribution Ltd) |
| 61155 | 18758211000001107 | Simvastatin | Simvastatin 40mg/ 5ml oral suspension sugar free (A A H Pharmaceuticals Ltd) |
| 61321 | 4579911000001105 | Simvastatin | Simvastatin 10mg tablets (Sandoz Ltd) |
| 61360 | 9805011000001104 | Simvastatin | Simvastatin 10mg tablets (Almus Pharmaceuticals Ltd) |
| 61665 | 21899011000001107 | Simvastatin | Simvastatin 10mg tablets (Waymade Healthcare Plc) |
| 62137 | 21899611000001100 | Simvastatin | Simvastatin 40mg tablets (Waymade Healthcare Plc) |
| 64104 | 28988511000001107 | Simvastatin | Simvastatin 20mg tablets (Crescent Pharma Ltd) |
| 64180 | 28918311000001104 | Simvastatin | Simvastatin 10mg tablets (Crescent Pharma Ltd) |
| 64307 | 28918111000001101 | Simvastatin | Simvastatin 40mg tablets (Crescent Pharma Ltd) |
| 64968 | 30097311000001108 | Simvastatin | Simvastatin 10mg tablets (DE Pharmaceuticals) |
| 65181 | 30097711000001107 | Simvastatin | Simvastatin 40mg tablets (DE Pharmaceuticals) |
| 65679 | 30097511000001102 | Simvastatin | Simvastatin 20mg tablets (DE Pharmaceuticals) |
| 65901 | 10733711000001109 | Simvastatin | Simvastatin 40mg tablets (Zentiva) |
| 65925 | 17841911000001100 | Simvastatin | Simvastatin 20mg/ 5ml oral suspension sugar free (Alliance Healthcare (Distribution) Ltd) |
| 67098 | 32494611000001109 | Simvastatin | Simvastatin 10mg tablets (Brown & Burk UK Ltd) |
| 67745 | 10733011000001107 | Simvastatin | Simvastatin 10mg tablets (Zentiva) |
| 67773 | 10733511000001104 | Simvastatin | Simvastatin 20mg tablets (Zentiva) |
| 68563 | 32495011000001103 | Simvastatin | Simvastatin 40mg tablets (Brown & Burk UK Ltd) |
| 68686 | 33555611000001103 | Simvastatin | Simvastatin 20mg tablets (Genesis Pharmaceuticals Ltd) |
| 69413 | 32494811000001108 | Simvastatin | Simvastatin 20mg tablets (Brown & Burk UK Ltd) |
| 71773 | 32495311000001100 | Simvastatin | Simvastatin 80mg tablets (Brown & Burk UK Ltd) |
| 72050 | 33654911000001100 | Simvastatin | Simvastatin 10mg tablets (Genesis Pharmaceuticals Ltd) |

| | | | |
|---|---|---|---|
| 75134 | 4464511000001100 | Simvastatin | Simvastatin 80mg tablets (Mylan) |
| 76481 | | Simvastatin | Simvastatin 80mg Tablet (Dexcel-Pharma Ltd) |
| 76594 | | Simvastatin | Simvastatin 40mg tablets (Tillomed Laboratories Ltd) |
| 77357 | | Simvastatin | Zocor 20mg tablets (Waymade Healthcare Plc) |
| 77358 | | Simvastatin | Zocor 10mg tablets (Waymade Healthcare Plc) |
| 77470 | | Simvastatin | Zocor 10mg tablets (Necessity Supplies Ltd) |
| 77471 | | Simvastatin | Zocor 20mg tablets (Necessity Supplies Ltd) |
| 713 | 408036003 | Rosuvastatin calcium | Rosuvastatin 10mg tablets |
| 6213 | 408037007 | Rosuvastatin calcium | Rosuvastatin 20mg tablets |
| 7347 | 4171011000001104 | Rosuvastatin calcium | Crestor 10mg tablets (AstraZeneca UK Ltd) |
| 7554 | 409108001 | Rosuvastatin calcium | Rosuvastatin 5mg tablets |
| 9897 | 408024009 | Rosuvastatin calcium | Rosuvastatin 40mg tablets |
| 9930 | 4172111000001108 | Rosuvastatin calcium | Crestor 40mg tablets (AstraZeneca UK Ltd) |
| 15252 | 4171311000001101 | Rosuvastatin calcium | Crestor 20mg tablets (AstraZeneca UK Ltd) |
| 17688 | 9747511000001107 | Rosuvastatin calcium | Crestor 5mg tablets (AstraZeneca UK Ltd) |
| 53460 | 13857911000001102 | Rosuvastatin calcium | Crestor 10mg tablets (DE Pharmaceuticals) |
| 57763 | 10769311000001104 | Rosuvastatin calcium | Rosuvastatin 10mg tablets (Waymade Healthcare Plc) |
| 57999 | 16155011000001105 | Rosuvastatin calcium | Crestor 40mg tablets (Lexon (UK) Ltd) |
| 58617 | 16075311000001101 | Rosuvastatin calcium | Rosuvastatin 20mg/ 5ml oral suspension |
| 59447 | 10513311000001102 | Rosuvastatin calcium | Crestor 20mg tablets (Waymade Healthcare Plc) |
| 59452 | 12561111000001105 | Rosuvastatin calcium | Rosuvastatin 5mg tablets (Waymade Healthcare Plc) |
| 60160 | 18202311000001102 | Rosuvastatin calcium | Rosuvastatin 5mg tablets (Mawdsley-Brooks & Company Ltd) |
| 70308 | 14212311000001103 | Rosuvastatin calcium | Crestor 20mg tablets (Sigma Pharmaceuticals Plc) |
| 71014 | 11580111000001101 | Rosuvastatin calcium | Rosuvastatin 20mg tablets (Waymade Healthcare Plc) |
| 73025 | 35027811000001104 | Rosuvastatin calcium | Rosuvastatin 20mg tablets (Mylan) |
| 74552 | 35183311000001102 | Rosuvastatin calcium | Rosuvastatin 10mg tablets (Milpharm Ltd) |
| 75971 | | Rosuvastatin calcium | Rosuvastatin 10mg tablets (Sandoz Ltd) |
| 76120 | | Rosuvastatin calcium | Rosuvastatin 10mg tablets (Teva UK Ltd) |
| 490 | 320012008 | Pravastatin sodium | Pravastatin 10mg tablets |
| 730 | 320013003 | Pravastatin sodium | Pravastatin 20mg tablets |
| 1219 | 320014009 | Pravastatin sodium | Pravastatin 40mg tablets |
| 1221 | 802411000001108 | Pravastatin sodium | Lipostat 10mg tablets (Bristol-Myers Squibb Pharmaceuticals Ltd) |
| 1223 | 535011000001102 | Pravastatin sodium | Lipostat 40mg tablets (Bristol-Myers Squibb Pharmaceuticals Ltd) |
| 3690 | 454111000001107 | Pravastatin sodium | Lipostat 20mg tablets (Bristol-Myers Squibb Pharmaceuticals Ltd) |
| 32921 | 163075001000027105 | Pravastatin sodium | Pravastatin 10mg Tablet (Dr Reddy's Laboratories (UK) Ltd) |
| 34820 | 7977111000001100 | Pravastatin sodium | Pravastatin 40mg tablets (A A H Pharmaceuticals Ltd) |
| 36377 | 7943411000001102 | Pravastatin sodium | Pravastatin 20mg tablets (Teva UK Ltd) |
| 40382 | 7976911000001100 | Pravastatin sodium | Pravastatin 20mg tablets (A A H Pharmaceuticals Ltd) |
| 43218 | 7943211000001101 | Pravastatin sodium | Pravastatin 10mg tablets (Teva UK Ltd) |
| 47988 | 8113311000001107 | Pravastatin sodium | Pravastatin 40mg tablets (Mylan) |
| 48097 | 7943611000001104 | Pravastatin sodium | Pravastatin 40mg tablets (Teva UK Ltd) |
| 50925 | 15174911000001105 | Pravastatin sodium | Pravastatin 10mg tablets (Sigma Pharmaceuticals Plc) |
| 51676 | 19732111000001103 | Pravastatin sodium | Pravastatin 40mg tablets (Medreich Plc) |

| | | | |
|---|---|---|---|
| 51890 | 19731911000001106 | Pravastatin sodium | Pravastatin 20mg tablets (Medreich Plc) |
| 52755 | 8027311000001103 | Pravastatin sodium | Pravastatin 20mg tablets (Alliance Healthcare (Distribution) Ltd) |
| 54435 | 9807511000001103 | Pravastatin sodium | Pravastatin 40mg tablets (Almus Pharmaceuticals Ltd) |
| 54607 | 9806911000001107 | Pravastatin sodium | Pravastatin 20mg tablets (Almus Pharmaceuticals Ltd) |
| 55912 | 8027611000001108 | Pravastatin sodium | Pravastatin 40mg tablets (Alliance Healthcare (Distribution) Ltd) |
| 56146 | 21850611000001104 | Pravastatin sodium | Pravastatin 10mg tablets (Waymade Healthcare Plc) |
| 56607 | 21850811000001100 | Pravastatin sodium | Pravastatin 20mg tablets (Waymade Healthcare Plc) |
| 56735 | 8113111000001105 | Pravastatin sodium | Pravastatin 20mg tablets (Mylan) |
| 56893 | 18465611000001100 | Pravastatin sodium | Pravastatin 40mg tablets (Accord Healthcare Ltd) |
| 56916 | 11410811000001100 | Pravastatin sodium | Pravastatin 40mg tablets (PLIVA Pharma Ltd) |
| 57108 | 21851011000001102 | Pravastatin sodium | Pravastatin 40mg tablets (Waymade Healthcare Plc) |
| 57137 | 9806311000001106 | Pravastatin sodium | Pravastatin 10mg tablets (Almus Pharmaceuticals Ltd) |
| 57296 | 17915711000001103 | Pravastatin sodium | Pravastatin 20mg tablets (Phoenix Healthcare Distribution Ltd) |
| 57397 | 18465211000001102 | Pravastatin sodium | Pravastatin 10mg tablets (Accord Healthcare Ltd) |
| 59508 | 18465411000001103 | Pravastatin sodium | Pravastatin 20mg tablets (Accord Healthcare Ltd) |
| 60251 | 7997711000001109 | Pravastatin sodium | Pravastatin 10mg tablets (Sandoz Ltd) |
| 61134 | 15175111000001106 | Pravastatin sodium | Pravastatin 20mg tablets (Sigma Pharmaceuticals Plc) |
| 62979 | 8099711000001100 | Pravastatin sodium | Pravastatin 40mg tablets (Kent Pharmaceuticals Ltd) |
| 63074 | 11410611000001104 | Pravastatin sodium | Pravastatin 20mg tablets (PLIVA Pharma Ltd) |
| 63787 | 13761111000001104 | Pravastatin sodium | Pravastatin 10mg tablets (Tillomed Laboratories Ltd) |
| 67829 | 7998011000001108 | Pravastatin sodium | Pravastatin 20mg tablets (Sandoz Ltd) |
| 68156 | 7976411000001108 | Pravastatin sodium | Pravastatin 10mg tablets (A A H Pharmaceuticals Ltd) |
| 71015 | 19731611000001100 | Pravastatin sodium | Pravastatin 10mg tablets (Medreich Plc) |
| 72048 | 7959111000001108 | Pravastatin sodium | Pravastatin 40mg tablets (Actavis UK Ltd) |
| 72149 | 14957811000001102 | Pravastatin sodium | Pravastatin 5mg/ 5ml oral suspension |
| 75826 | | Pravastatin sodium | Pravastatin 20mg tablets (Actavis UK Ltd) |
| 77394 | | Pravastatin sodium | Lipostat 20mg tablets (Dowelhurst Ltd) |
| 379 | 320022002 | Fluvastatin sodium | Fluvastatin 20mg capsules |
| 2137 | 320023007 | Fluvastatin sodium | Fluvastatin 40mg capsules |
| 5985 | 378111000001106 | Fluvastatin sodium | Lescol XL 80mg tablets (Novartis Pharmaceuticals UK Ltd) |
| 8380 | 84811000001104 | Fluvastatin sodium | Lescol 20mg capsules (Novartis Pharmaceuticals UK Ltd) |
| 9153 | 409611000001108 | Fluvastatin sodium | Lescol 40mg capsules (Novartis Pharmaceuticals UK Ltd) |
| 11627 | 36566411000001105 | Fluvastatin sodium | Fluvastatin 80mg modified-release tablets |
| 53770 | 14584511000001106 | Fluvastatin sodium | Fluvastatin 40mg capsules (A A H Pharmaceuticals Ltd) |
| 59278 | 14036511000001107 | Fluvastatin sodium | Fluvastatin 20mg capsules (Zentiva) |
| 62148 | 16237911000001102 | Fluvastatin sodium | Fluvastatin 20mg capsules (Actavis UK Ltd) |
| 67328 | 16499511000001107 | Fluvastatin sodium | Lescol XL 80mg tablets (Mawdsley-Brooks & Company Ltd) |
| 71029 | 20289511000001104 | Fluvastatin sodium | Fluvastatin 40mg capsules (Sandoz Ltd) |
| 72308 | 14037111000001100 | Fluvastatin sodium | Fluvastatin 20mg capsules (Alliance Healthcare (Distribution) Ltd) |
| 73383 | 16238111000001104 | Fluvastatin sodium | Fluvastatin 40mg capsules (Actavis UK Ltd) |
| 74085 | 14765111000001100 | Fluvastatin sodium | Lescol 40mg capsules (Sigma Pharmaceuticals Plc) |
| 77306 | | Fluvastatin sodium | Nandovar XL 80mg tablets (Sandoz Ltd) |
| 77425 | | Fluvastatin sodium | Lescol 20mg capsules (Sigma Pharmaceuticals Plc) |

| | | | |
|---|---|---|---|
| 77472 | | Fluvastatin sodium | Lescol 40mg capsules (Lexon (UK) Ltd) |
| 420 | 320035006 | Cerivastatin sodium | Cerivastatin 100microgram tablets |
| 4961 | 22624500100002710 1 | Cerivastatin sodium | Lipobay 300microgram Tablet (Bayer Plc) |
| 5009 | 320036007 | Cerivastatin sodium | Cerivastatin 200microgram tablets |
| 5251 | 320037003 | Cerivastatin sodium | Cerivastatin 300microgram tablets |
| 5278 | 320041004 | Cerivastatin sodium | Cerivastatin 400microgram tablets |
| 9315 | 226225001000027104 | Cerivastatin sodium | Lipobay 100microgram Tablet (Bayer Plc) |
| 9316 | 226235001000027102 | Cerivastatin sodium | Lipobay 200microgram Tablet (Bayer Plc) |
| 18442 | 201215001000027108 | Cerivastatin sodium | Lipobay 400microgram Tablet (Bayer Plc) |
| 31658 | 134491009 | Cerivastatin sodium | Cerivastatin 800microgram tablets |
| 53813 | 4535911000001106 | Cerivastatin sodium | Lipobay 100microgram tablets (Bayer Plc) |
| 55207 | 4537511000001108 | Cerivastatin sodium | Lipobay 200microgram tablets (Bayer Plc) |
| 58480 | 4566311000001105 | Cerivastatin sodium | Lipobay 300microgram tablets (Bayer Plc) |
| 62132 | 4538111000001103 | Cerivastatin sodium | Lipobay 400microgram tablets (Bayer Plc) |
| 28 | 320029006 | Atorvastatin calcium trihydrate | Atorvastatin 10mg tablets |
| 75 | 320030001 | Atorvastatin calcium trihydrate | Atorvastatin 20mg tablets |
| 745 | 320031002 | Atorvastatin calcium trihydrate | Atorvastatin 40mg tablets |
| 2955 | 484211000001108 | Atorvastatin calcium trihydrate | Lipitor 40mg tablets (Pfizer Ltd) |
| 3411 | 643911000001108 | Atorvastatin calcium trihydrate | Lipitor 10mg tablets (Pfizer Ltd) |
| 5775 | 134489001 | Atorvastatin calcium trihydrate | Atorvastatin 80mg tablets |
| 7374 | 232011000001102 | Atorvastatin calcium trihydrate | Lipitor 20mg tablets (Pfizer Ltd) |
| 17683 | 756111000001109 | Atorvastatin calcium trihydrate | Lipitor 80mg tablets (Pfizer Ltd) |
| 47065 | 19722511000001105 | Atorvastatin calcium trihydrate | Atorvastatin 20mg chewable tablets sugar free |
| 47090 | 19722411000001106 | Atorvastatin calcium trihydrate | Atorvastatin 10mg chewable tablets sugar free |
| 47630 | 19719611000001109 | Atorvastatin calcium trihydrate | Lipitor 20mg chewable tablets (Pfizer Ltd) |
| 47721 | 19719311000001104 | Atorvastatin calcium trihydrate | Lipitor 10mg chewable tablets (Pfizer Ltd) |
| 48346 | 20528611000001105 | Atorvastatin calcium trihydrate | Atorvastatin 60mg tablets |
| 48518 | 14158611000001100 | Atorvastatin calcium trihydrate | Atorvastatin 10mg/ 5ml oral solution |
| 48973 | 20528511000001106 | Atorvastatin calcium trihydrate | Atorvastatin 30mg tablets |
| 49558 | 20491911000001107 | Atorvastatin calcium trihydrate | Atorvastatin 20mg tablets (A A H Pharmaceuticals Ltd) |
| 49751 | 20508311000001106 | Atorvastatin calcium trihydrate | Atorvastatin 40mg tablets (Alliance Healthcare (Distribution) Ltd) |
| 50236 | 20576911000001102 | Atorvastatin calcium trihydrate | Atorvastatin 10mg tablets (Zentiva) |
| 50272 | 20978811000001102 | Atorvastatin calcium trihydrate | Atorvastatin 40mg tablets (Pfizer Ltd) |
| 50788 | 20978111000001109 | Atorvastatin calcium trihydrate | Atorvastatin 20mg tablets (Pfizer Ltd) |
| 50790 | 20448411000001102 | Atorvastatin calcium trihydrate | Atorvastatin 20mg tablets (Dexcel-Pharma Ltd) |
| 50963 | 20494711000001108 | Atorvastatin calcium trihydrate | Atorvastatin 40mg tablets (Teva UK Ltd) |
| 51134 | 20491711000001105 | Atorvastatin calcium trihydrate | Atorvastatin 10mg tablets (A A H Pharmaceuticals Ltd) |
| 51200 | 20570211000001109 | Atorvastatin calcium trihydrate | Atorvastatin 40mg tablets (Arrow Generics Ltd) |
| 51359 | 20569511000001105 | Atorvastatin calcium trihydrate | Atorvastatin 20mg tablets (Arrow Generics Ltd) |
| 51622 | 20529611000001101 | Atorvastatin calcium trihydrate | Atorvastatin 20mg tablets (Consilient Health Ltd) |
| 51876 | 20529811000001102 | Atorvastatin calcium trihydrate | Atorvastatin 40mg tablets (Consilient Health Ltd) |
| 52097 | 20573211000001104 | Atorvastatin calcium trihydrate | Atorvastatin 40mg tablets (Wockhardt UK Ltd) |
| 52168 | 20496611000001108 | Atorvastatin calcium trihydrate | Atorvastatin 20mg tablets (Aspire Pharma Ltd) |

| | | | |
|---|---|---|---|
| 52211 | 20482911000001102 | Atorvastatin calcium trihydrate | Atorvastatin 20mg tablets (Actavis UK Ltd) |
| 52397 | 20982611000001106 | Atorvastatin calcium trihydrate | Atorvastatin 40mg tablets (Dr Reddy's Laboratories (UK) Ltd) |
| 52398 | 20492311000001102 | Atorvastatin calcium trihydrate | Atorvastatin 40mg tablets (A A H Pharmaceuticals Ltd) |
| 52459 | 20483311000001108 | Atorvastatin calcium trihydrate | Atorvastatin 80mg tablets (Actavis UK Ltd) |
| 52460 | 20496911000001102 | Atorvastatin calcium trihydrate | Atorvastatin 40mg tablets (Aspire Pharma Ltd) |
| 52821 | 20982911000001100 | Atorvastatin calcium trihydrate | Atorvastatin 80mg tablets (Dr Reddy's Laboratories (UK) Ltd) |
| 53594 | 16507511000001107 | Atorvastatin calcium trihydrate | Lipitor 80mg tablets (Mawdsley-Brooks & Company Ltd) |
| 53772 | 20508511000001100 | Atorvastatin calcium trihydrate | Atorvastatin 80mg tablets (Alliance Healthcare (Distribution) Ltd) |
| 53887 | 20483111000001106 | Atorvastatin calcium trihydrate | Atorvastatin 40mg tablets (Actavis UK Ltd) |
| 53890 | 20979511000001106 | Atorvastatin calcium trihydrate | Atorvastatin 80mg tablets (Pfizer Ltd) |
| 54535 | 20977811000001101 | Atorvastatin calcium trihydrate | Atorvastatin 10mg tablets (Pfizer Ltd) |
| 54992 | 14158711000001109 | Atorvastatin calcium trihydrate | Atorvastatin 10mg/ 5ml oral suspension |
| 55032 | 20448211000001101 | Atorvastatin calcium trihydrate | Atorvastatin 10mg tablets (Dexcel-Pharma Ltd) |
| 55034 | 14158911000001106 | Atorvastatin calcium trihydrate | Atorvastatin 40mg/ 5ml oral suspension |
| 55444 | 20577411000001107 | Atorvastatin calcium trihydrate | Atorvastatin 40mg tablets (Zentiva) |
| 55727 | 20482711000001104 | Atorvastatin calcium trihydrate | Atorvastatin 10mg tablets (Actavis UK Ltd) |
| 56016 | 19719611000001109 | Atorvastatin calcium trihydrate | Lipitor 20mg chewable tablets (Pfizer Ltd) |
| 56097 | 19722411000001106 | Atorvastatin calcium trihydrate | Atorvastatin 10mg chewable tablets sugar free |
| 56165 | 19722511000001105 | Atorvastatin calcium trihydrate | Atorvastatin 20mg chewable tablets sugar free |
| 56182 | 20577611000001105 | Atorvastatin calcium trihydrate | Atorvastatin 80mg tablets (Zentiva) |
| 56248 | 14198211000001106 | Atorvastatin calcium trihydrate | Atorvastatin 20mg tablets (Sigma Pharmaceuticals Plc) |
| 56564 | 22047511000001102 | Atorvastatin calcium trihydrate | Atorvastatin 20mg tablets (Almus Pharmaceuticals Ltd) |
| 56841 | 20448611000001104 | Atorvastatin calcium trihydrate | Atorvastatin 40mg tablets (Dexcel-Pharma Ltd) |
| 57117 | 21779811000001108 | Atorvastatin calcium trihydrate | Atorvastatin 80mg tablets (Waymade Healthcare Plc) |
| 57348 | 20529411000001104 | Atorvastatin calcium trihydrate | Atorvastatin 10mg tablets (Consilient Health Ltd) |
| 57834 | 22613211000001109 | Atorvastatin calcium trihydrate | Atorvastatin 40mg tablets (DE Pharmaceuticals) |
| 57836 | 20495411000001101 | Atorvastatin calcium trihydrate | Atorvastatin 80mg tablets (Teva UK Ltd) |
| 58041 | 20494411000001102 | Atorvastatin calcium trihydrate | Atorvastatin 20mg tablets (Teva UK Ltd) |
| 58110 | 20577211000001108 | Atorvastatin calcium trihydrate | Atorvastatin 20mg tablets (Zentiva) |
| 58394 | 20508111000001109 | Atorvastatin calcium trihydrate | Atorvastatin 20mg tablets (Alliance Healthcare (Distribution) Ltd) |
| 58418 | 20492611000001107 | Atorvastatin calcium trihydrate | Atorvastatin 80mg tablets (A A H Pharmaceuticals Ltd) |
| 58742 | 20570911000001100 | Atorvastatin calcium trihydrate | Atorvastatin 80mg tablets (Arrow Generics Ltd) |
| 58834 | 13831111000001108 | Atorvastatin calcium trihydrate | Atorvastatin 10mg tablets (DE Pharmaceuticals) |
| 58868 | 14197911000001103 | Atorvastatin calcium trihydrate | Atorvastatin 10mg tablets (Sigma Pharmaceuticals Plc) |
| 59272 | 21779411000001106 | Atorvastatin calcium trihydrate | Atorvastatin 20mg tablets (Waymade Healthcare Plc) |
| 59331 | 13922611000001102 | Atorvastatin calcium trihydrate | Lipitor 10mg tablets (DE Pharmaceuticals) |
| 59357 | 22940511000001105 | Atorvastatin calcium trihydrate | Atorvastatin 10mg tablets (Ranbaxy (UK) Ltd) |
| 59446 | 22047711000001107 | Atorvastatin calcium trihydrate | Atorvastatin 40mg tablets (Almus Pharmaceuticals Ltd) |
| 59776 | 20497611000001105 | Atorvastatin calcium trihydrate | Atorvastatin 80mg tablets (Aspire Pharma Ltd) |
| 59859 | 20492811000001106 | Atorvastatin calcium trihydrate | Atorvastatin 10mg tablets (Teva UK Ltd) |
| 60464 | 14018311000001109 | Atorvastatin calcium trihydrate | Atorvastatin 20mg/ 5ml oral suspension |
| 60511 | 22941211000001101 | Atorvastatin calcium trihydrate | Atorvastatin 40mg tablets (Ranbaxy (UK) Ltd) |
| 60607 | 22613511000001107 | Atorvastatin calcium trihydrate | Atorvastatin 80mg tablets (DE Pharmaceuticals) |

| | | | |
|---|---|---|---|
| 60989 | 22202511000001100 | Atorvastatin calcium trihydrate | Atorvastatin 80mg tablets (Phoenix Healthcare Distribution Ltd) |
| 61149 | 21779211000001107 | Atorvastatin calcium trihydrate | Atorvastatin 10mg tablets (Waymade Healthcare Plc) |
| 62219 | 22613011000001104 | Atorvastatin calcium trihydrate | Atorvastatin 20mg tablets (DE Pharmaceuticals) |
| 62429 | 13831511000001104 | Atorvastatin calcium trihydrate | Atorvastatin 20mg tablets (DE Pharmaceuticals) |
| 62476 | 22047911000001109 | Atorvastatin calcium trihydrate | Atorvastatin 80mg tablets (Almus Pharmaceuticals Ltd) |
| 63140 | 20507911000001106 | Atorvastatin calcium trihydrate | Atorvastatin 10mg tablets (Alliance Healthcare (Distribution) Ltd) |
| 63249 | 20513411000001108 | Atorvastatin calcium trihydrate | Atorvastatin 80mg tablets (Consilient Health Ltd) |
| 63469 | 20512911000001107 | Atorvastatin calcium trihydrate | Atorvastatin 30mg tablets (Consilient Health Ltd) |
| 64067 | 14018211000001101 | Atorvastatin calcium trihydrate | Atorvastatin 20mg/ 5ml oral solution |
| 64702 | 21099211000001101 | Atorvastatin calcium trihydrate | Atorvastatin 30mg tablets (A A H Pharmaceuticals Ltd) |
| 64810 | 22202111000001109 | Atorvastatin calcium trihydrate | Atorvastatin 40mg tablets (Phoenix Healthcare Distribution Ltd) |
| 64825 | 22201511000001108 | Atorvastatin calcium trihydrate | Atorvastatin 10mg tablets (Phoenix Healthcare Distribution Ltd) |
| 64868 | 29772511000001108 | Atorvastatin calcium trihydrate | Atorvastatin 40mg tablets (Sigma Pharmaceuticals Plc) |
| 65193 | 22940911000001103 | Atorvastatin calcium trihydrate | Atorvastatin 20mg tablets (Ranbaxy (UK) Ltd) |
| 66963 | 29772911000001101 | Atorvastatin calcium trihydrate | Atorvastatin 80mg tablets (Sigma Pharmaceuticals Plc) |
| 67402 | 32395011000001108 | Atorvastatin calcium trihydrate | Atorvastatin 40mg tablets (Kent Pharmaceuticals Ltd) |
| 67573 | 22612811000001102 | Atorvastatin calcium trihydrate | Atorvastatin 10mg tablets (DE Pharmaceuticals) |
| 67660 | 22941511000001103 | Atorvastatin calcium trihydrate | Atorvastatin 80mg tablets (Ranbaxy (UK) Ltd) |
| 67846 | 22047311000001108 | Atorvastatin calcium trihydrate | Atorvastatin 10mg tablets (Almus Pharmaceuticals Ltd) |
| 68023 | 20496111000001100 | Atorvastatin calcium trihydrate | Atorvastatin 10mg tablets (Aspire Pharma Ltd) |
| 68048 | 22201711000001103 | Atorvastatin calcium trihydrate | Atorvastatin 20mg tablets (Phoenix Healthcare Distribution Ltd) |
| 68467 | 32394811000001103 | Atorvastatin calcium trihydrate | Atorvastatin 20mg tablets (Kent Pharmaceuticals Ltd) |
| 68785 | 33556411000001105 | Atorvastatin calcium trihydrate | Atorvastatin 10mg tablets (Mylan) |
| 68827 | 33556611000001108 | Atorvastatin calcium trihydrate | Atorvastatin 20mg tablets (Mylan) |
| 69093 | 20573911000001108 | Atorvastatin calcium trihydrate | Atorvastatin 80mg tablets (Wockhardt UK Ltd) |
| 69427 | 33556811000001107 | Atorvastatin calcium trihydrate | Atorvastatin 40mg tablets (Mylan) |
| 70693 | 29771911000001107 | Atorvastatin calcium trihydrate | Atorvastatin 10mg tablets (Sigma Pharmaceuticals Plc) |
| 70987 | 20982011000001104 | Atorvastatin calcium trihydrate | Atorvastatin 10mg tablets (Dr Reddy's Laboratories (UK) Ltd) |
| 71017 | 20982411000001108 | Atorvastatin calcium trihydrate | Atorvastatin 20mg tablets (Dr Reddy's Laboratories (UK) Ltd) |
| 72164 | 34961811000001104 | Atorvastatin calcium trihydrate | Atorvastatin 20mg tablets (Bristol Laboratories Ltd) |
| 72213 | 20572711000001103 | Atorvastatin calcium trihydrate | Atorvastatin 20mg tablets (Wockhardt UK Ltd) |
| 72641 | 14158811000001101 | Atorvastatin calcium trihydrate | Atorvastatin 40mg/ 5ml oral solution |
| 73520 | 13923011000001100 | Atorvastatin calcium trihydrate | Lipitor 20mg tablets (DE Pharmaceuticals) |
| 74518 | 20572511000001108 | Atorvastatin calcium trihydrate | Atorvastatin 10mg tablets (Wockhardt UK Ltd) |
| 75391 | 34682311000001101 | Atorvastatin calcium trihydrate | Atorvastatin 80mg/ 5ml oral suspension |
| 75622 | | Atorvastatin calcium trihydrate | Atorvastatin 60mg tablets (A A H Pharmaceuticals Ltd) |
| 77344 | | Atorvastatin calcium trihydrate | Atorvastatin 80mg tablets (Mylan) |
| 77427 | | Atorvastatin calcium trihydrate | Lipitor 10mg tablets (Waymade Healthcare Plc) |
| 77434 | | Atorvastatin calcium trihydrate | Lipitor 20mg tablets (Waymade Healthcare Plc) |
| 77782 | | Atorvastatin calcium trihydrate | Atorvastatin 40mg tablets (Bristol Laboratories Ltd) |
| 24509 | | | SIMVASTATIN |
| 29438 | | | SIMVASTATIN |
